# Progression Subtypes in Parkinson’s Disease: A Data-driven Multi-Cohort Analysis

**DOI:** 10.1101/2023.10.12.23296943

**Authors:** Tom Hähnel, Tamara Raschka, Stefano Sapienza, Jochen Klucken, Enrico Glaab, Jean-Christophe Corvol, Björn Falkenburger, Holger Fröhlich

## Abstract

**Background:** The progression of Parkinson’s disease (PD) is heterogeneous across patients. This heterogeneity complicates patients counseling and inflates the number of patients needed to test potential neuroprotective treatments. Moreover, disease subtypes might require different therapies. This work uses a data-driven approach to investigate how observed heterogeneity in PD can be explained by the existence of distinct PD progression subtypes.

**Methods:** To derive stable PD progression subtypes in an unbiased manner, we analyzed multimodal longitudinal data from three large PD cohorts. A latent time joint mixed-effects model (LTJMM) was used to align patients on a common disease timescale. Progression subtypes were identified by variational deep embedding with recurrence (VaDER). These subtypes were then characterized across the three cohorts using clinical scores, DaTSCAN imaging and digital gait biomarkers. To assign patients to progression subtypes from baseline data, we developed predictive models and performed extensive cross-cohort validation.

**Results:** In each cohort, we identified a fast-progressing and a slow-progressing subtype. These subtypes were reflected by different patterns of motor and non-motor symptoms progression, survival rates, treatment response and features extracted from DaTSCAN imaging and digital gait assessments. Predictive models achieved robust performance with ROC-AUC up to 0.79 for subtype identification. Simulations demonstrated that enriching clinical trials with fast-progressing patients based on predictions from baseline can reduce the required cohort size by 43%.

**Conclusion:** Our results show that heterogeneity in PD can be explained by two distinct subtypes of PD progression that are stable across cohorts and can be predicted from baseline data. These subtypes align with the brain-first vs. body-first concept, which potentially provides a biological explanation for subtype differences. The predictive models will enable clinical trials with significantly lower sample sizes by enriching fast-progressing patients.

## Introduction

Parkinson’s disease (PD) is the fastest-growing neurological disease and the second most common neurodegenerative disease.^1^ Recent randomized clinical trials (RCTs) have investigated potentially disease-modifying treatments, but have failed to reach their primary endpoints.^2–5^ This raises the question of whether our understanding of PD pathogenesis is insufficient or whether RCTs were inadequately designed to demonstrate treatment effects on disease progression. The high heterogeneity observed in people with PD (PwPD)^6^ limits the statistical power of clinical trials. Furthermore, the observed heterogeneity suggests the existence of PD subtypes which might show different treatment responses.

The construct of PD as a heterogeneous group of different subtypes has been proposed in several concepts. Some concepts categorize PwPD by single clinical features like age of onset, motor phenotype or onset of dementia.^7^ The brain-first vs. body-first concept explains heterogeneity observed in imaging data by different routes of alpha-synuclein spreading through the nervous system.^8^ This model is further extended by the alpha-synuclein origin site and connectome (SOC) model which suggests that alpha-synuclein spreading spreading from one brain hemisphere to the other is less common.^9^ Other researchers identified subtypes using data-driven methods and machine learning.^10–14^ These approaches have the advantages of being hypothesis-free and being able to capture more complex patterns from multivariate data. Subtypes were mostly inferred based on cross-sectional differences^10^, but some researchers also investigated differences in disease progression using longitudinal data from single cohorts.^11,12^

Our study aims to identify PD subtypes with a focus on differences in disease *progression,* inferred from multimodal longitudinal cohort data. We extensively characterized PD subtypes regarding differences in motor and non-motor symptom progression, mortality, treatment response, DaTSCAN imaging and digital gait biomarkers. In particular, we investigated the generalizability of our findings by external validation in additional and highly diverse cohorts. Further, we developed a strategy to enrich for PwPD of one subtype within a study cohort. We then analyzed how this enrichment reduces the required sample size and increases the statistical power of clinical trials.

## Material and methods

### Clinical cohorts

We analyzed PwPD from three cohort studies: (I) de-novo PwPD from the Parkinson’s Progression Markers Initiative (PPMI, NCT04477785), (II) early disease stage PwPD from the French ICEBERG cohort study (NCT02305147), and (III) PwPD from all disease stages from the Luxembourg Parkinson’s Study (LuxPARK, NCT05266872)^15^. Besides clinical scores, PwPD in PPMI underwent repeated DaTSCANs and LuxPARK PwPD had a single standardized gait assessment. Inclusion and exclusion criteria details are presented in the supplement. Ethical committee approval and subject’s consent according to the Declaration of Helsinki has been obtained by the individual study groups.

### Aligning PwPD trajectories on a common disease timescale

To address temporal heterogeneity between the cohorts, we aligned PwPD on a comparable *common disease timescale* using a latent time joint mixed-effects model (LTJMM).^16^ In brief, LTJMM models a linear progression of multiple clinical outcomes over time and estimates the deviance of an individual PwPD’s progression compared to a “mean PwPD”. Thereby, we estimated how much the timescale of each individual PwPD is shifted from the timescale of the mean PwPD, i.e. where the PwPD is aligned on a common disease timescale using the mean PwPD as reference. For instance, a PwPD with diagnosis in a very early PD stage may exhibit a negative time since diagnosis on the common disease timescale as a mean PwPD won’t be diagnosed at this time. In contrast, PwPD diagnosed at a more advanced stage of PD will present with a higher time since diagnosis at the common disease timescale. To achieve comparability, we used time since diagnosis as timescale in all cohorts. The following clinical scores were used as outcomes in LTJMM as they measure a wide variety of motor and non-motor symptoms and were assessed in all three cohorts: Unified Parkinson’s Disease Rating Scale (UPDRS) I-IV, Postural Instability and Gait Dysfunction score (PIGD), Montreal Cognitive Assessment (MoCA) and Scales for Outcomes in Parkinson’s Disease-Autonomic Dysfunction (SCOPA). We accounted for age and sex as covariates. Formulas and implementation details are provided in the supplement.

### Subtype identification using VaDER

We identified PD progression subtypes using variational deep embedding with recurrence (VaDER).^17^ VaDER implements long short-term memory (LSTM) networks to handle longitudinal data as input. Subtypes are identified on a low-dimensional representation of the data via a variational autoencoder. Using these techniques, VaDER identifies subtypes in multivariate longitudinal data for short time series. We used the predicted LTJMM outcome trajectories of UPDRS I-IV, PIGD, MoCA and SCOPA on the common disease timescale as inputs for VaDER. Thereby, we assign each PwPD to one disease progression subtype. Details of VaDER fitting and hyperparameter optimization are presented in the supplement (Table S1).

### Cross-cohort validation

To achieve optimal representation in each cohort, LTJMM and VaDER were applied separately to each cohort. Generalizability of our findings was evaluated by a cross-cohort validation using PPMI for training and validating the VaDER and predictive models on ICEBERG and LuxPARK. PPMI was chosen as training model as it is publicly available.

### Symptom domain comparisons

We modeled the progression of 114 different outcomes (including single questions, scores and sub-scores from questionnaires and clinical assessments) using linear mixed-effects models, binary mixed-effects models and ordinal mixed-effects models depending on the scale of the outcome. Standardized mean differences (SMD) of coefficients between subtypes were calculated, reflecting the subtype differences in progression speed. The 114 outcomes were grouped into 22 different symptom domains which were assessed in all three cohorts (Table S2). For each symptom domain, a three-level meta-analysis was performed, thereby providing an overall estimate of the symptom domain progression difference between subtypes across all cohorts.

To address baseline associations with the subtypes, we applied a similar approach with logistic regression models. Using three-level meta-analyses, we obtained overall regression coefficients for symptom domains across cohorts.

P-values and confidence intervals (CIs) were adjusted for multiple testing using the Benjamini-Hochberg procedure. Details are provided in the supplement.

### Survival analysis

To compare mortality between subtypes, we performed a survival analysis for LuxPARK using a Cox proportional hazards model with subtype, age and sex as covariates and the common disease timescale as time variable. The analysis was implemented using the python lifelines package.^18^

### Treatment response analysis

UPDRS III in PPMI is reported at annual visits in the OFF state defined by last medication intake at least 6 hours ago and after medication intake in the ON state. We calculated the treatment response as the relative improvement in UPDRS III after medication intake and averaged responses over all clinical visits at which UPDRS III was performed in ON and OFF state.

### DaTSCAN analysis

In PPMI, DaTSCAN analysis was performed at screening visit and up to three additional visits. We compared signal binding ratio (SBR) and asymmetry index between subtypes obtained at screening visits using a t-test. For PwPD with longitudinal DaTSCAN measurements available, we modeled SBR and asymmetry index changes over time using a linear mixed-effects model. Subsequently, we compared the obtained progression rates of SBR and asymmetry index between subtypes using a t-test.

### Gait analysis

In LuxPARK, PwPD completed a standardized gait assessment at one visit using the automated gait assessment system eGaIT.^19^ PwPD underwent a timed up and go (TUG) task using accelerometer and gyroscope sensors attached to their shoes. 15 digital gait parameters were calculated based on straight steps from the TUG task. Gait differences between subtypes were analyzed by conducting an ANCOVA and controlling for disease duration on the common disease timescale. P-values and CI were corrected for multiple testing using the Benjamini-Hochberg procedure. The digital gait parameters are described Table S3.

### Subtype prediction models

We predicted PwPD subtypes from (I) baseline and (II) baseline and one additional visit after one year using logistic regression, random forest and eXtreme Gradient Boosting (XGBoost) with repeated nested 5-fold cross-validation (Table S4). UPDRS I-III, PIGD, MoCA and SCOPA were used as predictors. Cross-cohort validation was performed using the model trained on PPMI and validating this model on ICEBERG and LuxPARK. Details regarding the predictive models are described in the supplement.

### Sample size estimation using subtype predictions

Based on the prediction obtained from the predictive models described above, we simulated PwPD cohorts with different percentages of fast-progressing PwPD and assessed the sample sizes which would be required for a RCT investigating a potentially disease-modifying drug. Our simulations are based on considerations from an ongoing trial^20^. Therefore, we assumed 30% treatment effect on disease progression, UPDRS I-III sum score as primary outcome, one year study duration, 80% power and a significance level of 0.1. Details regarding the sample size estimations are described in the supplement.

### Statistical analysis

For comparison of cohort characteristics, the following tests were applied: Sex was compared using Fisher’s exact test, Hoehn & Yahr using Kolmogorov-Smirnov test and all other characteristics were compared using Mann-Whitney U test. P-values were adjusted for multiple testing using the Benjamini-Hochberg procedure.

All statistical tests were conducted as two-tailed tests with significance level 0.05.

## Results

### Demographic and clinical characteristics

Overall, 1,124 PwPD were analyzed. In general, the cohorts exhibited different clinical characteristics related to disease duration at baseline: LuxPARK included advanced disease stages compared to ICEBERG and PPMI with mostly early disease stages (Table 1). Significant differences across cohorts were observed for age, disease duration, Hoehn & Yahr stage, UPDRS I-IV, PIGD, MoCA and SCOPA.

**Table 1:** Demographic and clinical characteristics. PwPD baseline characteristics and study characteristics for PPMI, ICEBERG and LuxPARK cohort. For sex and H&Y, relative and absolute frequencies are shown. For other characteristics, median and first/third quartiles are reported. Corresponding p-values were corrected for multiple testing. UPDRS IV was not assessed at baseline in PPMI. Significant p-values are emphasized in italic. Abbreviations: H&Y: Hoehn & Yahr, MoCA: Montreal Cognitive Assessment, PIGD: Postural Instability and Gait Dysfunction score, SCOPA: Scales for Outcomes in Parkinson’s Disease-Autonomic Dysfunction, UPDRS: Unified Parkinson’s Disease Rating Scale.

|  | PPMI | ICEBERG | LuxPARK | PPMI vs<br>ICEBERG<br>p-values | PPMI vs<br>LuxPARK<br>p-values | ICEBERG<br>vs LuxPARK<br>p-values |
| --- | --- | --- | --- | --- | --- | --- |
| **Number of PwPD** | 409 | 154 | 561 | .. | .. | .. |
| **Age, years** | 63.0 [55.2-69.3] | 63.7 [57.1-69.4] | 67.8 [59.4-73.1] | 1.0 | <0.0001 | <0.0001 |
| **Sex** | .. | .. | .. | 0.71 | 1.0 | 0.49 |
| Male | 66.7% [273] | 62.3% [96] | 67.6% [379] | .. | .. | .. |
| Female | 33.3% [136] | 37.7% [58] | 32.4% [182] | .. | .. | .. |
| **Hoehn & Yahr** | .. | .. | .. | <0.0001 | <0.0001 | 0.0076 |
| H&Y I | 43.8% [179] | 2.6% [4] | 18.7% [105] | .. | .. | .. |
| H&Y II | 56.2% [230] | 93.5% [144] | 67.0% [376] | .. | .. | .. |
| H&Y III | 0 | 3.9% [6] | 9.1% [51] | .. | .. | .. |
| H&Y IV | 0 | 0 | 3.7% [21] | .. | .. | .. |
| H&Y V | 0 | 0 | 1.4% [8] | .. | .. | .. |
| **Disease duration, years** | 0.3 [0.2-0.6] | 1.2 [0.6-2.3] | 2.9 [0.9-6.5] | <0.0001 | <0.0001 | <0.0001 |
| **Number of visits** | 14 [12-16] | 5 [4-5] | 4 [3-6] | <0.0001 | <0.0001 | 0.13 |
| **Follow up, years** | 7.0 [5.0-7.0] | 4.1 [3.0-4.4] | 4.0 [2.4-5.0] | <0.0001 | <0.0001 | 1.0 |
| **UPDRS I** | 5 [3-7] | 9 [6-12] | 9 [5-13] | <0.0001 | <0.0001 | 1.0 |
| **UPDRS II** | 5 [3-8] | 8 [5-10] | 10 [5-15] | <0.0001 | <0.0001 | 0.002 |
| **UPDRS III** | 20 [14-26] | 29 [24-35] | 32 [22-44] | <0.0001 | <0.0001 | 0.047 |
| **UPDRS IV** | .. | 0 [0-0] | 0 [0-1] | .. | .. | <0.0001 |
| **PIGD** | 0.2 [0.0-0.4] | 0.2 [0.0-0.4] | 0.4 [0.2-1.0] | 0.26 | <0.0001 | <0.0001 |
| **MoCA** | 28 [26-29] | 28 [26-29] | 25 [23-28] | 0.11 | <0.0001 | <0.0001 |
| **SCOPA** | 8 [6-12] | 11 [7-17] | 14 [9-20] | 0.00034 | <0.0001 | 0.0015 |

### Identification of PD progression subtypes

After aligning PwPD to a common disease timescale (Fig. S1), we identified two distinct PD progression subtypes in the PPMI, ICEBERG and LuxPARK cohorts, assigned each PwPD to one of the progression subtypes and predicted the PD subtypes from baseline data (Fig. 1). We repeated these steps in a cross-cohort validation fashion to explore the generalizability of our approach. This was done by training our model on the publicly available PPMI data and using this model for PwPD subtype assignments and predictions in ICEBERG and LuxPARK.

**Figure 1:**
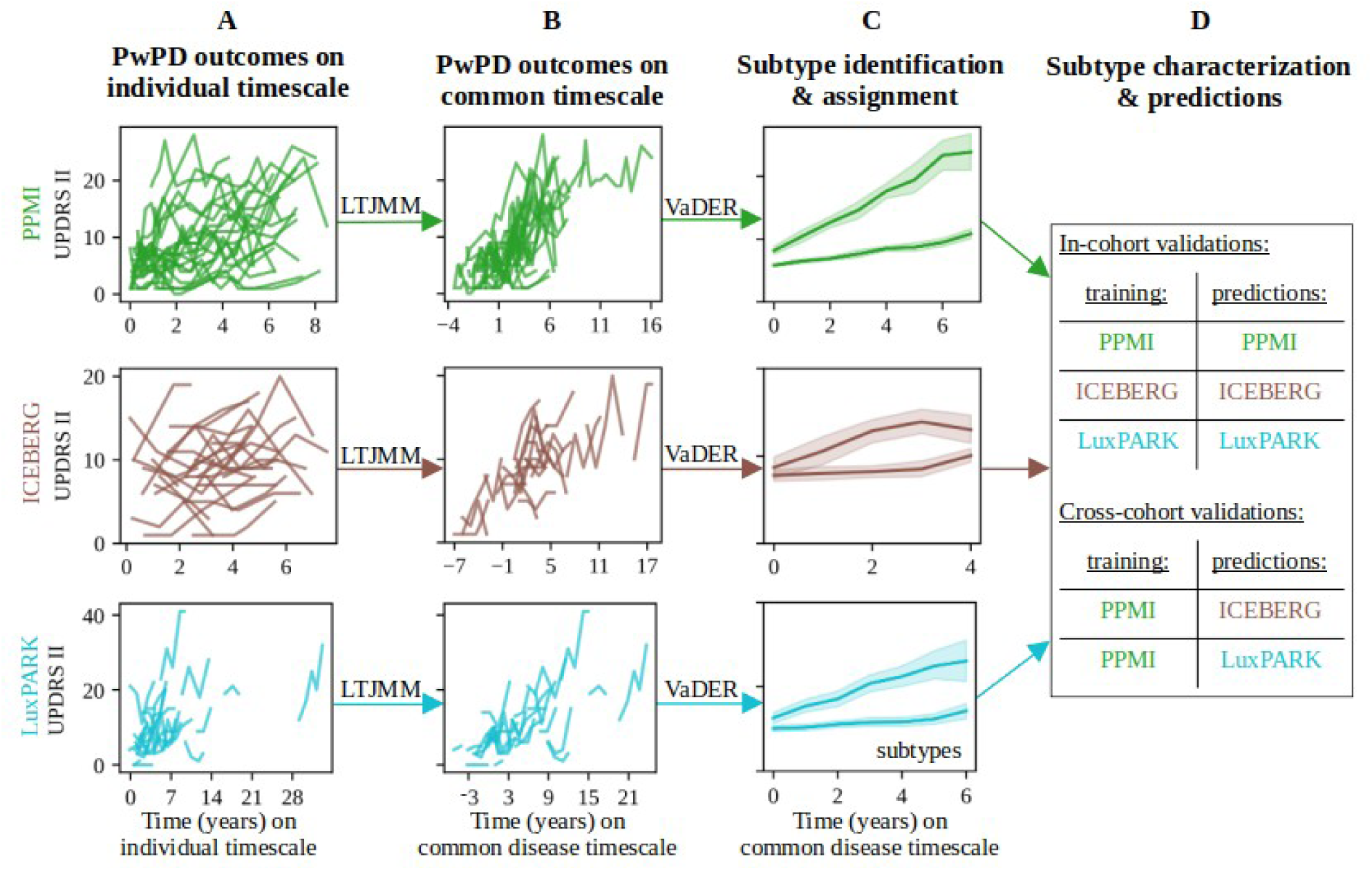
Model training and validation procedure for subtype identification and predictions. Individual PwPD outcomes (A) were aligned on a common timescale (B) using the latent time joint mixed-effects model (LTJMM). The UPDRS II values of 25 randomly sampled PwPD are shown for visualization. Subsequently, two distinct progression subtypes were identified (C) using a variational deep embedding with recurrence (VaDER). Subtypes were further characterized and models were trained to predict subtype associations from baseline (D). VaDER and predictive models were trained and evaluated on each cohort separately and results were compared across cohorts (in-cohort validation). Additionally, PPMI-trained models were applied to ICEBERG and LuxPARK and results were compared with results of the in-cohort approach (cross-cohort validation). Abbreviations: UPDRS: Unified Parkinson’s Disease Rating Scale

### PD subtypes exhibit different symptom characteristics

We explored baseline and progression characteristics of clinical symptoms between both PD subtypes. Focusing on the motor (UPDRS II/III/IV, PIGD) and non-motor (UPDRS I, MoCA, SCOPA) outcomes assessed in all three cohorts, we observed minor differences at baseline but large differences in progression speed. One subtype exhibited significantly faster progression for all symptoms and thereby was named *fast-progressing* subtype compared to the second *slow-progressing* subtype (Fig. 2A). Most PwPD were assigned to the slow-progressing subtype (PPMI: 335 slow/74 fast, ICEBERG: 112 slow/42 fast, LuxPARK: 408 slow/153 fast). While mean progression trajectories were clearly separated for most outcomes, we observed some overlap in ICEBERG for autonomic symptoms reported by SCOPA and also for cognition reported by MoCA. However, ICEBERG is the smallest cohort and PwPD in ICEBERG present with only minimal cognitive impairment at baseline. Also, the trend to more rapid progression in the fast-progressing subtype is still similar to PPMI and LuxPARK. We also observed some overlap of subtypes in terms of motor fluctuations reported by UPDRS IV in the LuxPARK cohort while there was a better separation for PPMI and ICEBERG.

**Figure 2:**
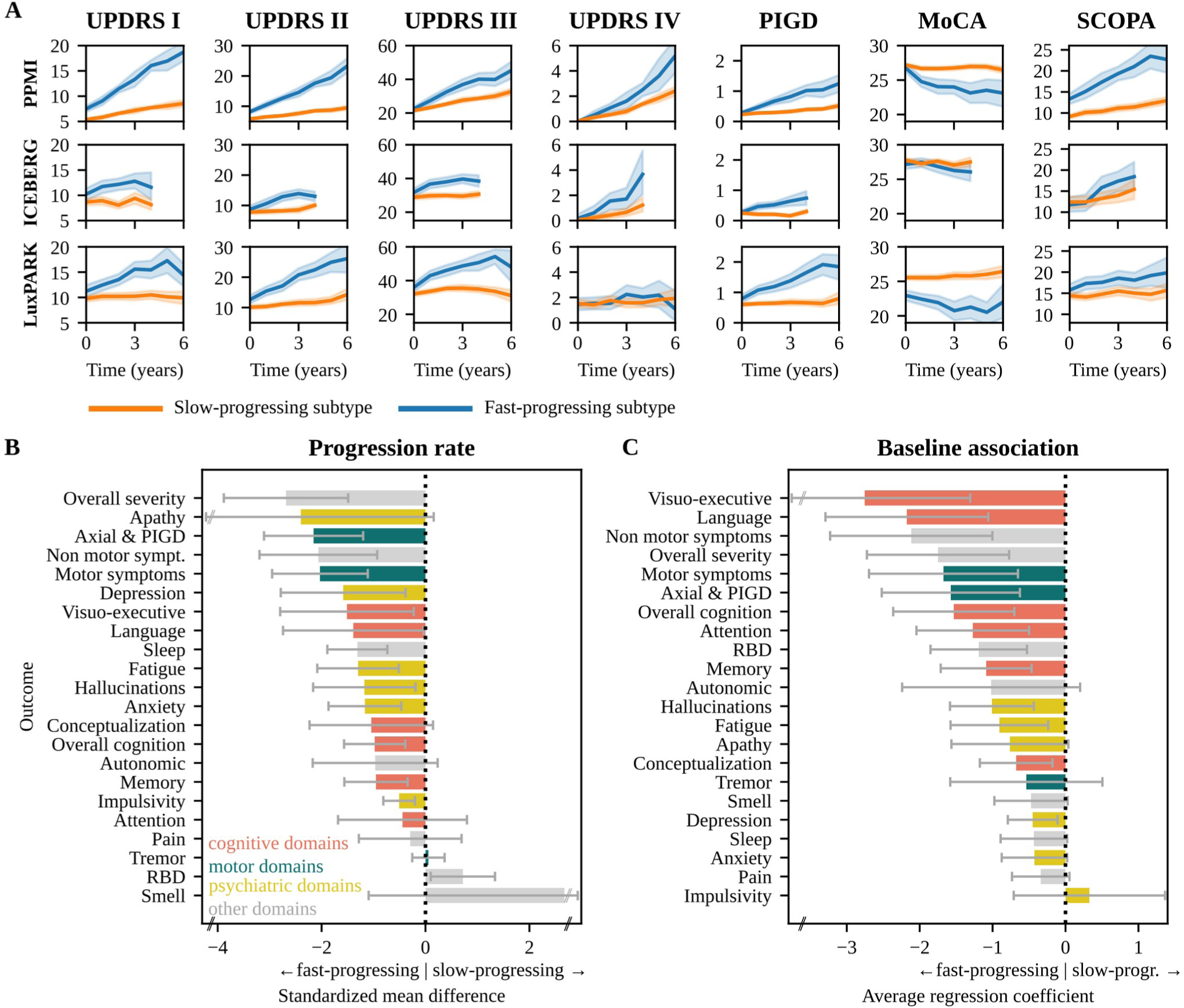
Disease progression and baseline characteristics of subtypes. **A:** Progression of motor scores (UPDRS II/III/IV, PIGD) and non-motor scores (UPDRS I, MoCA, SCOPA) for the slow-progressing subtype (orange) and fast-progressing subtype (blue) for PPMI, ICEBERG and LuxPARK. Mean and 95% confidence interval for each subtype are shown. ICEBERG data is shown up to four years as only a few ICEBERG PwPD had longer follow up. **B**: Standardized mean differences (SMD) of progression speed between both subtypes for different symptom domains (orange: cognition, green: motor, yellow: psychiatric, gray: other). Negative SMD values indicate that the fast-progressing subtype shows a faster progression. **C**: Average regression coefficients showing associations of symptom domains at baseline with subtypes. Negative values indicate that more severe symptoms at baseline are associated with the faster subtype. 95% confidence intervals are shown and were corrected for multiple testing. Abbreviations: MoCA: Montreal Cognitive Assessment, PIGD: Postural Instability and Gait Dysfunction score, SCOPA: Scales for Outcomes in Parkinson’s Disease-Autonomic Dysfunction, RBD: REM behavior sleep disorder, UPDRS: Unified Parkinson’s Disease Rating Scale.

The results shown in Fig. 2A comprise only a small subset of symptom domains affected in PD. We were wondering whether all symptoms progress more rapidly in the fast-progressing subtype, or whether the pattern of progression rates differs between the two subtypes. Therefore, we aggregated single questions, sub-scores and total scores from different assessments into 22 distinct symptom domains (Table S2). Indeed, overall disease severity, axial and PIGD symptoms progressed much more rapidly in the fast-progressing subtype as compared to the slow-progressing subtype (Fig. 2B). In contrast, the rate of progression for most cognitive domains differed less between both subtypes. Interestingly, there was no difference in tremor progression between subtypes, and the slow-progressing subtype exhibited faster progression of REM behavior sleep disorder (RBD) symptoms than the fast-progressing subtype.

Regarding subtype differences at baseline, cognitive domains exhibited more pronounced differences than other domains (Fig. 2C). Fast-progressing PwPD exhibited already higher RBD values at baseline, thereby providing an explanation for the slower RBD progression observed in the fast-progressing subtype. Similar to progression characteristics, tremor had no significant baseline association with subtypes. Visuo-executive function and language function exhibited the largest baseline and progression differences between both subtypes compared to the other cognitive domains.

Progression characteristics and baseline characteristics were mostly similar between the three cohorts and reproducible in the cross-cohort validation (Fig. S2-4), thereby supporting generalizability of these findings.

### Mortality and treatment response

Mortality data was only available for LuxPARK and showed an increased hazard ratio (HR) for death for the fast-progressing subtype (HR=3.4, 95% CI: 1.9 – 6.2, p<0.0001, Fig. 3A). Similar findings were obtained in the cross-cohort validation approach (HR=3.7, 95% CI: 1.9 – 7.1, p<0.0001, Fig. S5).

**Figure 3:**
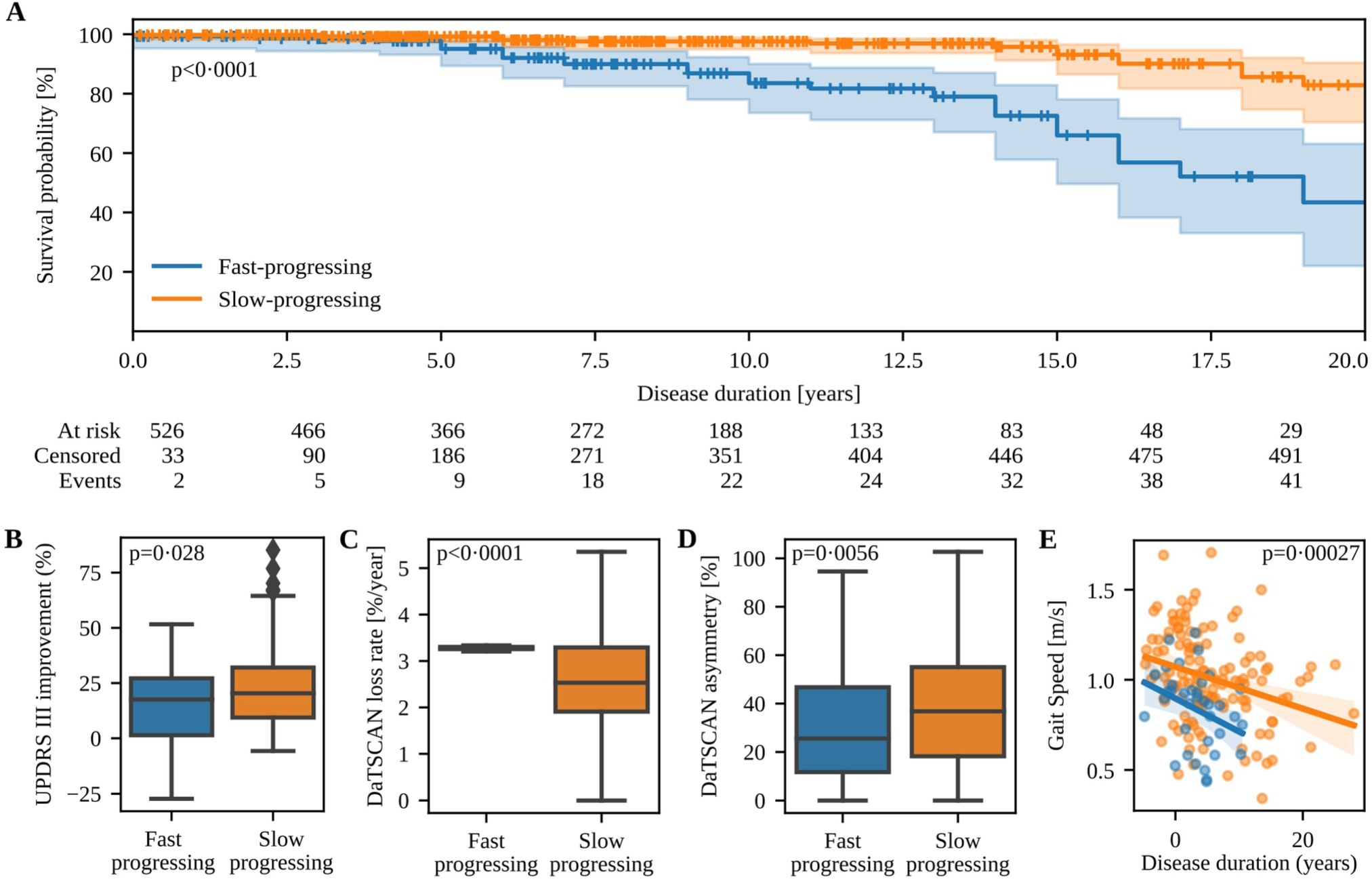
Mortality, treatment response and biomarker differences between subtypes. **A:** Kaplan-Meier estimator for survival probability on the common disease timescale for fast-progressing (blue) and slow-progressing (orange) PwPD in LuxPARK. Censored observations are indicated by small vertical ticks. The corresponding p-value for the subtype covariate from the cox proportional hazard model is reported. **B:** Mean UPDRS III improvement of PwPD in PPMI after dopaminergic drug intake compared to OFF state. **C:** Progression of DaTSCAN uptake loss for fast-progressing and slow-progressing progressing PwPD. **D:** DaTSCAN asymmetry index at baseline for slow-progressing and fast-progressing PwPD. **E:** Correlation of gait speed with disease duration on the common timescale for fast-progressing (blue) and slow-progressing (orange) PwPD. Only the most significant digital gait parameter is shown here while correlations of all gait parameters are presented in the supplement. The corresponding p-value from the ANCOVA analysis is shown and was corrected for multiple testing of all digital gait parameters. Abbreviations: UPDRS: Unified Parkinson’s Disease Rating Scale

Treatment responses were available for PPMI and indicated a worse response to dopaminergic treatment for fast-progressing PwPD (p=0.028, Fig. 3B).

### Imaging and gait biomarkers

In addition to clinical outcomes, we investigated whether subtype differences were also reflected by biomarkers. Baseline DaTSCAN imaging in PPMI showed no subtype difference (p=0.37), but DaTSCAN progression differed significantly (p<0.0001, Fig. 3C). DaTSCAN asymmetry was more pronounced for the slow-progressing subtype at baseline (p=0.0056, Fig. 3D). Over time, differences in DaTSCAN asymmetry between subtypes narrowed, as DaTSCAN asymmetry increased for fast-progressing PwPD (+1.1 %/year) and decreased for slow-progressing PwPD (−1.0 %/year) with high significance between subtypes (p<0.0001).

We analyzed 15 digital gait parameters, of which seven exhibited significant differences between subtypes. Specifically, the fast-progressing subtype expressed lower gait speed (p=0.00027, Fig. 3E), shorter stride length (p=0.00027), a larger toe off angle (p=0.002), lower toe clearance (p=0.006), shorter relative swing time (p=0.013), higher relative stance time (p=0.013) and a lower heel clearance (p=0.027). Correlations of all digital gait parameters are presented in the supplement (Fig. S6). Furthermore, gait speed (p=0.0055), stride length (p=0.0055) and toe off angle (p=0.0055) remained also significant in the cross-cohort validation using the PPMI-trained model (Fig. S7).

### Enriching clinical trials with fast-progressing PwPD

Finally, we assessed the feasibility of using PD subtypes for stratification in clinical trials based on machine learning subtype predictions. PD subtypes could be predicted from baseline data with ROC-AUC up to 68% for PPMI, 58% for ICEBERG and 67% for LuxPARK using nested cross-validation (Fig. 4, Fig. S8). Including data from one additional follow up visit for predictions, ROC-AUCs increased to 79% for PPMI, 79% for ICEBERG and 67% for LuxPARK. Cross-cohort validation resulted in marginally lower ROC-AUCs: 56% for ICEBERG and 61% for LuxPARK using baseline data, respectively 71% for ICEBERG and 70% for LuxPARK when including follow-up data. Altogether this demonstrates the generalization ability of subtype prediction using machine learning.

**Figure 4:**
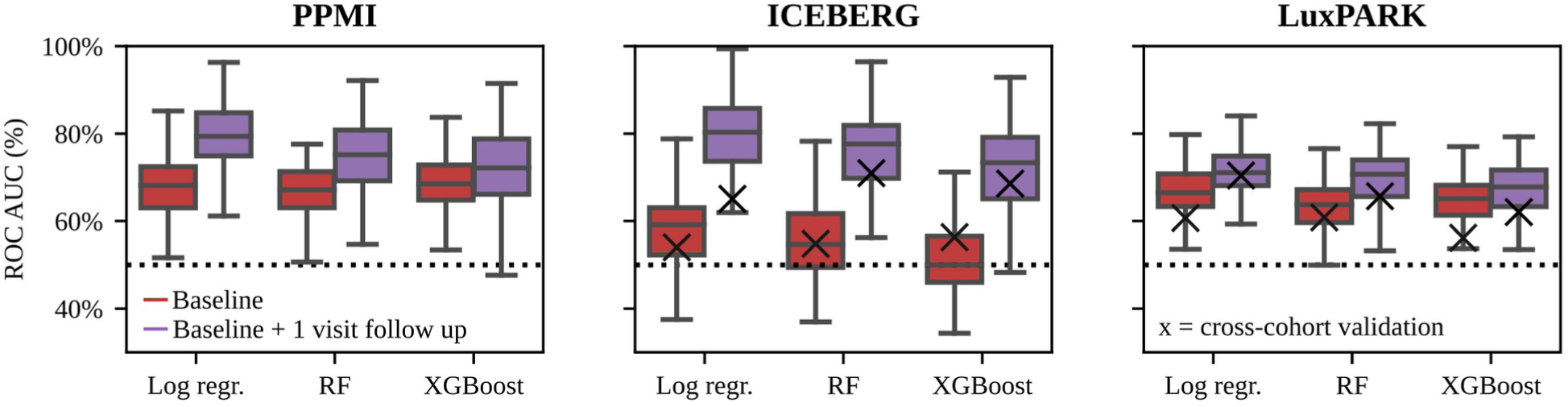
Evaluation of subtype predictions using different machine learning models. Subtypes of individual PwPD were predicted from baseline data (red) or baseline data with one follow up visit (purple) using three different predictive models (Logistic Regression, Random Forest, XGBoost). Models were trained using repeated nested cross-fold validation. ROC-AUC of the subtype predictions is shown for PPMI, ICEBERG and LuxPARK. Additionally, cross-cohort validation was performed using the PPMI-trained model for ICEBERG and LuxPARK predictions (black cross for ICEBERG and LuxPARK figures). Abbreviations: Log Regr: Logistic Regression, RF: Random Forrest, ROC-AUC: receiver operating characteristics-area under the curve, XGBoost: eXtreme Gradient Boosting.

When enrolling PwPD with a high predicted probability for the fast-progressing subtype in a clinical study, there is a trade-off between desired percentage of fast-progressing PwPD in the study and the number of eligible PwPD. If a high percentage of fast-progressing PwPD is desired, fewer PwPD will be eligible for the study (Fig. S9). Without enrichment, 18% of PwPD in PPMI belong to the fast-progressing subtype. Using our predictive models, a percentage of 47% fast-progressing PwPD in a study cohort can be achieved by still allowing inclusion of 30% of all PwPD of the PPMI cohort (Fig. 5A).

**Figure 5:**
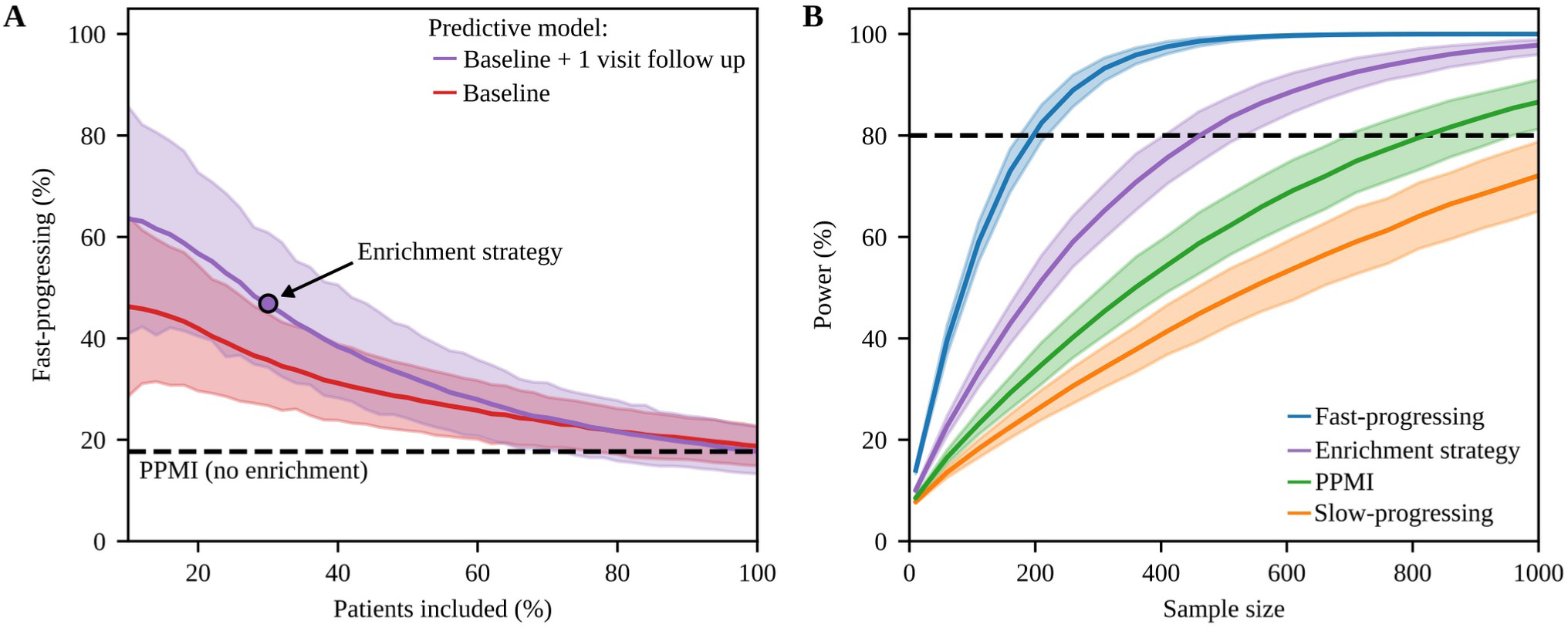
Subtype enrichment for sample size reduction in clinical trials. **A:** Probabilities for PwPD in PPMI of belonging to the fast-progressing subtype predicted from baseline (red) and baseline with one follow up visit (purple) were calculated using the logistic regression model. The figure depicts the percentage of fast-progressing PwPD and the number of PwPD which would be still eligible for study inclusion depending on the threshold applied to the predicted probabilities. The black dashed line indicates the percentage of fast-progressing PwPD observed in the complete PPMI cohort. When using the predictions from baseline + 1 visit follow up data, 47% enrichment can be achieved with still 30% of PwPD being eligible for study inclusion (purple circle). **B**: Estimated power and sample sizes required for a clinical trial depending on percentage of fast-progressing PwPD, assuming the same treatment effect on disease progression for both subtypes: theoretical cohort of only fast-progressing PwPD (blue), enrichment strategy presented in A (purple), default PPMI cohort without enrichment (green), theoretical cohort of only slow-progressing PwPD (orange). A treatment effect of 30% on progression rate of UPDRS I-III, one year observation period and significance level α=0.1 were assumed. The dashed black line indicates 80% power. 95% confidence intervals are shown for both figures.

Subsequently, we simulated a RCT inspired by a currently ongoing trial^20^, demonstrating that such an enrichment strategy, can reduce the required sample size by 43% without losing statistical power. A – theoretical – cohort of only fast-progressing PwPD would reduce the sample size by even 76% (Fig. 5B). Thereby, using the UPDRS I-III sum as primary study outcome resulted in lowest required sample size along all analyzed outcomes with our enrichment strategy reducing required sample sizes for many clinical scores (Fig. S10).

## Discussion

### Progression subtype identification on a common disease timescale

In this study, we identified subtypes of PD *progression* and demonstrated generalizability in multiple external cohorts by using a combination of LTJMM and VaDER. This approach offers several advantages over traditional methods. By aligning PwPD on a common disease timescale, we accommodated for heterogeneity and uncertainty in diagnosis time and prevented an important bias in the data. For example, fast-progressing PwPD or PwPD with tremor as diagnosis-leading symptom are likely to be diagnosed earlier. In this situation, the bias in diagnosis time would be corrected automatically by shifting the PwPD to an earlier disease time on the common disease timescale. Moreover, we include non-motor symptoms for time-aligning to reflect that neurodegeneration does not systematically start in the substantia nigra^21^ and non-motor symptoms precede motor symptoms in PwPD.^7^ By using this approach, we were able to demonstrate the high generalizability of our progression subtypes across heterogeneous cohorts with significant differences, e.g. differences in disease stages, disease severity, age and PD diagnosis criteria. Benefits in disease progression modeling using such a temporal synchronization technique have recently been demonstrated for other neurodegenerative diseases.^16,22^

Compared to other data-driven clustering approaches, our work is the first study that identifies subtypes of PD *progression* for which generalizability in external cohorts could be demonstrated.^11–14^

### Relating PD progression subtypes with the body-first vs brain-first concept

Interestingly, the results from our hypothesis-free data-driven approach are surprisingly consistent with the brain-first vs body-first subtype concept.^8,9^ The fast-progressing subtype exhibits higher portion of RBD symptoms, more severe non-motor symptoms and cognitive impairment at baseline and a more rapid progression of most symptoms, thereby aligning with the body-first subtype. Contrarily, slow-progressing PwPD exhibit more RBD progression, reflecting the fact that brain-first PwPD develop RBD after onset of parkinsonism. Indeed, the fact that more PwPD were classified into the slow-progressing subtype aligns well with the fact that only a minority of PwPD present RBD at PD onset. ^23^ The DaTSCAN uptake ratio was similar in both subtypes at baseline. This can be explained by the fact that PD diagnosis depends on the onset of motor symptoms and thus a specific degree of nigrostriatal degeneration. Similar findings have been observed for the brain-first vs body-first concept.^8^ DaTSCAN was more asymmetric in slow-progressing PwPD – as proposed for the brain-first subtype.^9^ The fact that hyposmia was more abundant in fast-progressing PwPD supports the hypothesis that hyposmia is related to the body-first subtype, which has been the subject of controversy.^9^ The higher portion of gait impairment in fast-progressing PwPD is reflected by differences in a variety of digital gait markers and is consistent with the idea that the brain stem is earlier affected in body-first PwPD^8^, thus confirming our subtype concept by digital biomarkers.

### Imaging and digital gait biomarkers

Our analysis shows significant differences across progression subtypes in features extracted from DaTSCANs and digital gait assessments. This confirms the biological basis of the two progression subtypes and suggests these methods effectively report disease progression. In particular, sensor-based gait assessments are rather inexpensive and could be performed in an at-home setting. Hence, our results contribute to the growing body of literature suggesting the idea to systematically monitor motor symptoms via such technologies, opening up the possibility for a better individualized treatment of PD in the future.^24^ However, opposed to most authors we base this conclusion not on a discrimination of PD versus healthy controls, but on a differentiation between PD progression subtypes.

### Subtype stratification for clinical trials

Enriching fast-progressing PwPD in a cohort reduces the variance of progression rates. In addition, neuroprotective effects are potentially higher in fast-progressing PwPD. Both factors enhance the statistical power of clinical trials. Yet, the presumed biological difference between PD subtypes suggests that they may require different treatments. Our enrichment strategy reduces the required sample size in RCTs by approximately 43%. This is in a similar range as demonstrated for Huntington’s disease using a comparable stratification approach.^22^ Among the outcomes we investigated, the MDS-UPDRS I-III sum score achieved the highest statistical power compared to other potential primary outcomes, in line with the design of ongoing PD trials.^20^

### Limitations and open research questions

Using LTJMM, we assume an approximately linear outcome progression. Other researchers have shown that at least some markers of PD progression demonstrate a non-linear progression profile.^25^ Therefore, using an exponential or sigmoid function could be indeed more realistic for some markers, but would at the same time result in a significantly more complex model requiring also more visits per PwPD to accurately estimate model parameters. Despite this simplification, LTJMM has been applied successfully for disease progression modeling in other neurodegenerative disorders.^16^

Another limitation arises from the heterogeneous set of outcomes measured in PPMI, ICEBERG and LuxPARK. Our choice of outcomes for model training was a trade-off between assessing relevant symptoms and having outcomes measured across all three cohorts. Other outcomes choices may be advantageous but would have hampered cross-cohort validation and thus, the question of generalizability.

Unanswered questions involve how biomarkers like alpha-synuclein pathology assessed by real-time quaking-induced conversion (RT-QuiC) and other digital biomarkers relate to the subtypes. Recently, it has been discussed if different alpha-synuclein strain types may depict the biological basis of the brain-first and body-first subtype.^26^ There is also a need to explore how different genetic mutations are related to these subtypes. For example, GBA mutation carriers are suggested to have a shorter PD prodromal phase and present more often RBD, thus relating potentially to the fast-progressing and body-first subtype.^27–29^ On the other hand, PwPD with LRRK2 G2019S mutation show less RBD symptoms and hyposmia^27,30^, which may be related to the brain-first and slow-progressing subtype.

Ideally, new clinical trials should assess these biological and digital markers along with a comparable set of clinical markers including the outcomes used in this publication for subtype identification. This will allow researchers to relate the biomarkers to the slow-progressing and fast-progressing subtype, thereby leading to an even more precise description and prediction of PD subtypes.

## Conclusion

We provide compelling evidence for the existence of a fast- and slow-progressing subtype in PD as our conclusions are derived from prospective, longitudinal cohorts including more than 1,100 PwPD and were replicated in three distinct PD cohorts. Our findings are in accordance with the body-first vs brain-first and the SOC model, which provide a biological explanation for the subtypes. Our results enhance the understanding of PD progression heterogeneity and highlight the potential of digital gait assessments to objectively monitor motor symptom progression. Finally, we offer a promising strategy to optimize clinical trial designs or investigate new therapeutic strategies in PD subtypes.

## Data availability

The source code used for training LTJMM, VaDER and all relevant statistical analyses will be published at https://github.com/t-haehnel/PD-Progression-Subtypes upon acceptance of the paper under the MIT license, thereby providing free access for anyone. As this study is a retrospective analysis, availability of the clinical data depends on the individual study groups (PPMI: www.ppmi-info.org, ICEBERG, LuxPARK).

## Supplementary material

Supplementary material including details about methods, datasets and additional figures is available online.

## Appendix

### ICEBERG study group

#### Steering committee

Marie Vidailhet, MD, PhD, (Pitié-Salpêtrière Hospital, Paris, principal investigator of ICEBERG), Jean-Christophe Corvol, MD, PhD (Pitié-Salpêtrière Hospital, Paris, scientific lead), Isabelle Arnulf, MD, PhD (Pitié-Salpêtrière Hospital, Paris, member of the steering committee), Stéphane Lehericy, MD, PhD (Pitié-Salpêtrière Hospital, Paris, member of the steering committee);

#### Clinical data

Marie Vidailhet, MD, PhD, (Pitié-Salpêtrière Hospital, Paris, coordination), Graziella Mangone, MD, PhD (Pitié-Salpêtrière Hospital, Paris, co-coordination), Jean-Christophe Corvol, MD, PhD (Pitié-Salpêtrière Hospital, Paris), Isabelle Arnulf, MD, PhD (Pitié-Salpêtrière Hospital, Paris), Sara Sambin, MD (Pitié-Salpêtrière Hospital, Paris), Poornima Menon, MD (Pitié-Salpêtrière Hospital, Paris, Jonas Ihle, MD (Pitié-Salpêtrière Hospital, Paris), Caroline Weill, MD, (Pitié-Salpêtrière Hospital, Paris), David Grabli, MD, PhD (Pitié-Salpêtrière Hospital, Paris); Florence Cormier-Dequaire, MD (Pitié-Salpêtrière Hospital, Paris); Louise Laure Mariani, MD, PhD (Pitié-Salpêtrière Hospital, Paris), Bertrand Degos, MD, PhD (Avicenne Hospital, Bobigny);

#### Neuropsychological data

Richard Levy, MD (Pitié-Salpêtrière Hospital, Paris, coordination), Fanny Pineau, MS (Pitié-Salpêtrière Hospital, Paris, neuropsychologist), Julie Socha, MS (Pitié-Salpêtrière Hospital, Paris, neuropsychologist), Eve Benchetrit, MS (La Timone Hospital, Marseille, neuropsychologist), Virginie Czernecki, MS (Pitié-Salpêtrière Hospital, Paris, neuropsychologist), Marie-Alexandrine, MS (Pitié-Salpêtrière Hospital, Paris, neuropsychologist);

#### Eye movement

Sophie Rivaud-Pechoux, PhD (ICM, Paris, coordination); Elodie Hainque, MD, PhD (Pitié-Salpêtrière Hospital, Paris);

#### Sleep assessment

Isabelle Arnulf, MD, PhD (Pitié-Salpêtrière Hospital, Paris, coordination), Smaranda Leu Semenescu, MD (Pitié-Salpêtrière Hospital, Paris), Pauline Dodet, MD (Pitié-Salpêtrière Hospital, Paris);

#### Genetic data

Jean-Christophe Corvol, MD, PhD (Pitié-Salpêtrière Hospital, Paris, coordination), Graziella Mangone, MD, PhD (Pitié-Salpêtrière Hospital, Paris, co-coordination), Samir Bekadar, MS (Pitié-Salpêtrière Hospital, Paris, biostatistician), Alexis Brice, MD (ICM, Pitié-Salpêtrière Hospital, Paris), Suzanne Lesage, PhD (INSERM, ICM, Paris, genetic analyses);

#### Metabolomics

Fanny Mochel, MD, PhD (Pitié-Salpêtrière Hospital, Paris, coordination), Farid Ichou, PhD (ICAN, Pitié-Salpêtrière Hospital, Paris), Vincent Perlbarg, PhD, Pierre and Marie Curie University), Benoit Colsch, PhD (CEA, Saclay), Arthur Tenenhaus, PhD (Supelec, Gif-sur-Yvette, data integration);

#### Brain MRI data

Stéphane Lehericy, MD, PhD (Pitié-Salpêtrière Hospital, Paris, coordination), Rahul Gaurav, MS, (Pitié-Salpêtrière Hospital, Paris, data analysis), Nadya Pyatigorskaya, MD, PhD, (Pitié-Salpêtrière Hospital, Paris, data analysis); Lydia Yahia-Cherif, PhD (ICM, Paris, Biostatistics), Romain Valabregue, PhD (ICM, Paris, data analysis), Cécile Galléa, PhD (ICM, Paris);

#### Datscan imaging data

Marie-Odile Habert, MCU-PH (Pitié-Salpêtrière Hospital, Paris, coordination);

#### Voice recording

Dijana Petrovska, PhD (Telecom Sud Paris, Evry, coordination), Laetitia Jeancolas, MS (Telecom Sud Paris, Evry);

#### Study management

Alizé Chalançon (Pitié-Salpêtrière Hospital, Paris, Project manager), Carole Dongmo-Kenfack (Pitié-Salpêtrière Hospital, Paris, clinical research assistant); Christelle Laganot (Pitié-Salpêtrière Hospital, Paris, clinical research assistant), Valentine Maheo (Pitié-Salpêtrière Hospital, Paris, clinical research assistant), Manon Gomes (Pitié-Salpêtrière Hospital, Paris, clinical research assistant)

#### Study sponsoring

The ICEBERG Study was funded by the Programme d’investissements d’avenir (ANR-10-IAIHU-06), the Paris Institute of Neurosciences – IHU (IAIHU-06), the Agence Nationale de la Recherche (ANR-11-INBS-0006), and Électricité de France (Fondation d’Entreprise EDF).

### NCER-PD/LuxPARK consortium

We would like to thank all participants of the Luxembourg Parkinson’s Study for their important support to our research. Furthermore, we acknowledge the joint effort of the National Centre of Excellence in Research on Parkinson’s Disease (NCER-PD) Consortium members from the partner institutions Luxembourg Centre for Systems Biomedicine, Luxembourg Institute of Health, Centre Hospitalier de Luxembourg, and Laboratoire National de Santé generally contributing to the Luxembourg Parkinson’s Study as listed below:

Geeta ACHARYA^2^, Gloria AGUAYO^2^, Myriam ALEXANDRE^2^, Muhammad ALI^1^, Wim AMMERLANN^2^, Giuseppe ARENA^1^, Rudi BALLING^1^, Michele BASSIS^1^, Katy BEAUMONT^2^, Regina BECKER^1^, Camille BELLORA^2^, Guy BERCHEM^3^, Daniela BERG^11^, Alexandre BISDORFF^5^, Ibrahim BOUSSAAD^1^, Kathrin BROCKMANN^11^, Jessica CALMES^2^, Lorieza CASTILLO^2^, Gessica CONTESOTTO^2^, Nico DIEDERICH^3^, Rene DONDELINGER^5^, Daniela ESTEVES^2^, Guy FAGHERAZZI^2^, Jean-Yves FERRAND^2^, Manon GANTENBEIN^2^, Thomas GASSER^11^, Piotr GAWRON^1^, Soumyabrata GHOSH^1^, Marijus GIRAITIS^2,3^, Enrico GLAAB^1^, Elisa GÓMEZ DE LOPE^1^, Jérôme GRAAS^2^, Mariella GRAZIANO^17^, Valentin GROUES^1^, Anne GRÜNEWALD^1^, Wei GU^1^, Gaël HAMMOT^2^, Anne-Marie HANFF^2,20,21^, Linda HANSEN^1,3^, Michael HENEKA^1^, Estelle HENRY^2^, Sylvia HERBRINK^6^, Sascha HERZINGER^1^, Michael HEYMANN^2^, Michele HU^8^, Alexander HUNDT^2^, Nadine JACOBY^18^, Jacek JAROSLAW LEBIODA^1^, Yohan JAROSZ^1^, Sonja JÓNSDÓTTIR^2^, Quentin KLOPFENSTEIN^1^, Jochen KLUCKEN^1,2,3^, Rejko KRÜGER^1,2,3^, Pauline LAMBERT^2^, Zied LANDOULSI^1^, Roseline LENTZ^7^, Inga LIEPELT^11^, Robert LISZKA^14^, Laura LONGHINO^3^, Victoria LORENTZ^2^, Paula Cristina LUPU^2^, Tainá M. MARQUES^1^, Clare MACKAY^10^, Walter MAETZLER^15^, Katrin MARCUS^13^, Guilherme MARQUES^2^, Patricia MARTINS CONDE^1^, Patrick MAY^1^, Deborah MCINTYRE^2^, Chouaib MEDIOUNI^2^, Francoise MEISCH^1^, Myriam MENSTER^2^, Maura MINELLI^2^, Michel MITTELBRONN^1,4^, Brit MOLLENHAUER^12^, Friedrich MÜHLSCHLEGEL^4^, Romain NATI^3^, Ulf NEHRBASS^2^, Sarah NICKELS^1^, Beatrice NICOLAI^3^, Jean-Paul NICOLAY^19^, Fozia NOOR^2^, Marek OSTASZEWSKI^1^, Clarissa P. C. GOMES^1^, Sinthuja PACHCHEK^1^, Claire PAULY^1,3^, Laure PAULY^2,20^, Lukas PAVELKA^1,3^, Magali PERQUIN^2^, Nancy E. RAMIA^1^, Rosalina RAMOS LIMA^2^, Armin RAUSCHENBERGER^1^, Rajesh RAWAL^1^, Dheeraj REDDY BOBBILI^1^, Kirsten ROOMP^1^, Eduardo ROSALES^2^, Isabel ROSETY^1^, Estelle SANDT^2^, Stefano SAPIENZA^1^, Venkata SATAGOPAM^1^, Margaux SCHMITT^2^, Sabine SCHMITZ^1^, Reinhard SCHNEIDER^1^, Jens SCHWAMBORN^1^, Amir SHARIFY^2^, Ekaterina SOBOLEVA^1^, Kate SOKOLOWSKA^2^, Hermann THIEN^2^, Elodie THIRY^3^, Rebecca TING JIIN LOO^1^, Christophe TREFOIS^1^, Johanna TROUET^2^, Olena TSURKALENKO^2^, Michel VAILLANT^2^, Mesele VALENTI^2^, Gilles VAN CUTSEM^1,3^, Carlos VEGA^1^, Liliana VILAS BOAS^3^, Maharshi VYAS^1^, Richard WADE-MARTINS^9^, Paul WILMES^1^, Evi WOLLSCHEID-LENGELING^1^, Gelani ZELIMKHANOV^3^

1. Luxembourg Centre for Systems Biomedicine, University of Luxembourg, Esch-sur-Alzette, Luxembourg
2. Luxembourg Institute of Health, Strassen, Luxembourg
3. Centre Hospitalier de Luxembourg, Strassen, Luxembourg
4. Laboratoire National de Santé, Dudelange, Luxembourg
5. Centre Hospitalier Emile Mayrisch, Esch-sur-Alzette, Luxembourg
6. Centre Hospitalier du Nord, Ettelbrück, Luxembourg
7. Parkinson Luxembourg Association, Leudelange, Luxembourg
8. Oxford Parkinson’s Disease Centre, Nuffield Department of Clinical Neurosciences, University of Oxford, Oxford, UK
9. Oxford Parkinson’s Disease Centre, Department of Physiology, Anatomy and Genetics, University of Oxford, South Parks Road, Oxford, UK
10. Oxford Centre for Human Brain Activity, Wellcome Centre for Integrative Neuroimaging, Department of Psychiatry, University of Oxford, Oxford, UK
11. Center of Neurology and Hertie Institute for Clinical Brain Research, Department of Neurodegenerative Diseases, University Hospital Tübingen, Germany
12. Paracelsus-Elena-Klinik, Kassel, Germany
13. Ruhr-University of Bochum, Bochum, Germany
14. Westpfalz-Klinikum GmbH, Kaiserslautern, Germany
15. Department of Neurology, University Medical Center Schleswig-Holstein, Kiel, Germany
16. Department of Neurology Philipps, University Marburg, Marburg, Germany
17. Association of Physiotherapists in Parkinson’s Disease Europe, Esch-sur-Alzette, Luxembourg
18. Private practice, Ettelbruck, Luxembourg
19. Private practice, Luxembourg, Luxembourg
20. Faculty of Science, Technology and Medicine, University of Luxembourg, Esch-sur-Alzette, Luxembourg
21. Department of Epidemiology, CAPHRI School for Public Health and Primary Care, Maastricht University Medical Centre+, Maastricht, the Netherlands

