## Supplementary Material for "Progression Subtypes in Parkinson’s Disease: A Data-driven Multi-Cohort Analysis"

### Clinical cohorts

#### PPMI

We analyzed 409 people with Parkinson's Disease (PwPD) from the publicly available Parkinson's Progression Markers Initiative (PPMI) with clinical visits between 2011 and 2020. All PwPD had a clinical diagnosis of Parkinson's Disease (PD) and a pathological dopamine transporter SPECT (DaTSCAN). We restricted our analysis to untreated de-novo PwPD. Therefore, we included only PwPD with clinical diagnosis not more than two years before baseline visit, Hoehn & Yahr stage 0-2 and no dopaminergic treatment at baseline visit. We further restricted our analysis to PwPD with age > 30 years and at least one additional visit as we require longitudinal information. In addition to clinical scores, DaTSCANs were performed at screening visit and up to three additional visits. Informed consent to data collection and sharing was obtained from all PwPD by PPMI. Ethical guidelines on human data collection were adhered to.

#### ICEBERG

We analyzed 154 PwPD from the ICEBERG cohort study (NCT02305147), an ongoing four-year observational study of PwPD with recent onset of PD conducted at the Paris Brain Institute (Institut du Cerveau-ICM, Pitié-Salpêtrière Hospital, Paris, France). Visits were performed between 2014 and 2022. PD was diagnosed according to UK Parkinson's Disease Society Brain Bank criteria and PwPD with DaTSCANs showing no dopaminergic deficit were excluded. Inclusion was restricted to disease onset not more than three years before baseline visit. We further restricted our analysis to PwPD with at least two visits as we require longitudinal information. Informed consent was obtained and ethical guidelines were adhered to.

#### LuxPARK

We analyzed 561 PwPD from the LuxPARK trial, an ongoing observational study of all disease stages PwPD from Luxembourg and the Greater Region with up to four years follow up. Visits were performed between 2015 and 2022. PD was diagnosed according to UK Parkinson's Disease Society Brain Bank criteria. We restricted our analysis to PwPD with at least two visits as we require longitudinal information. In addition to clinical scores, digital gait measurements were performed at one visit for a subset of 177 patients. Informed consent was obtained and ethical guidelines were adhered to.

### Latent time joint mixed-effects model

We modeled disease progression in each cohort as a linear process using a latent time joint mixed-effects model (LTJMM) as proposed from Li et. al.[1]

$$y_{ijk} = x_i \beta_k + \gamma_k (t_{ijk} + \delta_i) + \alpha_{0ik} + \alpha_{1ik} t_{ijk} + \epsilon_{ijk}$$

Thereby, we denote $y_{ijk}$ as outcome $k$ observed at measurement $j$ for an individual $i$. We account for age and sex differences by including age at diagnosis and sex as covariates $x_i$ into the model with $\beta_k$ as corresponding coefficient shared across all individuals. The coefficient $\gamma_k$ represents the mean slope of the cohort for each outcome $k$ and is thereby shared across all individuals. We use the time since diagnosis as $t_{ijk}$ and shift all measurements of an individual by a PwPD specific time shift $\delta_i$ shared across all outcomes. Additionally, we include random intercepts $\alpha_{0ik}$ and random slopes $\alpha_{1ik}$ for each individual and outcome. As usual, measurement errors $\varepsilon_{ijk}$ and time shifts $\delta_i$ are both assumed to be drawn from normal distributions with a mean of zero. Random intercepts and slopes follow a multivariate normal distribution with mean of zero. Fitting was performed using a Markov chain Monte Carlo (MCMC) algorithm with 4 chains, 25000 iterations and 12500 warm up steps. Analyses were performed using the R packages ltjmm[2] and rstan.[3]

Unified Parkinson's Disease Rating Scale (UPDRS) I-IV, Postural Instability and Gait Dysfunction score (PIGD), Montreal Cognitive Assessment (MoCA) and Scales for Outcomes in Parkinson's Disease-Autonomic Dysfunction (SCOPA) were used as outcomes and min-max-normalized on the theoretical range of the scores. MoCA scores were inverted to ensure positive slopes for all outcomes.

Convergence of MCMCs and normal distribution of parameter estimates were inspected manually. In addition, $\hat{R}$ statistics were calculated and ensured to be below 1.05.

To visualize and validate the effect of aligning PwPD on a common timescale, we inspected the distributions of Hoehn & Yahr (H&Y) stages which were not used for fitting the LTJMM model. Thereby, we observed a clearer separation of H&Y stages after applying LTJMM to the data (Supplementary Fig. 1).

Further, we inspected the accuracy of our LTJMM approach in predicting outcomes at the next visit. Therefore, we re-trained LTJMM, but excluded the last measurement of all outcomes. Using this LTJMM model, we predicted these last measurements of all outcomes and calculated the coefficient of determination ($R^2$) for these predictions. Thereby, we obtained reasonable $R^2$ values: 55% (PPMI), 53% (ICEBERG), 61% (LuxPARK).

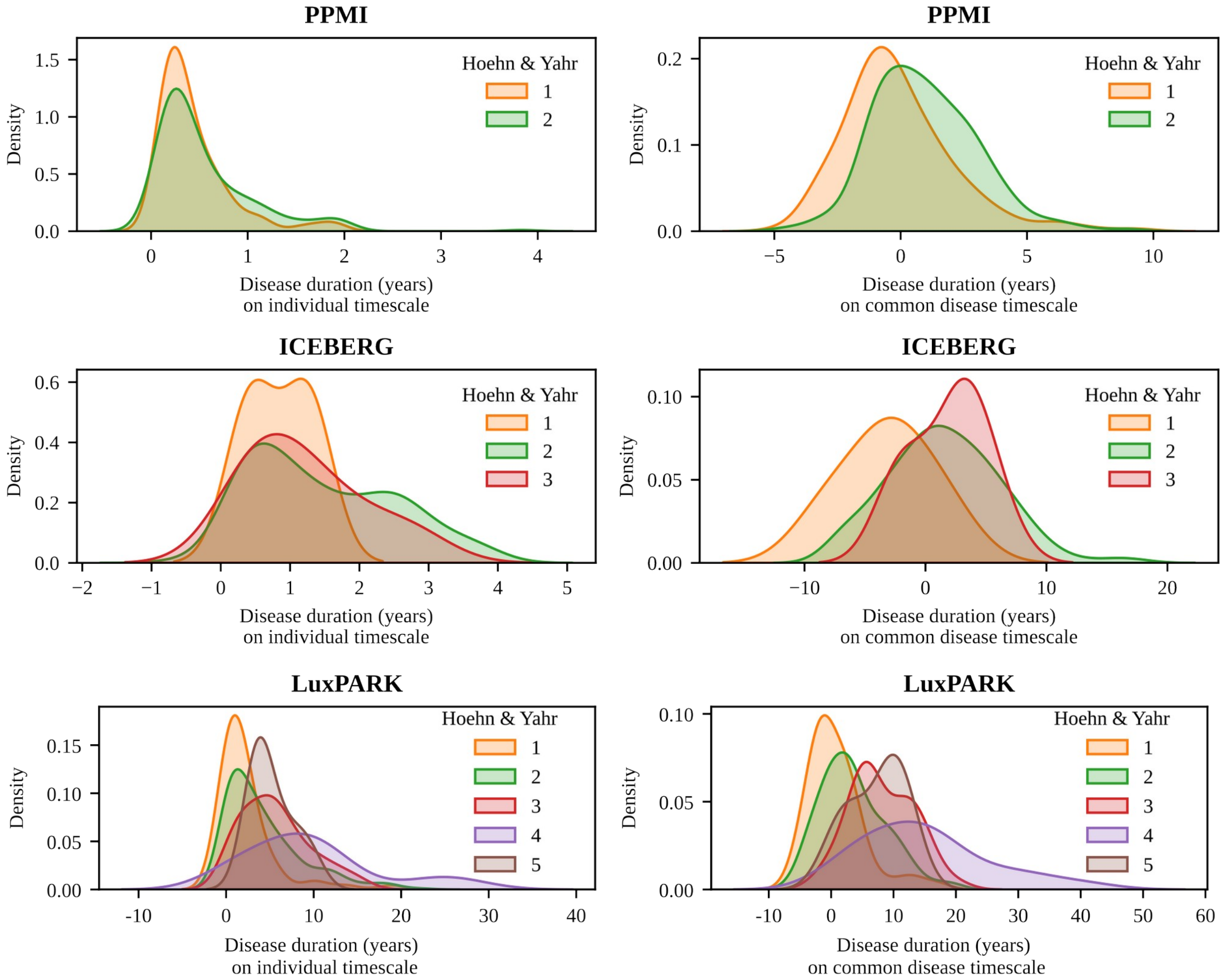

***Supplementary Figure 1: Effect of time-aligning PwPD on distributions of H&Y stages***

*The H&Y distributions from PPMI, ICEBERG and LuxPARK at baseline are depicted. On the left side, H&Y stages are plotted against the original time scale. On the right side, H&Y stages are plotted against the common timescale calculated from the LTJMM.*

*Abbreviations: H&Y: Hoehn&Yahr, LTJMM: latent time joint mixed-effects model*

### Variational deep embedding with recurrence (VaDER)

Parkinson's disease (PD) progression subtypes were identified using variational deep embedding with recurrence (VaDER). Briefly, VaDER implements a recurrent variational autoencoder, in which each data point in the latent space (representing a multivariate patient trajectory) is mapped to a mixture of Gaussians rather than to a single Gaussian. Hence, patient trajectories are clustered. A further distinction point of VaDER is that the input data is passed through an imputation layer. That means that VaDER can directly deal with longitudinal data containing missing values (including those that may occur not at random) and does not require any error prone pre-imputation. A detailed technical description of the VaDER algorithm can be obtained from the original publication.[4]

We used the predicted LTJMM outcomes $\hat{y_{ijk}}$ of UPDRS I-IV, PIGD, MoCA and SCOPA on the common timescale to calculate *outcome progression scores*. The outcomes used are slightly different from our recent work to allow comparability between the different cohorts.[5] Outcome progression scores were calculated for each PwPD by subtracting the outcome at $t$=0 from all outcomes and dividing it by the standard deviation of the outcome at $t$=0. Outcome progression scores were used as input for VaDER.

Hyperparameter optimization was performed using a random search through the following grid: learning rate = {0.0001, 0.001, 0.01, 0.1}, batch size = {16, 32, 64} number of nodes in the hidden layers = {1, 2, 4, 8, 16, 32, 64}. 360 hyperparameter sets were sampled from the grid and evaluated for k={2, 3, 4, 5} number of clusters. 50 epochs were used for VaDER training.

We compared the prediction strength of the fitted VaDER models versus a null model of random cluster assignments as described in our original publication.[4] The increase in prediction strength compared to the null model was already significant for $k$=2 in all cohorts, thus we considered two clusters as appropriate. The final models were trained 20 times and consensus clustering was used to finally assign PwPD to the clusters. The hyperparameters obtained from this approach are presented in Supplementary Table 1 and were used for all following calculations.

| | PPMI | ICEBERG | LuxPARK |
|---|---|---|---|
| number of clusters | 2 | 2 | 2 |
| learning rate | 0.0001 | 0.001 | 0.0001 |
| batch size | 16 | 32 | 16 |
| number of nodes (first hidden layer) | 64 | 32 | 32 |
| number of nodes (second hidden layer) | 1 | 1 | 2 |

***Supplementary Table 1: Results of VaDER hyperparameter optimization for PPMI, ICEBERG and LuxPARK***

### Symptom domain comparisons

To allow a more comprehensive comparison and validation of the variety of outcomes captured across the three cohorts, we grouped 114 outcomes (including single questions, scores and sub-scores from questionnaires and clinical assessments) into 22 symptom domains (Supplementary Table 2). The choice of the 22 symptom domains represents a trade-off between capturing most clinically relevant motor and non-motor symptoms and which outcomes had been assessed in the three cohorts. For each cohort, we included only outcomes where at least 30 PwPD in total and 5 PwPD per subtype were assessed. In addition, at least two measurements per PwPD were required for the progression analysis. For the baseline characteristics analysis, we only evaluated values at baseline and screening visit.

To assess the progression characteristics of both subtypes regarding the defined symptom domains, we applied the following steps: Outcomes were normalized using min-max-normalization. Scales where high values report a low symptom severity were inverted, thus we ensured that high outcome scores always correspond to a high symptom severity. Depending on the scale of each outcome, we modeled the outcome progression using a linear mixed-effects model, a binary mixed-effects model or an ordinal mixed-effects model on the common disease timescale. We used one model per subtype. For each outcome, coefficients depicting outcome progression were extracted for each PwPD and standardized mean differences (SMDs) between subtypes were calculated. Next, we conducted a three-level meta-analysis with random effects for each symptom domain. Thereby, we first calculated for each cohort an overall SMD estimate across all outcomes of one symptom domain. Subsequently, we calculated an overall SMD estimate of the symptom domain across the three cohorts (see forest plots at end of the supplement). P-values and 95% confidence intervals (CI) were corrected for multiple testing across the 22 symptom domains using the Benjamini-Hochberg procedure.[6]

To assess the association of baseline characteristics with the subtypes regarding the defined symptom domains, we applied the following steps: Outcomes were normalized using min-max-normalization. Scales where high values report a low symptom severity were inverted, thus we ensured that high outcome scores always correspond to a high symptom severity. For each outcome, we trained a logistic regression model to predict the subtype based on the baseline outcome value. For each outcome, the logistic regression coefficient was extracted. Next, we conducted a three-level meta-analysis with random effects for each symptom domain. Thereby, we first calculated an overall regression coefficient estimate across all outcomes of one symptom domain per cohort. Subsequently, we calculated an overall regression coefficient estimate of the symptom domain across the three cohorts (see forest plots at end of the supplement). P-values and 95% confidence intervals (CI) were corrected for multiple testing across the 22 symptom domains using the Benjamini-Hochberg procedure[6].

Both analyses were repeated as a cross-cohort validation (see forest plots at end of the supplement).

Analyses were performed using the R-packages lme4[7], ordinal[8] and meta[9].

| Symptom domain | Outcome | Definition/calculation of the outcome |
|---|---|---|
| Anxiety | NMSQ Anxiety | NMSQ item 17 |
| | HADS anxiety | HADS anxiety sub-score |
| | STA | STA sum score |
| | PDQ39 Anxiety | PDQ39 item 21 |
| | UPDRS I Anxiety | UPDRS I item 4 |
| Apathy | DAS | DAS sum score |
| | SAS | SAS sum score |
| | UPDRS I Apathy | UPDRS I item 5 |
| Autonomic symptoms | NMSQ Autonomic | NMSQ sum of items 4, 5, 6, 7, 8, 9, 19, 20, 28 |
| | SCOPA-AUT | SCOPA sum score |
| | UPDRS I Autonomic | UPDRS I sum of items 10, 11, 12 |
| Attention | NMSQ Attention | NMSQ item 15 |
| | Letter Number Sequencing | Letter Number Sequencing score |
| | Digit Span | Digit Span Score: sum of forward scores and backward scores |
| | MATTIS Attention | MATTIS attention sub-score |
| | MOCA Attention | MoCA sum of: digits, letters, subtraction points |
| | MMSE Attention | MMSE attention sub-score |
| | PDQ39 Attention | PDQ39 item 31 |
| | SIQCDE Attention | Short IQCODE score item 11 |
| Conceptualization | MATTIS Conceptualization | MATTIS conceptualization sub-score |
| | MOCA Abstraction | MoCA abstraction sub-score |
| | FAB Conceptualization | FAB item 1 |
| Language | Boston Naming Test | Boston Naming Test sum score |
| | MOCA Language + Naming | MoCA sum of items: naming, repeat and verbal fluency task |
| | VFT phonematic F | phonematic VFT F total word count |
| | VFT phonematic S | phonematic VFT S total word count |
| | VFT semantic animal | semantic VFT animal total word count |
| | VFT semantic sum | semantic VFT total word count (sum of tasks colors, fruits, towns, animals) |
| | VFT semantic supermarket | semantic VFT supermarket total word count |
| | MMSE Language | MMSE language sub-score |
| | FAB VFT | FAB lexical fluency item |
| Memory | NMSQ Memory | NMSQ item 12 |
| | CERAD Words DR | CERAD word count immediate recall |
| | CERAD Words IR | CERAD word count delayed recall |
| | CERAD Words Recognition | CERAD recognition (number of correct, Yes + No) |
| | Hopkins Verbal Learning Test DR | Hopkins Verbal Learning Test delayed recall |
| | Hopkins Verbal Learning Test IR | Hopkins Verbal Learning Test immediate recall |
| | MOCA Orientation + Memory | MoCA sum of: memory (uncued only), orientation |
| | MATTIS Memory | MATTIS memory sub-score |
| | MMSE Memory | MMSE sum of: orientation (location + time), words memorization |
| | SIQCDE Memory | Short IQCODE score sum of items 1, 2, 3, 4, 5, 6, 7 |
| | PDQ39 Memory | PDQ39 item 32 |
| Overall Cognition | MATTIS | MATTIS sum score |
| | MMSE | MMSE sum score |
| | MoCA | MoCA sum score |
| | SIQCDE | Short IQCODE score sum score |
| | FAB | FAB sum score |

| Symptom domain | Outcome | Definition/calculation of the outcome |
|---|---|---|
| | PDQ39 Cognition | PDQ39 sum of items 31, 32 |
| | UPDRS I Cognition | UPDRS I item 1 |
| | NMSQ Cognition | NMSQ sum of items 12, 15 |
| Visu-executive function | MMSE Construction | MMSE item 30 |
| | Judgment Line Orientation | Judgment of Line Orientation sum score |
| | Symbol Digit Modalities | Symbol Digital Modalities sum score |
| | Stroop Errors | Stroop test number of errors |
| | Stroop Time | Stroop test required time |
| | Trailmaking A | Trail Making Test A time |
| | Trailmaking B | Trail Making Test B time |
| | MATTIS Initiation + Construction | MATTIS sum of: sub-score initiation, sub-score construction |
| | FAB 3-6 | FAB sum of items 3, 4, 5, 6 |
| | MOCA Visuspatial/Executive | MoCA visuospatial/executive sub-score |
| Depression | BDI | BDI sum score |
| | GDS | GDS sum score |
| | HADS depression | HADS depression sub-score |
| | PDQ39 Depression | PDQ39 sum of items 17, 18, 19, 20, 22 |
| | NMSQ Depression | NMSQ sum of items 13, 16 |
| | UPDRS I Depression | UPDRS 1 item 3 |
| Fatigue | UPDRS I Fatigue | UPDRS 1 item 13 |
| Hallucinations | NMSQ Hallucination | NMSQ sum of items 14, 30 |
| | UPDRS I Hallucinations | UPDRS 1 item 2 |
| Impulsivity | QUIP | QUIP sum score |
| | QUIP-RS | QUIPRS sum score |
| Motor symptoms (overall) | PDQ39 ADL | PDQ39 ADL sub-score |
| | Pegboard | PEGBoard sum of: average of left hand, right hand and both hands |
| | UPDRS II | UPDRS II sum score |
| | UPDRS III off | UPDRS III sum score (OFF only) |
| | UPDRS III on | UPDRS III sum score (ON only) |
| | UPDRS IV | UPDRS IV sum score |
| Non motor symptoms (overall) | NMSQ | NMSQ sum score |
| | UPDRS I | UPDRS I sum score |
| Overall disease severity | UPDRS I-III on | UPDRS I, II, III sum (ON only) |
| | UPDRS I-III off | UPDRS I, II, III sum (OFF only) |
| | FAQ | FAQ sum score |
| | PDQ39 | PDQ39 sum score |
| | SEADL | SEADL score |
| | H&Y | Hoehn & Yahr |
| | CGIS | CGI-S score |
| Pain | NMSQ Pain | NMSQ item 10 |
| | PDQ39 Pain | PDQ39 sum of items 37, 38 |
| | UPDRS I Pain | UPDRS 1 item 9 |
| Axial & PIGD symptoms | UPDRS III axial off | UPDRS III axial score (OFF only) |
| | UPDRS III axial on | UPDRS III axial score (ON only) |
| | FOGAC | FOGAC sum score |
| | FOGQ | FOGQ sum score |

| Symptom domain | Outcome | Definition/calculation of the outcome |
|---|---|---|
| | GABS Examination | GABS sum of items 8-24 |
| | GABS Questionnaire | GABS sum of items 1-7 |
| | NFOGQ | NFOGQ sum score |
| | PDQ39 Mobility | PDQ39 mobility sub-score |
| | PIGD off | PIGD score (OFF only) |
| | PIGD on | PIGD score (ON only) |
| | TUG | Timed Up and Go time |
| Sleep (general) | ESS | ESS sum score |
| | PDSS | PDSS sum score |
| | UPDRS I Sleep | UPDRS I sum of items 7, 8 |
| | NMSQ Sleep | NMSQ sub of items 22, 23 |
| RBD Sleep | RBD-HK | RBD-HK sum score |
| | RBD-SQ | RBD-SQ sum score |
| | NMSQ RBD | NMSQ sum of items 24, 25 |
| Smell | NMSQ Smell | NMSQ item 2 |
| | Sniffin Test | Sniffin Test score |
| | UPSIT | UPSIT sum score |
| Tremor | TD off | TD score (OFF only) |
| | TD on | TD score (ON only) |

***Supplementary Table 2: Construction of symptom domains***

*Abbreviations: BDI: Beck Depression Inventory, CERAD: Consortium to Establish a Registry for Alzheimer's Disease, CGIS: Clinical Global Impression-Severity, DAS: Dimensional Apathy Scale, ESS: Epworth Sleepiness Scale, FAB: Frontal Assessment Battery, FAQ: Functional Activities Questionnaire, FOGAC: Freezing of Gait AC, FOGQ: Freezing of Gait Questionnaire, GABS: Clinical Gait and Balance Scale, GDS: Geriatric Depression Scale, H&Y: Hoehn & Yahr scale, HADS: Hospital Anxiety and Depression Scale, MATTIS: Mattis Dementia Rating Scale, MMSE: Mini Mental Status Examination, MOCA: Montreal Cognitive Assessment, NFOGQ: New Freezing of Gait Questionnaire, NMSQ: Non-Motor Symptoms Questionnaire, PDQ39: Parkinson's Disease Questionnaire-39, PDSS: Parkinson's Disease Sleep Scale, PIGD: Postural Instability and Gait Disorder score, QUIP: Questionnaire for Impulsive-Compulsive Disorders, QUIP-RS: QUIP-Rating Scale, RBD-HK: REM Sleep Behavior Disorder Questionnaire-Hong Kong, RBD-SQ: REM Sleep Behavior Disorder Screening Questionnaire, SAS: Starkstein Apathy Scale, SCOPA-AUT: Scales for Outcomes in Parkinson's Disease-Autonomic Dysfunction, SEADL: Schwab and England Activities of Daily Living Scale, SIQCDE: Short Informant Questionnaire on Cognitive Decline in the Elderly, STA: State-Trait Anxiety Inventory, TD: Tremor Dominance Score, TUG: Timed Up and Go, UPDRS: MDS-Unified Parkinson's Disease Rating Scale, UPSIT: University of Pennsylvania Smell Identification Test, VFT: Verbal Fluency Task*

### Digital gait measurements

| Gait parameter | Unit | Description |
|---|---|---|
| Gait speed | m/s | The average walking speed. |
| Heel strike angle | degree | The angle between the toes and the surface when the foot lands. |
| Landing impact | g | The maximum vertical acceleration during landing of the foot. |
| Max. foot clearance | cm | The maximum elevation of the foot from the ground during the swing phase. |
| Max heel clearance | cm | The maximum elevation of the heel from the ground during the swing phase. |
| Max. toe clearance | cm | The maximum elevation of the toe from the ground during the swing phase. |
| Max. lateral excursion | cm | The maximum lateral deviation of the foot in the swing phase, measured from an imaginary line between the foot’s position at start and end of the swing phase. |
| Stance time (relative) | % | Duration from initial contact of the foot with the surface until start of next swing phase of the foot. |
| Stance time (absolute) | s | Proportion of stance time divided by the total duration of the stride. |
| Stride length | cm | The length of one stride. |
| Stride time | s | Sum of stance time and swing time. |
| Swing time (relative) | % | Duration from start of swing until next foot contact with the surface. |
| Swing time (absolute) | s | Proportion of swing time divided by the total duration of the stride. |
| Toe off angle | degree | The angle between the heel and the surface at the beginning of the swing phase. |
| Turning angle | degree | The angle between the direction of the last swing phase (imaginary line between foot position at the beginning and end of the swing phase) and the orientation of the foot in the next stance phase. |

***Supplementary Table 3: Description of digital gait biomarkers.***
*Gait parameters were calculated as mean of all straight steps from the Timed Up and Go task. Turning steps were excluded from the calculation.*
*Abbreviations: max: maximum, min: minimum*

### Predictive models

We developed several models to predict PwPD subtypes from (I) baseline and (II) baseline and one additional visit. Therefore, we used penalized Logistic Regression with L2 regularization, Random Forest[10] and eXtreme Gradient Boosting (XGBoost)[11]. These predictive models were implemented using the python packages scikit-learn[12] and xgboost.[13] We used UPDRS I-III, PIGD, MoCA and SCOPA as baseline predictors as they capture a variety of motor and non-motor symptoms and were measured across the three cohorts. UPDRS IV was not included as it was mostly not assessed at baseline. Hyperparameter optimization was performed using grid search (Logistic Regression) or randomized search (Random Forest, XGBoost) with 50 iterations in an inner repeated cross-fold validation using 5 folds with 20 repeats (Supplementary Table 4). Class weights were used to accommodate for unbalanced classes. Estimates for receiver operating characteristics-area under the curve (ROC-AUC) were obtained from an outer repeated cross-fold validation using 5 folds with 20 repeats. Cross-cohort validation was performed by training the predictive model on the complete PPMI dataset using the best parameters from the hyperparameter optimization and predicting the subtype assignments of ICEBERG/LuxPARK. Furthermore, we assessed how much these predictions can be improved if short follow-up data is included into the models. Therefore, we repeated the steps above using outcomes at baseline and at one year follow up.

| **Model** | **Parameter** | **Grid values** | **Baseline PPMI / ICEBERG / LuxPARK** | **Baseline + FU PPMI / ICEBERG / LuxPARK** |
|---|---|---|---|---|
| Logistic regression | Lambda | 0.001, 0.01, 0.1, 1, 10, 100, 1000 | 0.01 / 0.01 / 100 | 0.01 / 10 / 10 |
| Random Forest | Bootstrap | True, False | True / True / True | True / True / True |
| | Maximum depth | 10, 20, 30, 40 | 40 / 20 / 20 | 20 / 10 / 10 |
| | Minimum samples per leaf | 1, 2, 4 | 4 / 1 / 1 | 4 / 2 / 1 |
| | Minimum samples per split | 2, 5, 10 | 10 / 5 / 10 | 10 / 10 / 10 |
| | Number of Estimators | 200, 600, 800, 1000, 1200, 1400, 1600, 1800, 2000 | 800 / 1400 / 1800 | 1800 / 2000 / 200 |
| XGBoost | Minimum Child Weight | 1, 5, 10 | 10 / 1 / 1 | 5 / 1 / 5 |
| | Gamma | 0.5, 1, 1.5, 2, 5 | 2 / 0.5 / 5 | 0.5 / 1.5 / 1.5 |
| | Subsample | 0.6, 0.8, 1.0 | 0.8 / 0.6 / 0.8 | 0.8 / 0.6 / 1.0 |
| | Colsample by tree | 0.6, 0.8, 1.0 | 0.8 / 0.6 / 0.6 | 0.6 / 1.0 / 1.0 |
| | Maximum Depth | 3, 4, 5 | 4 / 4 / 3 | 4 / 3 / 3 |

***Supplementary Table 4: Predictive model hyperparameter optimization***

*Hyperparameter space used for hyperparameter optimization. For Logistic regression, L2 penalization and hyperparameter grid search was used. For Random Forest and XGBoost, randomized hyperparameter search with 50 samples was used. Repeated stratified k-fold cross validation was performed using 5 splits and 20 repeats. Results of hyperparameter training for the baseline-only and baseline with one visit follow-up models are presented.*

*Abbreviations: FU: follow up*

### Sample size estimation

To assess the effect of enriching fast-progressing PwPD in clinical trials on required sample sizes and power, we simulated a randomized controlled trial (RCT) for a potentially disease modifying drug based on considerations of an ongoing clinical trial.[14] We assumed measurements of UPDRS I-III sum score each 60 days for a total of one year observation time. A power of 80% and significance level of 0.1 was chosen. We assumed a treatment effect of 30% reduction in disease progression speed.[14] For simplification, we used equally sized control and treatment groups without different treatment dosage arms. Power and sample size were calculated based on PPMI data using a linear mixed-effects model, thereby assuming a linear UPDRS I-III increase over time. Calculations were based on a method from Edland et al[15] and implemented using the R package longpower.[16] Sample size and power calculations were performed for the complete PPMI cohort and different percentages of fast-progressing PwPD. Enrichment of fast-progressing PwPD was simulated using predictions of the PPMI logistic regression models as this model outperformed Random Forest and XGBoost in predicting PwPD subtypes.

### Additional plots

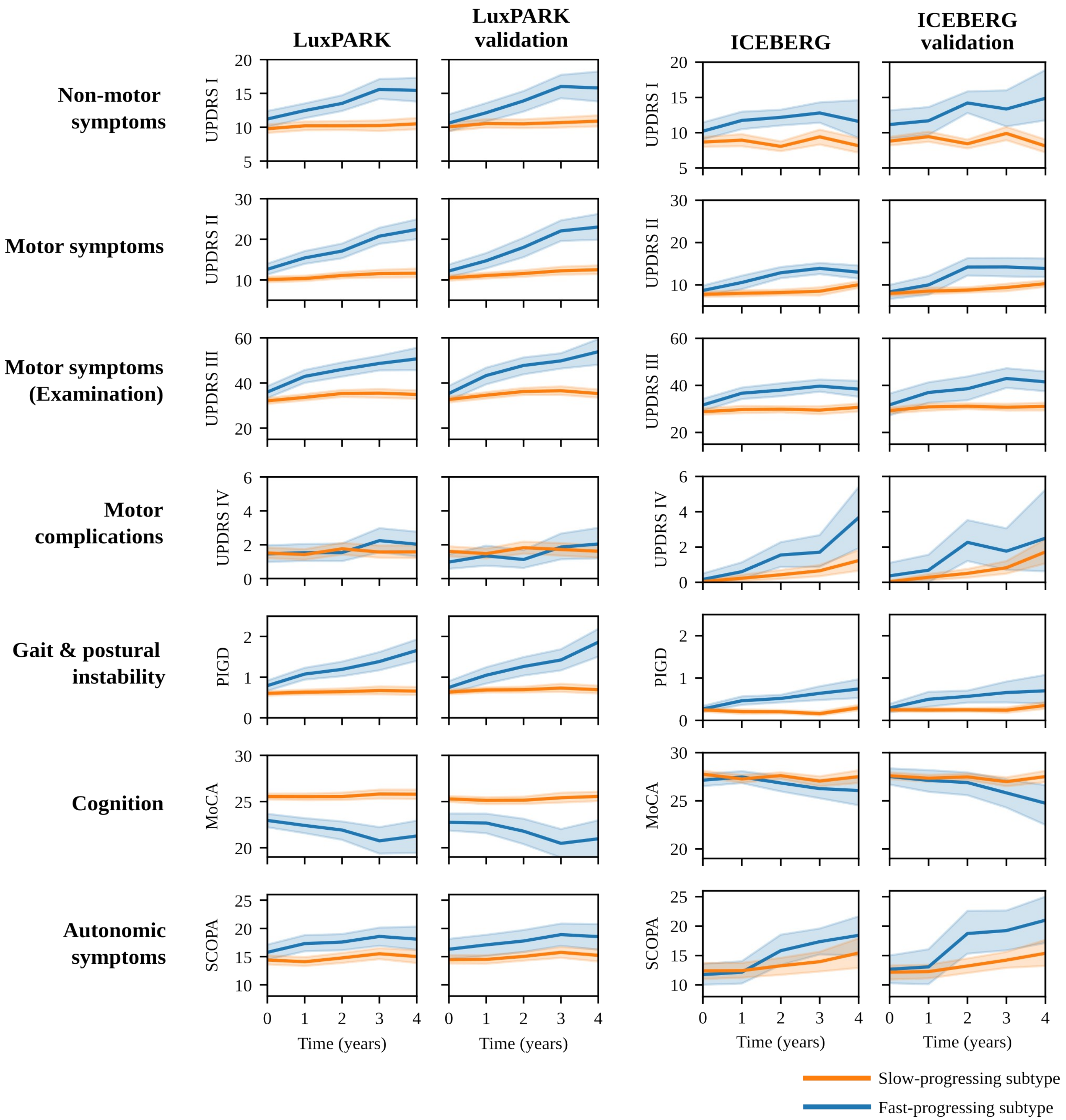

***Supplementary Figure 2: Progression trajectories of subtypes for motor and non-motor symptoms (validation)***

*Progression of motor symptoms (UPDRS II/III/IV, PIGD) and non-motor symptoms (UPDRS I, MoCA, SCOPA) for the slow-progressing subtype (orange) and fast-progressing subtype (blue) for the ICEBERG and LuxPARK cohort. The in-cohort training results and the cross-cohort validation results (models trained on PPMI) are shown side by side. Mean and 95% confidence interval for each subtype are shown.*

*Abbreviations: MoCA: Montreal Cognitive Assessment, PIGD: Postural Instability and Gait Dysfunction score, SCOPA: Scales for Outcomes in Parkinson's Disease-Autonomic Dysfunction, UPDRS: Unified Parkinson's Disease Rating Scale.*

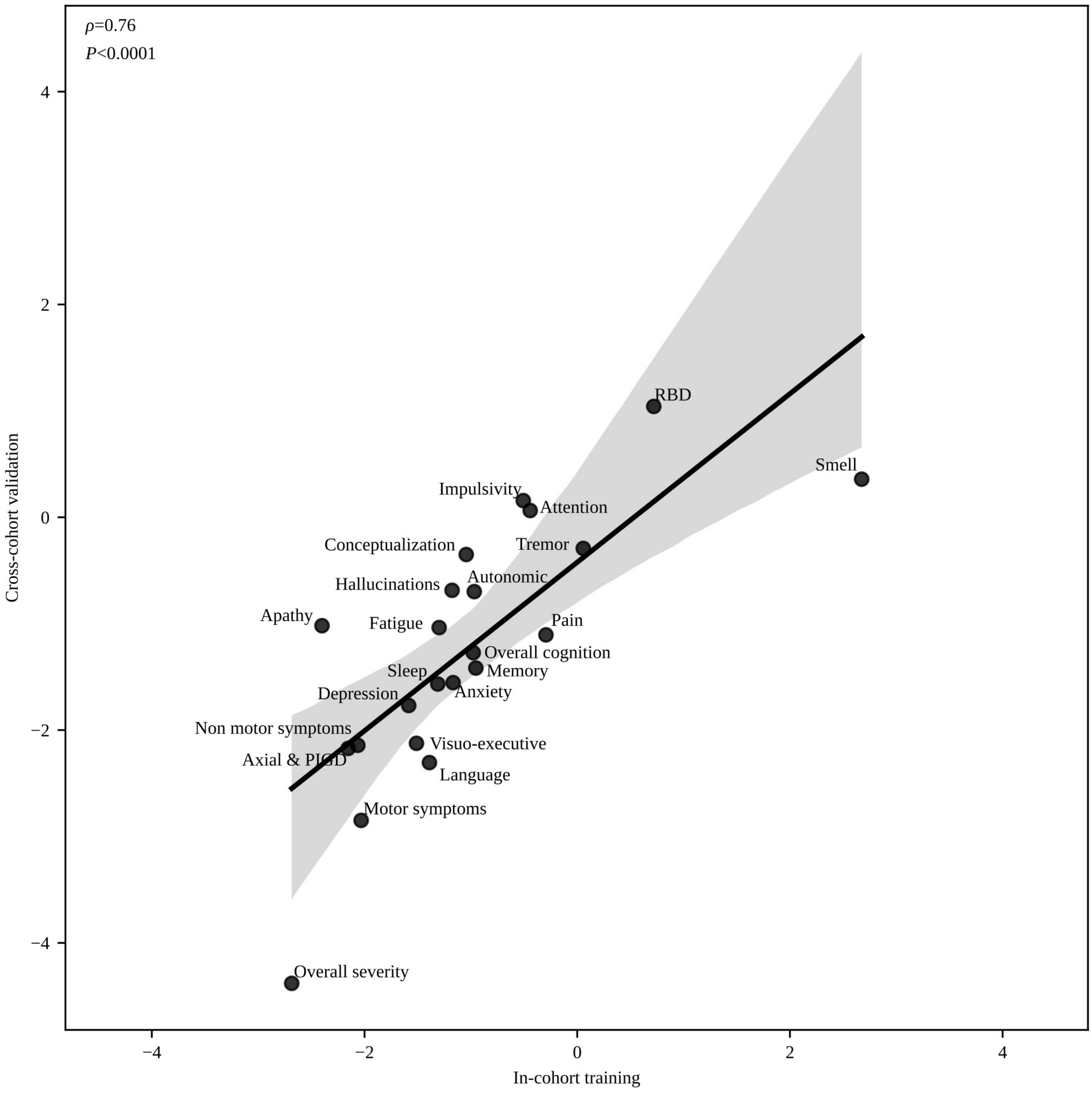

***Supplementary Figure 3: Symptom domain progression rate validation***

*The figure depicts the correlation of standardized mean differences (SMDs) of progression rates calculated for each symptom domain using the in-cohort training approach and the cross-cohort validation approach (i.e. models trained on PPMI). The 95% confidence interval of the regression line is depicted in gray. The Pearson correlation coefficient with corresponding p-value is shown.*

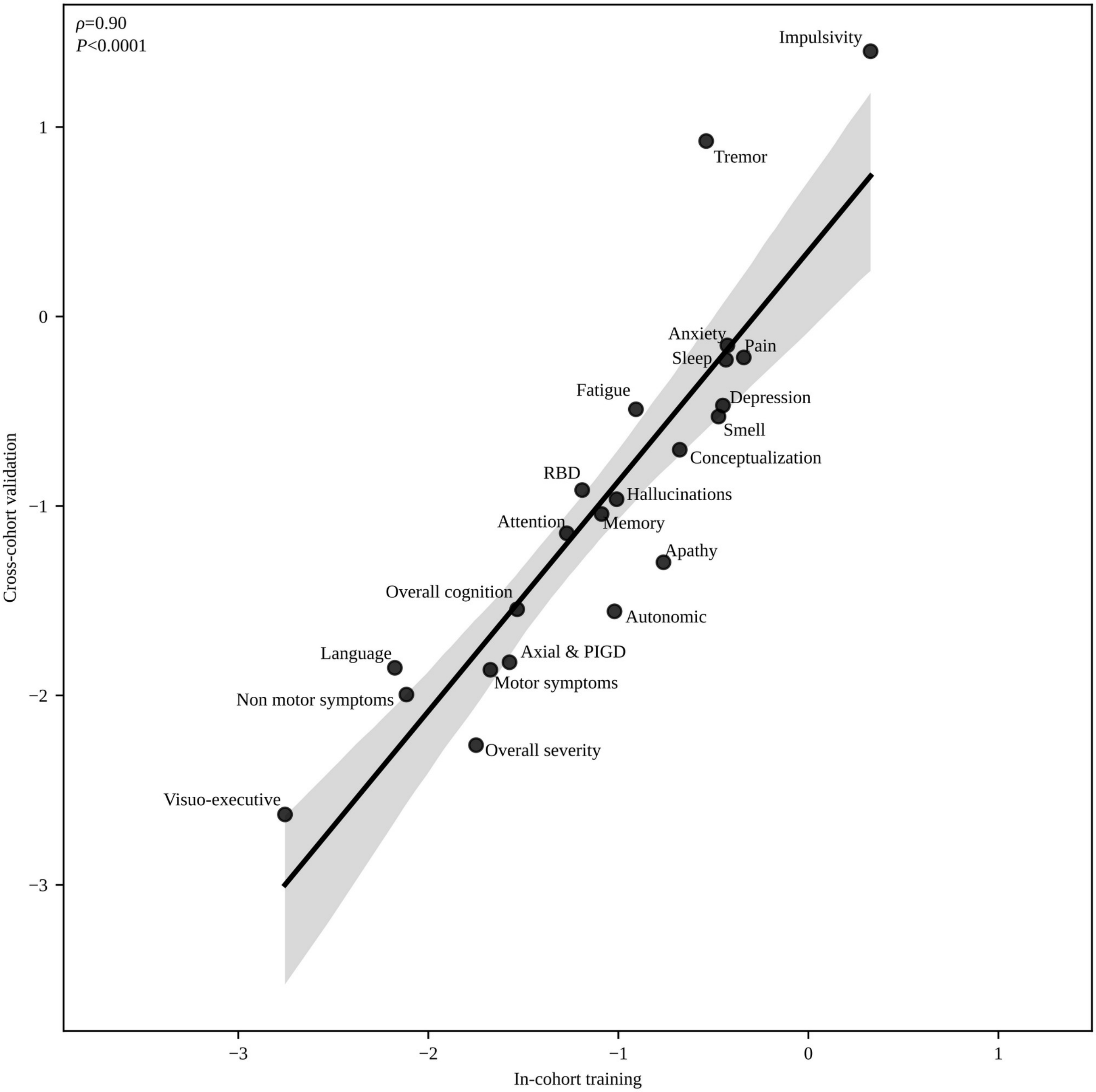

***Supplementary Figure 4: Symptom domain baseline associations validation***
*The figure depicts the correlation of average regression coefficients for baseline outcomes calculated for each symptom domain using the in-cohort training approach and the cross-cohort validation approach (i.e. models trained on PPMI). The 95% confidence interval of the regression line is depicted in gray. The Pearson correlation coefficient with corresponding p-value is shown.*

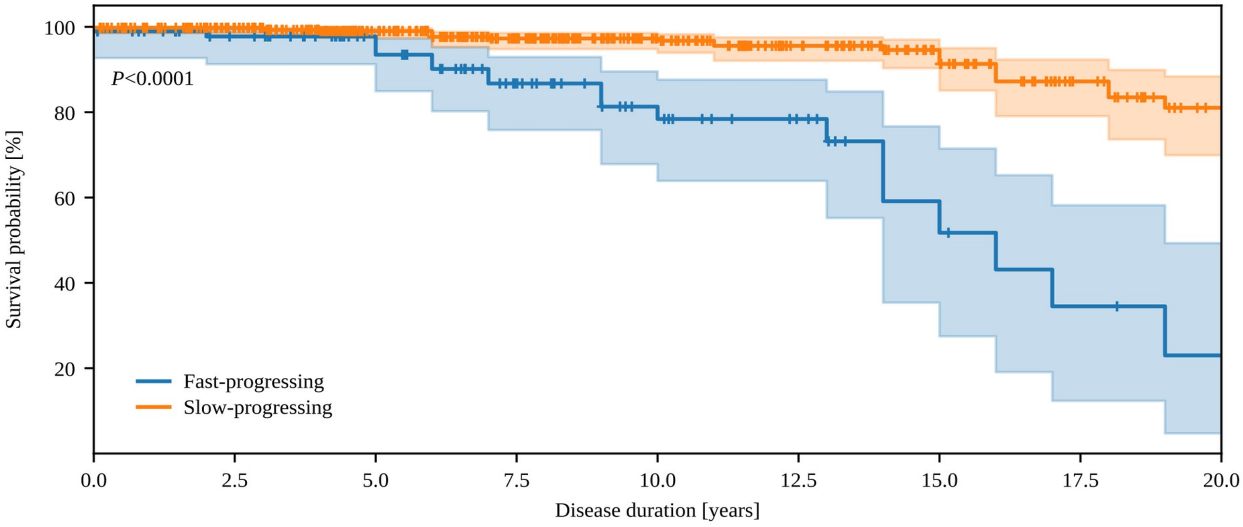

| | | | | | | | | | |
|---|---|---|---|---|---|---|---|---|---|
| At risk | 526 | 466 | 366 | 272 | 188 | 133 | 83 | 48 | 29 |
| Censored | 33 | 90 | 186 | 271 | 351 | 404 | 446 | 475 | 491 |
| Events | 2 | 5 | 9 | 18 | 22 | 24 | 32 | 38 | 41 |

***Supplementary Figure 5: Survival curves (validation)***
*Kaplan-Meier estimator for survival probability on the common disease timescale for fast-progressing (blue) and slow-progressing (orange) PwPD in LuxPARK. Right-censored observations are indicated by a small vertical tick. The corresponding p-value for the subtype covariate from the cox proportional hazard model is reported. The analysis was done using the PPMI-trained model as cross-cohort validation.*

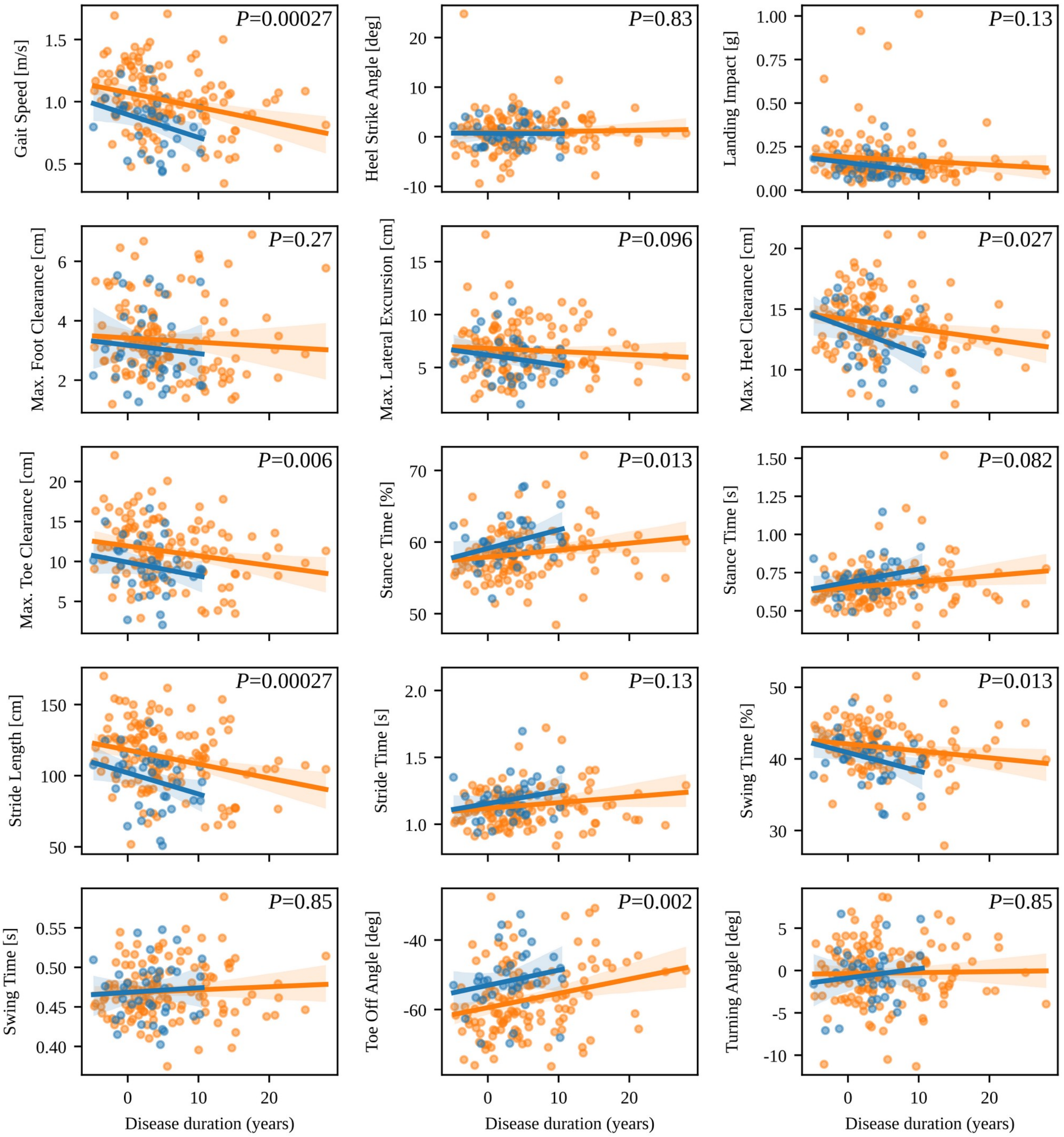

***Supplementary Figure 6: Digital gait biomarkers***

*Correlation of all digital gait markers with disease duration on the common disease timescale for fast-progressing (blue) and slow-progressing (orange) PwPD. The corresponding p-values from the ANCOVA analyses are shown and were corrected for multiple testing.*

*Abbreviations: deg: degree.*

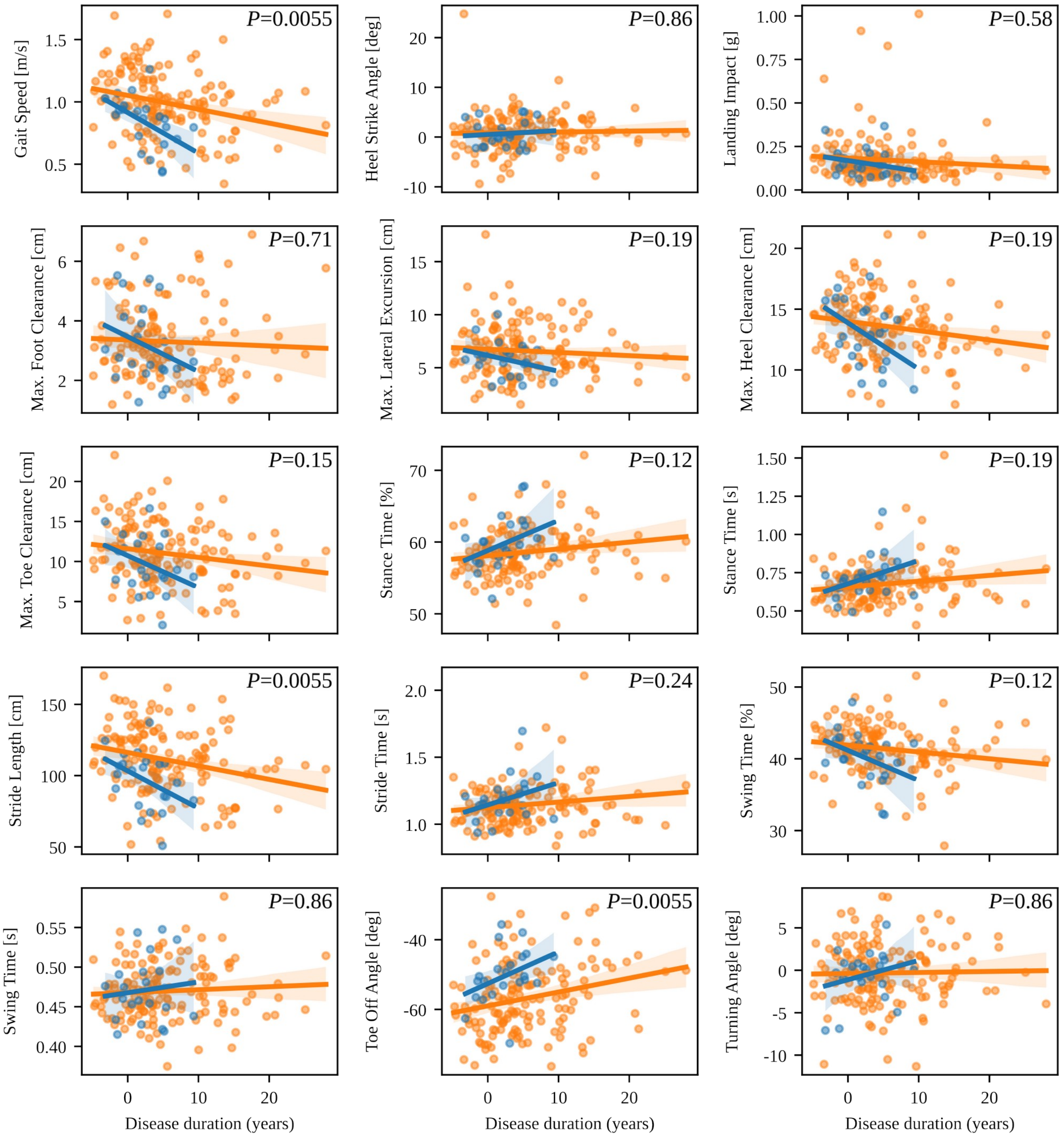

***Supplementary Figure 7: Digital gait biomarkers (validation)***

*Correlation of all digital gait markers with disease duration on the common disease timescale for fast-progressing (blue) and slow-progressing (orange) PwPD. The corresponding p-values from the ANCOVA analyses are shown and were corrected for multiple testing. The analysis was done using the PPMI-trained model as cross-cohort validation.*
*Abbreviations: deg: degree.*

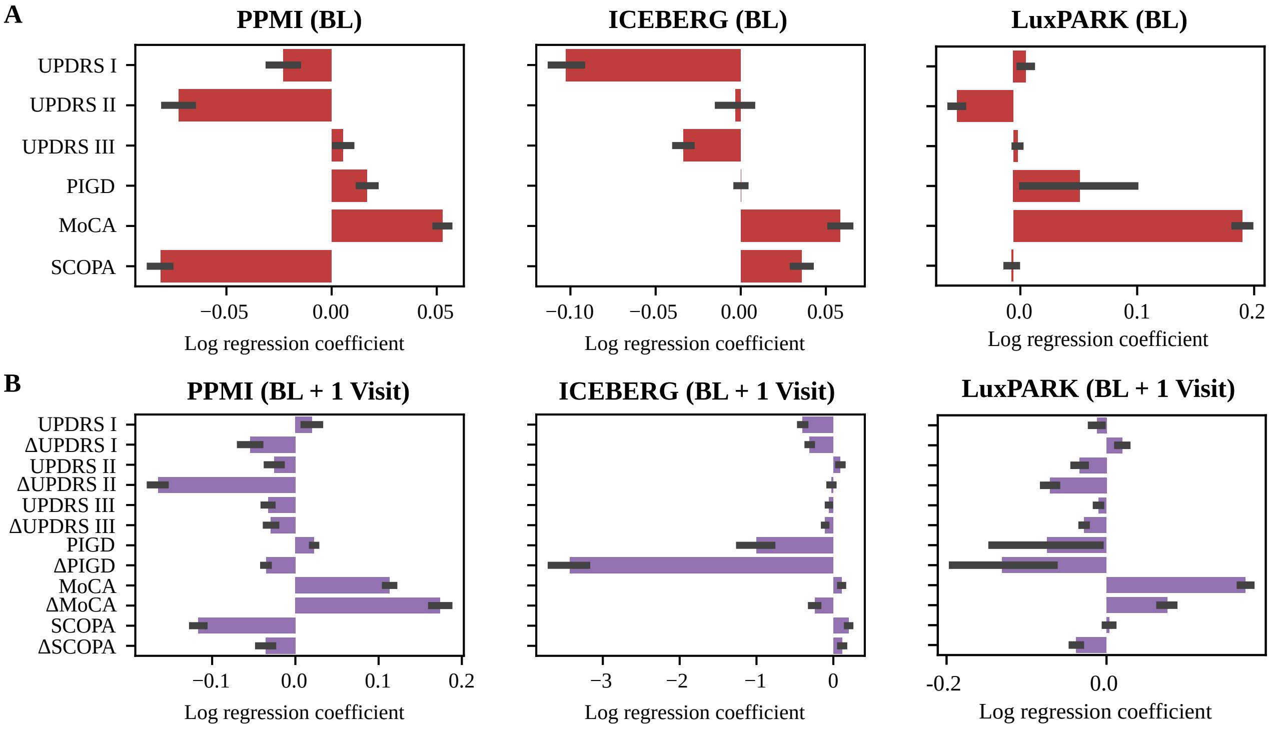

***Supplementary Figure 8: Coefficients of logistic regression models for subtype predictions***
*Logistic regression coefficients with 95% confidence interval using baseline data (**A**, red) or baseline data with one follow up visit (**B**, purple) from the predictive logistic regression model are shown. Higher outcome scores together with positive coefficients influence the prediction towards the slow-progressing subtype and towards the fast-progressing subtype if coefficients are negative.*
*Abbreviations: BL: Baseline, Δ: Difference from first visit to baseline, MoCA: Montreal Cognitive Assessment, PIGD: Postural Instability and Gait Dysfunction score, SCOPA: Scales for Outcomes in Parkinson's Disease-Autonomic Dysfunction, UPDRS: Unified Parkinson's Disease Rating Scale.*

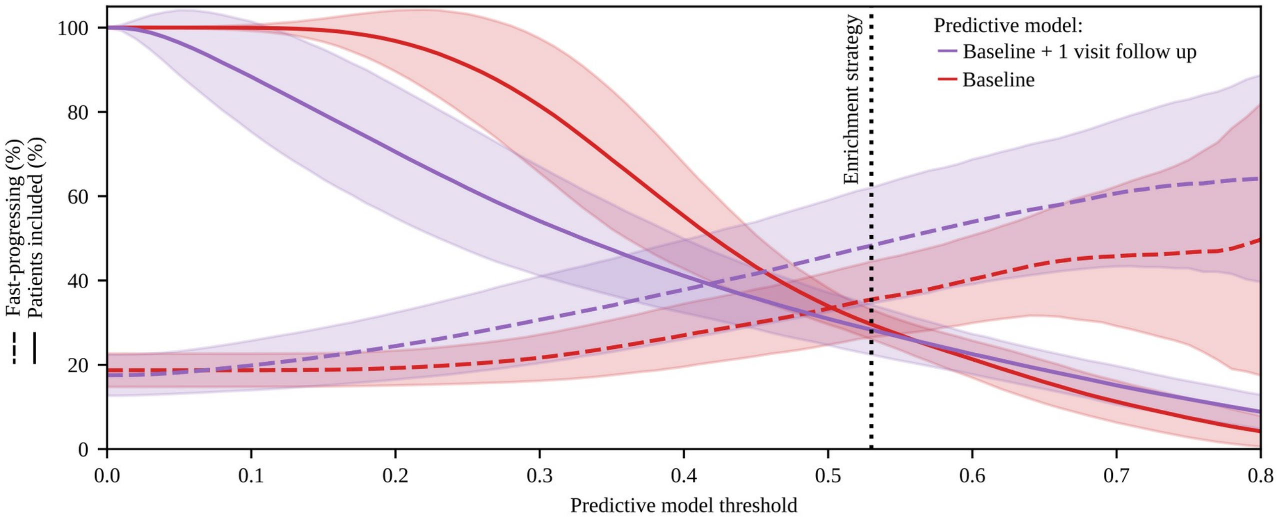

***Supplementary Figure 9: Trade-off between enriching fast-progressing PwPD and number of eligible PwPD***
*Depending on the chosen threshold applied to the logistic predictive model, the percentage of fast-progressing PwPD (dashed lines) and the percentage of PwPD being still eligible for study inclusion (solid lines) changes. Curves are displayed for the predictive model using baseline data (red) and the predictive model using data from baseline and one follow-up visit (purple). The vertical dotted line indicates the threshold of the predictive model, where still 30% of PwPD are eligible for study inclusion and a 47% enrichment of fast-progressing PwPD is achieved using the predictive model based on baseline data and one follow-up visit.*

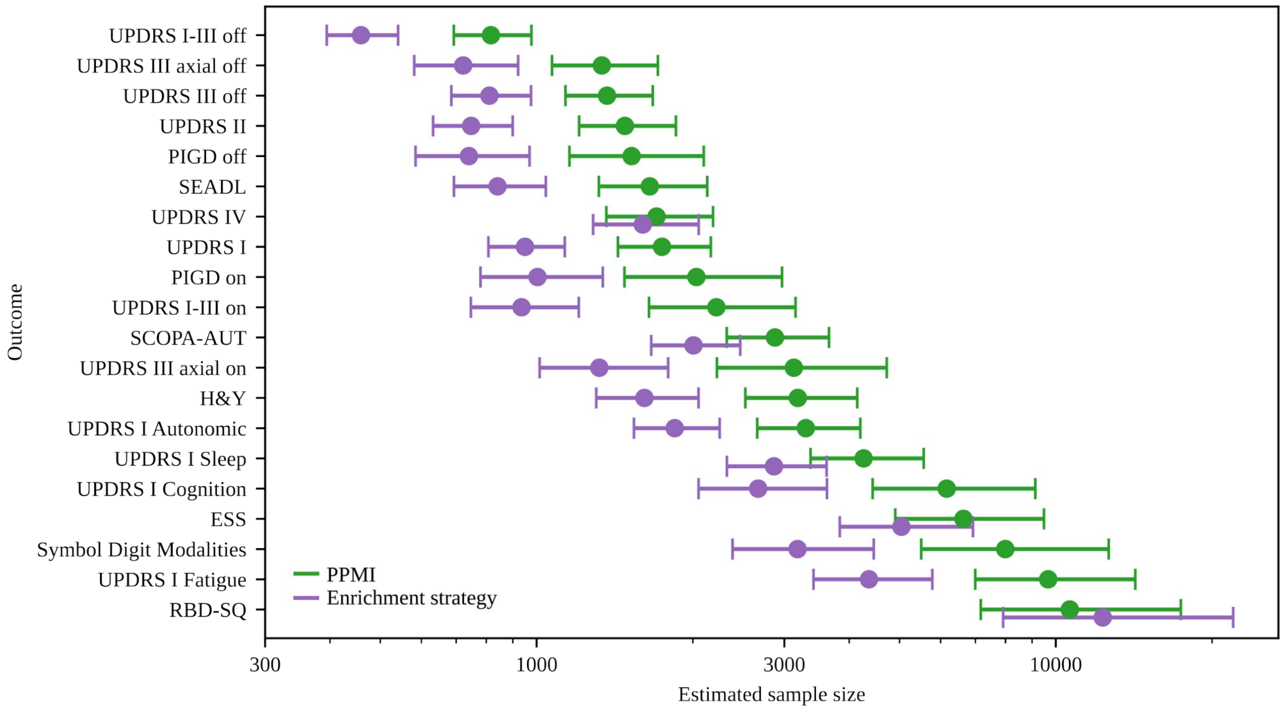

***Supplementary Figure 10: Subtype enrichment for sample size reduction in clinical trials using different outcomes***
*Required sample sizes for a clinical trial depending on the clinical score used as primary outcome in the trial. The required sample sizes calculated using the default PPMI cohort are shown in green. The required sample sizes using the enrichment strategy from Fig. 6A are shown in purple. Mean estimate and 95% confidence intervals are shown. Sample sizes were calculated for all outcomes listed in Supplementary Table 2 and assessed in PPMI. From these outcomes, only the 20 outcomes with lowest required sample sizes are shown. The estimated sample size is shown on a logarithmic axis.*
*Abbreviations: ESS: Epworth Sleepiness Scale, H&Y: Hoehn & Yahr, PIGD: Postural Instability and Gait Disorder score, RBD-SQ: REM Sleep Behavior Disorder Screening Questionnaire, SCOPA-AUT: Scales for Outcomes in Parkinson's Disease-Autonomic Dysfunction, SEADL: Schwab and England Activities of Daily Living Scale, UPDRS: Unified Parkinson's Disease Rating Scale.*

### Forest plots for symptom domain progression (in cohort)

### Forest plot for progression characteristics of symptom domain Axial & PIGD

| Study | Slow progressing Total | Slow progressing Mean | Slow progressing SD | Fast progressing Total | Fast progressing Mean | Fast progressing SD | SMD | 95%−CI |
|---|---|---|---|---|---|---|---|---|
| **Cohort = PPMI** | | | | | | | | |
| UPDRS III axial off | 335 | 0.16 | 0.0777 | 74 | 0.52 | 0.2251 | −3.04 | [−3.37; −2.71] |
| UPDRS III axial on | 300 | 0.09 | 0.0855 | 66 | 0.43 | 0.2922 | −2.33 | [−2.64; −2.01] |
| PIGD off | 335 | 0.13 | 0.0917 | 74 | 0.61 | 0.3265 | −2.97 | [−3.30; −2.65] |
| PIGD on | 300 | 0.10 | 0.0823 | 66 | 0.48 | 0.3223 | −2.49 | [−2.81; −2.16] |
| **Random effects model** | **1270** | | | **280** | | | **−2.71** | **[−3.05; −2.36]** |
| **Cohort = ICEBERG** | | | | | | | | |
| UPDRS III axial off | 111 | 0.09 | 0.0004 | 42 | 0.36 | 0.1002 | −5.16 | [−5.85; −4.48] |
| UPDRS III axial on | 105 | 0.10 | 0.0011 | 41 | 0.17 | 0.1309 | −1.02 | [−1.40; −0.64] |
| GABS Examination | 112 | 0.04 | 0.0191 | 42 | 0.12 | 0.1273 | −1.26 | [−1.64; −0.87] |
| GABS Questionaire | 112 | 0.03 | 0.0138 | 42 | 0.15 | 0.0932 | −2.41 | [−2.86; −1.96] |
| PIGD off | 111 | 0.06 | 0.0154 | 42 | 0.24 | 0.0735 | −4.52 | [−5.14; −3.89] |
| PIGD on | 105 | 0.07 | 0.0042 | 41 | 0.23 | 0.1364 | −2.22 | [−2.67; −1.78] |
| **Random effects model** | **656** | | | **250** | | | **−2.75** | **[−4.10; −1.39]** |
| **Cohort = LuxPARK** | | | | | | | | |
| UPDRS III axial on | 380 | 0.13 | 0.0510 | 140 | 0.30 | 0.1008 | −2.44 | [−2.68; −2.19] |
| FOGAC | 63 | 0.11 | 0.0767 | 17 | 0.11 | 0.1963 | 0.03 | [−0.51; 0.57] |
| FOGQ | 166 | 0.17 | 0.0954 | 64 | 0.31 | 0.2075 | −0.97 | [−1.28; −0.67] |
| NFOGQ | 19 | 0.24 | 0.0939 | 5 | 0.91 | 1.3814 | −1.10 | [−2.14; −0.05] |
| PDQ39 Mobility | 407 | 0.18 | 0.0835 | 152 | 0.44 | 0.1897 | −2.12 | [−2.34; −1.89] |
| PIGD on | 380 | 0.14 | 0.0843 | 140 | 0.33 | 0.1427 | −1.89 | [−2.11; −1.66] |
| TuG | 154 | 0.03 | 0.0318 | 42 | 0.05 | 0.0269 | −0.77 | [−1.12; −0.42] |
| **Random effects model** | **1569** | | | **560** | | | **−1.35** | **[−2.00; −0.70]** |
| **Random effects model** | **3495** | | | **1090** | | | **−2.15** | **[−2.77; −1.54]** |

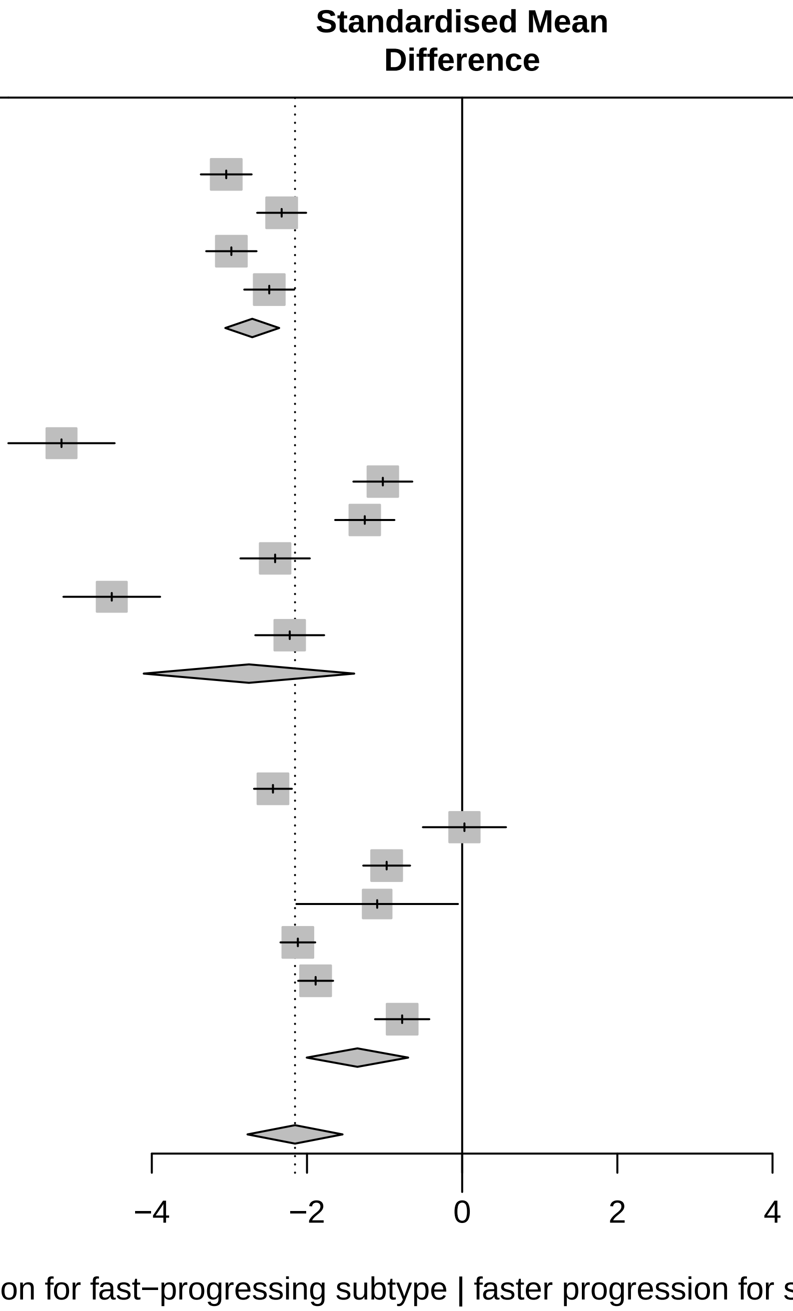

Three−level metaanalysis using random effects to calculate an overall SMD estimate for Axial & PIGD across cohorts.
The dashed line indicates the overall mean SMD estimate. The solid line indicates no effect.

### Forest plot for progression characteristics of symptom domain Depression

| Study | Slow progressing Total | Slow progressing Mean | Slow progressing SD | Fast progressing Total | Fast progressing Mean | Fast progressing SD | SMD | 95%−CI |
|---|---|---|---|---|---|---|---|---|
| **Cohort = LuxPARK** | | | | | | | | |
| BDI | 406 | 0.04 | 0.0176 | 149 | 0.14 | 0.0358 | −4.19 | [−4.50; −3.88] |
| NMSQ Depression | 408 | 1.04 | 0.5080 | 152 | 2.15 | 0.2609 | −2.43 | [−2.67; −2.20] |
| PDQ39 Depression | 407 | 0.06 | 0.0499 | 152 | 0.18 | 0.0605 | −2.31 | [−2.54; −2.08] |
| UPDRS I Depression | 408 | 0.84 | 0.1665 | 153 | 1.53 | 0.4505 | −2.51 | [−2.75; −2.28] |
| **Random effects model** | **1629** | | | **606** | | | **−2.86** | **[−3.72; −1.99]** |
| **Cohort = PPMI** | | | | | | | | |
| GDS | 324 | 0.05 | 0.1681 | 73 | 0.35 | 0.1441 | −1.78 | [−2.06; −1.50] |
| UPDRS I Depression | 335 | 1.16 | 1.4484 | 74 | 2.83 | 2.1275 | −1.04 | [−1.31; −0.78] |
| **Random effects model** | **659** | | | **147** | | | **−1.41** | **[−2.13; −0.69]** |
| **Cohort = ICEBERG** | | | | | | | | |
| HADS depression | 112 | 0.10 | 0.0186 | 42 | 0.16 | 0.0578 | −1.60 | [−1.99; −1.20] |
| NMSQ Depression | 112 | 2.00 | 0.4071 | 42 | 1.42 | 1.4046 | 0.72 | [ 0.35; 1.08] |
| UPDRS I Depression | 112 | 1.54 | 0.5269 | 42 | 0.83 | 1.2172 | 0.91 | [ 0.54; 1.28] |
| **Random effects model** | **336** | | | **126** | | | **0.01** | **[−1.56; 1.59]** |
| **Random effects model** | **2624** | | | **879** | | | **−1.58** | **[−2.64; −0.53]** |

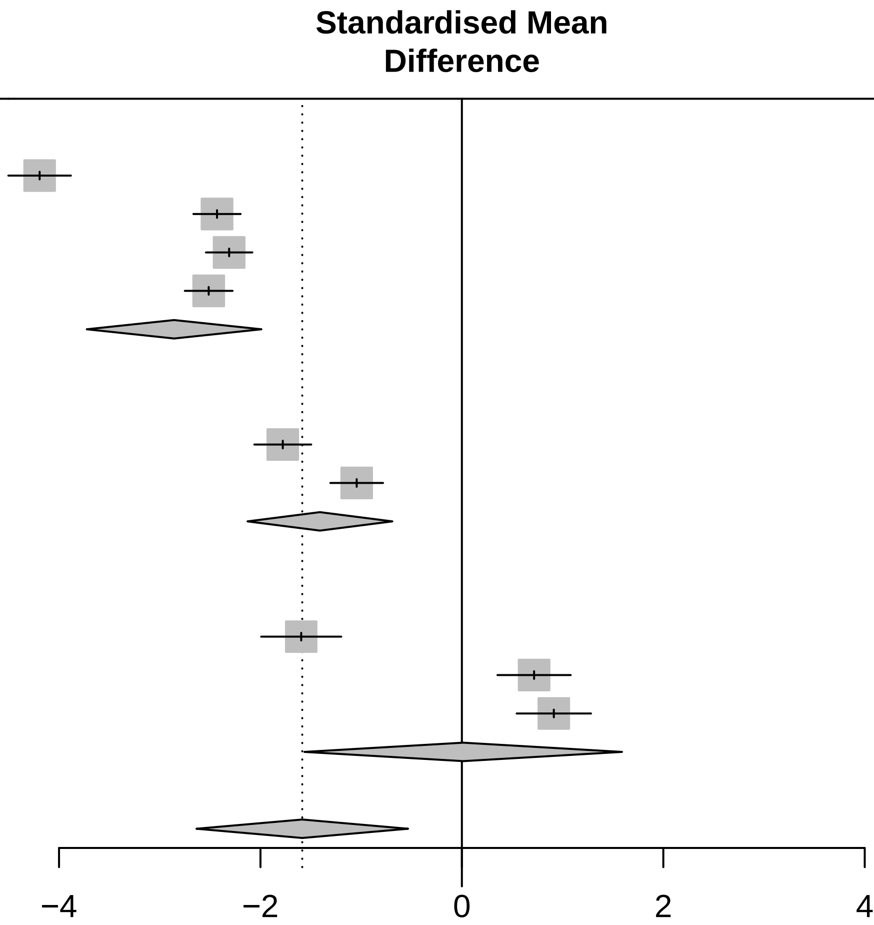

<− Faster progression for fast−progressing subtype | faster progression for slow−progressing subtype −>

Three−level metaanalysis using random effects to calculate an overall SMD estimate for Depression across cohorts.
The dashed line indicates the overall mean SMD estimate. The solid line indicates no effect.

### Forest plot for progression characteristics of symptom domain Overall severity

| Study | Slow progressing Total | Slow progressing Mean | Slow progressing SD | Fast progressing Total | Fast progressing Mean | Fast progressing SD | SMD | 95%−CI |
|---|---|---|---|---|---|---|---|---|
| **Cohort = ICEBERG** | | | | | | | | |
| CGIS | 112 | 1.07 | 0.0666 | 42 | 2.39 | 0.8408 | −2.99 | [−3.48; −2.50] |
| H&Y | 112 | −0.40 | 0.3209 | 42 | 0.57 | 0.1069 | −3.45 | [−3.97; −2.92] |
| SEADL | 112 | 0.05 | 0.0260 | 42 | 0.13 | 0.1255 | −1.17 | [−1.55; −0.79] |
| UPDRS I−III off | 111 | 0.14 | 0.0092 | 42 | 0.30 | 0.0654 | −4.65 | [−5.28; −4.01] |
| UPDRS I−III on | 105 | 0.11 | 0.0103 | 41 | 0.23 | 0.0362 | −5.75 | [−6.51; −4.99] |
| **Random effects model** | **552** | | | **209** | | | **−3.58** | **[−5.10; −2.06]** |
| **Cohort = LuxPARK** | | | | | | | | |
| FAQ | 354 | 0.09 | 0.0549 | 124 | 0.30 | 0.1342 | −2.47 | [−2.73; −2.22] |
| H&Y | 408 | 2.37 | 1.5849 | 153 | 2.93 | 2.3835 | −0.31 | [−0.49; −0.12] |
| PDQ39 | 403 | 0.10 | 0.0470 | 149 | 0.28 | 0.1213 | −2.33 | [−2.56; −2.10] |
| UPDRS I−III on | 377 | 0.14 | 0.0350 | 138 | 0.34 | 0.1151 | −2.99 | [−3.26; −2.72] |
| **Random effects model** | **1542** | | | **564** | | | **−2.02** | **[−3.18; −0.86]** |
| **Cohort = PPMI** | | | | | | | | |
| H&Y | 335 | 4.88 | 1.8738 | 74 | 4.44 | 3.7381 | 0.19 | [−0.06; 0.44] |
| SEADL | 335 | 0.14 | 0.0788 | 74 | 0.58 | 0.2890 | −3.09 | [−3.42; −2.76] |
| UPDRS I−III off | 335 | 0.26 | 0.1092 | 74 | 0.64 | 0.1259 | −3.38 | [−3.72; −3.04] |
| UPDRS I−III on | 300 | 0.11 | 0.0952 | 66 | 0.46 | 0.2094 | −2.77 | [−3.10; −2.44] |
| **Random effects model** | **1305** | | | **288** | | | **−2.26** | **[−3.88; −0.63]** |
| **Random effects model** | **3399** | | | **1061** | | | **−2.68** | **[−3.55; −1.82]** |

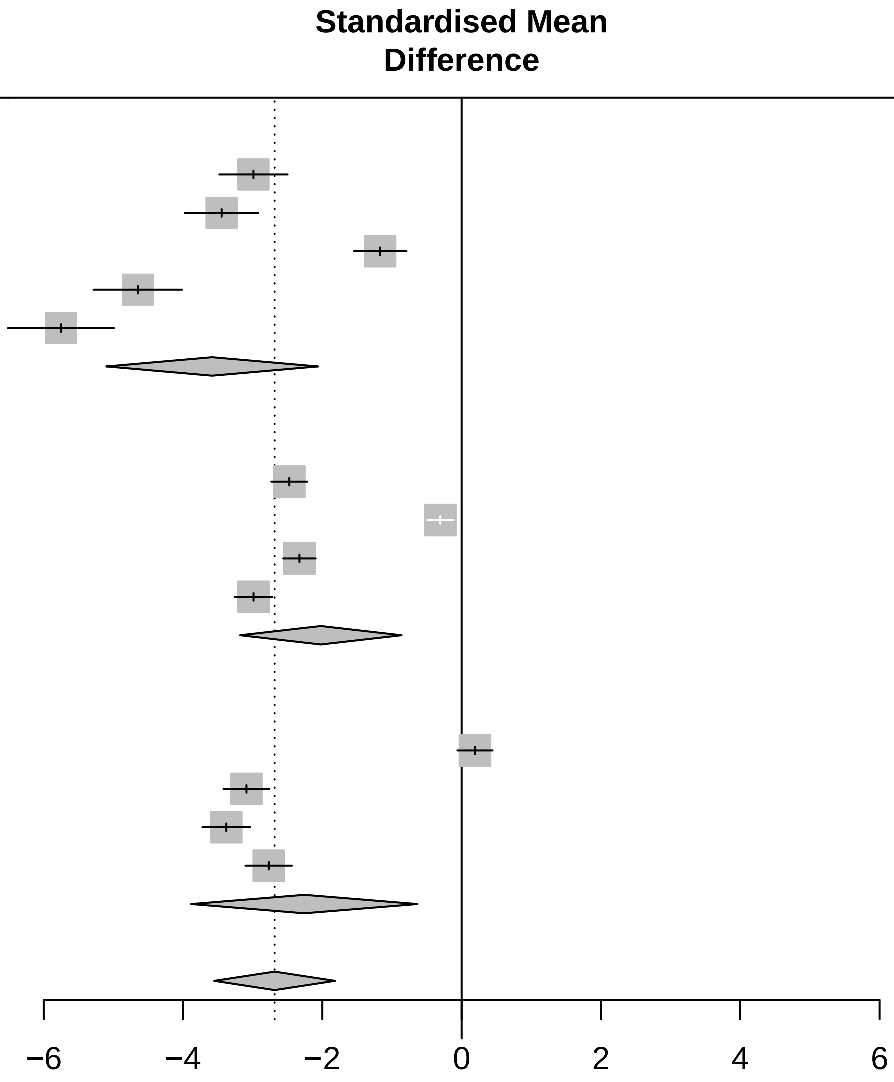

Three−level metaanalysis using random effects to calculate an overall SMD estimate for Overall severity across cohorts. The dashed line indicates the overall mean SMD estimate. The solid line indicates no effect.

### Forest plot for progression characteristics of symptom domain Apathy

| Study | Slow progressing Total | Slow progressing Mean | Slow progressing SD | Fast progressing Total | Fast progressing Mean | Fast progressing SD | SMD | 95%−CI |
|---|---|---|---|---|---|---|---|---|
| Cohort = ICEBERG | | | | | | | | |
| DAS | 103 | 0.08 | 0.0179 | 38 | 0.11 | 0.0428 | −1.11 | [ −1.50; −0.71] |
| SAS | 104 | 0.08 | 0.0303 | 41 | 0.12 | 0.0322 | −1.39 | [ −1.78; −0.99] |
| UPDRS I Apathy | 112 | 1.18 | 0.4818 | 42 | 1.44 | 0.8770 | −0.41 | [ −0.77; −0.06] |
| Random effects model | 319 | | | 121 | | | −0.96 | [ −1.54; −0.39] |
| Cohort = LuxPARK | | | | | | | | |
| SAS | 405 | 0.05 | 0.0480 | 153 | 0.12 | 0.0369 | −1.52 | [ −1.73; −1.31] |
| UPDRS I Apathy | 408 | 0.73 | 0.0431 | 153 | 1.86 | 0.2459 | −8.48 | [ −9.01; −7.95] |
| Random effects model | 813 | | | 306 | | | −4.99 | [−11.81; 1.82] |
| Cohort = PPMI | | | | | | | | |
| UPDRS I Apathy | 335 | 2.14 | 1.2773 | 74 | 4.63 | 2.7266 | −1.52 | [ −1.79; −1.25] |
| **Random effects model** | **1467** | | | **501** | | | **−2.40** | **[ −4.80; −0.00]** |

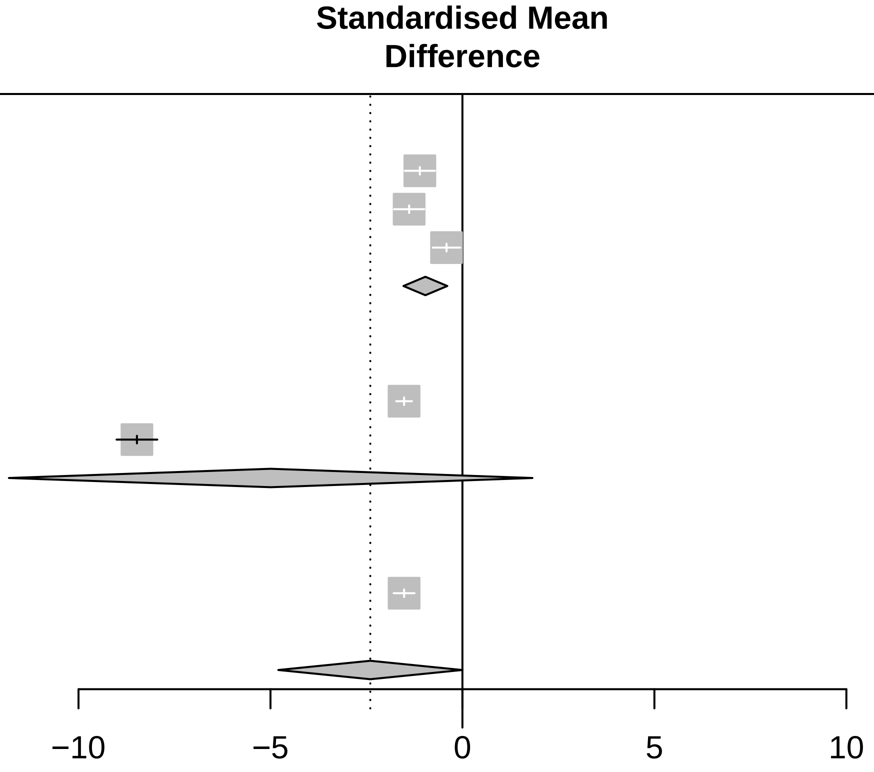

<− Faster progression for fast−progressing subtype | faster progression for slow−progressing subtype −>

Three−level metaanalysis using random effects to calculate an overall SMD estimate for Apathy across cohorts.
The dashed line indicates the overall mean SMD estimate. The solid line indicates no effect.

### Forest plot for progression characteristics of symptom domain Sleep

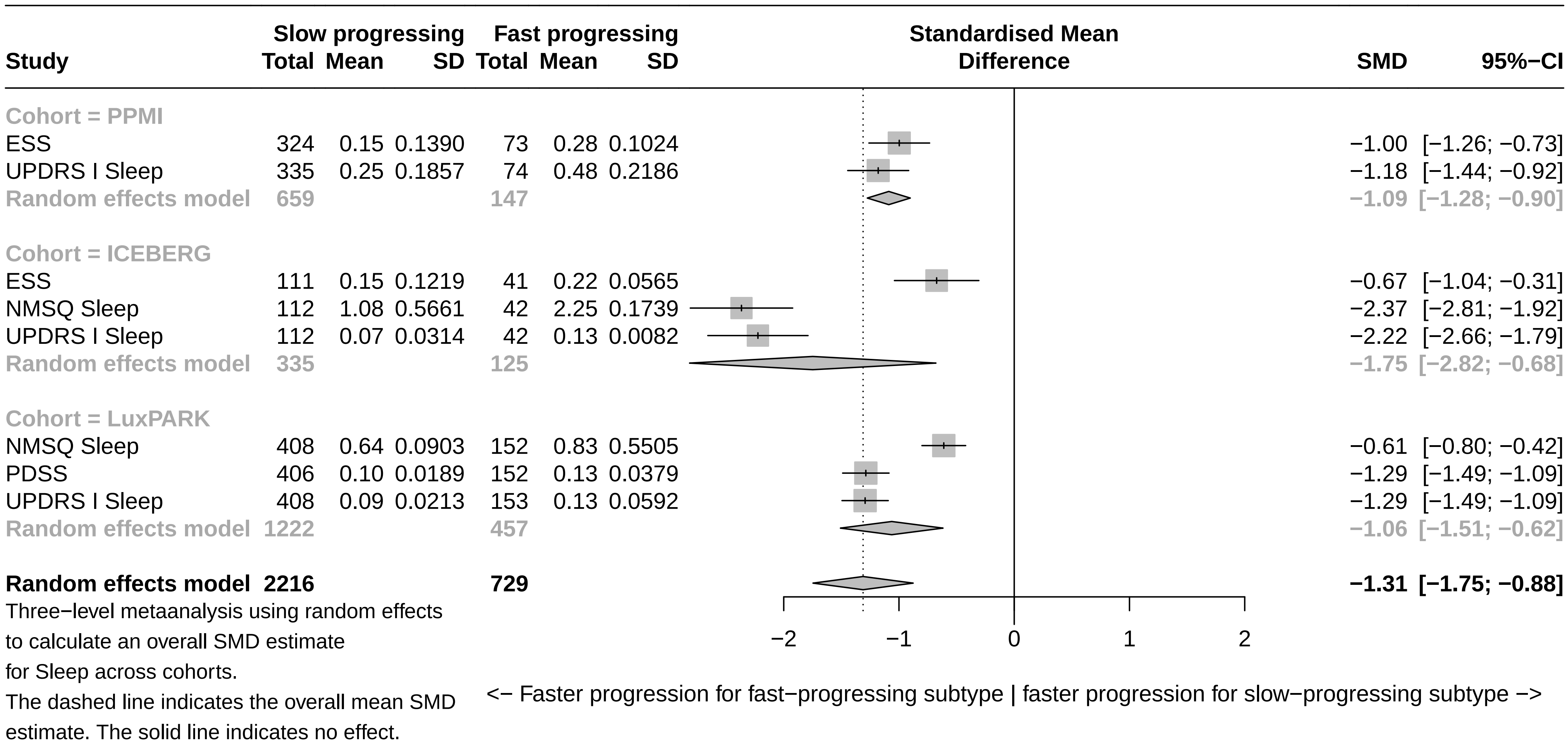

### Forest plot for progression characteristics of symptom domain Overall cognition

| Study | Slow progressing Total | Slow progressing Mean | Slow progressing SD | Fast progressing Total | Fast progressing Mean | Fast progressing SD | SMD | 95%−CI |
|---|---|---|---|---|---|---|---|---|
| **Cohort = LuxPARK** | | | | | | | | |
| FAB | 42 | 0.02 | 0.0108 | 15 | 0.13 | 0.1039 | −1.98 | [−2.68; −1.28] |
| MoCA | 408 | 0.03 | 0.0236 | 153 | 0.18 | 0.1102 | −2.41 | [−2.65; −2.18] |
| NMSQ Cognition | 408 | 1.31 | 0.6814 | 151 | 2.55 | 1.0627 | −1.54 | [−1.75; −1.33] |
| PDQ39 Cognition | 408 | 0.08 | 0.0242 | 152 | 0.16 | 0.0971 | −1.47 | [−1.68; −1.27] |
| SIQCDE | 351 | 0.03 | 0.0424 | 122 | 0.08 | 0.0683 | −0.99 | [−1.21; −0.78] |
| UPDRS I Cognition | 408 | 1.10 | 1.1392 | 153 | 2.17 | 1.5897 | −0.84 | [−1.03; −0.65] |
| **Random effects model** | **2025** | | | **746** | | | **−1.52** | **[−2.00; −1.04]** |
| **Cohort = ICEBERG** | | | | | | | | |
| FAB | 104 | 0.05 | 0.0378 | 41 | 0.02 | 0.0744 | 0.57 | [ 0.20; 0.94] |
| MATTIS | 104 | 0.05 | 0.0452 | 41 | 0.02 | 0.0664 | 0.58 | [ 0.22; 0.95] |
| MoCA | 112 | 0.05 | 0.0549 | 42 | 0.10 | 0.0770 | −0.80 | [−1.17; −0.43] |
| NMSQ Cognition | 112 | 1.69 | 0.2667 | 42 | 1.90 | 0.3774 | −0.70 | [−1.07; −0.34] |
| UPDRS I Cognition | 112 | 0.86 | 0.3592 | 42 | 1.21 | 1.2353 | −0.49 | [−0.85; −0.13] |
| **Random effects model** | **544** | | | **208** | | | **−0.17** | **[−0.77; 0.43]** |
| **Cohort = PPMI** | | | | | | | | |
| MoCA | 320 | 0.02 | 0.0720 | 71 | 0.29 | 0.2697 | −2.08 | [−2.37; −1.78] |
| UPDRS I Cognition | 335 | 1.64 | 3.2067 | 74 | 3.73 | 5.1644 | −0.57 | [−0.83; −0.32] |
| **Random effects model** | **655** | | | **145** | | | **−1.32** | **[−2.80; 0.15]** |
| **Random effects model** | **3224** | | | **1099** | | | **−0.98** | **[−1.48; −0.48]** |

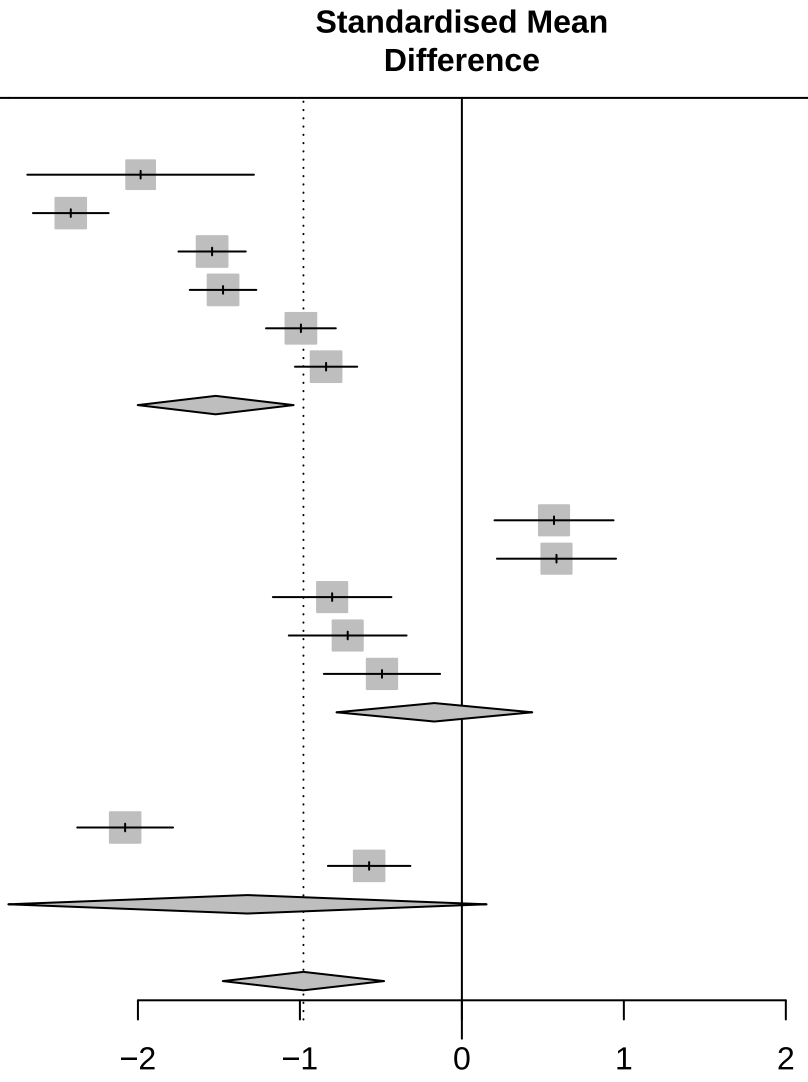

Three−level metaanalysis using random effects
to calculate an overall SMD estimate
for Overall cognition across cohorts.
The dashed line indicates the overall mean SMD
estimate. The solid line indicates no effect.

<− Faster progression for fast−progressing subtype | faster progression for slow−progressing subtype −>

### Forest plot for progression characteristics of symptom domain Conceptualization

| Study | Slow progressing Total | Slow progressing Mean | Slow progressing SD | Fast progressing Total | Fast progressing Mean | Fast progressing SD | SMD | 95%−CI |
|---|---|---|---|---|---|---|---|---|
| **Cohort = LuxPARK** | | | | | | | | |
| FAB Conceptualization | 43 | −1.31 | 0.2084 | 16 | −2.15 | 1.9839 | 0.80 | [ 0.21; 1.40] |
| MOCA Abstraction | 408 | 0.16 | 0.2392 | 153 | 0.43 | 0.1078 | −1.29 | [−1.49; −1.09] |
| **Random effects model** | **451** | | | **169** | | | **−0.26** | **[−2.31; 1.79]** |
| **Cohort = ICEBERG** | | | | | | | | |
| FAB Conceptualization | 104 | 0.97 | 1.5829 | 41 | 0.87 | 0.2338 | 0.08 | [−0.28; 0.44] |
| MATTIS Conzeptualization | 104 | 0.00 | 0.0741 | 41 | 0.05 | 0.0337 | −0.74 | [−1.11; −0.37] |
| MOCA Abstraction | 112 | 0.53 | 0.6543 | 42 | 1.72 | 0.5240 | −1.90 | [−2.31; −1.48] |
| **Random effects model** | **320** | | | **124** | | | **−0.85** | **[−1.97; 0.27]** |
| **Cohort = PPMI** | | | | | | | | |
| MOCA Abstraction | 320 | −1.90 | 0.9625 | 71 | 1.57 | 1.5654 | −3.16 | [−3.50; −2.82] |
| **Random effects model** | **1091** | | | **364** | | | **−1.04** | **[−2.17; 0.08]** |

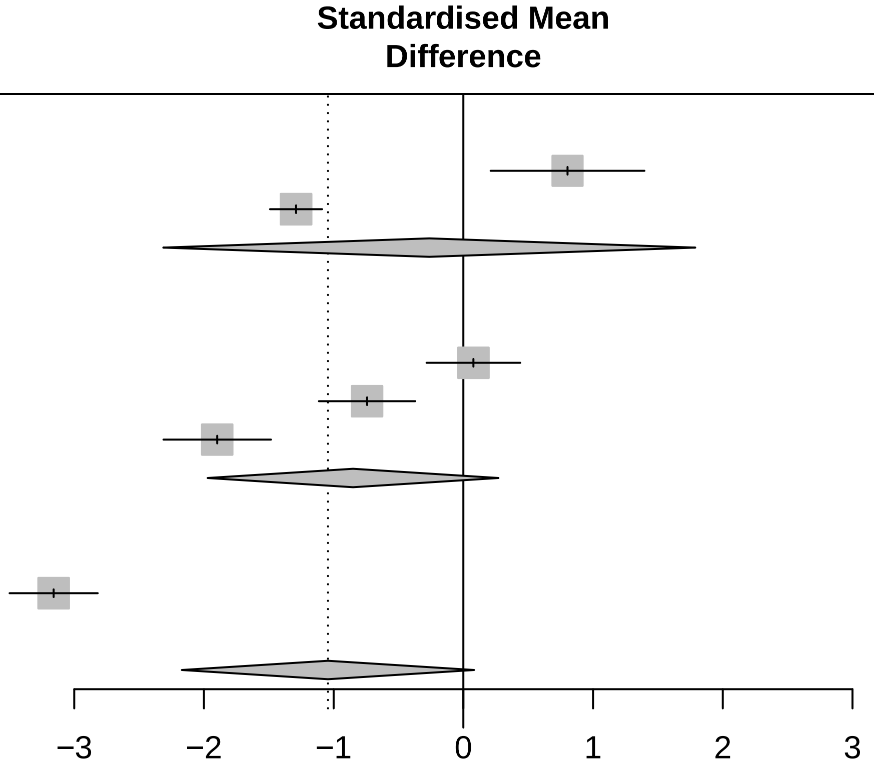

Three−level metaanalysis using random effects to calculate an overall SMD estimate for Conceptualization across cohorts. The dashed line indicates the overall mean SMD estimate. The solid line indicates no effect.

### Forest plot for progression characteristics of symptom domain Visuo−executive

| Study | Slow progressing Total | Slow progressing Mean | Slow progressing SD | Fast progressing Total | Fast progressing Mean | Fast progressing SD | SMD | 95%−CI |
|---|---|---|---|---|---|---|---|---|
| Cohort = ICEBERG | | | | | | | | |
| FAB 3−6 | 104 | 0.06 | 0.0473 | 41 | 0.02 | 0.1003 | 0.64 | [ 0.27; 1.01] |
| MATTIS Initiation + Construction | 104 | 0.04 | 0.0456 | 41 | 0.03 | 0.0611 | 0.33 | [−0.03; 0.69] |
| MOCA Visuspatial/Executive | 112 | 0.81 | 0.5368 | 42 | 1.37 | 0.1911 | −1.20 | [−1.58; −0.82] |
| Random effects model | 320 | | | 124 | | | −0.08 | [−1.19; 1.04] |
| Cohort = LuxPARK | | | | | | | | |
| FAB 3−6 | 42 | 0.07 | 0.0238 | 15 | 0.09 | 0.0408 | −0.74 | [−1.34; −0.13] |
| MOCA Visuspatial/Executive | 408 | 0.86 | 0.0973 | 153 | 2.12 | 0.3547 | −6.23 | [−6.64; −5.82] |
| Trailmaking A | 392 | 0.02 | 0.0225 | 134 | 0.07 | 0.0747 | −1.17 | [−1.38; −0.96] |
| Trailmaking B | 357 | 0.04 | 0.0371 | 99 | 0.09 | 0.0523 | −1.20 | [−1.44; −0.97] |
| Random effects model | 1199 | | | 401 | | | −2.34 | [−4.89; 0.22] |
| Cohort = PPMI | | | | | | | | |
| Judgement Line Orientation | 320 | 0.04 | 0.0357 | 71 | 0.17 | 0.1483 | −1.84 | [−2.13; −1.55] |
| MOCA Visuspatial/Executive | 320 | 0.77 | 1.2703 | 71 | 4.01 | 2.9805 | −1.89 | [−2.18; −1.60] |
| Symbol Digit Modalities | 320 | 0.05 | 0.0727 | 71 | 0.21 | 0.1285 | −1.81 | [−2.10; −1.52] |
| Random effects model | 960 | | | 213 | | | −1.85 | [−2.02; −1.68] |
| **Random effects model** | **2479** | | | **738** | | | **−1.51** | **[−2.67; −0.35]** |

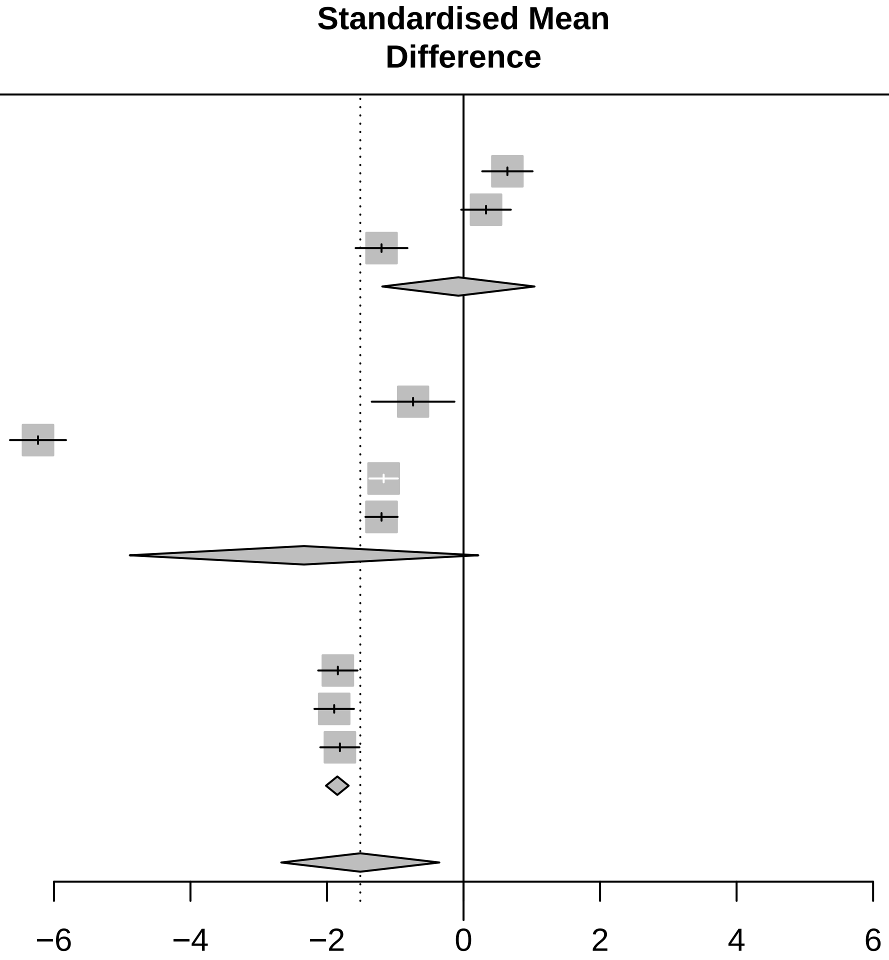

<− Faster progression for fast−progressing subtype | faster progression for slow−progressing subtype −>

Three−level metaanalysis using random effects to calculate an overall SMD estimate for Visuo−executive across cohorts. The dashed line indicates the overall mean SMD estimate. The solid line indicates no effect.

### Forest plot for progression characteristics of symptom domain Language

| Study | Slow progressing Total | Slow progressing Mean | Slow progressing SD | Fast progressing Total | Fast progressing Mean | Fast progressing SD | SMD | 95%−CI |
|---|---|---|---|---|---|---|---|---|
| **Cohort = LuxPARK** | | | | | | | | |
| FAB VFT | 43 | −0.41 | 1.4483 | 16 | 2.00 | 0.2269 | −1.90 | [−2.58; −1.23] |
| MOCA Language + Naming | 408 | 0.03 | 0.0199 | 153 | 0.12 | 0.0251 | −4.32 | [−4.63; −4.00] |
| VFT phonematic F | 332 | −0.01 | 0.0148 | 129 | 0.03 | 0.0095 | −2.89 | [−3.16; −2.61] |
| VFT phonematic S | 50 | −0.08 | 0.0356 | 21 | 0.11 | 0.0389 | −5.18 | [−6.19; −4.17] |
| **Random effects model** | **833** | | | **319** | | | **−3.55** | **[−4.94; −2.15]** |
| **Cohort = ICEBERG** | | | | | | | | |
| FAB VFT | 104 | −0.35 | 0.5878 | 41 | −1.33 | 1.6903 | 0.95 | [ 0.57; 1.33] |
| MOCA Language + Naming | 112 | 0.03 | 0.0362 | 42 | 0.05 | 0.0513 | −0.60 | [−0.96; −0.24] |
| VFT phonematic F | 112 | 0.03 | 0.0353 | 42 | 0.03 | 0.0767 | −0.12 | [−0.47; 0.24] |
| VFT phonematic S | 100 | −0.00 | 0.0072 | 41 | 0.03 | 0.0256 | −2.05 | [−2.49; −1.61] |
| VFT semantic supermarket | 99 | 0.06 | 0.0162 | 41 | 0.01 | 0.0160 | 3.22 | [ 2.69; 3.74] |
| **Random effects model** | **527** | | | **207** | | | **0.28** | **[−1.44; 1.99]** |
| **Cohort = PPMI** | | | | | | | | |
| MOCA Language + Naming | 320 | −0.04 | 0.0632 | 71 | 0.06 | 0.0917 | −1.44 | [−1.72; −1.16] |
| VFT phonematic F | 320 | −0.05 | 0.0624 | 71 | 0.04 | 0.0871 | −1.34 | [−1.61; −1.06] |
| VFT semantic sum | 320 | 0.02 | 0.0793 | 71 | 0.12 | 0.1057 | −1.15 | [−1.42; −0.88] |
| **Random effects model** | **960** | | | **213** | | | **−1.31** | **[−1.48; −1.14]** |
| **Random effects model** | **2320** | | | **739** | | | **−1.39** | **[−2.64; −0.14]** |

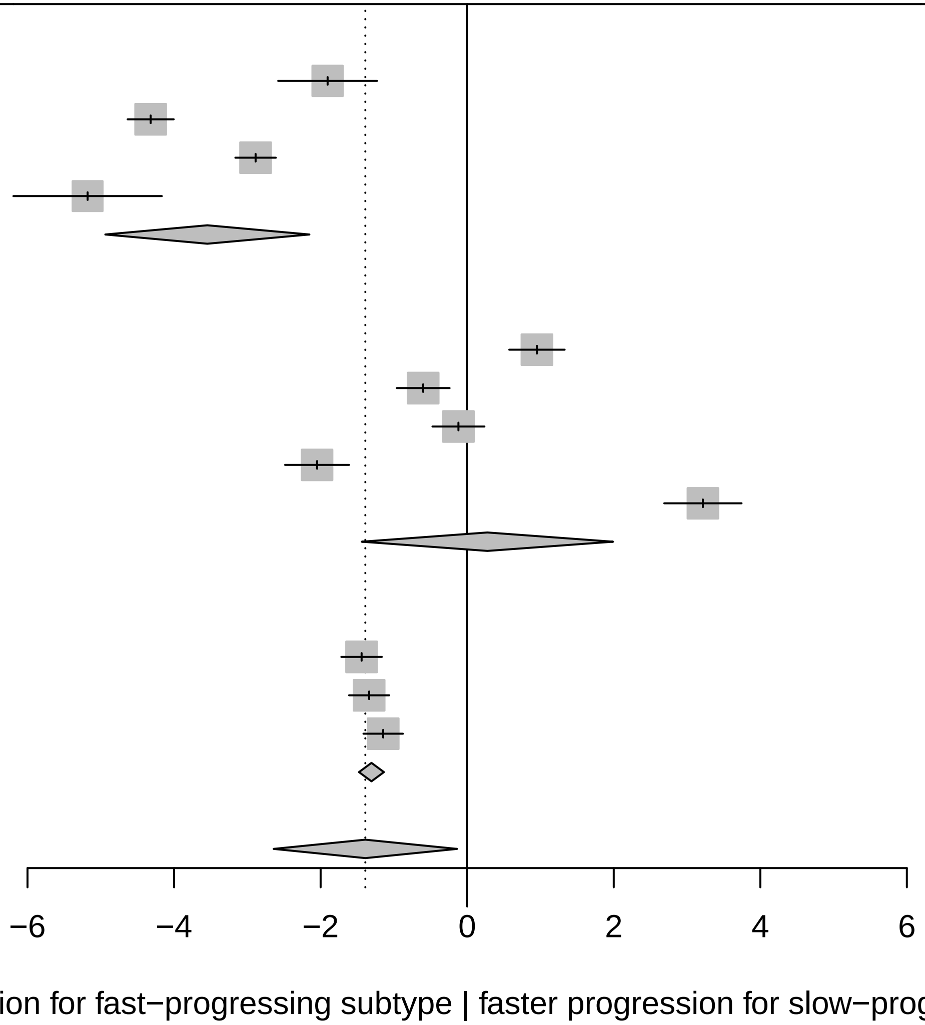

Three−level metaanalysis using random effects to calculate an overall SMD estimate for Language across cohorts.
The dashed line indicates the overall mean SMD estimate. The solid line indicates no effect.

### Forest plot for progression characteristics of symptom domain Anxiety

| Study | Slow progressing Total | Slow progressing Mean | Slow progressing SD | Fast progressing Total | Fast progressing Mean | Fast progressing SD | SMD | 95%−CI |
|---|---|---|---|---|---|---|---|---|
| **Cohort = ICEBERG** | | | | | | | | |
| HADS anxiety | 112 | 0.02 | 0.0221 | 42 | 0.08 | 0.0598 | −1.76 | [−2.16; −1.35] |
| NMSQ Anxiety | 111 | 1.38 | 0.8609 | 42 | 1.29 | 1.9033 | 0.07 | [−0.29; 0.42] |
| UPDRS I Anxiety | 112 | 0.90 | 0.0762 | 42 | 0.84 | 1.9574 | 0.06 | [−0.29; 0.42] |
| **Random effects model** | **335** | | | **126** | | | **−0.54** | **[−1.73; 0.65]** |
| **Cohort = LuxPARK** | | | | | | | | |
| NMSQ Anxiety | 408 | 1.08 | 0.3337 | 151 | 1.74 | 0.3269 | −2.00 | [−2.22; −1.77] |
| PDQ39 Anxiety | 408 | 1.03 | 0.3142 | 153 | 1.67 | 0.5120 | −1.70 | [−1.91; −1.49] |
| UPDRS I Anxiety | 408 | 0.64 | 0.1651 | 153 | 1.25 | 1.1298 | −1.01 | [−1.20; −0.81] |
| **Random effects model** | **1224** | | | **457** | | | **−1.57** | **[−2.14; −0.99]** |
| **Cohort = PPMI** | | | | | | | | |
| STA | 323 | 0.02 | 0.1127 | 72 | 0.23 | 0.0952 | −1.94 | [−2.23; −1.65] |
| UPDRS I Anxiety | 335 | 1.05 | 2.2489 | 74 | 3.37 | 2.2233 | −1.04 | [−1.30; −0.77] |
| **Random effects model** | **658** | | | **146** | | | **−1.48** | **[−2.37; −0.60]** |
| **Random effects model** | **2217** | | | **729** | | | **−1.17** | **[−1.75; −0.59]** |

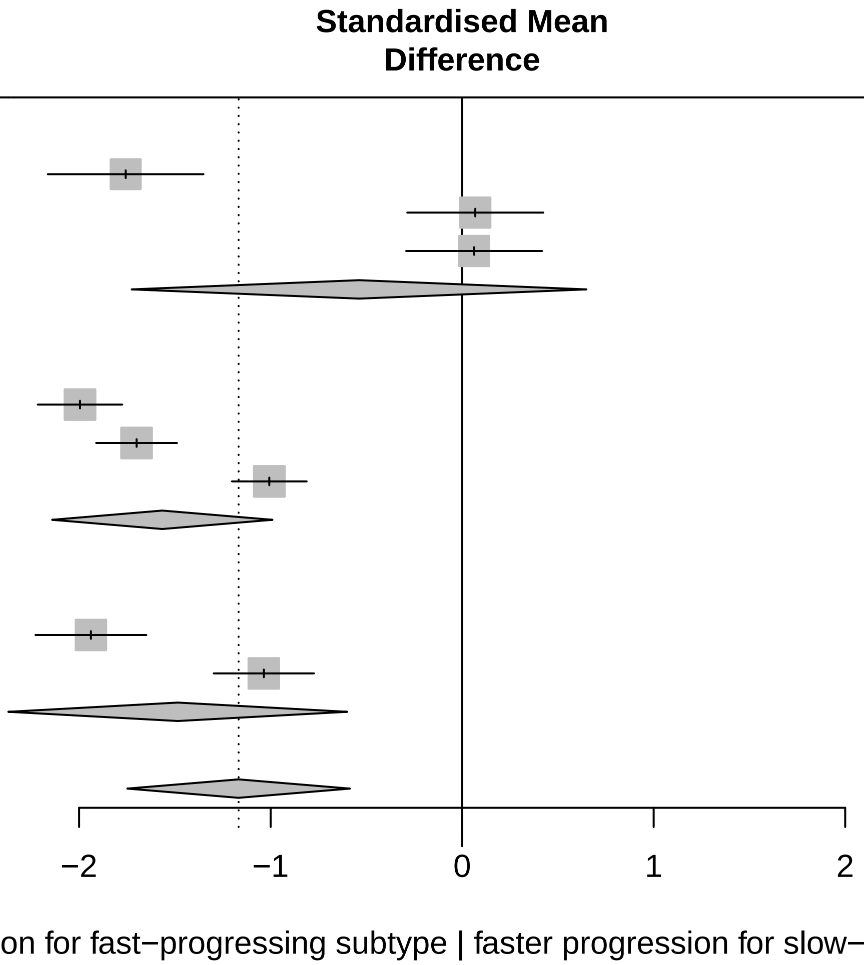

Three−level metaanalysis using random effects to calculate an overall SMD estimate for Anxiety across cohorts.
The dashed line indicates the overall mean SMD estimate. The solid line indicates no effect.

### Forest plot for progression characteristics of symptom domain Memory

| Study | Slow progressing Total | Slow progressing Mean | Slow progressing SD | Fast progressing Total | Fast progressing Mean | Fast progressing SD | SMD | 95%−CI |
|---|---|---|---|---|---|---|---|---|
| **Cohort = PPMI** | | | | | | | | |
| Hopkins Verbal Learning Test DR | 320 | 0.01 | 0.0245 | 71 | −0.00 | 0.0144 | 0.59 | [ 0.33; 0.85] |
| Hopkins Verbal Learning Test IR | 320 | 0.04 | 0.0872 | 71 | 0.17 | 0.1296 | −1.32 | [−1.60; −1.05] |
| MOCA Orientation + Memory | 320 | −0.01 | 0.0912 | 71 | 0.21 | 0.1801 | −1.97 | [−2.26; −1.68] |
| **Random effects model** | **960** | | | **213** | | | **−0.90** | **[−2.41; 0.61]** |
| **Cohort = ICEBERG** | | | | | | | | |
| MATTIS Memory | 104 | 0.07 | 0.0605 | 41 | 0.08 | 0.0382 | −0.21 | [−0.57; 0.16] |
| MOCA Orientation + Memory | 112 | 0.02 | 0.0179 | 42 | 0.03 | 0.0961 | −0.37 | [−0.72; −0.01] |
| NMSQ Memory | 111 | 1.19 | 1.4812 | 42 | 2.28 | 1.4312 | −0.74 | [−1.10; −0.37] |
| **Random effects model** | **327** | | | **125** | | | **−0.44** | **[−0.74; −0.13]** |
| **Cohort = LuxPARK** | | | | | | | | |
| MOCA Orientation + Memory | 408 | 0.01 | 0.0204 | 153 | 0.11 | 0.0920 | −2.05 | [−2.27; −1.83] |
| NMSQ Memory | 408 | 1.13 | 0.0606 | 151 | 2.30 | 1.2830 | −1.75 | [−1.96; −1.53] |
| PDQ39 Memory | 408 | 0.74 | 0.2966 | 152 | 1.58 | 1.5416 | −1.01 | [−1.20; −0.81] |
| SIQCDE Memory | 352 | 0.03 | 0.0459 | 122 | 0.07 | 0.0527 | −0.68 | [−0.89; −0.47] |
| **Random effects model** | **1576** | | | **578** | | | **−1.37** | **[−1.99; −0.75]** |
| **Random effects model** | **2863** | | | **916** | | | **−0.95** | **[−1.48; −0.43]** |

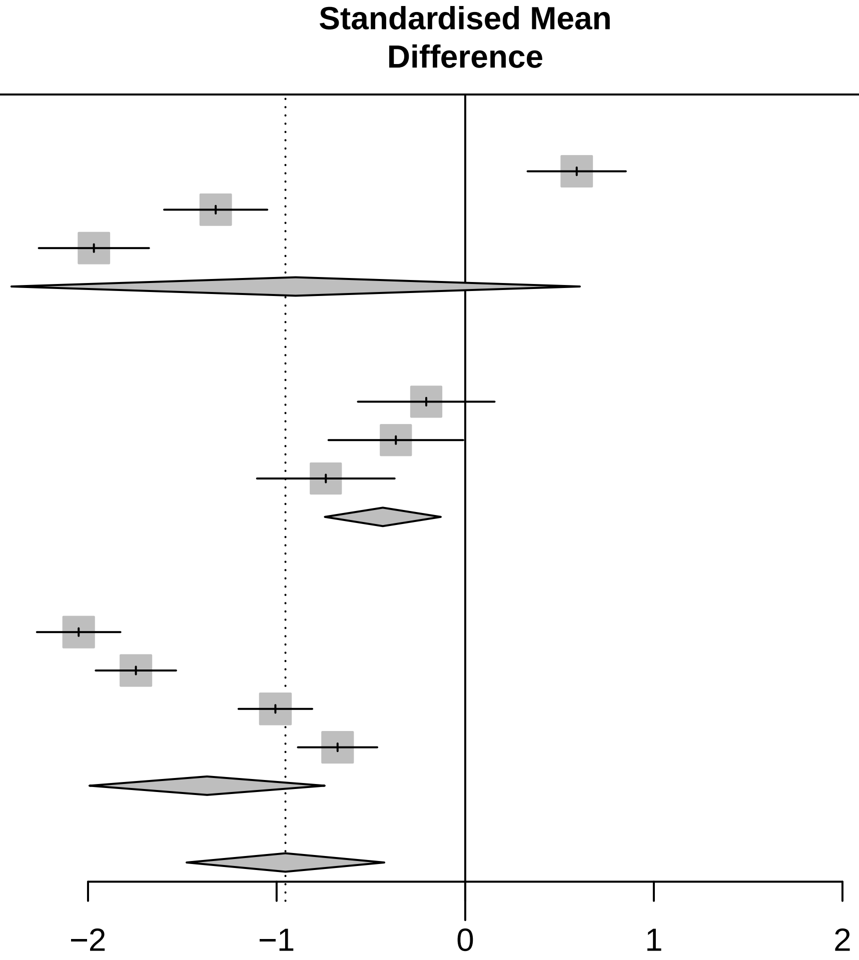

<− Faster progression for fast−progressing subtype | faster progression for slow−progressing subtype −>

Three−level metaanalysis using random effects to calculate an overall SMD estimate for Memory across cohorts.
The dashed line indicates the overall mean SMD estimate. The solid line indicates no effect.

### Forest plot for progression characteristics of symptom domain Attention

| Study | Slow progressing Total | Slow progressing Mean | Slow progressing SD | Fast progressing Total | Fast progressing Mean | Fast progressing SD | SMD | 95%−CI |
|---|---|---|---|---|---|---|---|---|
| **Cohort = PPMI** | | | | | | | | |
| Letter Number Sequencing | 56 | 0.03 | 0.1650 | 5 | −0.04 | 0.0202 | 0.40 | [−0.52; 1.31] |
| MOCA Attention | 320 | 1.33 | 0.7561 | 71 | 3.97 | 2.4125 | −2.15 | [−2.44; −1.85] |
| **Random effects model** | **376** | | | **76** | | | **−0.91** | **[−3.40; 1.58]** |
| **Cohort = ICEBERG** | | | | | | | | |
| MATTIS Attention | 104 | 0.00 | 0.0367 | 41 | −0.02 | 0.0090 | 0.71 | [ 0.34; 1.08] |
| MOCA Attention | 112 | 0.52 | 0.2299 | 42 | 0.94 | 0.5589 | −1.18 | [−1.56; −0.80] |
| NMSQ Attention | 111 | 2.20 | 0.0771 | 42 | 1.79 | 0.1678 | 3.71 | [ 3.16; 4.26] |
| **Random effects model** | **327** | | | **125** | | | **1.07** | **[−1.71; 3.86]** |
| **Cohort = LuxPARK** | | | | | | | | |
| MOCA Attention | 408 | 0.58 | 0.3055 | 153 | 1.52 | 0.5164 | −2.49 | [−2.73; −2.25] |
| NMSQ Attention | 408 | 1.47 | 1.1380 | 152 | 2.09 | 0.1286 | −0.63 | [−0.82; −0.44] |
| PDQ39 Attention | 408 | 1.34 | 0.2652 | 153 | 1.77 | 0.4815 | −1.29 | [−1.49; −1.09] |
| SIQCDE Attention | 353 | 0.49 | 0.3458 | 122 | 1.00 | 0.8884 | −0.94 | [−1.15; −0.72] |
| **Random effects model** | **1577** | | | **580** | | | **−1.34** | **[−2.13; −0.54]** |
| **Random effects model** | **2280** | | | **781** | | | **−0.44** | **[−1.66; 0.77]** |

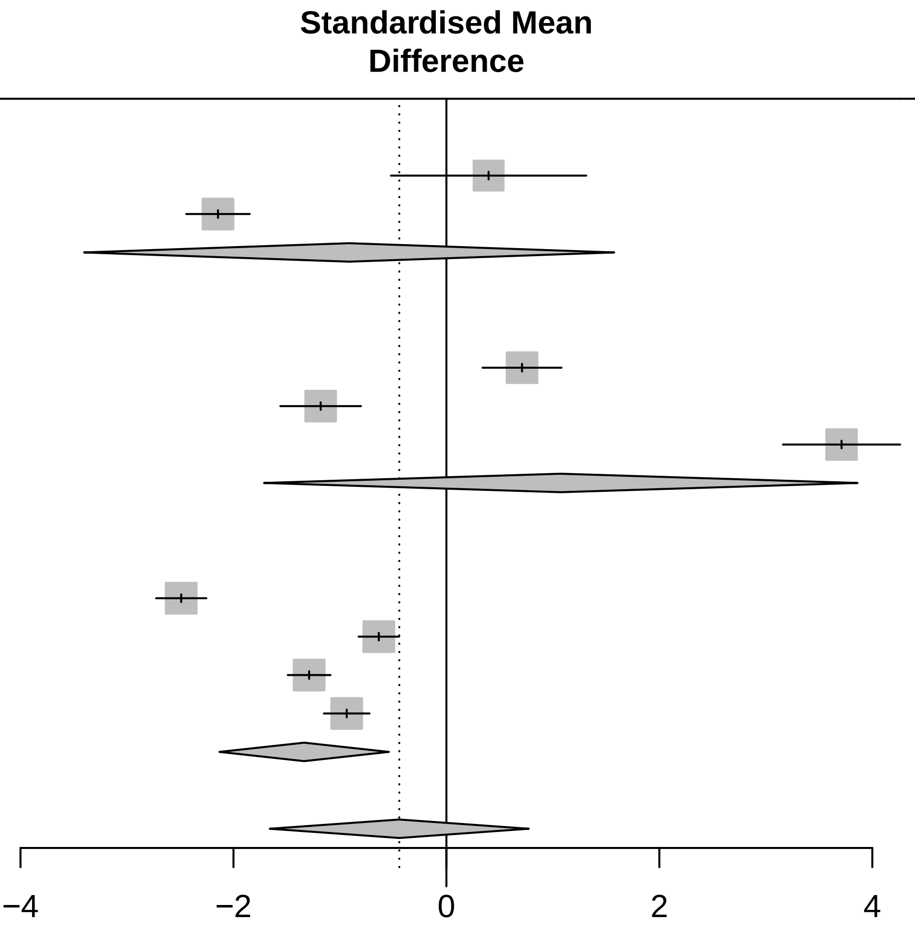

<− Faster progression for fast−progressing subtype | faster progression for slow−progressing subtype −>

Three−level metaanalysis using random effects to calculate an overall SMD estimate for Attention across cohorts.
The dashed line indicates the overall mean SMD estimate. The solid line indicates no effect.

### Forest plot for progression characteristics of symptom domain Non motor symptoms

| Study | Slow progressing Total | Slow progressing Mean | Slow progressing SD | Fast progressing Total | Fast progressing Mean | Fast progressing SD | SMD | 95%−CI |
|---|---|---|---|---|---|---|---|---|
| **Cohort = LuxPARK** | | | | | | | | |
| NMSQ | 402 | 0.11 | 0.0262 | 148 | 0.20 | 0.0522 | −2.75 | [−3.00; −2.50] |
| UPDRS I | 408 | 0.10 | 0.0268 | 153 | 0.22 | 0.0835 | −2.41 | [−2.64; −2.17] |
| **Random effects model** | **810** | | | **301** | | | **−2.58** | **[−2.91; −2.24]** |
| **Cohort = ICEBERG** | | | | | | | | |
| NMSQ | 111 | 0.20 | 0.0447 | 42 | 0.22 | 0.0016 | −0.50 | [−0.86; −0.14] |
| UPDRS I | 112 | 0.14 | 0.0182 | 42 | 0.19 | 0.0535 | −1.66 | [−2.06; −1.25] |
| **Random effects model** | **223** | | | **84** | | | **−1.07** | **[−2.21; 0.06]** |
| **Cohort = PPMI** | | | | | | | | |
| UPDRS I | 335 | 0.19 | 0.1170 | 74 | 0.55 | 0.1480 | −2.97 | [−3.30; −2.65] |
| **Random effects model** | **1368** | | | **459** | | | **−2.06** | **[−2.94; −1.19]** |

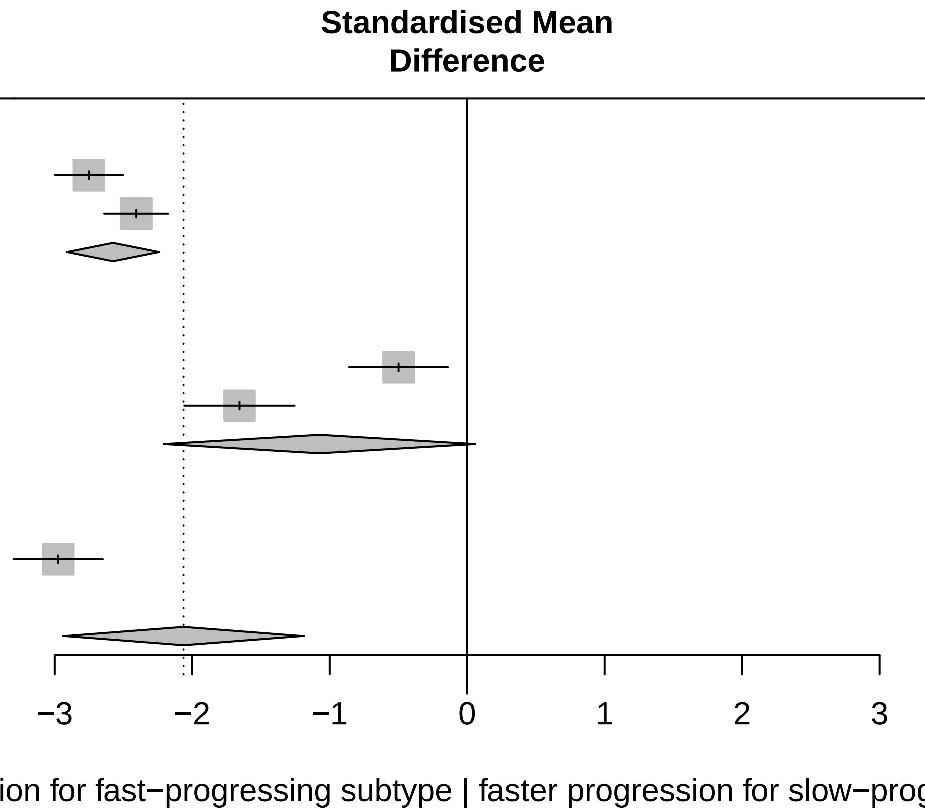

Three−level metaanalysis using random effects to calculate an overall SMD estimate for Non motor symptoms across cohorts. The dashed line indicates the overall mean SMD estimate. The solid line indicates no effect.

### Forest plot for progression characteristics of symptom domain Autonomic

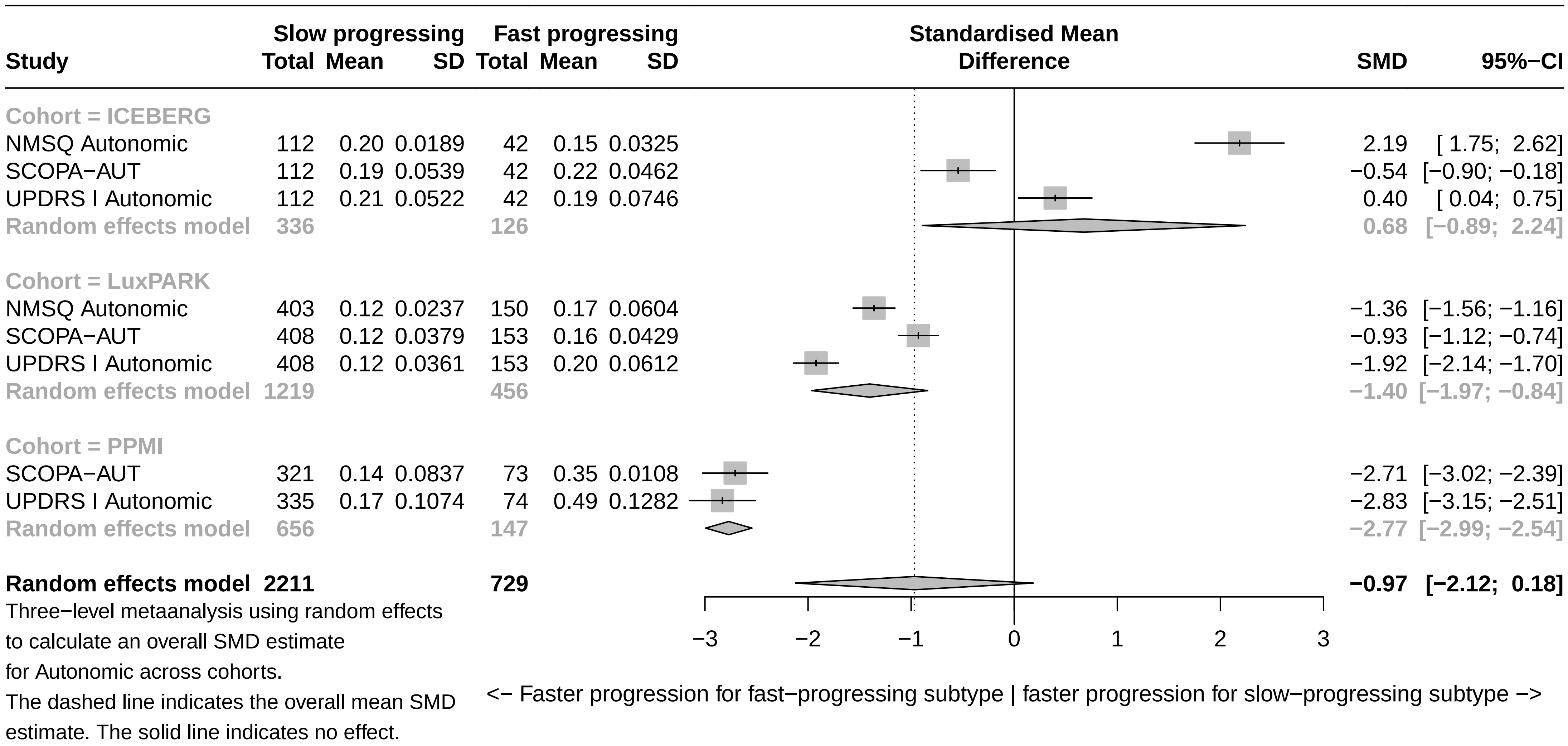

### Forest plot for progression characteristics of symptom domain Hallucinations

| Study | Slow progressing Total | Slow progressing Mean | Slow progressing SD | Fast progressing Total | Fast progressing Mean | Fast progressing SD | SMD | 95%−CI |
|---|---|---|---|---|---|---|---|---|
| **Cohort = ICEBERG** | | | | | | | | |
| NMSQ Hallucination | 112 | −2.07 | 1.7091 | 42 | 2.19 | 1.0470 | −2.72 | [−3.19; −2.25] |
| UPDRS I Hallucinations | 112 | 0.72 | 1.4359 | 42 | 4.74 | 6.1700 | −1.17 | [−1.55; −0.79] |
| **Random effects model** | **224** | | | **84** | | | **−1.94** | **[−3.46; −0.42]** |
| **Cohort = LuxPARK** | | | | | | | | |
| NMSQ Hallucination | 408 | 1.63 | 0.3010 | 152 | 1.79 | 0.6111 | −0.39 | [−0.58; −0.21] |
| UPDRS I Hallucinations | 408 | 1.23 | 0.2917 | 153 | 1.33 | 0.8760 | −0.19 | [−0.38; −0.00] |
| **Random effects model** | **816** | | | **305** | | | **−0.29** | **[−0.49; −0.09]** |
| **Cohort = PPMI** | | | | | | | | |
| UPDRS I Hallucinations | 335 | 3.04 | 0.7427 | 74 | 6.04 | 4.4997 | −1.48 | [−1.76; −1.21] |
| **Random effects model** | **1375** | | | **463** | | | **−1.18** | **[−2.05; −0.30]** |

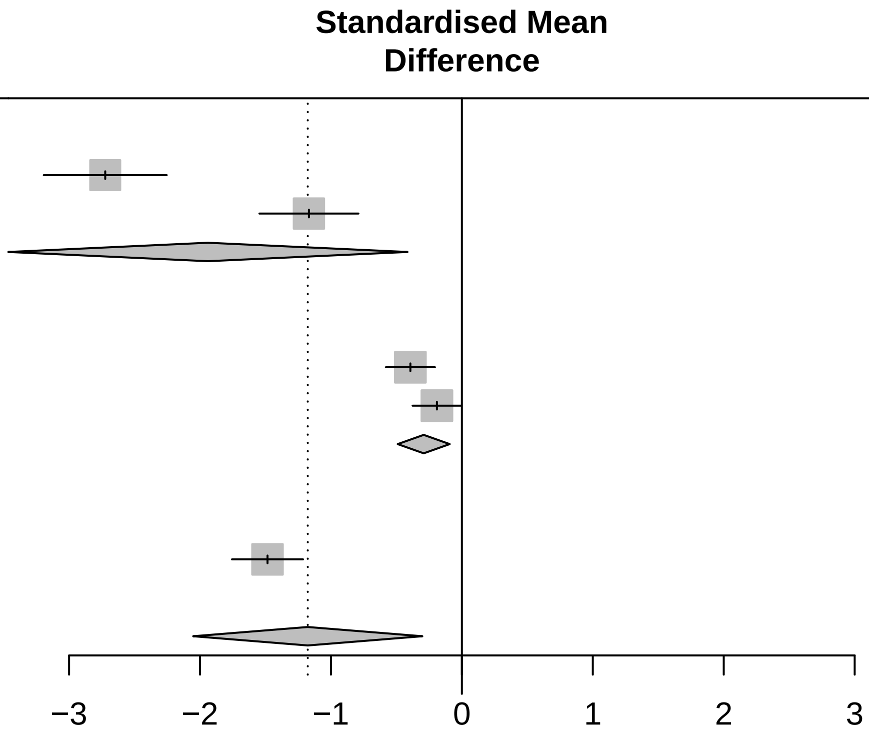

<− Faster progression for fast−progressing subtype | faster progression for slow−progressing subtype −>

Three−level metaanalysis using random effects to calculate an overall SMD estimate for Hallucinations across cohorts.
The dashed line indicates the overall mean SMD estimate. The solid line indicates no effect.

### Forest plot for progression characteristics of symptom domain Pain

| Study | Slow progressing Total | Slow progressing Mean | Slow progressing SD | Fast progressing Total | Fast progressing Mean | Fast progressing SD | SMD | 95%−CI |
|---|---|---|---|---|---|---|---|---|
| **Cohort = ICEBERG** | | | | | | | | |
| NMSQ Pain | 111 | 0.75 | 0.0044 | 42 | 0.59 | 0.6328 | 0.48 | [ 0.12; 0.84] |
| UPDRS I Pain | 112 | 1.04 | 0.1276 | 42 | 0.88 | 0.0334 | 1.44 | [ 1.05; 1.83] |
| **Random effects model** | **223** | | | **84** | | | **0.96** | **[ 0.01; 1.90]** |
| **Cohort = LuxPARK** | | | | | | | | |
| NMSQ Pain | 408 | 0.88 | 0.1017 | 152 | 0.84 | 0.1111 | 0.40 | [ 0.22; 0.59] |
| PDQ39 Pain | 407 | 0.10 | 0.0208 | 153 | 0.14 | 0.0468 | −1.10 | [−1.30; −0.90] |
| UPDRS I Pain | 408 | 1.07 | 0.1576 | 153 | 1.69 | 0.7281 | −1.55 | [−1.75; −1.34] |
| **Random effects model** | **1223** | | | **458** | | | **−0.75** | **[−1.90; 0.41]** |
| **Cohort = PPMI** | | | | | | | | |
| UPDRS I Pain | 335 | 1.88 | 1.6805 | 74 | 4.36 | 2.1147 | −1.40 | [−1.67; −1.13] |
| **Random effects model** | **1781** | | | **616** | | | **−0.29** | **[−1.27; 0.68]** |

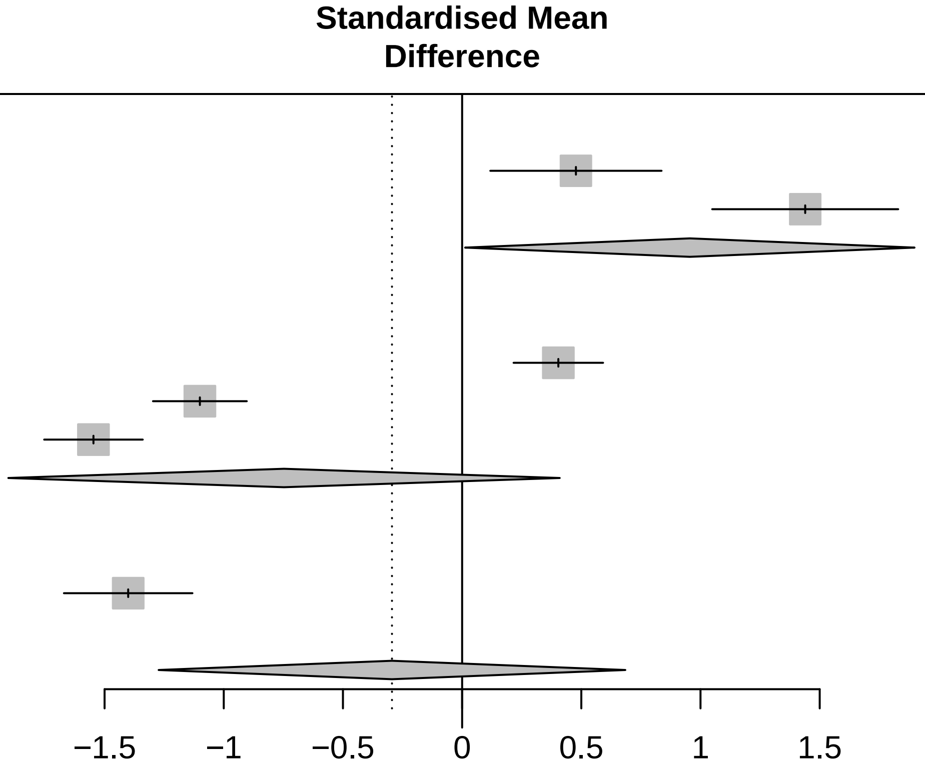

<− Faster progression for fast−progressing subtype | faster progression for slow−progressing subtype −>

Three−level metaanalysis using random effects to calculate an overall SMD estimate for Pain across cohorts.
The dashed line indicates the overall mean SMD estimate. The solid line indicates no effect.

### Forest plot for progression characteristics of symptom domain RBD

| Study | Slow progressing Total | Slow progressing Mean | Slow progressing SD | Fast progressing Total | Fast progressing Mean | Fast progressing SD | SMD | 95%−CI |
|---|---|---|---|---|---|---|---|---|
| **Cohort = ICEBERG** | | | | | | | | |
| NMSQ RBD | 112 | 1.73 | 0.3193 | 42 | 0.72 | 1.1740 | 1.50 | [ 1.11; 1.89] |
| RBD−HK | 111 | 0.07 | 0.0573 | 41 | 0.03 | 0.1471 | 0.37 | [ 0.01; 0.73] |
| **Random effects model** | **223** | | | **83** | | | **0.93** | **[−0.18; 2.04]** |
| **Cohort = LuxPARK** | | | | | | | | |
| NMSQ RBD | 407 | 1.58 | 0.3502 | 152 | 1.11 | 0.3609 | 1.33 | [ 1.12; 1.53] |
| RBD−SQ | 405 | 0.10 | 0.0627 | 152 | 0.07 | 0.0916 | 0.40 | [ 0.21; 0.59] |
| **Random effects model** | **812** | | | **304** | | | **0.86** | **[−0.04; 1.77]** |
| **Cohort = PPMI** | | | | | | | | |
| RBD−SQ | 324 | 0.18 | 0.2022 | 72 | 0.18 | 0.1810 | 0.03 | [−0.23; 0.28] |
| **Random effects model** | **1359** | | | **459** | | | **0.72** | **[ 0.15; 1.28]** |

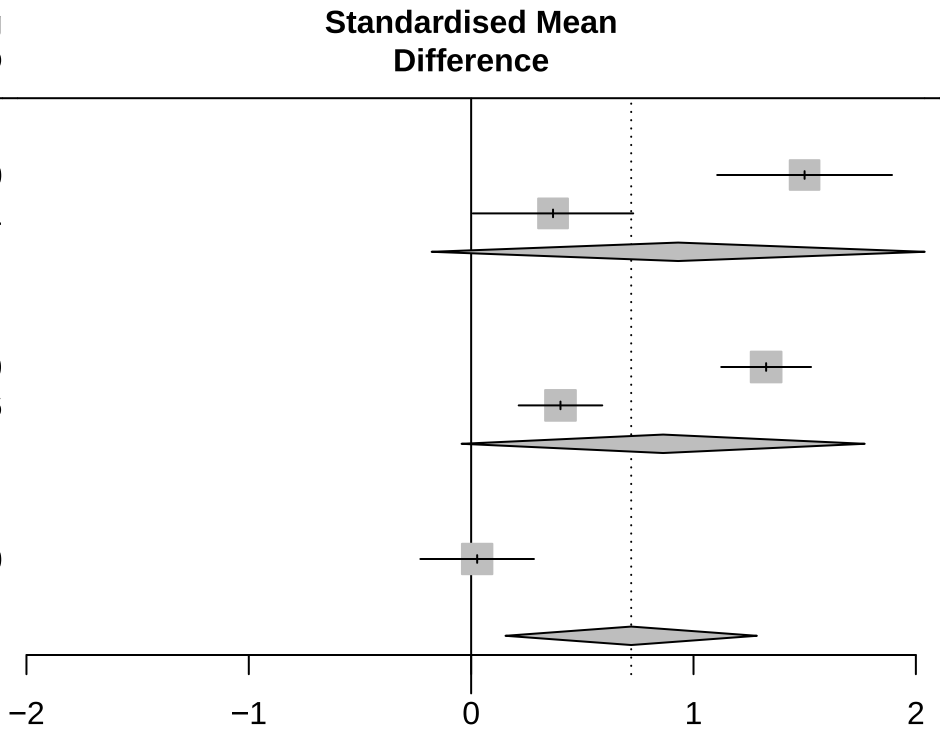

Three−level metaanalysis using random effects to calculate an overall SMD estimate for RBD across cohorts.
The dashed line indicates the overall mean SMD estimate. The solid line indicates no effect.

### Forest plot for progression characteristics of symptom domain Smell

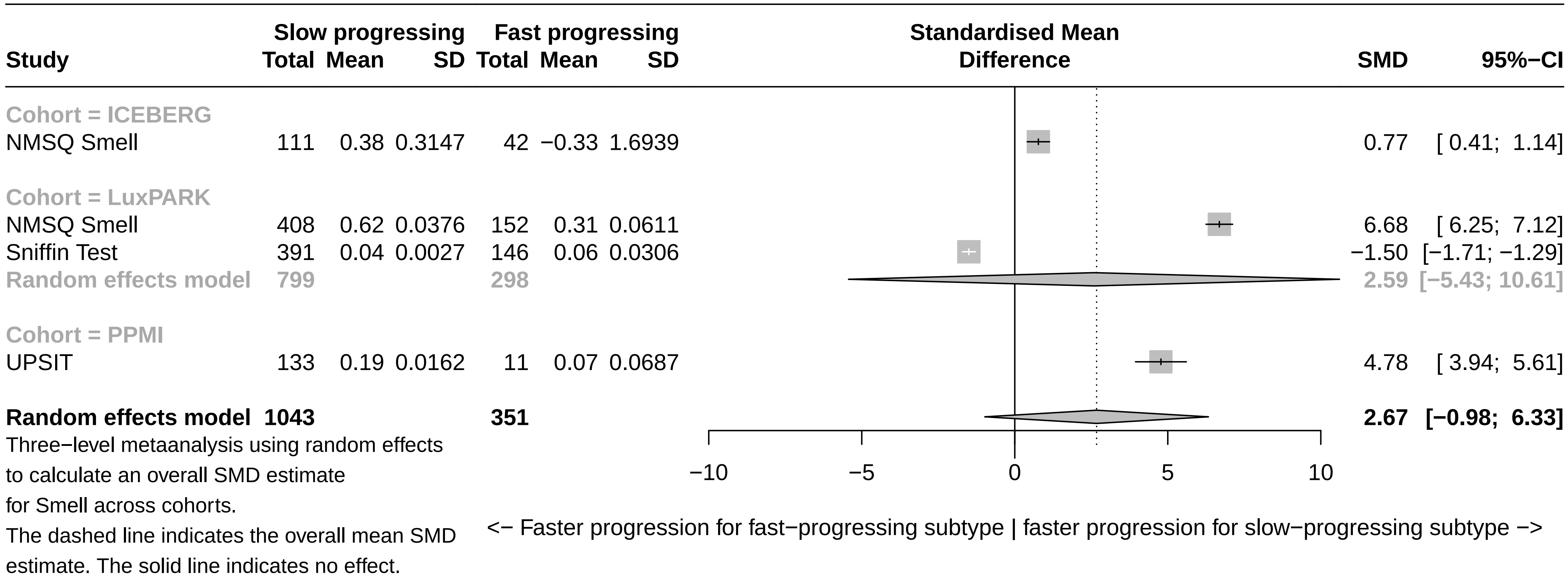

### Forest plot for progression characteristics of symptom domain Motor symptoms

| Study | Slow progressing Total | Slow progressing Mean | Slow progressing SD | Fast progressing Total | Fast progressing Mean | Fast progressing SD | SMD | 95%−CI |
|---|---|---|---|---|---|---|---|---|
| **Cohort = LuxPARK** | | | | | | | | |
| PDQ39 ADL | 408 | 0.15 | 0.0652 | 152 | 0.36 | 0.1909 | −1.85 | [−2.06; −1.63] |
| Pegboard | 386 | 0.06 | 0.0171 | 128 | 0.12 | 0.0374 | −2.43 | [−2.68; −2.18] |
| UPDRS II | 408 | 0.15 | 0.0555 | 153 | 0.42 | 0.1606 | −2.75 | [−2.99; −2.50] |
| UPDRS III on | 380 | 0.09 | 0.0212 | 139 | 0.23 | 0.0707 | −3.45 | [−3.74; −3.17] |
| UPDRS IV | 408 | 0.09 | 0.0759 | 153 | 0.11 | 0.0872 | −0.18 | [−0.36; 0.01] |
| **Random effects model** | **1990** | | | **725** | | | **−2.13** | **[−3.21; −1.04]** |
| **Cohort = PPMI** | | | | | | | | |
| UPDRS II | 335 | 0.17 | 0.1047 | 74 | 0.59 | 0.1604 | −3.57 | [−3.92; −3.21] |
| UPDRS III off | 335 | 0.26 | 0.1491 | 74 | 0.58 | 0.1757 | −2.02 | [−2.30; −1.73] |
| UPDRS III on | 300 | 0.03 | 0.1291 | 66 | 0.31 | 0.2480 | −1.77 | [−2.06; −1.47] |
| UPDRS III untreated | 335 | 0.55 | 0.1221 | 74 | 0.61 | 0.3608 | −0.36 | [−0.62; −0.11] |
| UPDRS IV | 301 | 0.26 | 0.1800 | 68 | 0.37 | 0.2256 | −0.59 | [−0.86; −0.33] |
| **Random effects model** | **1606** | | | **356** | | | **−1.66** | **[−2.78; −0.53]** |
| **Cohort = ICEBERG** | | | | | | | | |
| UPDRS II | 112 | 0.20 | 0.0313 | 42 | 0.39 | 0.0694 | −4.20 | [−4.79; −3.61] |
| UPDRS III off | 111 | 0.04 | 0.0082 | 42 | 0.16 | 0.0983 | −2.23 | [−2.66; −1.79] |
| UPDRS III on | 105 | 0.01 | 0.0023 | 41 | 0.09 | 0.0623 | −2.55 | [−3.02; −2.09] |
| UPDRS IV | 112 | 0.07 | 0.1174 | 42 | 0.16 | 0.1752 | −0.64 | [−1.00; −0.27] |
| **Random effects model** | **440** | | | **167** | | | **−2.39** | **[−3.82; −0.96]** |
| **Random effects model** | **4036** | | | **1248** | | | **−2.03** | **[−2.69; −1.38]** |

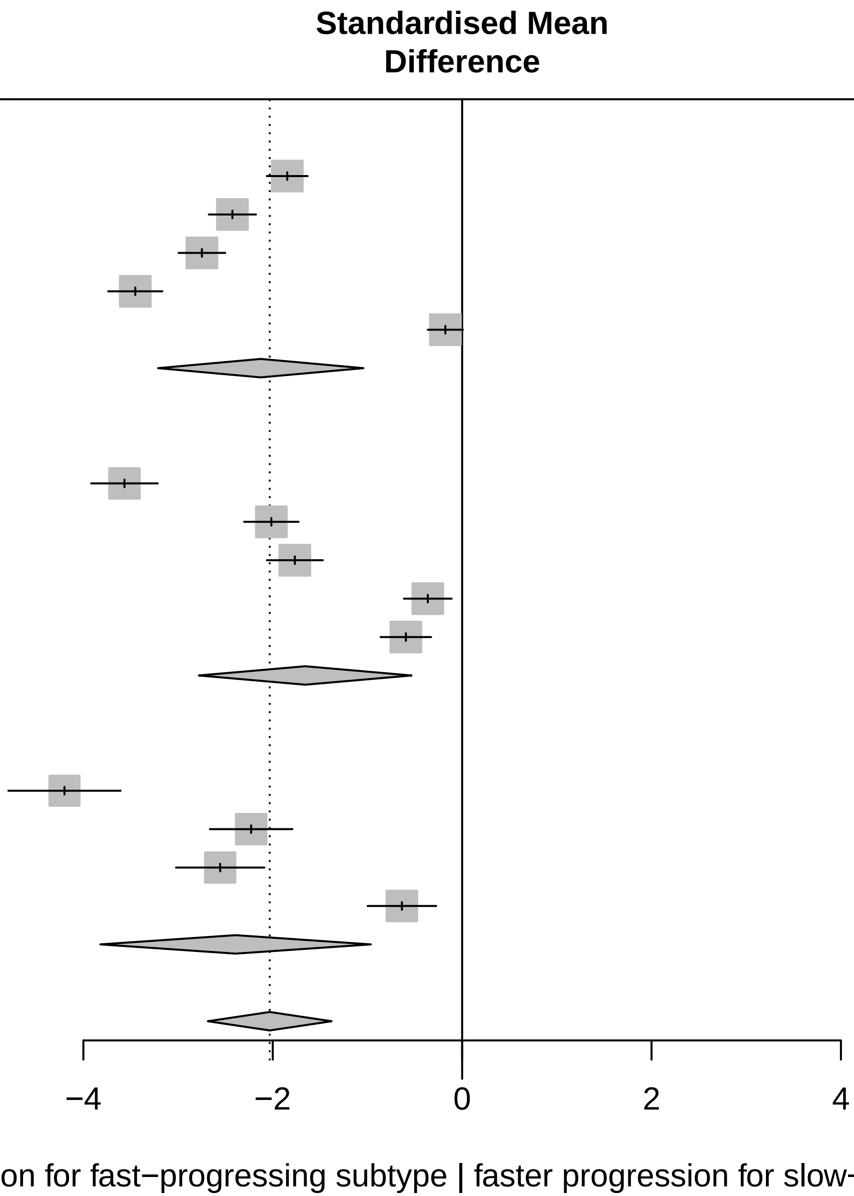

Three−level metaanalysis using random effects
to calculate an overall SMD estimate
for Motor symptoms across cohorts.
The dashed line indicates the overall mean SMD
estimate. The solid line indicates no effect.

### Forest plot for progression characteristics of symptom domain Impulsivity

| Study | Slow progressing Total | Slow progressing Mean | Slow progressing SD | Fast progressing Total | Fast progressing Mean | Fast progressing SD | SMD | 95%−CI |
|---|---|---|---|---|---|---|---|---|
| **Cohort = PPMI** | | | | | | | | |
| QUIP | 324 | 0.04 | 0.0547 | 72 | 0.07 | 0.1027 | −0.41 | [−0.67; −0.16] |
| **Cohort = ICEBERG** | | | | | | | | |
| QUIP−RS | 112 | 0.04 | 0.0690 | 42 | 0.09 | 0.0820 | −0.67 | [−1.03; −0.31] |
| **Random effects model** | **436** | | | **114** | | | **−0.51** | **[−0.75; −0.27]** |

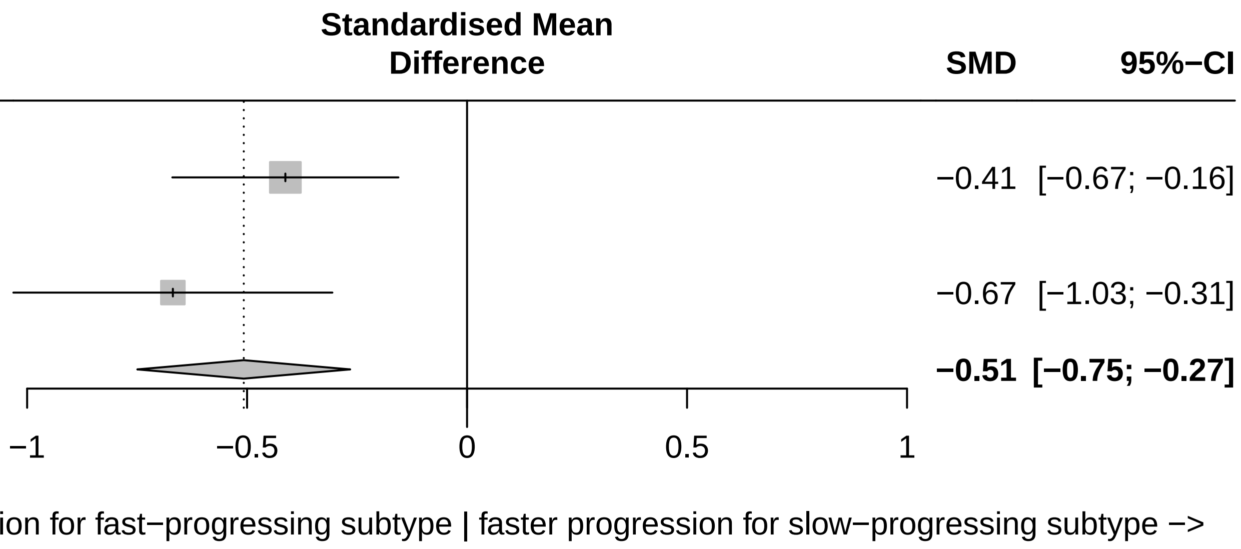

Three−level metaanalysis using random effects
to calculate an overall SMD estimate
for Impulsivity across cohorts.
The dashed line indicates the overall mean SMD
estimate. The solid line indicates no effect.

### Forest plots for symptom domain progression (cross-cohort validation)

### Forest plot for progression characteristics of symptom domain Axial & PIGD (validation)

| Study | Slow progressing Total | Slow progressing Mean | Slow progressing SD | Fast progressing Total | Fast progressing Mean | Fast progressing SD | SMD | 95%−CI |
|---|---|---|---|---|---|---|---|---|
| **Cohort = ICEBERG (validation)** | | | | | | | | |
| UPDRS III axial off | 134 | 0.12 | 0.0154 | 19 | 0.54 | 0.2700 | −4.47 | [−5.16; −3.77] |
| UPDRS III axial on | 127 | 0.13 | 0.0067 | 19 | 0.16 | 0.1915 | −0.53 | [−1.01; −0.04] |
| GABS Examination | 135 | 0.05 | 0.0213 | 19 | 0.27 | 0.2062 | −3.01 | [−3.60; −2.42] |
| GABS Questionaire | 135 | 0.04 | 0.0637 | 19 | 0.32 | 0.2745 | −2.49 | [−3.04; −1.93] |
| PIGD off | 134 | 0.09 | 0.0457 | 19 | 0.37 | 0.1549 | −4.13 | [−4.80; −3.46] |
| PIGD on | 127 | 0.09 | 0.0434 | 19 | 0.23 | 0.1583 | −1.97 | [−2.50; −1.44] |
| **Random effects model** | **792** | | | **114** | | | **−2.75** | **[−3.91; −1.59]** |
| **Cohort = LuxPARK (validation)** | | | | | | | | |
| UPDRS III axial on | 430 | 0.15 | 0.0629 | 90 | 0.34 | 0.1203 | −2.59 | [−2.87; −2.31] |
| FOGAC | 70 | 0.09 | 0.0702 | 10 | 0.15 | 0.3365 | −0.43 | [−1.10; 0.23] |
| FOGQ | 188 | 0.18 | 0.1030 | 42 | 0.43 | 0.3730 | −1.36 | [−1.72; −1.01] |
| PDQ39 Mobility | 461 | 0.21 | 0.1023 | 98 | 0.51 | 0.2099 | −2.40 | [−2.66; −2.14] |
| PIGD on | 430 | 0.15 | 0.0996 | 90 | 0.39 | 0.1639 | −2.06 | [−2.32; −1.80] |
| TuG | 172 | 0.03 | 0.0320 | 24 | 0.07 | 0.1145 | −0.76 | [−1.19; −0.32] |
| **Random effects model** | **1751** | | | **354** | | | **−1.63** | **[−2.33; −0.93]** |
| **Random effects model** | **2543** | | | **468** | | | **−2.17** | **[−2.89; −1.45]** |

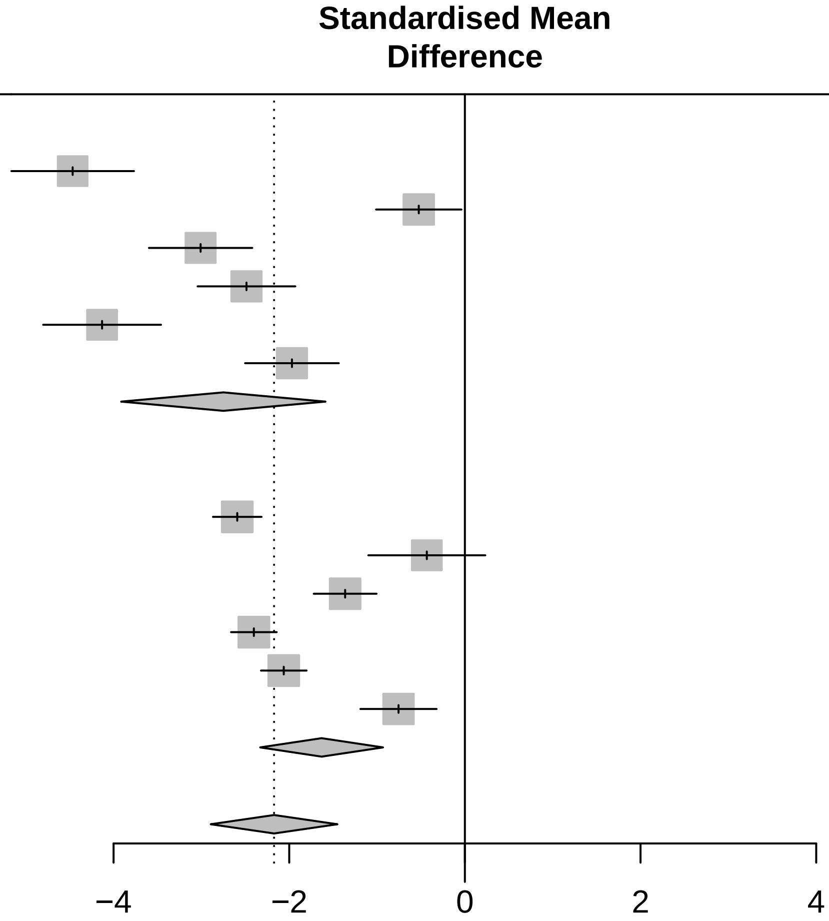

Three−level metaanalysis using random effects to calculate an overall SMD estimate for Axial & PIGD across cohorts. The dashed line indicates the overall mean SMD estimate. The solid line indicates no effect.

<− Faster progression for fast−progressing subtype | faster progression for slow−progressing subtype −>

### Forest plot for progression characteristics of symptom domain Depression (validation)

| Study | Slow progressing Total | Slow progressing Mean | Slow progressing SD | Fast progressing Total | Fast progressing Mean | Fast progressing SD | SMD | 95%−CI |
|---|---|---|---|---|---|---|---|---|
| **Cohort = LuxPARK (validation)** | | | | | | | | |
| BDI | 459 | 0.06 | 0.0305 | 96 | 0.14 | 0.0120 | −2.80 | [−3.07; −2.52] |
| NMSQ Depression | 462 | 1.19 | 0.4669 | 98 | 2.22 | 0.2138 | −2.37 | [−2.63; −2.11] |
| PDQ39 Depression | 461 | 0.08 | 0.0500 | 98 | 0.16 | 0.0658 | −1.60 | [−1.84; −1.36] |
| UPDRS I Depression | 462 | 0.92 | 0.2209 | 99 | 1.83 | 0.7230 | −2.51 | [−2.77; −2.25] |
| **Random effects model** | **1844** | | | **391** | | | **−2.32** | **[−2.82; −1.81]** |
| **Cohort = ICEBERG (validation)** | | | | | | | | |
| HADS depression | 135 | 0.11 | 0.0026 | 19 | 0.17 | 0.0615 | −2.86 | [−3.44; −2.28] |
| NMSQ Depression | 135 | 1.83 | 0.2900 | 19 | 1.59 | 2.8964 | 0.22 | [−0.26; 0.71] |
| UPDRS I Depression | 135 | 1.32 | 0.7801 | 19 | 1.64 | 0.6916 | −0.42 | [−0.91; 0.06] |
| **Random effects model** | **405** | | | **57** | | | **−1.01** | **[−2.85; 0.82]** |
| **Random effects model** | **2249** | | | **448** | | | **−1.77** | **[−2.67; −0.87]** |

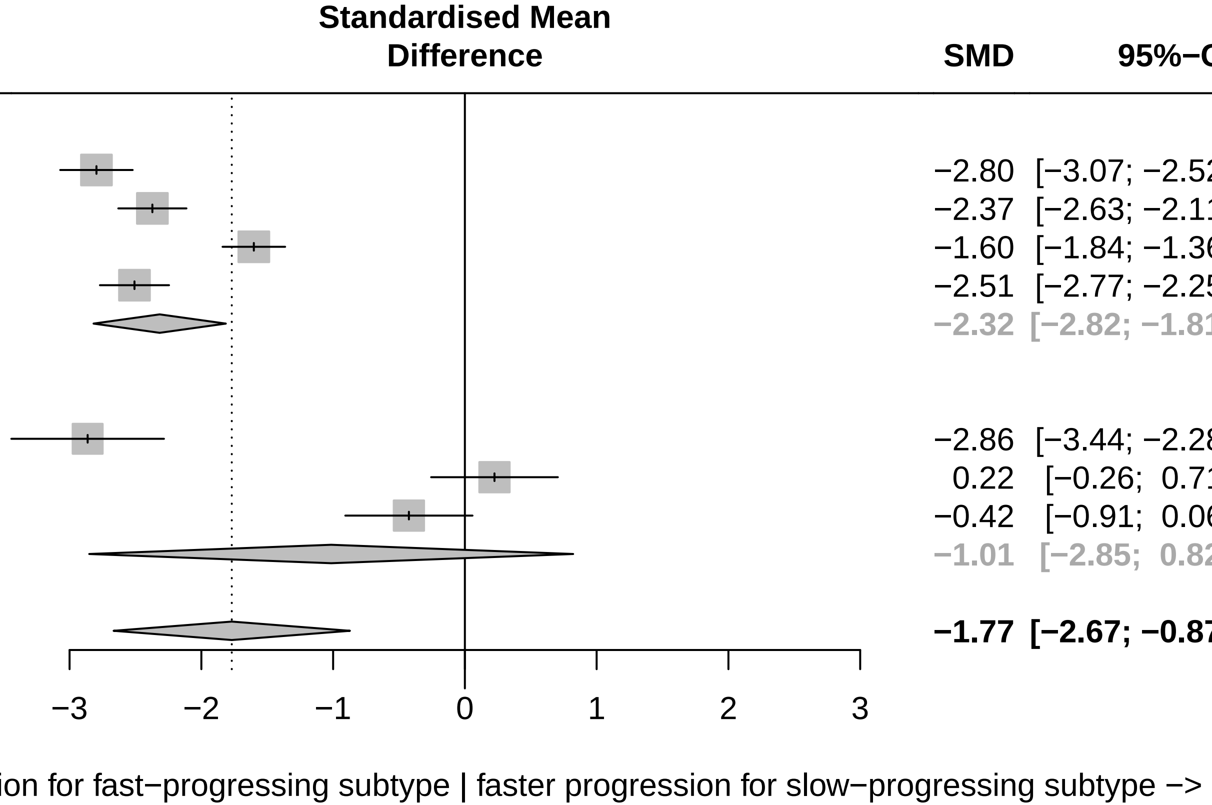

Three−level metaanalysis using random effects to calculate an overall SMD estimate for Depression across cohorts.
The dashed line indicates the overall mean SMD estimate. The solid line indicates no effect.

### Forest plot for progression characteristics of symptom domain Overall severity (validation)

| Study | Slow progressing Total | Slow progressing Mean | Slow progressing SD | Fast progressing Total | Fast progressing Mean | Fast progressing SD | SMD | 95%−CI |
|---|---|---|---|---|---|---|---|---|
| **Cohort = ICEBERG (validation)** | | | | | | | | |
| CGIS | 135 | 1.40 | 0.2906 | 19 | 2.38 | 1.5224 | −1.65 | [ −2.16; −1.13] |
| H&Y | 135 | −0.47 | 0.0485 | 19 | 0.81 | 0.6415 | −5.68 | [ −6.49; −4.88] |
| SEADL | 135 | 0.06 | 0.0267 | 19 | 0.20 | 0.1218 | −2.84 | [ −3.42; −2.26] |
| UPDRS I−III off | 134 | 0.17 | 0.0261 | 19 | 0.61 | 0.1236 | −8.93 | [−10.05; −7.81] |
| UPDRS I−III on | 127 | 0.14 | 0.0236 | 19 | 0.43 | 0.0561 | −9.86 | [−11.11; −8.62] |
| Random effects model | 666 | | | 95 | | | −5.76 | [ −8.93; −2.60] |
| **Cohort = LuxPARK (validation)** | | | | | | | | |
| FAQ | 399 | 0.11 | 0.0810 | 79 | 0.33 | 0.1342 | −2.39 | [ −2.68; −2.11] |
| H&Y | 462 | 2.74 | 0.5202 | 99 | 4.83 | 1.2833 | −2.92 | [ −3.20; −2.64] |
| PDQ39 | 456 | 0.12 | 0.0548 | 96 | 0.32 | 0.1556 | −2.38 | [ −2.64; −2.12] |
| UPDRS I−III on | 426 | 0.16 | 0.0502 | 89 | 0.40 | 0.1484 | −3.14 | [ −3.44; −2.84] |
| Random effects model | 1743 | | | 363 | | | −2.70 | [ −3.08; −2.33] |
| **Random effects model** | **2409** | | | **458** | | | **−4.38** | **[ −6.33; −2.43]** |

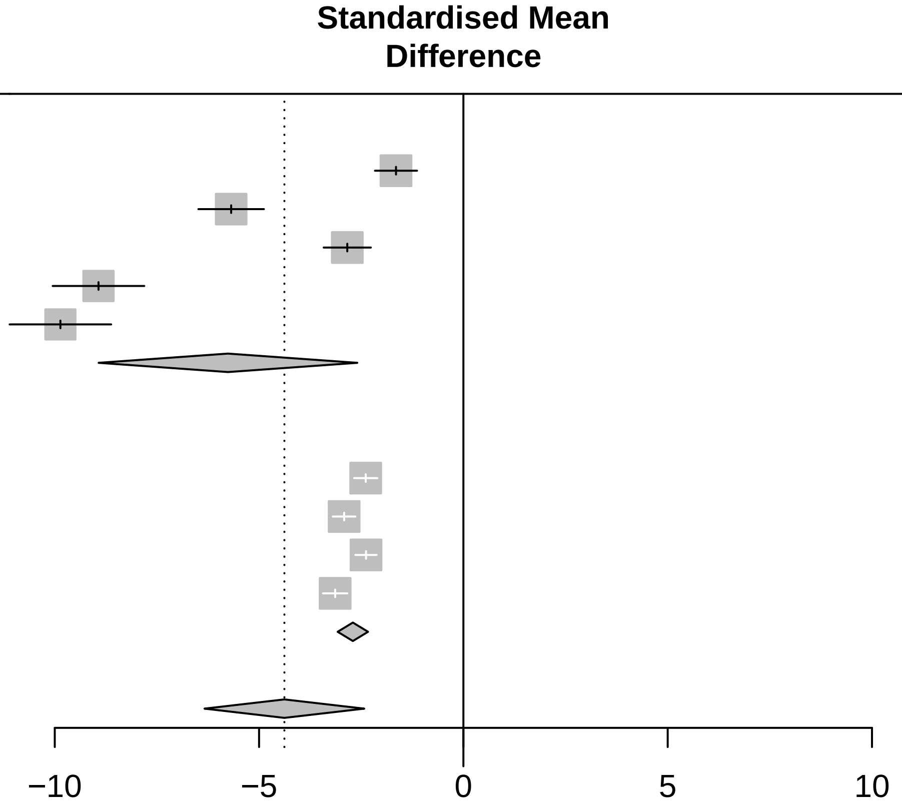

<− Faster progression for fast−progressing subtype | faster progression for slow−progressing subtype −>

Three−level metaanalysis using random effects to calculate an overall SMD estimate for Overall severity across cohorts. The dashed line indicates the overall mean SMD estimate. The solid line indicates no effect.

### Forest plot for progression characteristics of symptom domain Apathy (validation)

| Study | Slow progressing Total | Slow progressing Mean | Slow progressing SD | Fast progressing Total | Fast progressing Mean | Fast progressing SD | SMD | 95%−CI |
|---|---|---|---|---|---|---|---|---|
| **Cohort = ICEBERG (validation)** | | | | | | | | |
| DAS | 124 | 0.09 | 0.0128 | 17 | 0.09 | 0.0490 | 0.22 | [−0.28; 0.73] |
| SAS | 127 | 0.09 | 0.0224 | 18 | 0.09 | 0.0462 | 0.07 | [−0.42; 0.56] |
| UPDRS I Apathy | 135 | 1.11 | 0.1876 | 19 | 1.17 | 1.1351 | −0.14 | [−0.62; 0.34] |
| **Random effects model** | **386** | | | **54** | | | **0.05** | **[−0.24; 0.33]** |
| **Cohort = LuxPARK (validation)** | | | | | | | | |
| SAS | 459 | 0.05 | 0.0433 | 99 | 0.14 | 0.0457 | −2.05 | [−2.30; −1.80] |
| UPDRS I Apathy | 462 | 0.88 | 0.0486 | 99 | 2.08 | 0.9082 | −3.14 | [−3.42; −2.85] |
| **Random effects model** | **921** | | | **198** | | | **−2.59** | **[−3.66; −1.53]** |
| **Random effects model** | **1307** | | | **252** | | | **−1.02** | **[−2.34; 0.31]** |

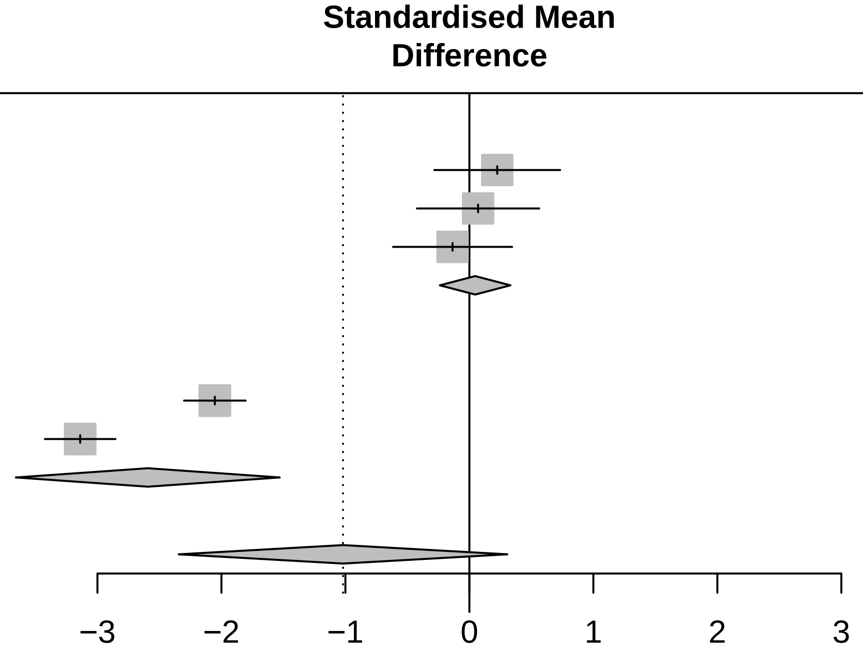

<− Faster progression for fast−progressing subtype | faster progression for slow−progressing subtype −>

Three−level metaanalysis using random effects to calculate an overall SMD estimate for Apathy across cohorts.
The dashed line indicates the overall mean SMD estimate. The solid line indicates no effect.

### Forest plot for progression characteristics of symptom domain Sleep (validation)

| Study | Slow progressing Total | Slow progressing Mean | Slow progressing SD | Fast progressing Total | Fast progressing Mean | Fast progressing SD | SMD | 95%−CI |
|---|---|---|---|---|---|---|---|---|
| **Cohort = ICEBERG (validation)** | | | | | | | | |
| ESS | 133 | 0.17 | 0.1063 | 19 | 0.16 | 0.0548 | 0.09 | [−0.39; 0.58] |
| NMSQ Sleep | 135 | 1.21 | 0.2211 | 19 | 3.35 | 1.1076 | −4.89 | [−5.62; −4.16] |
| UPDRS I Sleep | 135 | 0.08 | 0.0224 | 19 | 0.15 | 0.0255 | −3.05 | [−3.64; −2.45] |
| **Random effects model** | **403** | | | **57** | | | **−2.60** | **[−5.46; 0.25]** |
| **Cohort = LuxPARK (validation)** | | | | | | | | |
| NMSQ Sleep | 462 | 0.68 | 0.0575 | 98 | 0.76 | 0.3560 | −0.53 | [−0.75; −0.31] |
| PDSS | 460 | 0.11 | 0.0237 | 98 | 0.12 | 0.0355 | −0.51 | [−0.73; −0.29] |
| UPDRS I Sleep | 462 | 0.10 | 0.0325 | 99 | 0.12 | 0.0353 | −0.64 | [−0.86; −0.42] |
| **Random effects model** | **1384** | | | **295** | | | **−0.56** | **[−0.69; −0.43]** |
| **Random effects model** | **1787** | | | **352** | | | **−1.57** | **[−3.11; −0.03]** |

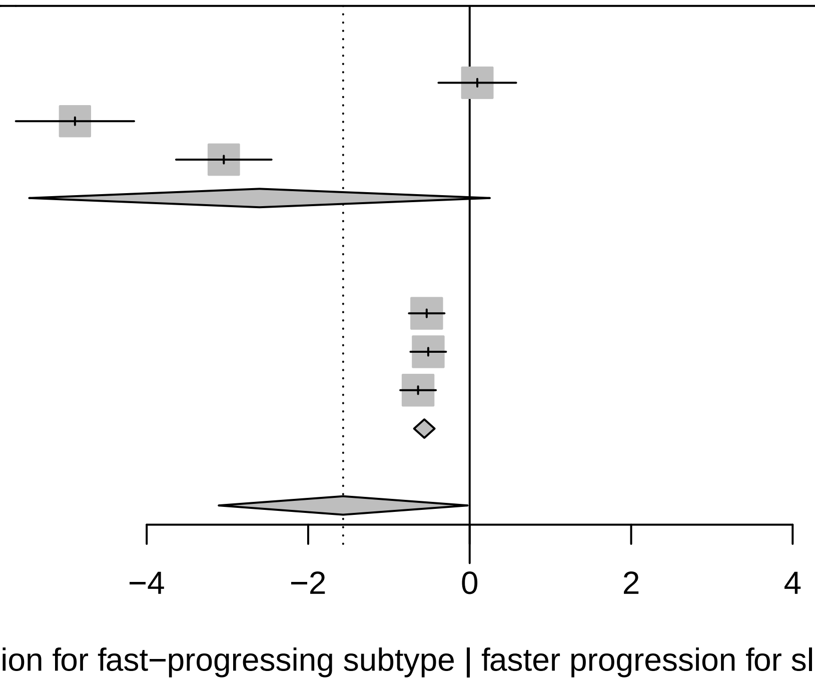

Three−level metaanalysis using random effects to calculate an overall SMD estimate for Sleep across cohorts.
The dashed line indicates the overall mean SMD estimate. The solid line indicates no effect.

### Forest plot for progression characteristics of symptom domain Overall cognition (validation)

| Study | Slow progressing Total | Slow progressing Mean | Slow progressing SD | Fast progressing Total | Fast progressing Mean | Fast progressing SD | SMD | 95%−CI |
|---|---|---|---|---|---|---|---|---|
| **Cohort = LuxPARK (validation)** | | | | | | | | |
| FAB | 48 | 0.03 | 0.0435 | 9 | 0.23 | 0.2836 | −1.69 | [−2.47; −0.91] |
| MoCA | 462 | 0.04 | 0.0278 | 99 | 0.21 | 0.1602 | −2.31 | [−2.57; −2.06] |
| NMSQ Cognition | 462 | 1.36 | 0.5848 | 97 | 3.17 | 1.3498 | −2.34 | [−2.60; −2.08] |
| PDQ39 Cognition | 461 | 0.08 | 0.0310 | 99 | 0.18 | 0.1010 | −1.95 | [−2.19; −1.70] |
| SIQCDE | 396 | 0.04 | 0.0408 | 77 | 0.10 | 0.0812 | −1.16 | [−1.42; −0.91] |
| UPDRS I Cognition | 462 | 1.21 | 0.1909 | 99 | 3.13 | 1.2061 | −3.58 | [−3.88; −3.28] |
| **Random effects model** | **2291** | | | **480** | | | **−2.18** | **[−2.85; −1.52]** |
| **Cohort = ICEBERG (validation)** | | | | | | | | |
| FAB | 127 | 0.05 | 0.0383 | 18 | 0.01 | 0.0697 | 1.03 | [ 0.53; 1.54] |
| MATTIS | 127 | 0.04 | 0.0346 | 18 | 0.07 | 0.0661 | −0.71 | [−1.22; −0.21] |
| MoCA | 135 | 0.05 | 0.0503 | 19 | 0.15 | 0.0817 | −1.80 | [−2.33; −1.28] |
| NMSQ Cognition | 135 | 1.85 | 0.4908 | 19 | 1.30 | 1.3777 | 0.83 | [ 0.34; 1.32] |
| UPDRS I Cognition | 135 | 0.87 | 0.8917 | 19 | 1.09 | 1.5180 | −0.22 | [−0.70; 0.26] |
| **Random effects model** | **659** | | | **93** | | | **−0.17** | **[−1.19; 0.85]** |
| **Random effects model** | **2950** | | | **573** | | | **−1.27** | **[−2.10; −0.44]** |

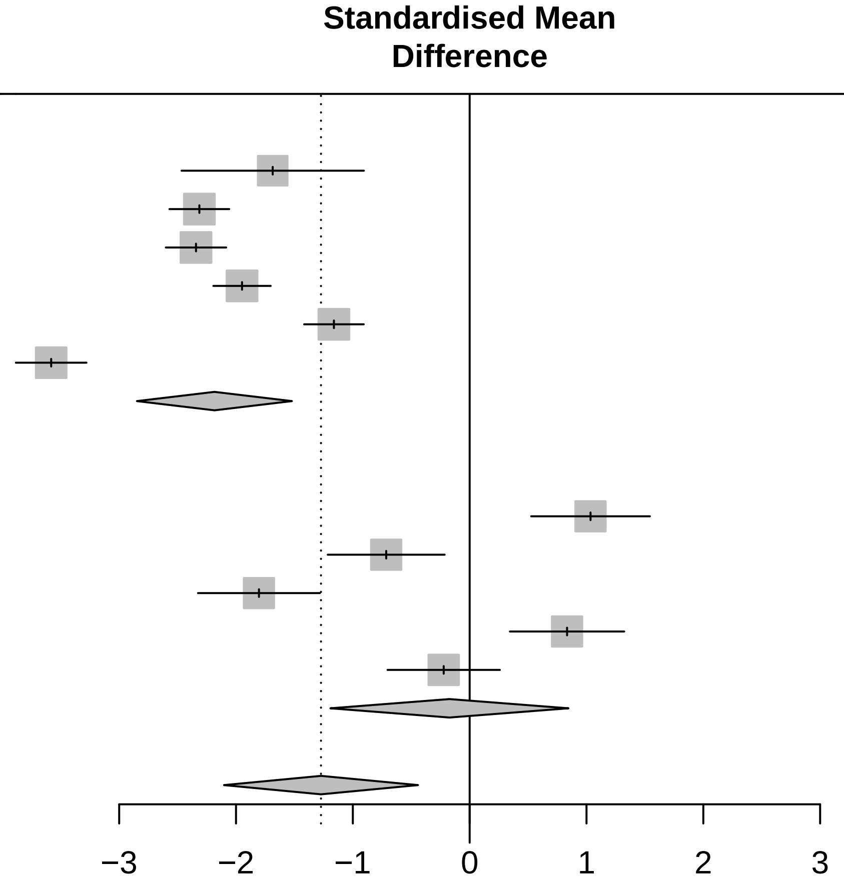

Three−level metaanalysis using random effects to calculate an overall SMD estimate for Overall cognition across cohorts. The dashed line indicates the overall mean SMD estimate. The solid line indicates no effect.

<− Faster progression for fast−progressing subtype | faster progression for slow−progressing subtype −>

### Forest plot for progression characteristics of symptom domain Conceptualization (validation)

| Study | Slow progressing Total | Slow progressing Mean | Slow progressing SD | Fast progressing Total | Fast progressing Mean | Fast progressing SD | SMD | 95%−CI |
|---|---|---|---|---|---|---|---|---|
| **Cohort = ICEBERG (validation)** | | | | | | | | |
| FAB Conceptualization | 127 | 2.45 | 1.5170 | 18 | 0.60 | 0.2456 | 1.29 | [ 0.78; 1.81] |
| MATTIS Conzeptualization | 127 | 0.01 | 0.0596 | 18 | 0.07 | 0.0491 | −1.17 | [−1.68; −0.65] |
| MOCA Abstraction | 135 | 1.01 | 0.9387 | 19 | 0.99 | 0.2560 | 0.02 | [−0.46; 0.50] |
| **Random effects model** | **389** | | | **55** | | | **0.05** | **[−1.34; 1.44]** |
| **Cohort = LuxPARK (validation)** | | | | | | | | |
| FAB Conceptualization | 49 | −1.64 | 0.9250 | 10 | −0.35 | 2.5093 | −0.97 | [−1.67; −0.26] |
| MOCA Abstraction | 462 | 0.17 | 0.2063 | 99 | 0.41 | 0.4000 | −0.93 | [−1.16; −0.71] |
| **Random effects model** | **511** | | | **109** | | | **−0.94** | **[−1.15; −0.72]** |
| **Random effects model** | **900** | | | **164** | | | **−0.35** | **[−1.25; 0.55]** |

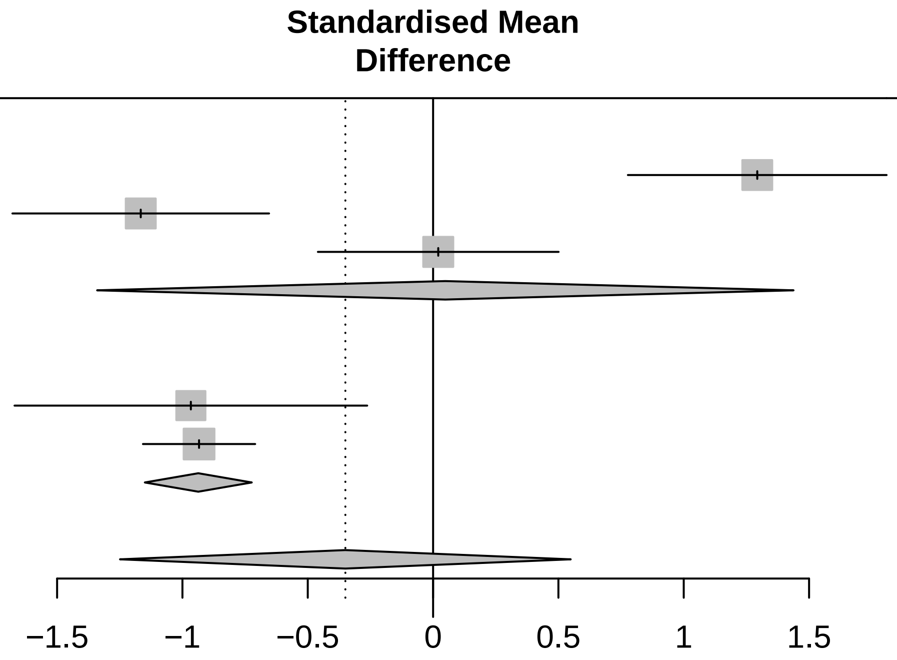

<− Faster progression for fast−progressing subtype | faster progression for slow−progressing subtype −>

Three−level metaanalysis using random effects to calculate an overall SMD estimate for Conceptualization across cohorts. The dashed line indicates the overall mean SMD estimate. The solid line indicates no effect.

### Forest plot for progression characteristics of symptom domain Visuo−executive (validation)

| Study | Slow progressing Total | Slow progressing Mean | Slow progressing SD | Fast progressing Total | Fast progressing Mean | Fast progressing SD | SMD | 95%−CI |
|---|---|---|---|---|---|---|---|---|
| **Cohort = LuxPARK (validation)** | | | | | | | | |
| FAB 3−6 | 48 | 0.06 | 0.0214 | 9 | 0.22 | 0.1676 | −2.32 | [−3.16; −1.48] |
| MOCA Visuspatial/Executive | 462 | 1.00 | 0.0008 | 99 | 2.73 | 0.5962 | −6.92 | [−7.38; −6.46] |
| Trailmaking A | 440 | 0.02 | 0.0161 | 86 | 0.10 | 0.1102 | −1.72 | [−1.97; −1.46] |
| Trailmaking B | 396 | 0.04 | 0.0216 | 60 | 0.10 | 0.0701 | −1.75 | [−2.05; −1.46] |
| **Random effects model** | **1346** | | | **254** | | | **−3.18** | **[−5.64; −0.71]** |
| **Cohort = ICEBERG (validation)** | | | | | | | | |
| FAB 3−6 | 127 | 0.06 | 0.0507 | 18 | 0.01 | 0.0948 | 0.89 | [ 0.39; 1.40] |
| MATTIS Initiation + Construction | 127 | 0.03 | 0.0602 | 18 | 0.07 | 0.0242 | −0.64 | [−1.14; −0.14] |
| MOCA Visuspatial/Executive | 135 | 0.69 | 0.6431 | 19 | 2.93 | 2.0152 | −2.42 | [−2.97; −1.87] |
| **Random effects model** | **389** | | | **55** | | | **−0.72** | **[−2.59; 1.16]** |
| **Random effects model** | **1735** | | | **309** | | | **−2.12** | **[−3.91; −0.34]** |

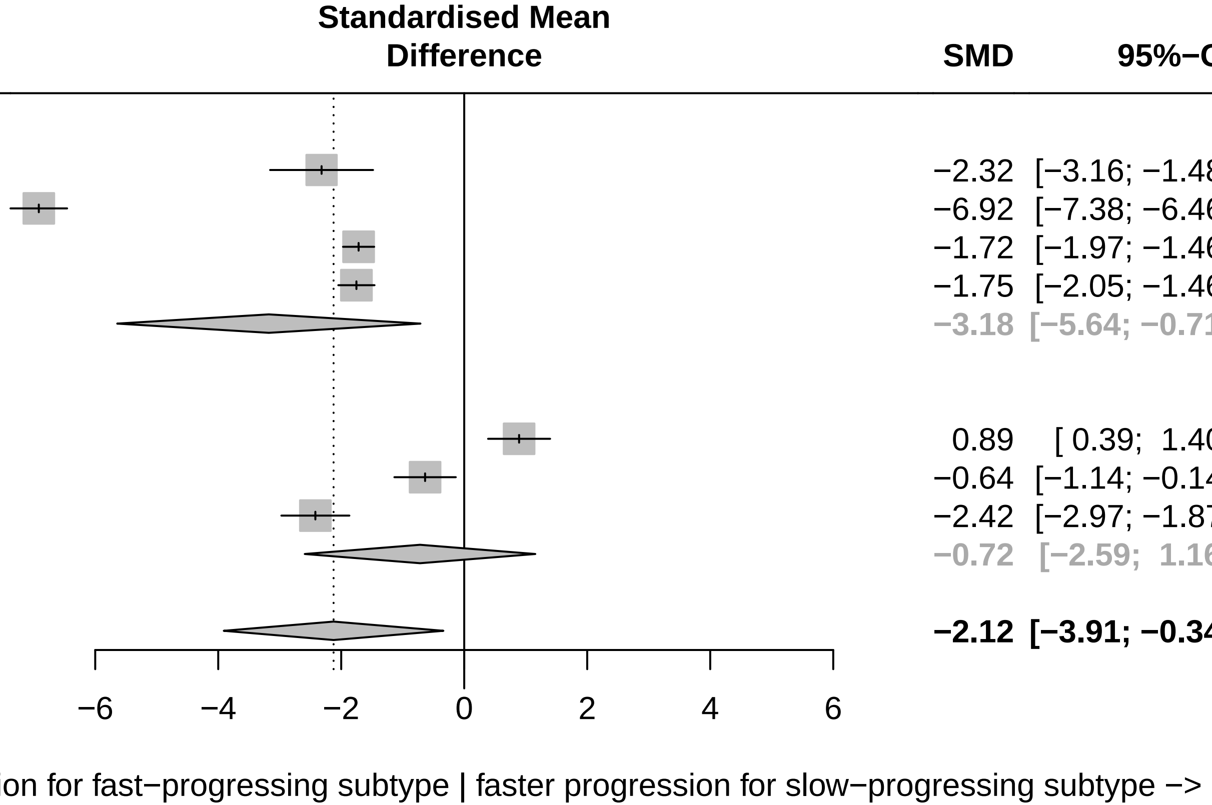

Three−level metaanalysis using random effects to calculate an overall SMD estimate for Visuo−executive across cohorts. The dashed line indicates the overall mean SMD estimate. The solid line indicates no effect.

### Forest plot for progression characteristics of symptom domain Language (validation)

| Study | Slow progressing Total | Slow progressing Mean | Slow progressing SD | Fast progressing Total | Fast progressing Mean | Fast progressing SD | SMD | 95%−CI |
|---|---|---|---|---|---|---|---|---|
| **Cohort = LuxPARK (validation)** | | | | | | | | |
| FAB VFT | 49 | 0.61 | 2.3727 | 10 | 2.27 | 1.7866 | −0.71 | [−1.41; −0.02] |
| MOCA Language + Naming | 462 | 0.04 | 0.0173 | 99 | 0.18 | 0.0729 | −4.19 | [−4.52; −3.86] |
| VFT phonematic F | 374 | −0.00 | 0.0089 | 87 | 0.06 | 0.0011 | −7.27 | [−7.80; −6.75] |
| VFT phonematic S | 57 | −0.02 | 0.0422 | 14 | 0.17 | 0.1257 | −2.94 | [−3.71; −2.18] |
| **Random effects model** | **942** | | | **210** | | | **−3.79** | **[−6.47; −1.11]** |
| **Cohort = ICEBERG (validation)** | | | | | | | | |
| FAB VFT | 127 | −0.43 | 0.5594 | 18 | 0.38 | 0.0389 | −1.53 | [−2.06; −1.01] |
| MOCA Language + Naming | 135 | 0.03 | 0.0221 | 19 | 0.08 | 0.0620 | −1.62 | [−2.14; −1.11] |
| VFT phonematic F | 135 | 0.03 | 0.0352 | 19 | 0.04 | 0.0665 | −0.27 | [−0.75; 0.22] |
| VFT phonematic S | 123 | 0.00 | 0.0105 | 18 | 0.03 | 0.0003 | −2.59 | [−3.17; −2.01] |
| VFT semantic supermarket | 122 | 0.05 | 0.0317 | 18 | 0.04 | 0.0017 | 0.40 | [−0.10; 0.89] |
| **Random effects model** | **642** | | | **92** | | | **−1.12** | **[−2.15; −0.08]** |
| **Random effects model** | **1584** | | | **302** | | | **−2.31** | **[−3.84; −0.77]** |

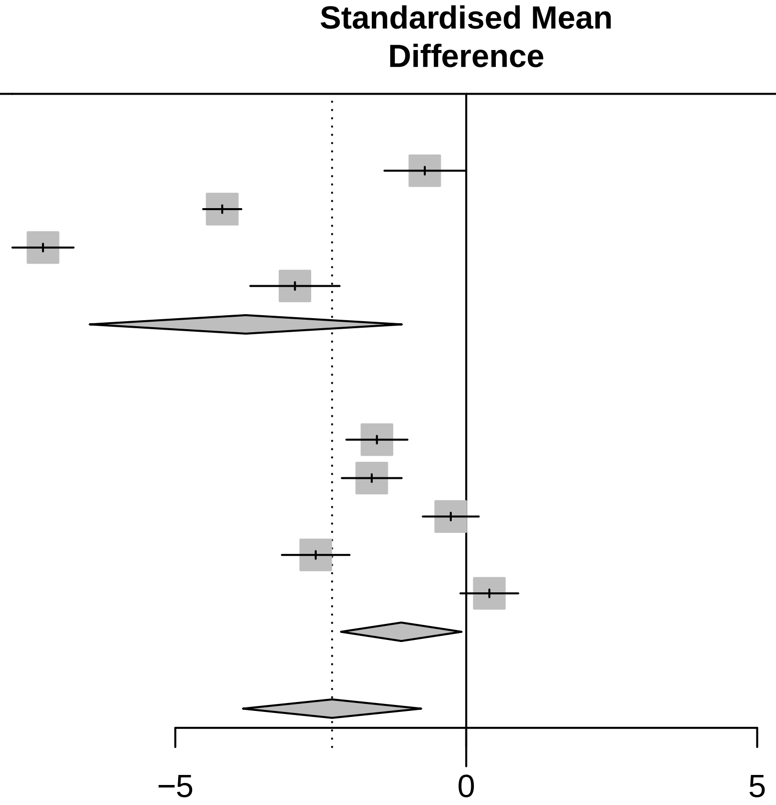

<− Faster progression for fast−progressing subtype | faster progression for slow−progressing subtype −>

Three−level metaanalysis using random effects to calculate an overall SMD estimate for Language across cohorts.
The dashed line indicates the overall mean SMD estimate. The solid line indicates no effect.

### Forest plot for progression characteristics of symptom domain Anxiety (validation)

| Study | Slow progressing Total | Slow progressing Mean | Slow progressing SD | Fast progressing Total | Fast progressing Mean | Fast progressing SD | SMD | 95%−CI |
|---|---|---|---|---|---|---|---|---|
| **Cohort = ICEBERG (validation)** | | | | | | | | |
| HADS anxiety | 135 | 0.03 | 0.0129 | 19 | 0.09 | 0.0045 | −4.57 | [−5.28; −3.87] |
| NMSQ Anxiety | 134 | 1.14 | 0.3856 | 19 | 2.46 | 2.1622 | −1.58 | [−2.09; −1.07] |
| UPDRS I Anxiety | 135 | 0.89 | 0.0973 | 19 | 1.66 | 2.5099 | −0.88 | [−1.37; −0.39] |
| **Random effects model** | **404** | | | **57** | | | **−2.33** | **[−4.54; −0.12]** |
| **Cohort = LuxPARK (validation)** | | | | | | | | |
| NMSQ Anxiety | 462 | 1.15 | 0.3264 | 97 | 1.40 | 0.2377 | −0.79 | [−1.02; −0.57] |
| PDQ39 Anxiety | 462 | 1.10 | 0.2517 | 99 | 1.78 | 1.2006 | −1.23 | [−1.46; −1.00] |
| UPDRS I Anxiety | 462 | 0.73 | 0.2466 | 99 | 0.89 | 0.7241 | −0.41 | [−0.63; −0.19] |
| **Random effects model** | **1386** | | | **295** | | | **−0.81** | **[−1.27; −0.34]** |
| **Random effects model** | **1790** | | | **352** | | | **−1.55** | **[−2.74; −0.37]** |

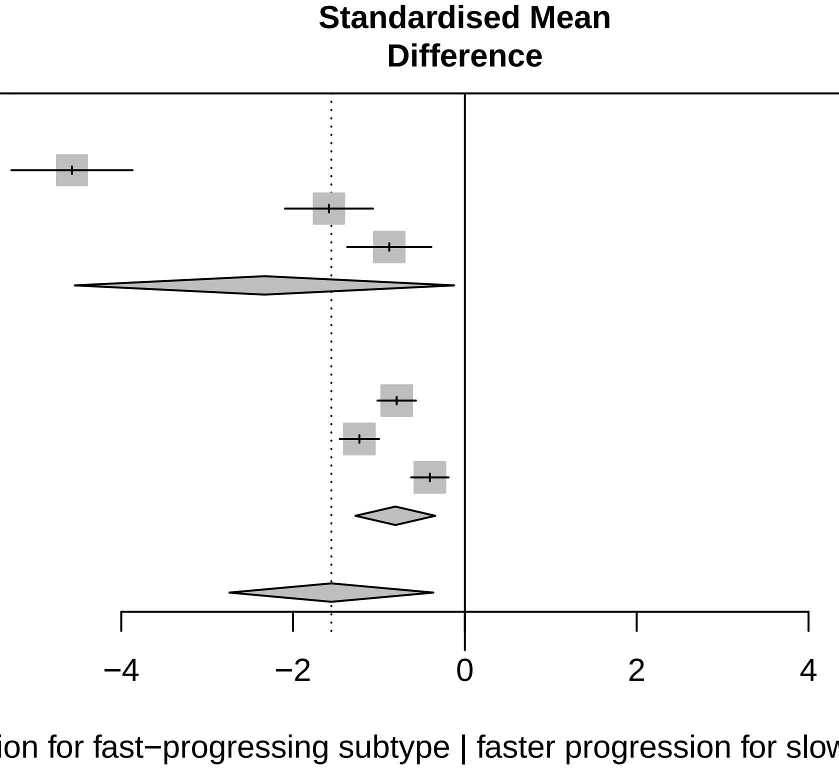

Three−level metaanalysis using random effects to calculate an overall SMD estimate for Anxiety across cohorts.
The dashed line indicates the overall mean SMD estimate. The solid line indicates no effect.

### Forest plot for progression characteristics of symptom domain Memory (validation)

| Study | Slow progressing Total | Slow progressing Mean | Slow progressing SD | Fast progressing Total | Fast progressing Mean | Fast progressing SD | SMD | 95%−CI |
|---|---|---|---|---|---|---|---|---|
| **Cohort = ICEBERG (validation)** | | | | | | | | |
| MATTIS Memory | 127 | 0.07 | 0.0498 | 18 | 0.09 | 0.0571 | −0.48 | [−0.98; 0.01] |
| MOCA Orientation + Memory | 135 | 0.01 | 0.0232 | 19 | 0.06 | 0.0612 | −1.66 | [−2.18; −1.14] |
| NMSQ Memory | 134 | 1.37 | 1.2394 | 19 | 1.95 | 2.0623 | −0.43 | [−0.91; 0.06] |
| **Random effects model** | **396** | | | **56** | | | **−0.85** | **[−1.64; −0.07]** |
| **Cohort = LuxPARK (validation)** | | | | | | | | |
| MOCA Orientation + Memory | 462 | 0.01 | 0.0199 | 99 | 0.14 | 0.1483 | −1.91 | [−2.15; −1.67] |
| NMSQ Memory | 462 | 1.18 | 0.1844 | 97 | 2.77 | 1.1996 | −3.02 | [−3.30; −2.74] |
| SIQCDE Memory | 397 | 0.04 | 0.0417 | 77 | 0.08 | 0.0691 | −0.93 | [−1.18; −0.68] |
| **Random effects model** | **1321** | | | **273** | | | **−1.95** | **[−3.14; −0.77]** |
| **Random effects model** | **1717** | | | **329** | | | **−1.42** | **[−2.22; −0.62]** |

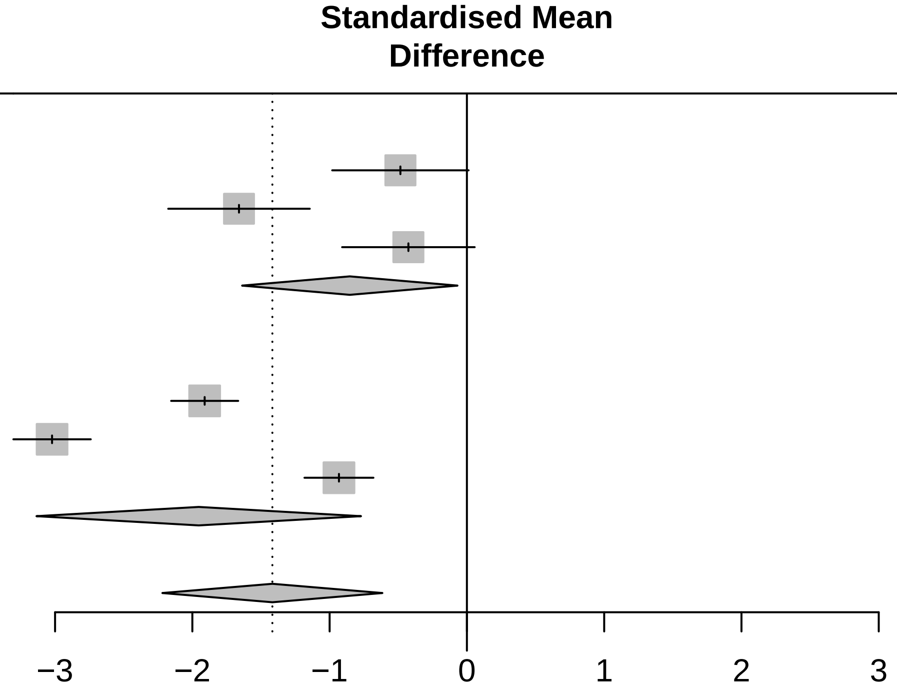

<− Faster progression for fast−progressing subtype | faster progression for slow−progressing subtype −>

Three−level metaanalysis using random effects to calculate an overall SMD estimate for Memory across cohorts.
The dashed line indicates the overall mean SMD estimate. The solid line indicates no effect.

### Forest plot for progression characteristics of symptom domain Attention (validation)

| Study | Slow progressing Total | Slow progressing Mean | Slow progressing SD | Fast progressing Total | Fast progressing Mean | Fast progressing SD | SMD | 95%−CI |
|---|---|---|---|---|---|---|---|---|
| **Cohort = ICEBERG (validation)** | | | | | | | | |
| MATTIS Attention | 127 | 0.01 | 0.0320 | 18 | −0.07 | 0.0070 | 2.63 | [ 2.05; 3.21] |
| MOCA Attention | 135 | 0.63 | 0.1829 | 19 | 1.40 | 1.5398 | −1.37 | [−1.88; −0.87] |
| NMSQ Attention | 134 | 2.26 | 0.0973 | 19 | 0.47 | 0.8163 | 6.02 | [ 5.18; 6.85] |
| **Random effects model** | **396** | | | **56** | | | **2.41** | **[−1.77; 6.60]** |
| **Cohort = LuxPARK (validation)** | | | | | | | | |
| MOCA Attention | 462 | 0.67 | 0.2059 | 99 | 2.00 | 0.8758 | −3.22 | [−3.50; −2.93] |
| NMSQ Attention | 462 | 1.53 | 0.9835 | 98 | 2.55 | 0.6917 | −1.07 | [−1.30; −0.85] |
| PDQ39 Attention | 462 | 1.41 | 0.1286 | 99 | 1.88 | 0.9219 | −1.17 | [−1.40; −0.94] |
| SIQCDE Attention | 398 | 0.58 | 0.4896 | 77 | 1.30 | 0.8721 | −1.26 | [−1.52; −1.00] |
| **Random effects model** | **1784** | | | **373** | | | **−1.68** | **[−2.68; −0.67]** |
| **Random effects model** | **2180** | | | **429** | | | **0.06** | **[−2.25; 2.38]** |

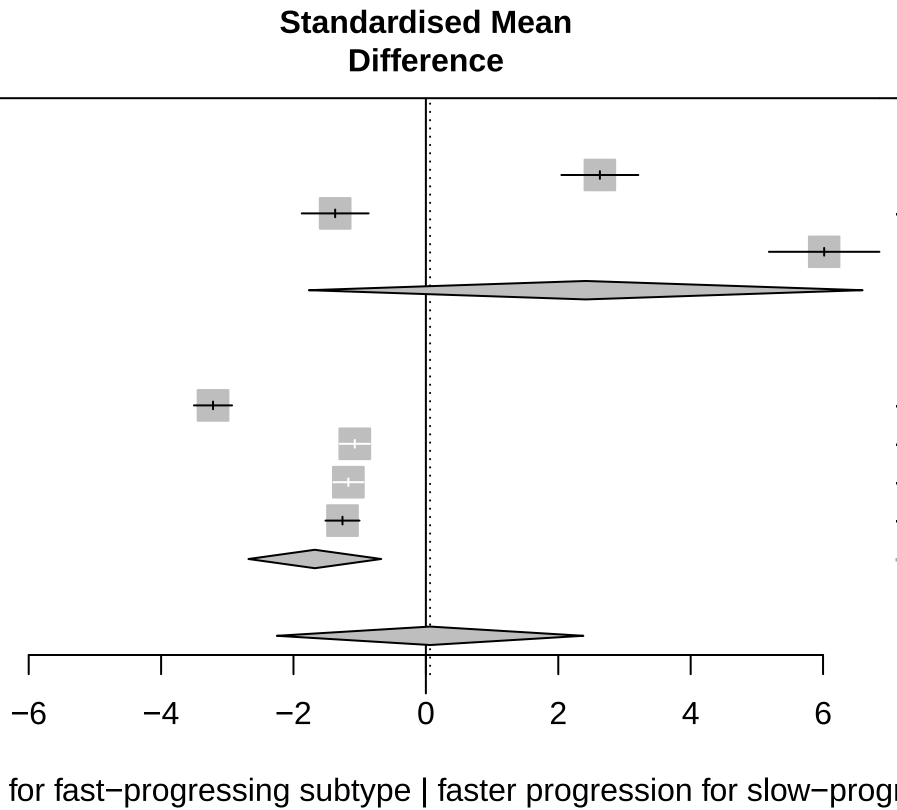

Three−level metaanalysis using random effects to calculate an overall SMD estimate for Attention across cohorts.
The dashed line indicates the overall mean SMD estimate. The solid line indicates no effect.

### Forest plot for progression characteristics of symptom domain Non motor symptoms (validation)

| Study | Slow progressing Total | Slow progressing Mean | Slow progressing SD | Fast progressing Total | Fast progressing Mean | Fast progressing SD | SMD | 95%−CI |
|---|---|---|---|---|---|---|---|---|
| **Cohort = ICEBERG (validation)** | | | | | | | | |
| NMSQ | 134 | 0.20 | 0.0347 | 19 | 0.22 | 0.0014 | −0.69 | [−1.18; −0.20] |
| UPDRS I | 135 | 0.14 | 0.0230 | 19 | 0.23 | 0.0397 | −3.28 | [−3.89; −2.67] |
| **Random effects model** | **269** | | | **38** | | | **−1.98** | **[−4.52; 0.56]** |
| **Cohort = LuxPARK (validation)** | | | | | | | | |
| NMSQ | 455 | 0.12 | 0.0300 | 95 | 0.21 | 0.0641 | −2.42 | [−2.68; −2.16] |
| UPDRS I | 462 | 0.12 | 0.0412 | 99 | 0.24 | 0.0886 | −2.20 | [−2.46; −1.95] |
| **Random effects model** | **917** | | | **194** | | | **−2.31** | **[−2.52; −2.10]** |
| **Random effects model** | **1186** | | | **232** | | | **−2.14** | **[−3.17; −1.11]** |

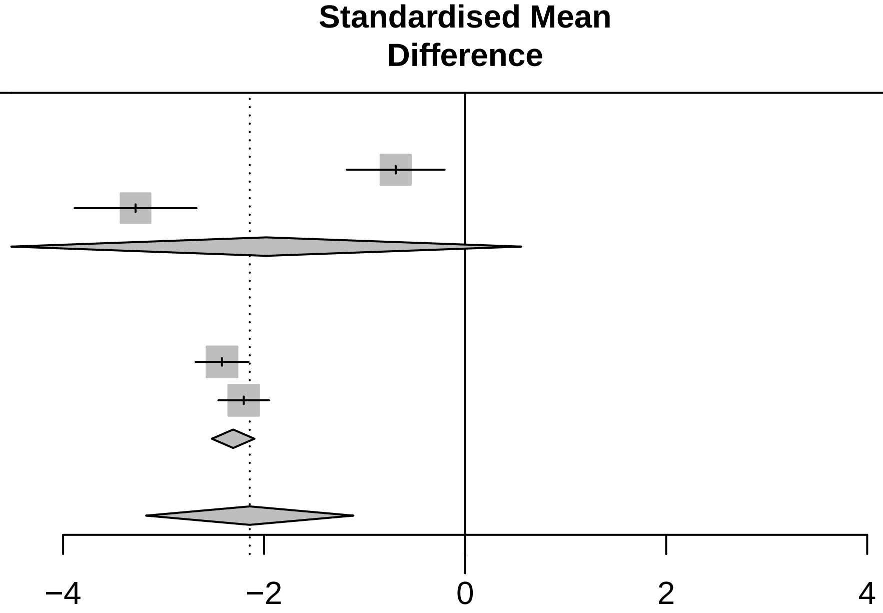

<− Faster progression for fast−progressing subtype | faster progression for slow−progressing subtype −>

Three−level metaanalysis using random effects to calculate an overall SMD estimate for Non motor symptoms across cohorts. The dashed line indicates the overall mean SMD estimate. The solid line indicates no effect.

### Forest plot for progression characteristics of symptom domain Autonomic (validation)

| Study | Slow progressing Total | Slow progressing Mean | Slow progressing SD | Fast progressing Total | Fast progressing Mean | Fast progressing SD | SMD | 95%−CI |
|---|---|---|---|---|---|---|---|---|
| **Cohort = ICEBERG (validation)** | | | | | | | | |
| NMSQ Autonomic | 135 | 0.19 | 0.0132 | 19 | 0.16 | 0.0734 | 1.21 | [ 0.71; 1.71] |
| SCOPA−AUT | 135 | 0.19 | 0.0481 | 19 | 0.28 | 0.0830 | −1.71 | [−2.22; −1.19] |
| UPDRS I Autonomic | 135 | 0.20 | 0.0600 | 19 | 0.21 | 0.0328 | −0.28 | [−0.76; 0.20] |
| **Random effects model** | **405** | | | **57** | | | **−0.26** | **[−1.91; 1.39]** |
| **Cohort = LuxPARK (validation)** | | | | | | | | |
| NMSQ Autonomic | 456 | 0.13 | 0.0288 | 97 | 0.15 | 0.0716 | −0.62 | [−0.84; −0.39] |
| SCOPA−AUT | 462 | 0.13 | 0.0363 | 99 | 0.17 | 0.0566 | −0.91 | [−1.13; −0.68] |
| UPDRS I Autonomic | 462 | 0.13 | 0.0428 | 99 | 0.22 | 0.0693 | −1.85 | [−2.09; −1.60] |
| **Random effects model** | **1380** | | | **295** | | | **−1.12** | **[−1.85; −0.39]** |
| **Random effects model** | **1785** | | | **352** | | | **−0.70** | **[−1.58; 0.18]** |

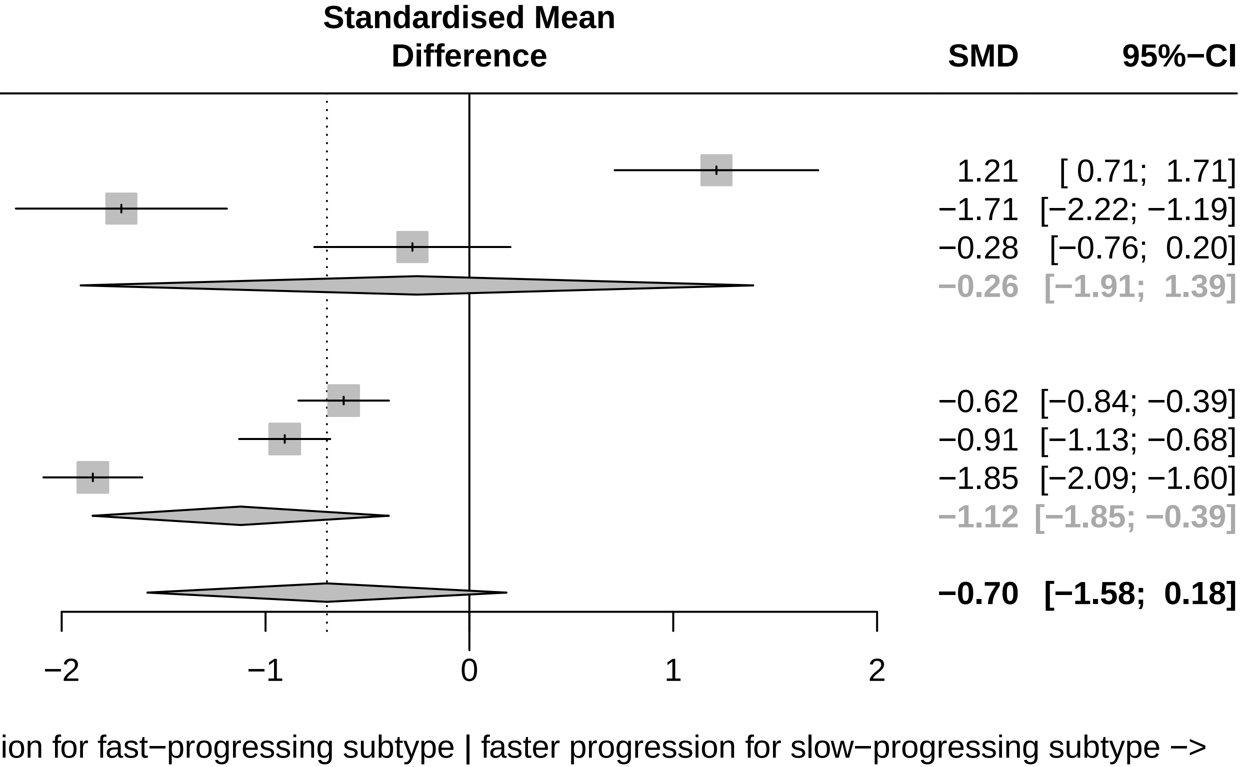

Three−level metaanalysis using random effects to calculate an overall SMD estimate for Autonomic across cohorts.
The dashed line indicates the overall mean SMD estimate. The solid line indicates no effect.

### Forest plot for progression characteristics of symptom domain Hallucinations (validation)

| Study | Slow progressing Total | Slow progressing Mean | Slow progressing SD | Fast progressing Total | Fast progressing Mean | Fast progressing SD | SMD | 95%−CI |
|---|---|---|---|---|---|---|---|---|
| **Cohort = LuxPARK (validation)** | | | | | | | | |
| NMSQ Hallucination | 462 | 1.67 | 0.2022 | 98 | 1.81 | 1.0142 | −0.30 | [−0.52; −0.09] |
| UPDRS I Hallucinations | 462 | 1.31 | 0.2786 | 99 | 1.38 | 0.8747 | −0.16 | [−0.38; 0.06] |
| **Random effects model** | **924** | | | **197** | | | **−0.23** | **[−0.39; −0.08]** |
| **Cohort = ICEBERG (validation)** | | | | | | | | |
| NMSQ Hallucination | 135 | 0.93 | 0.9204 | 19 | 1.38 | 0.7406 | −0.50 | [−0.99; −0.02] |
| UPDRS I Hallucinations | 135 | 1.03 | 2.0070 | 19 | 8.38 | 9.9115 | −1.88 | [−2.40; −1.35] |
| **Random effects model** | **270** | | | **38** | | | **−1.19** | **[−2.53; 0.16]** |
| **Random effects model** | **1194** | | | **235** | | | **−0.69** | **[−1.44; 0.06]** |

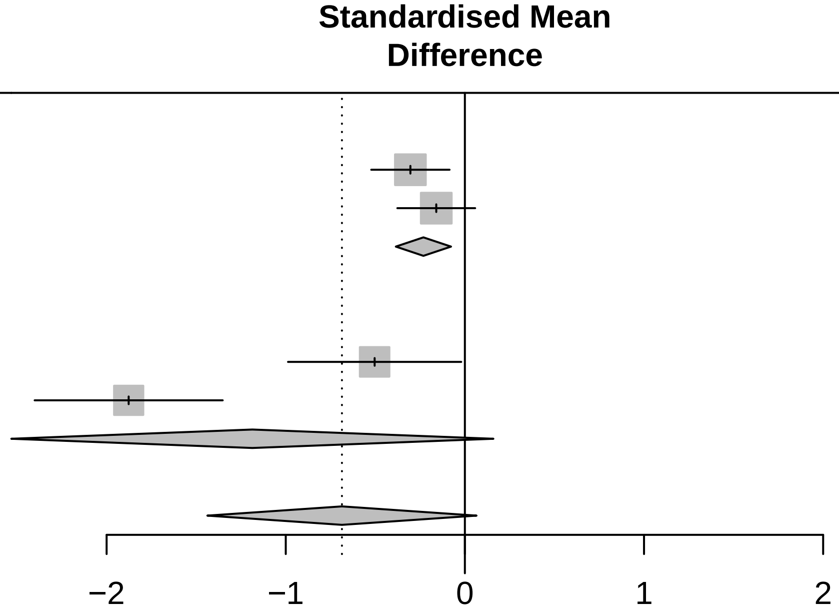

<− Faster progression for fast−progressing subtype | faster progression for slow−progressing subtype −>

Three−level metaanalysis using random effects to calculate an overall SMD estimate for Hallucinations across cohorts. The dashed line indicates the overall mean SMD estimate. The solid line indicates no effect.

### Forest plot for progression characteristics of symptom domain Pain (validation)

| Study | Slow progressing Total | Slow progressing Mean | Slow progressing SD | Fast progressing Total | Fast progressing Mean | Fast progressing SD | SMD | 95%−CI |
|---|---|---|---|---|---|---|---|---|
| Cohort = LuxPARK (validation) | | | | | | | | |
| NMSQ Pain | 462 | 0.79 | 0.0950 | 98 | 1.35 | 0.0765 | −6.03 | [−6.45; −5.62] |
| PDQ39 Pain | 461 | 0.11 | 0.0314 | 99 | 0.12 | 0.0169 | −0.29 | [−0.51; −0.08] |
| UPDRS I Pain | 462 | 1.17 | 0.2438 | 99 | 1.69 | 0.8251 | −1.26 | [−1.49; −1.03] |
| Random effects model | 1385 | | | 296 | | | −2.53 | [−6.00; 0.95] |
| Cohort = ICEBERG (validation) | | | | | | | | |
| NMSQ Pain | 134 | 0.68 | 0.3044 | 19 | 0.81 | 0.4887 | −0.39 | [−0.88; 0.09] |
| UPDRS I Pain | 135 | 1.06 | 0.1449 | 19 | 0.69 | 0.1723 | 2.47 | [ 1.92; 3.03] |
| Random effects model | 269 | | | 38 | | | 1.04 | [−1.77; 3.85] |
| **Random effects model** | **1654** | | | **334** | | | **−1.11** | **[−3.81; 1.60]** |

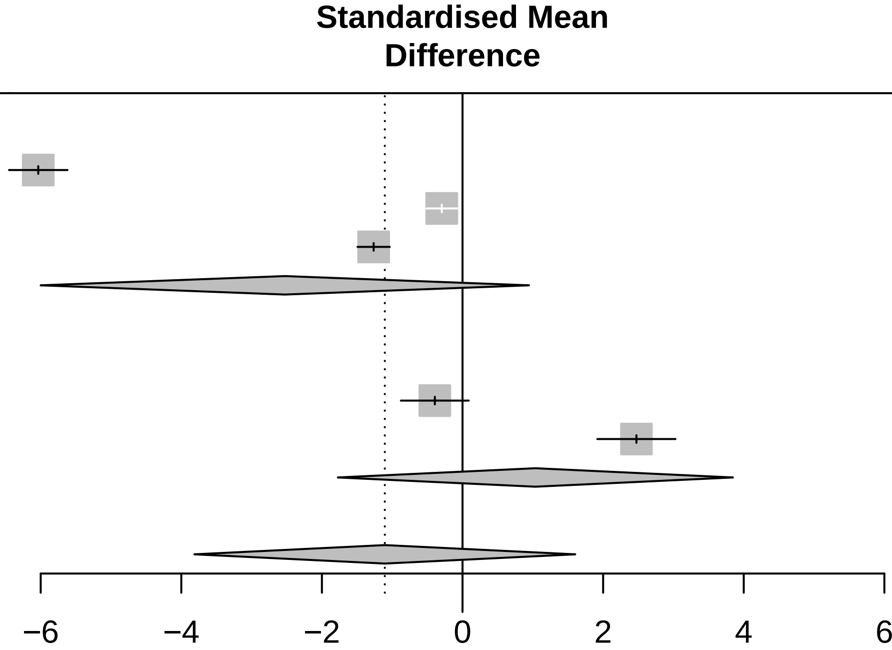

<− Faster progression for fast−progressing subtype | faster progression for slow−progressing subtype −>

Three−level metaanalysis using random effects to calculate an overall SMD estimate for Pain across cohorts. The dashed line indicates the overall mean SMD estimate. The solid line indicates no effect.

### Forest plot for progression characteristics of symptom domain RBD (validation)

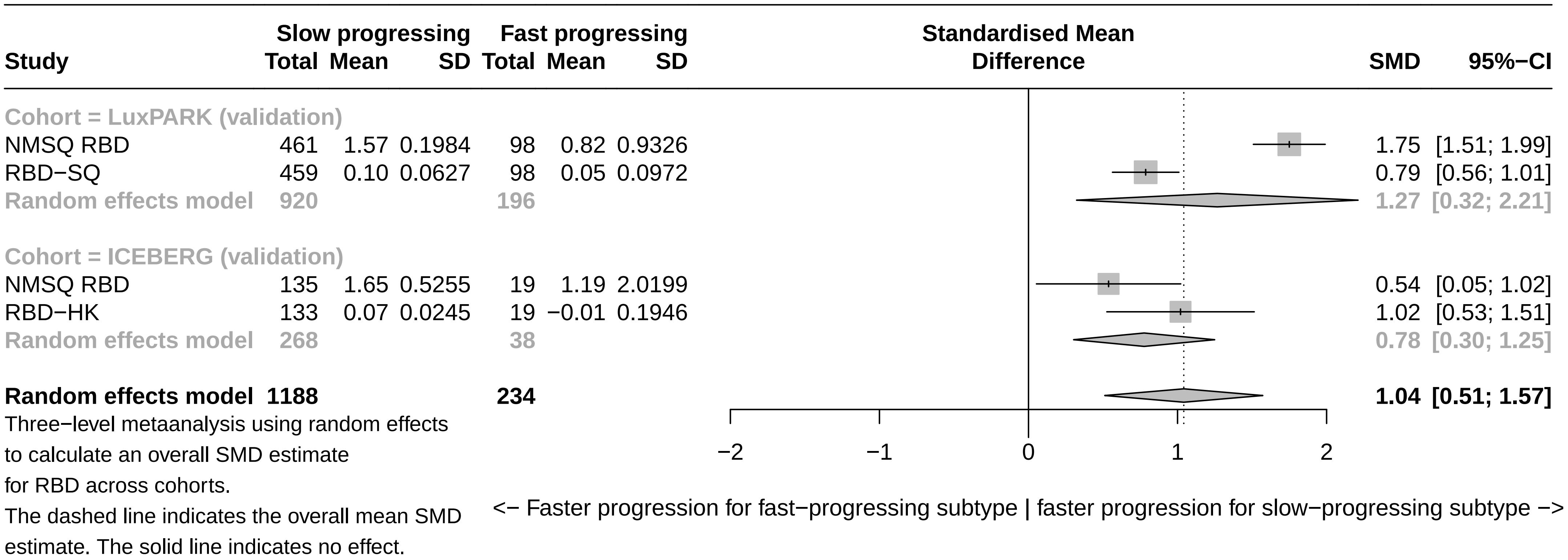

### Forest plot for progression characteristics of symptom domain Smell (validation)

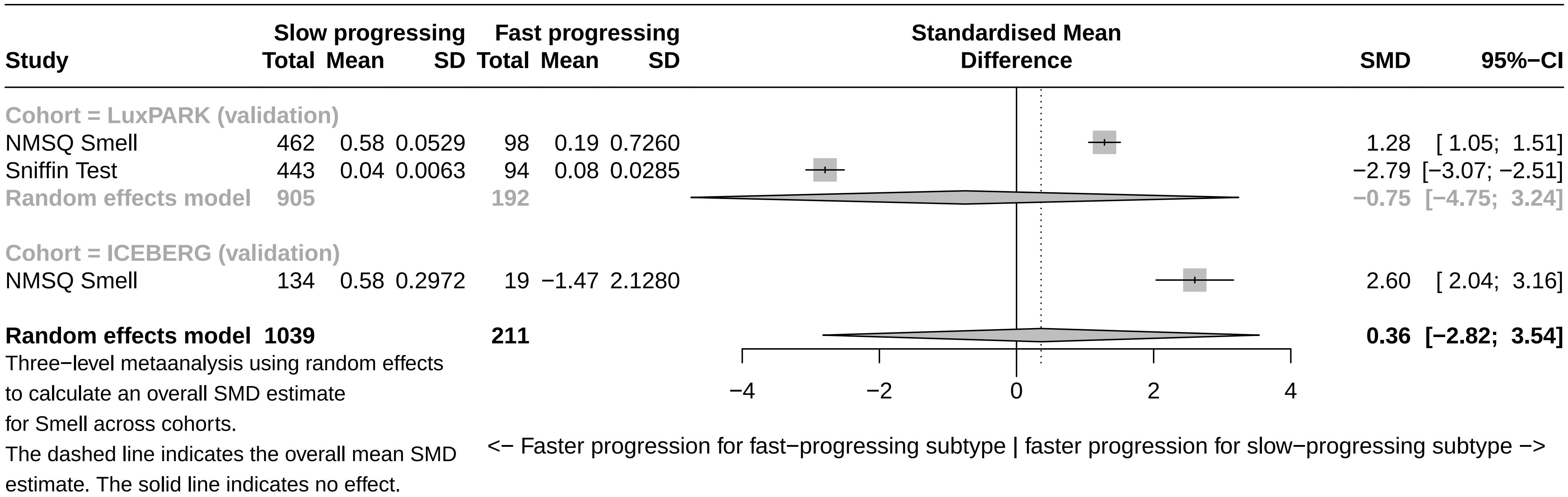

### Forest plot for progression characteristics of symptom domain Motor symptoms (validation)

| Study | Slow progressing Total | Slow progressing Mean | Slow progressing SD | Fast progressing Total | Fast progressing Mean | Fast progressing SD | SMD | 95%−CI |
|---|---|---|---|---|---|---|---|---|
| **Cohort = LuxPARK (validation)** | | | | | | | | |
| PDQ39 ADL | 462 | 0.17 | 0.0828 | 98 | 0.40 | 0.2464 | −1.85 | [−2.09; −1.60] |
| Pegboard | 429 | 0.06 | 0.0171 | 85 | 0.17 | 0.0578 | −3.85 | [−4.18; −3.52] |
| UPDRS II | 462 | 0.17 | 0.0671 | 99 | 0.49 | 0.2085 | −2.99 | [−3.27; −2.72] |
| UPDRS III on | 429 | 0.10 | 0.0289 | 90 | 0.29 | 0.0919 | −4.21 | [−4.56; −3.87] |
| UPDRS IV | 462 | 0.10 | 0.0806 | 99 | 0.09 | 0.0819 | 0.12 | [−0.09; 0.34] |
| **Random effects model** | **2244** | | | **471** | | | **−2.55** | **[−4.09; −1.01]** |
| **Cohort = ICEBERG (validation)** | | | | | | | | |
| UPDRS II | 135 | 0.22 | 0.0408 | 19 | 0.65 | 0.1997 | −5.47 | [−6.26; −4.69] |
| UPDRS III off | 134 | 0.07 | 0.0104 | 19 | 0.31 | 0.1850 | −3.64 | [−4.28; −3.01] |
| UPDRS III on | 127 | 0.03 | 0.0038 | 19 | 0.12 | 0.0846 | −2.94 | [−3.54; −2.35] |
| UPDRS IV | 135 | 0.09 | 0.1311 | 19 | 0.21 | 0.1229 | −0.93 | [−1.42; −0.44] |
| **Random effects model** | **531** | | | **76** | | | **−3.23** | **[−5.07; −1.39]** |
| **Random effects model** | **2775** | | | **547** | | | **−2.85** | **[−3.98; −1.72]** |

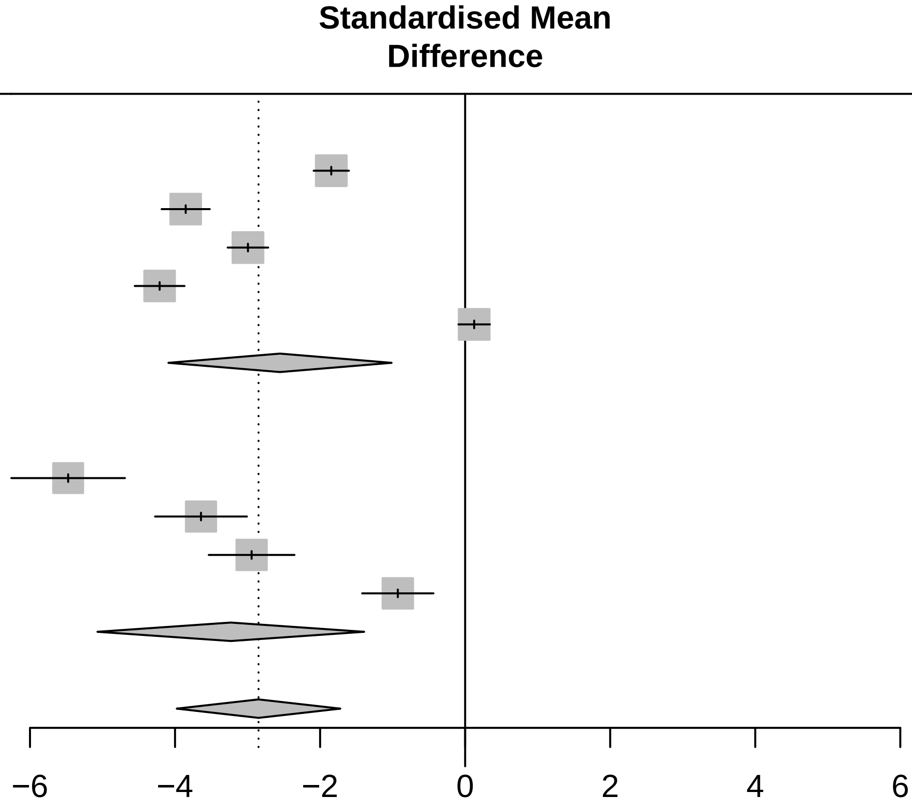

<− Faster progression for fast−progressing subtype | faster progression for slow−progressing subtype −>

Three−level metaanalysis using random effects to calculate an overall SMD estimate for Motor symptoms across cohorts. The dashed line indicates the overall mean SMD estimate. The solid line indicates no effect.

Forest plot for progression characteristics of symptom domain Impulsivity (validation)

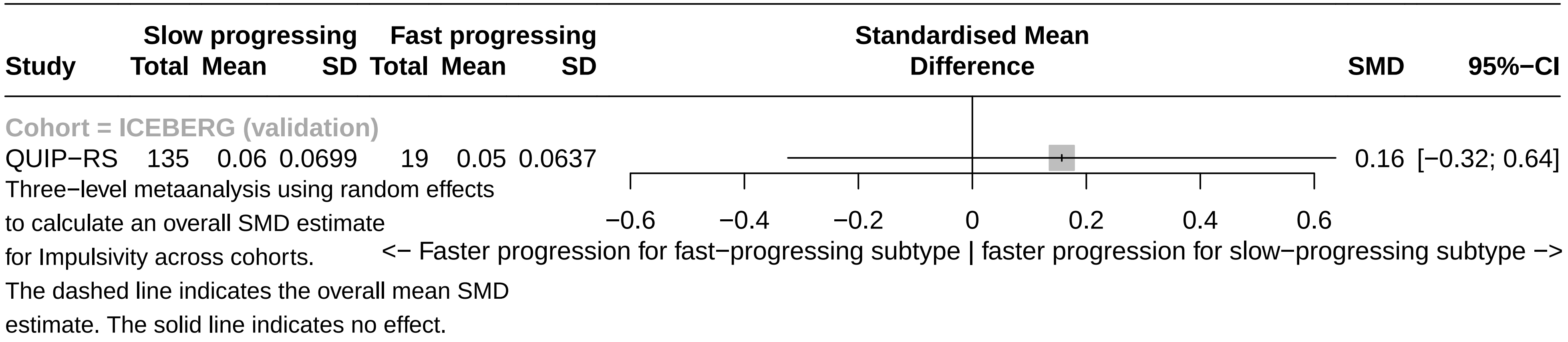

### Forest plots for symptom domain baseline associations (in cohort)

### Forest plot for baseline characteristics of symptom domain Axial & PIGD

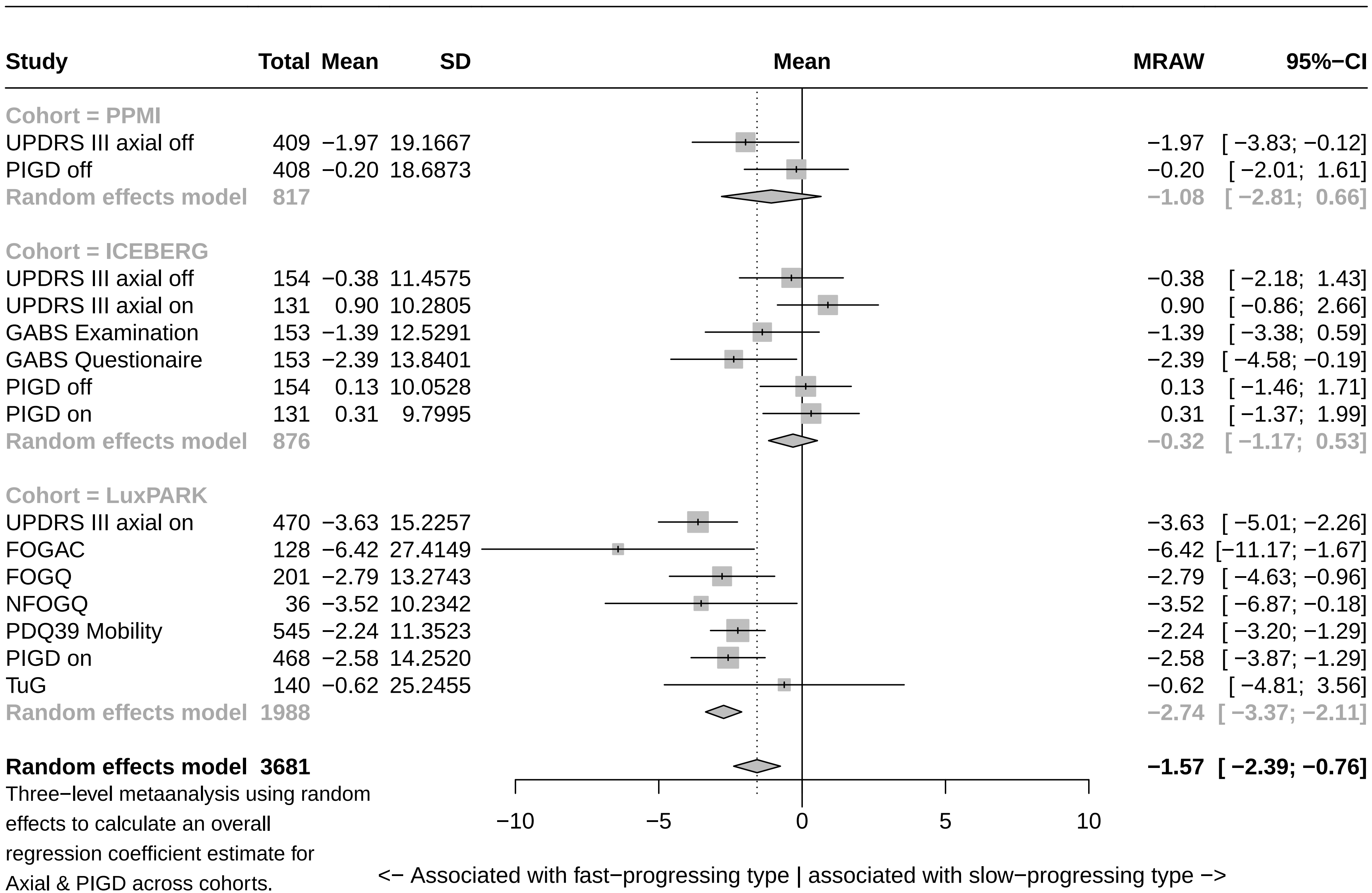

Three−level metaanalysis using random effects to calculate an overall regression coefficient estimate for Axial & PIGD across cohorts. The dashed line indicates the overall mean estimate. The solid line indicates no effect.

### Forest plot for baseline characteristics of symptom domain Depression

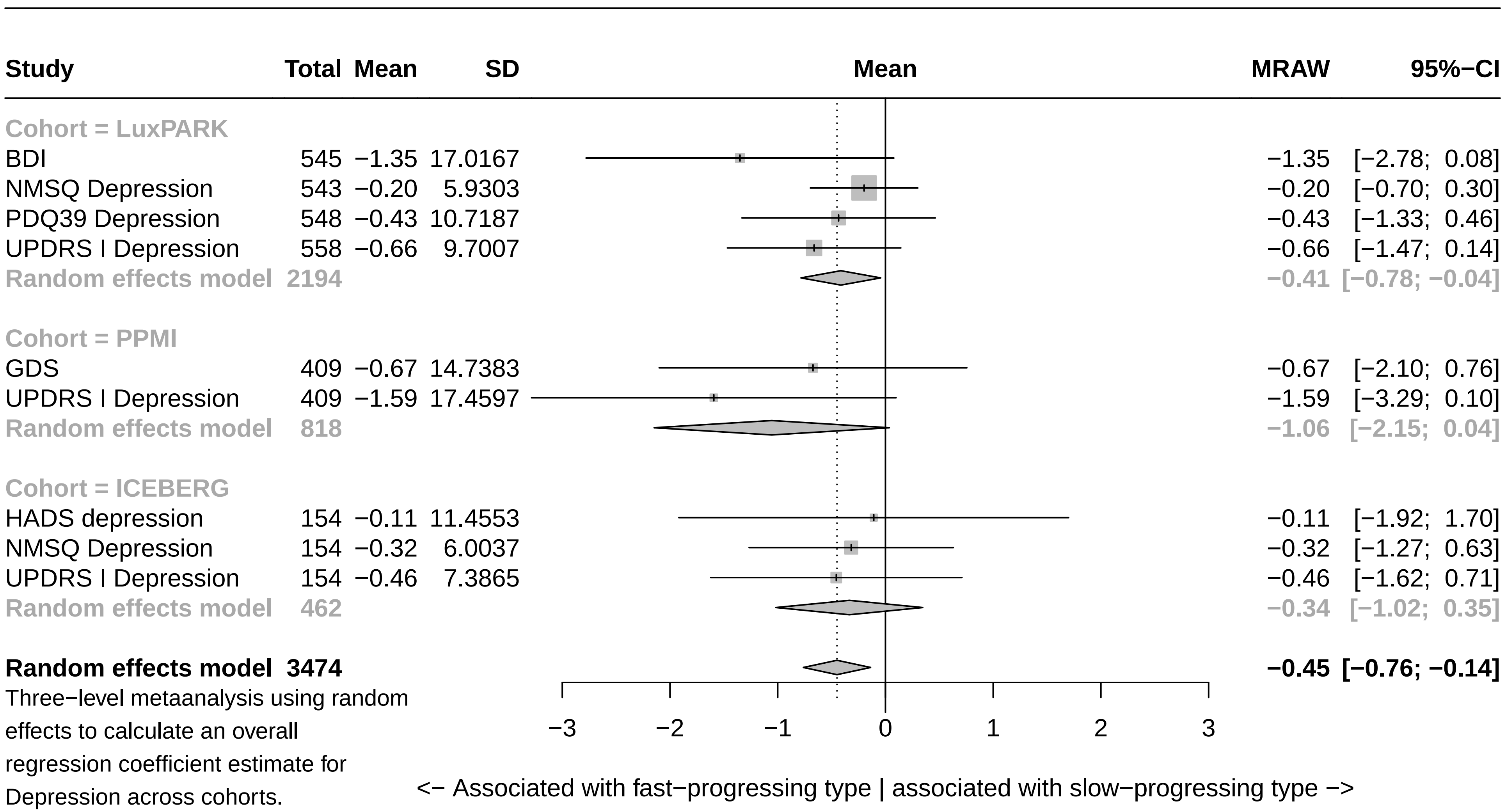

Three−level metaanalysis using random effects to calculate an overall regression coefficient estimate for Depression across cohorts.
The dashed line indicates the overall mean estimate. The solid line indicates no effect.

### Forest plot for baseline characteristics of symptom domain Memory

| Study | Total | Mean | SD | MRAW | 95%−CI |
|---|---|---|---|---|---|
| **Cohort = LuxPARK** | | | | | |
| CERAD Words DR | 212 | −0.99 | 9.7183 | −0.99 | [−2.30; 0.31] |
| CERAD Words IR | 216 | −1.63 | 12.8291 | −1.63 | [−3.35; 0.08] |
| CERAD Words Recognition | 217 | 0.59 | 18.0617 | 0.59 | [−1.81; 2.99] |
| MOCA Orientation + Memory | 559 | −2.54 | 12.4261 | −2.54 | [−3.57; −1.51] |
| NMSQ Memory | 545 | −0.39 | 4.6017 | −0.39 | [−0.77; 0.00] |
| PDQ39 Memory | 549 | −0.90 | 8.6625 | −0.90 | [−1.62; −0.17] |
| SIQCDE Memory | 234 | −1.96 | 18.3043 | −1.96 | [−4.30; 0.39] |
| **Random effects model** | **2532** | | | **−1.12** | **[−1.83; −0.40]** |
| **Cohort = PPMI** | | | | | |
| Hopkins Verbal Learning Test DR | 407 | −4.56 | 25.5606 | −4.56 | [−7.05; −2.08] |
| Hopkins Verbal Learning Test IR | 408 | −2.53 | 14.5925 | −2.53 | [−3.95; −1.11] |
| MOCA Orientation + Memory | 409 | −0.95 | 12.3831 | −0.95 | [−2.15; 0.25] |
| **Random effects model** | **1224** | | | **−2.44** | **[−4.35; −0.54]** |
| **Cohort = ICEBERG** | | | | | |
| MATTIS Memory | 154 | −0.29 | 9.4798 | −0.29 | [−1.79; 1.20] |
| MMSE Memory | 154 | −0.57 | 7.2104 | −0.57 | [−1.71; 0.57] |
| MOCA Orientation + Memory | 154 | 0.17 | 10.2795 | 0.17 | [−1.46; 1.79] |
| NMSQ Memory | 152 | −0.61 | 4.8031 | −0.61 | [−1.37; 0.16] |
| **Random effects model** | **614** | | | **−0.47** | **[−1.02; 0.08]** |
| **Random effects model** | **4370** | | | **−1.09** | **[−1.61; −0.57]** |

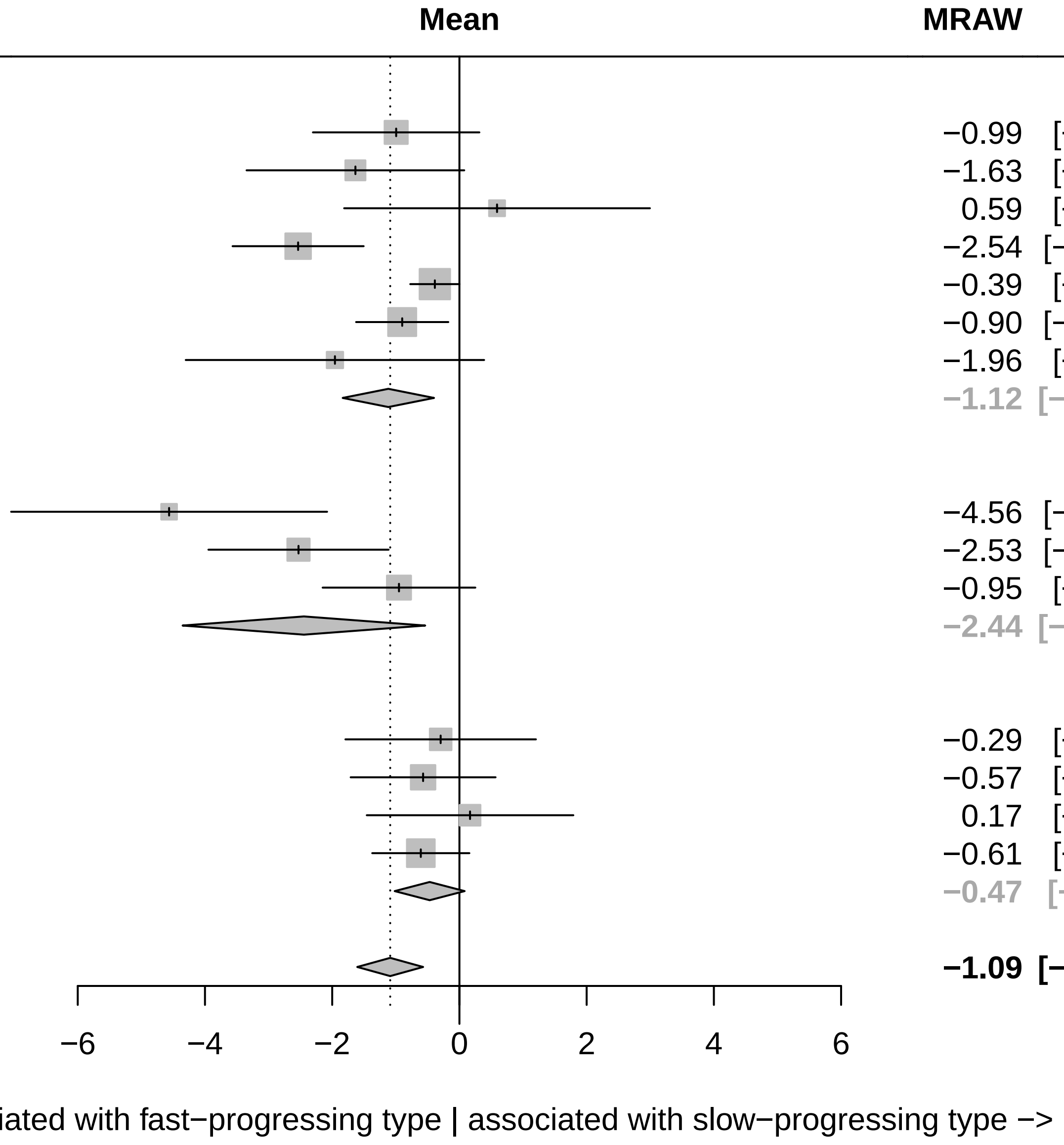

Three−level metaanalysis using random effects to calculate an overall regression coefficient estimate for Memory across cohorts.
The dashed line indicates the overall mean estimate. The solid line indicates no effect.

### Forest plot for baseline characteristics of symptom domain Overall severity

| Study | Total | Mean | SD | MRAW | 95%−CI |
|---|---|---|---|---|---|
| **Cohort = ICEBERG** | | | | | |
| CGIS | 154 | −0.69 | 10.7670 | −0.69 | [−2.39; 1.01] |
| H&Y | 154 | −2.42 | 19.1051 | −2.42 | [−5.44; 0.60] |
| SEADL | 154 | −1.45 | 15.6540 | −1.45 | [−3.93; 1.02] |
| UPDRS I−III off | 154 | −2.13 | 15.3270 | −2.13 | [−4.55; 0.29] |
| UPDRS I−III on | 131 | −1.14 | 12.3432 | −1.14 | [−3.26; 0.97] |
| **Random effects model** | **747** | | | **−1.34** | **[−2.34; −0.35]** |
| **Cohort = LuxPARK** | | | | | |
| FAQ | 230 | −1.73 | 11.0542 | −1.73 | [−3.16; −0.30] |
| H&Y | 560 | −3.24 | 17.6985 | −3.24 | [−4.70; −1.77] |
| PDQ39 | 531 | −2.52 | 13.7787 | −2.52 | [−3.69; −1.35] |
| UPDRS I−III on | 461 | −3.38 | 16.1434 | −3.38 | [−4.85; −1.91] |
| **Random effects model** | **1782** | | | **−2.68** | **[−3.36; −2.00]** |
| **Cohort = PPMI** | | | | | |
| H&Y | 409 | 0.12 | 5.4184 | 0.12 | [−0.40; 0.65] |
| SEADL | 409 | −0.78 | 14.0054 | −0.78 | [−2.14; 0.57] |
| UPDRS I−III off | 408 | −2.63 | 18.9616 | −2.63 | [−4.47; −0.80] |
| **Random effects model** | **1226** | | | **−0.90** | **[−2.41; 0.62]** |
| **Random effects model** | **3755** | | | **−1.75** | **[−2.50; −0.99]** |

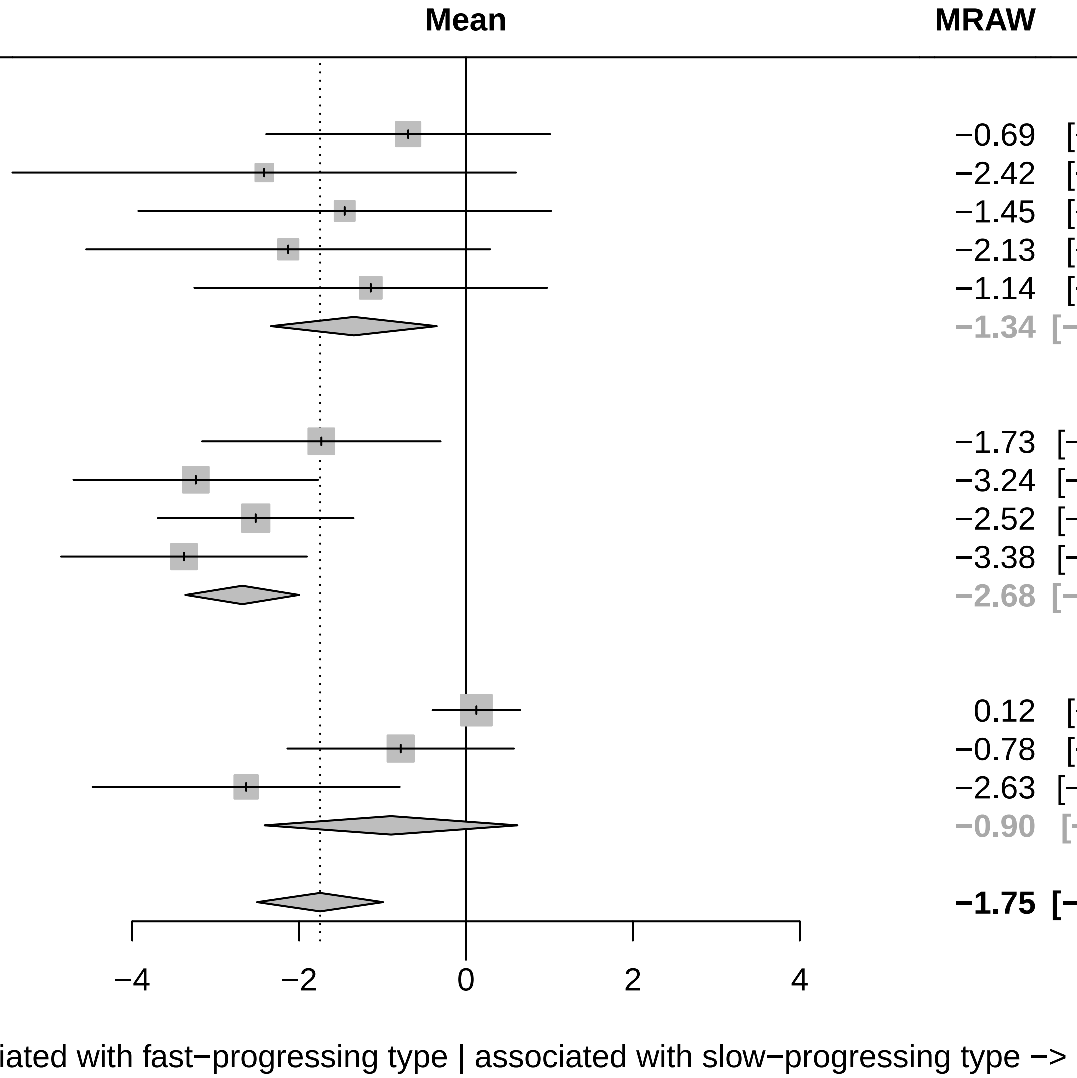

Three−level metaanalysis using random effects to calculate an overall regression coefficient estimate for Overall severity across cohorts. The dashed line indicates the overall mean estimate. The solid line indicates no effect.

### Forest plot for baseline characteristics of symptom domain Apathy

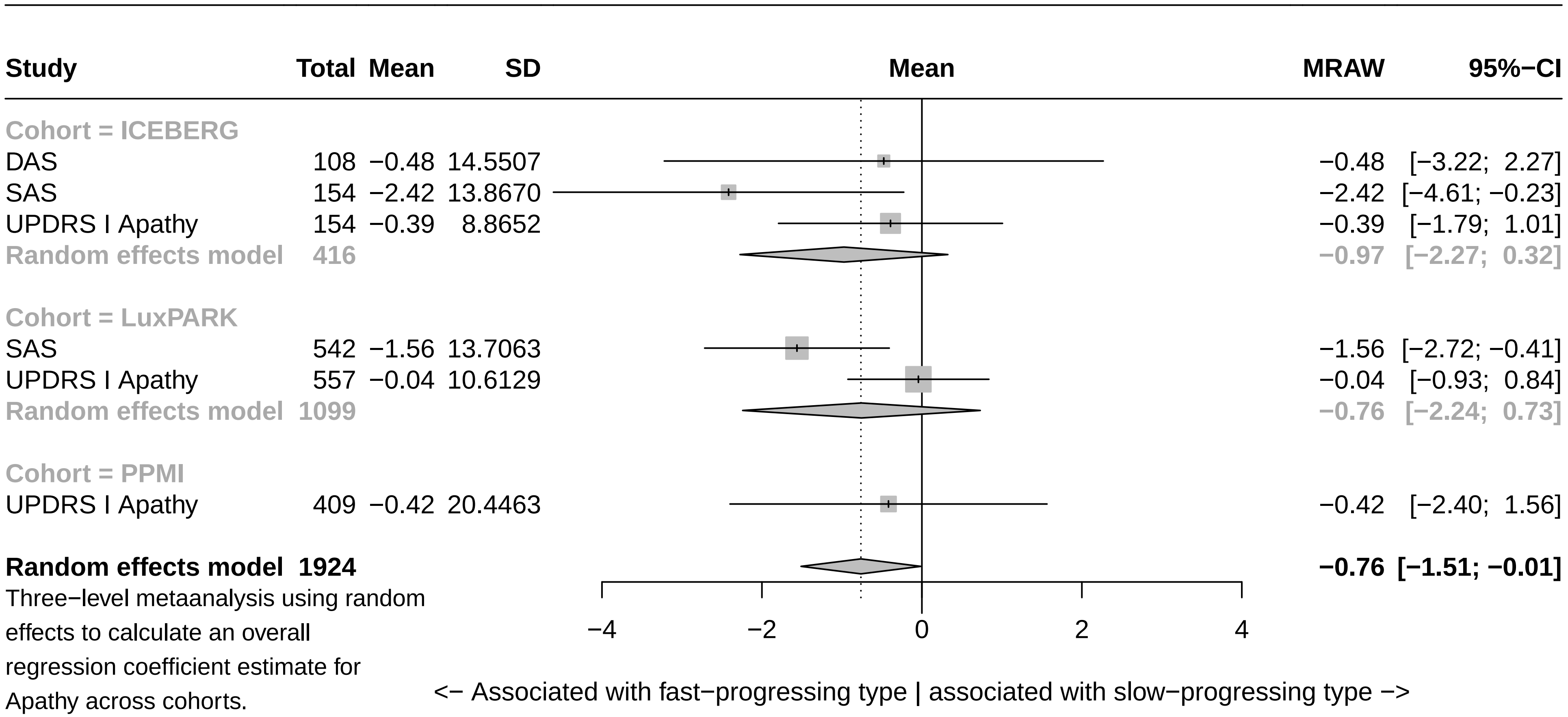

Three−level metaanalysis using random effects to calculate an overall regression coefficient estimate for Apathy across cohorts.
The dashed line indicates the overall mean estimate. The solid line indicates no effect.

### Forest plot for baseline characteristics of symptom domain Sleep

| Study | Total | Mean | SD | MRAW | 95%−CI |
|---|---|---|---|---|---|
| **Cohort = PPMI** | | | | | |
| ESS | 408 | −1.38 | 12.9174 | −1.38 | [−2.63; −0.12] |
| UPDRS I Sleep | 409 | −1.26 | 13.3186 | −1.26 | [−2.55; 0.03] |
| **Random effects model** | **817** | | | **−1.32** | **[−2.22; −0.42]** |
| **Cohort = ICEBERG** | | | | | |
| ESS | 143 | −0.62 | 11.0335 | −0.62 | [−2.43; 1.19] |
| NMSQ Sleep | 154 | −0.28 | 7.4813 | −0.28 | [−1.46; 0.91] |
| UPDRS I Sleep | 154 | 0.45 | 9.1040 | 0.45 | [−0.99; 1.89] |
| **Random effects model** | **451** | | | **−0.11** | **[−0.93; 0.70]** |
| **Cohort = LuxPARK** | | | | | |
| NMSQ Sleep | 545 | 0.03 | 6.7698 | 0.03 | [−0.54; 0.60] |
| PDSS | 541 | −0.88 | 13.5002 | −0.88 | [−2.02; 0.26] |
| UPDRS I Sleep | 558 | −0.37 | 10.4775 | −0.37 | [−1.24; 0.50] |
| **Random effects model** | **1644** | | | **−0.23** | **[−0.71; 0.24]** |
| **Random effects model** | **2912** | | | **−0.43** | **[−0.86; −0.01]** |

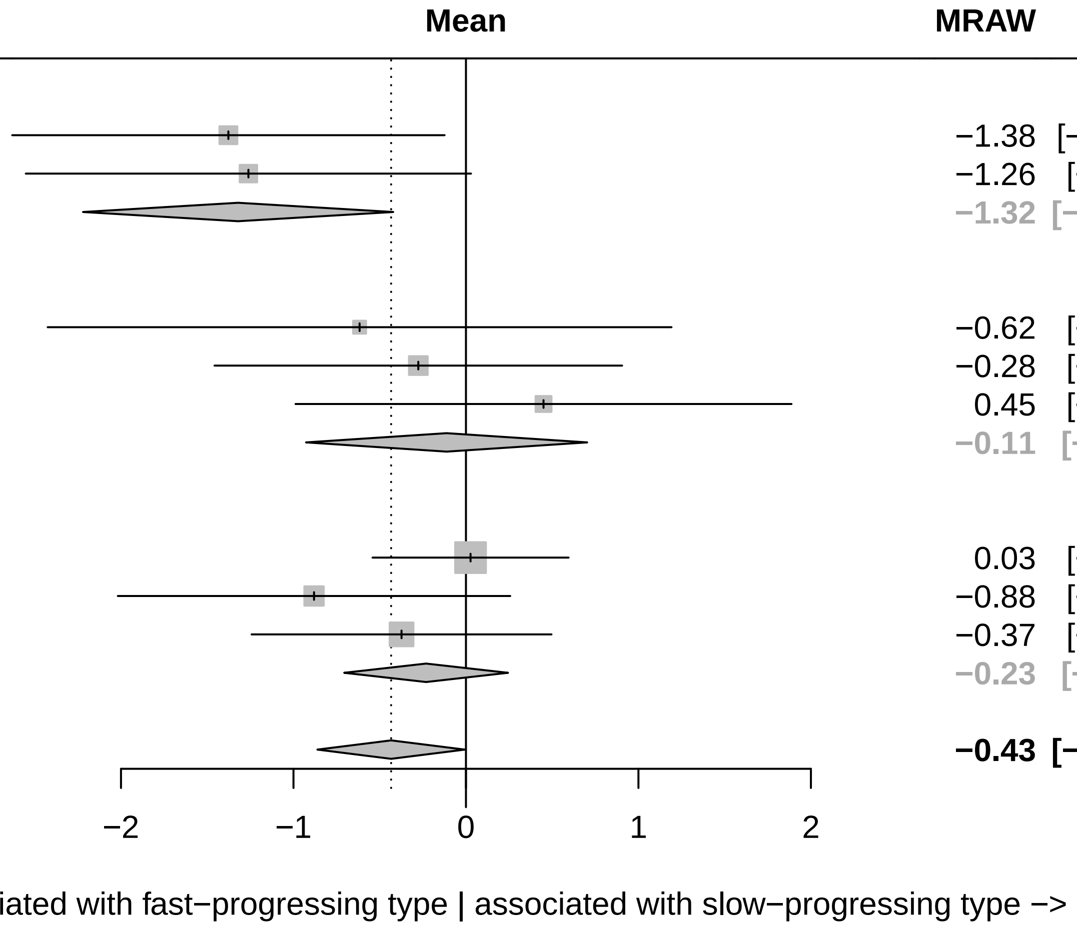

Three−level metaanalysis using random effects to calculate an overall regression coefficient estimate for Sleep across cohorts.
The dashed line indicates the overall mean estimate. The solid line indicates no effect.

### Forest plot for baseline characteristics of symptom domain Overall cognition

| Study | Total | Mean | SD | MRAW | 95%−CI |
|---|---|---|---|---|---|
| **Cohort = LuxPARK** | | | | | |
| FAB | 235 | −3.29 | 10.8757 | −3.29 | [−4.68; −1.90] |
| MoCA | 559 | −3.97 | 13.9238 | −3.97 | [−5.13; −2.82] |
| NMSQ Cognition | 544 | −0.59 | 5.5913 | −0.59 | [−1.06; −0.12] |
| PDQ39 Cognition | 549 | −1.23 | 9.8271 | −1.23 | [−2.05; −0.41] |
| SIQCDE | 232 | −1.85 | 18.8396 | −1.85 | [−4.27; 0.58] |
| UPDRS I Cognition | 558 | −1.92 | 10.6148 | −1.92 | [−2.80; −1.04] |
| **Random effects model** | **2677** | | | **−2.08** | **[−3.17; −0.99]** |
| **Cohort = ICEBERG** | | | | | |
| FAB | 154 | −1.38 | 9.2690 | −1.38 | [−2.85; 0.08] |
| MATTIS | 154 | −1.74 | 11.4295 | −1.74 | [−3.54; 0.07] |
| MMSE | 154 | −1.07 | 6.5563 | −1.07 | [−2.10; −0.03] |
| MoCA | 154 | −1.19 | 10.1073 | −1.19 | [−2.79; 0.40] |
| NMSQ Cognition | 154 | −0.85 | 6.1851 | −0.85 | [−1.82; 0.13] |
| UPDRS I Cognition | 154 | −1.20 | 10.4489 | −1.20 | [−2.85; 0.45] |
| **Random effects model** | **924** | | | **−1.13** | **[−1.66; −0.60]** |
| **Cohort = PPMI** | | | | | |
| MoCA | 409 | −0.59 | 14.1844 | −0.59 | [−1.97; 0.78] |
| UPDRS I Cognition | 409 | −1.27 | 9.1717 | −1.27 | [−2.16; −0.38] |
| **Random effects model** | **818** | | | **−1.07** | **[−1.82; −0.33]** |
| **Random effects model** | **4419** | | | **−1.53** | **[−2.07; −1.00]** |

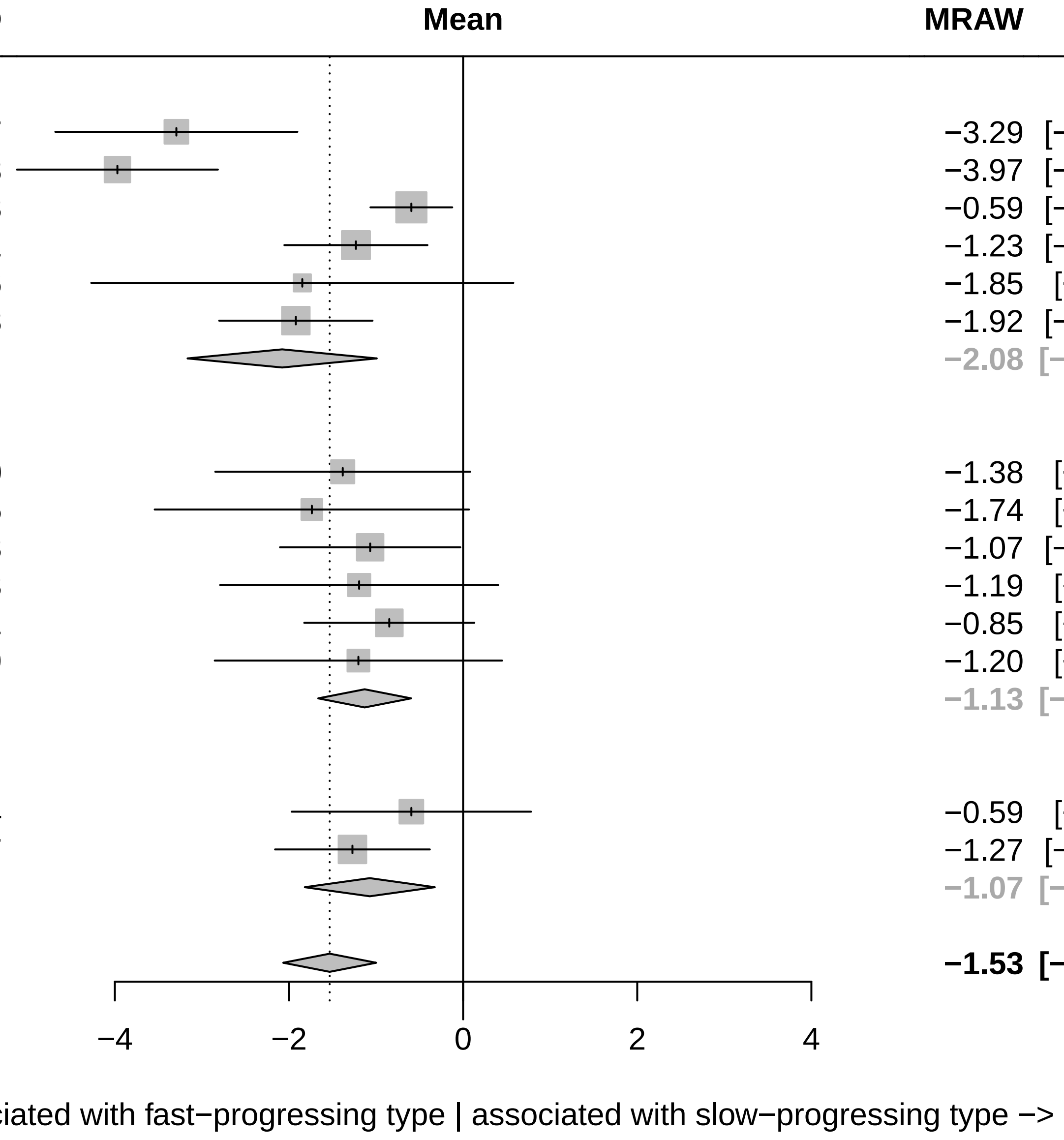

Three−level metaanalysis using random effects to calculate an overall regression coefficient estimate for Overall cognition across cohorts. The dashed line indicates the overall mean estimate. The solid line indicates no effect.

### Forest plot for baseline characteristics of symptom domain Conceptualization

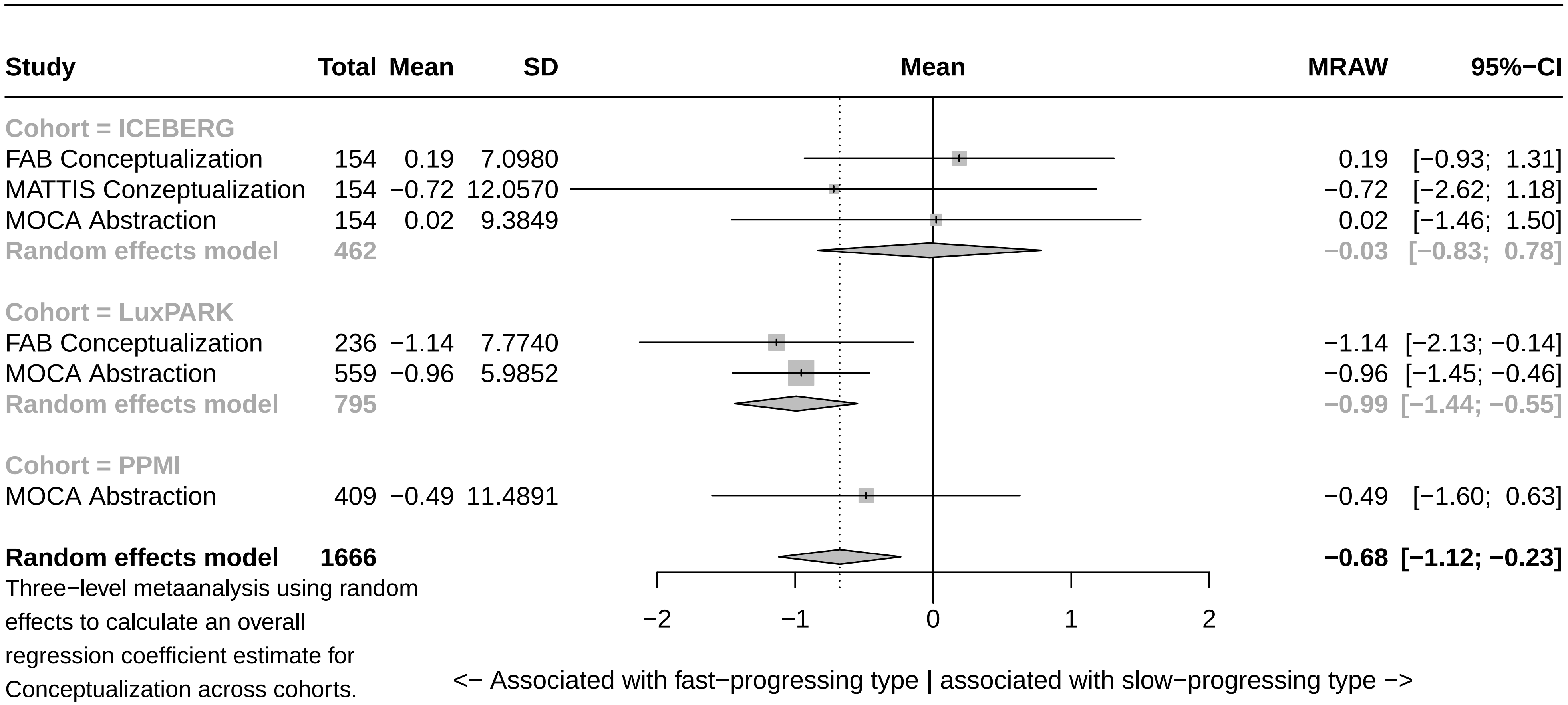

Three−level metaanalysis using random effects to calculate an overall regression coefficient estimate for Conceptualization across cohorts. The dashed line indicates the overall mean estimate. The solid line indicates no effect.

### Forest plot for baseline characteristics of symptom domain Visuo−executive

| Study | Total | Mean | SD | MRAW | 95%−CI |
|---|---|---|---|---|---|
| **Cohort = ICEBERG** | | | | | |
| FAB 3−6 | 154 | −1.18 | 9.2951 | −1.18 | [ −2.65; 0.29] |
| MATTIS Initiation + Construction | 154 | −1.16 | 11.8424 | −1.16 | [ −3.03; 0.71] |
| MMSE Construction | 154 | −2.06 | 15.8255 | −2.06 | [ −4.56; 0.44] |
| MOCA Visuspatial/Executive | 154 | −1.15 | 7.7696 | −1.15 | [ −2.38; 0.08] |
| **Random effects model** | **616** | | | **−1.25** | **[ −2.05; −0.46]** |
| **Cohort = LuxPARK** | | | | | |
| FAB 3−6 | 235 | −3.08 | 10.8871 | −3.08 | [ −4.47; −1.69] |
| Judgement Line Orientation | 214 | −2.53 | 11.8539 | −2.53 | [ −4.12; −0.94] |
| MOCA Visuspatial/Executive | 560 | −2.36 | 9.4592 | −2.36 | [ −3.15; −1.58] |
| Stroop Errors | 208 | −7.66 | 45.0379 | −7.66 | [−13.78; −1.54] |
| Stroop Time | 208 | −3.35 | 15.9332 | −3.35 | [ −5.51; −1.18] |
| Trailmaking A | 549 | −6.75 | 26.6864 | −6.75 | [ −8.98; −4.51] |
| Trailmaking B | 480 | −5.51 | 18.8538 | −5.51 | [ −7.20; −3.83] |
| **Random effects model** | **2454** | | | **−3.94** | **[ −5.30; −2.59]** |
| **Cohort = PPMI** | | | | | |
| Judgement Line Orientation | 408 | −1.19 | 11.5763 | −1.19 | [ −2.31; −0.07] |
| MOCA Visuspatial/Executive | 409 | −0.50 | 12.3462 | −0.50 | [ −1.69; 0.70] |
| Symbol Digit Modalities | 408 | −5.02 | 21.9826 | −5.02 | [ −7.15; −2.89] |
| **Random effects model** | **1225** | | | **−2.10** | **[ −4.71; 0.52]** |
| **Random effects model** | **4295** | | | **−2.75** | **[ −3.77; −1.74]** |

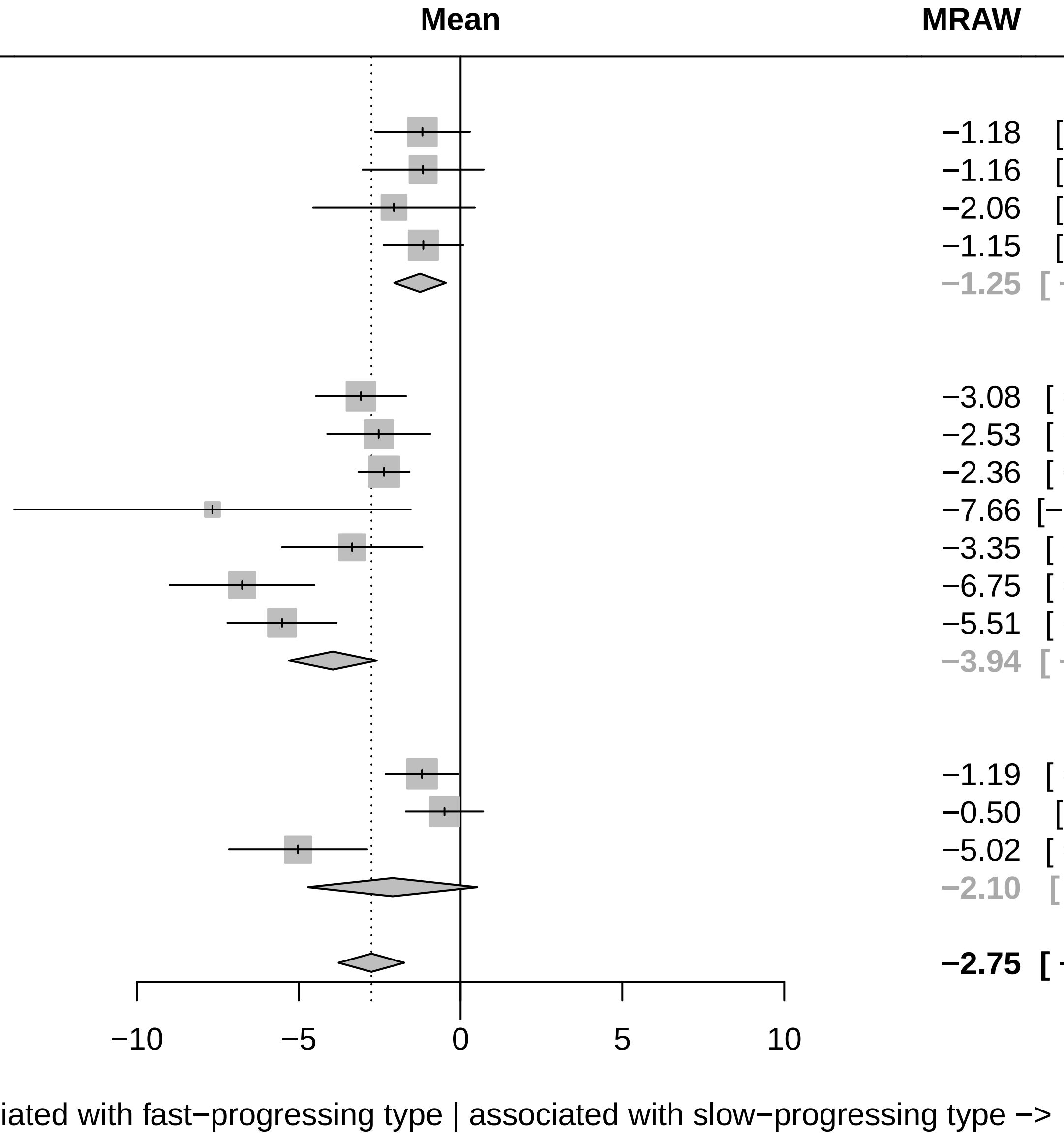

Three−level metaanalysis using random effects to calculate an overall regression coefficient estimate for Visuo−executive across cohorts. The dashed line indicates the overall mean estimate. The solid line indicates no effect.

#### Forest plot for baseline characteristics of symptom domain Language

| Study | Total | Mean | SD | MRAW | 95%−CI |
|---|---|---|---|---|---|
| **Cohort = ICEBERG** | | | | | |
| FAB VFT | 154 | −1.18 | 8.0533 | −1.18 | [−2.45; 0.09] |
| MMSE Language | 154 | 0.02 | 9.3212 | 0.02 | [−1.46; 1.49] |
| MOCA Language + Naming | 154 | −0.82 | 9.2009 | −0.82 | [−2.28; 0.63] |
| VFT phonematic F | 151 | −1.97 | 11.6530 | −1.97 | [−3.83; −0.11] |
| VFT phonematic S | 143 | −1.10 | 11.5611 | −1.10 | [−2.99; 0.80] |
| VFT semantic supermarket | 151 | −2.15 | 14.1365 | −2.15 | [−4.41; 0.10] |
| **Random effects model** | **907** | | | **−1.04** | **[−1.70; −0.38]** |
| **Cohort = LuxPARK** | | | | | |
| FAB VFT | 236 | −1.82 | 8.6299 | −1.82 | [−2.92; −0.72] |
| MOCA Language + Naming | 559 | −2.99 | 14.1846 | −2.99 | [−4.16; −1.81] |
| VFT phonematic F | 548 | −2.90 | 14.7073 | −2.90 | [−4.13; −1.66] |
| VFT phonematic S | 221 | −4.26 | 16.7491 | −4.26 | [−6.47; −2.05] |
| VFT semantic sum | 210 | −3.86 | 14.1298 | −3.86 | [−5.77; −1.94] |
| **Random effects model** | **1774** | | | **−2.89** | **[−3.67; −2.11]** |
| **Cohort = PPMI** | | | | | |
| MOCA Language + Naming | 409 | −0.96 | 16.1720 | −0.96 | [−2.53; 0.60] |
| VFT semantic sum | 408 | −5.86 | 23.6273 | −5.86 | [−8.15; −3.57] |
| **Random effects model** | **817** | | | **−3.34** | **[−8.13; 1.46]** |
| **Random effects model** | **3498** | | | **−2.18** | **[−2.98; −1.37]** |

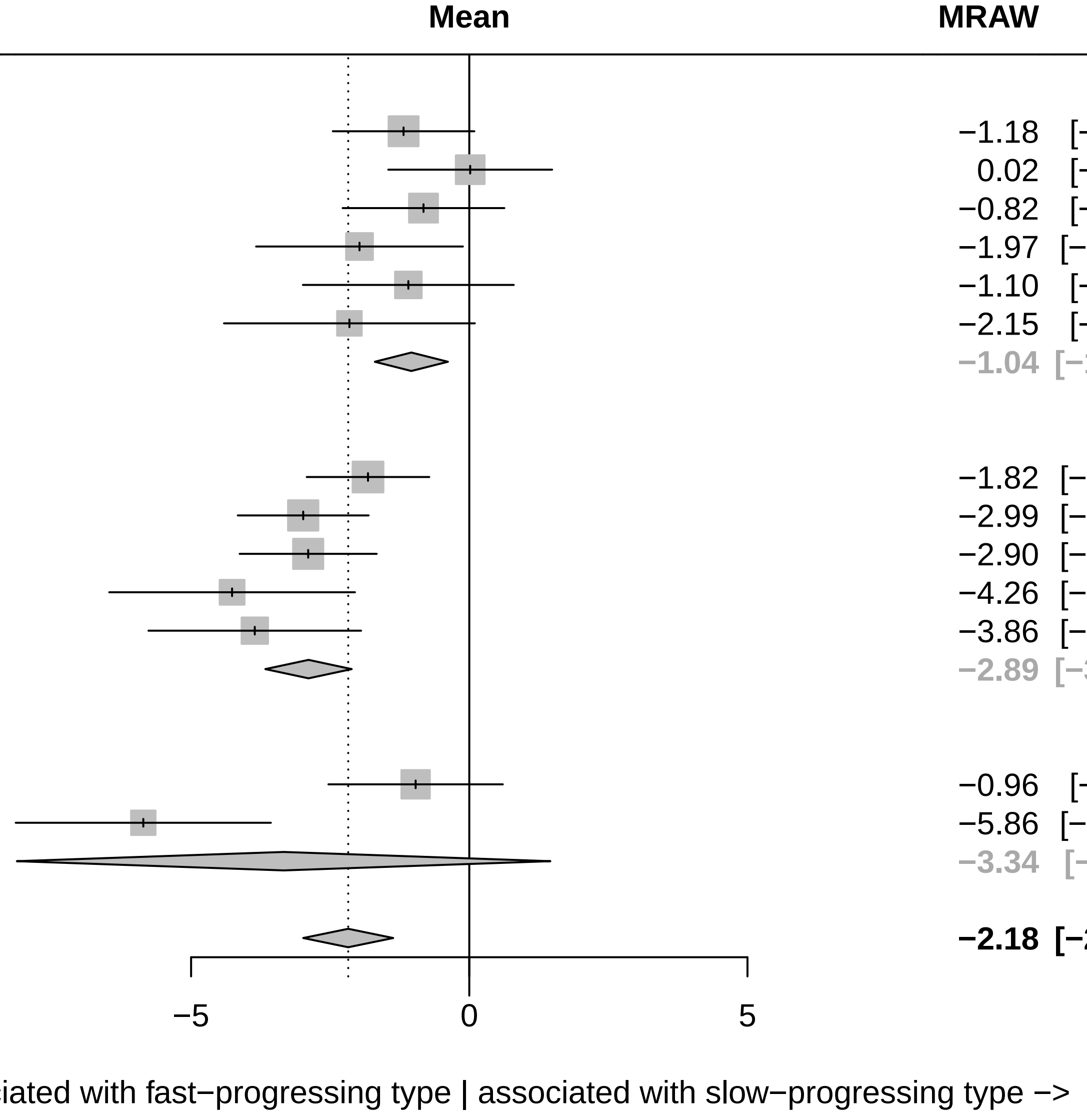

Three−level metaanalysis using random effects to calculate an overall regression coefficient estimate for Language across cohorts.
The dashed line indicates the overall mean estimate. The solid line indicates no effect.

### Forest plot for baseline characteristics of symptom domain Anxiety

| Study | Total | Mean | SD | MRAW | 95%−CI |
|---|---|---|---|---|---|
| **Cohort = ICEBERG** | | | | | |
| HADS anxiety | 154 | 1.37 | 14.0575 | 1.37 | [−0.85; 3.59] |
| NMSQ Anxiety | 152 | −1.03 | 4.7079 | −1.03 | [−1.78; −0.28] |
| UPDRS I Anxiety | 154 | −0.94 | 9.1543 | −0.94 | [−2.38; 0.51] |
| **Random effects model** | **460** | | | **−0.58** | **[−1.65; 0.49]** |
| **Cohort = LuxPARK** | | | | | |
| NMSQ Anxiety | 543 | 0.09 | 5.1173 | 0.09 | [−0.34; 0.52] |
| PDQ39 Anxiety | 550 | −0.79 | 8.5075 | −0.79 | [−1.50; −0.07] |
| UPDRS I Anxiety | 558 | −0.48 | 10.2466 | −0.48 | [−1.33; 0.37] |
| **Random effects model** | **1651** | | | **−0.32** | **[−0.89; 0.25]** |
| **Cohort = PPMI** | | | | | |
| STA | 405 | −0.62 | 13.8380 | −0.62 | [−1.97; 0.73] |
| UPDRS I Anxiety | 409 | 0.10 | 16.6865 | 0.10 | [−1.52; 1.71] |
| **Random effects model** | **814** | | | **−0.33** | **[−1.36; 0.71]** |
| **Random effects model** | **2925** | | | **−0.43** | **[−0.85; −0.00]** |

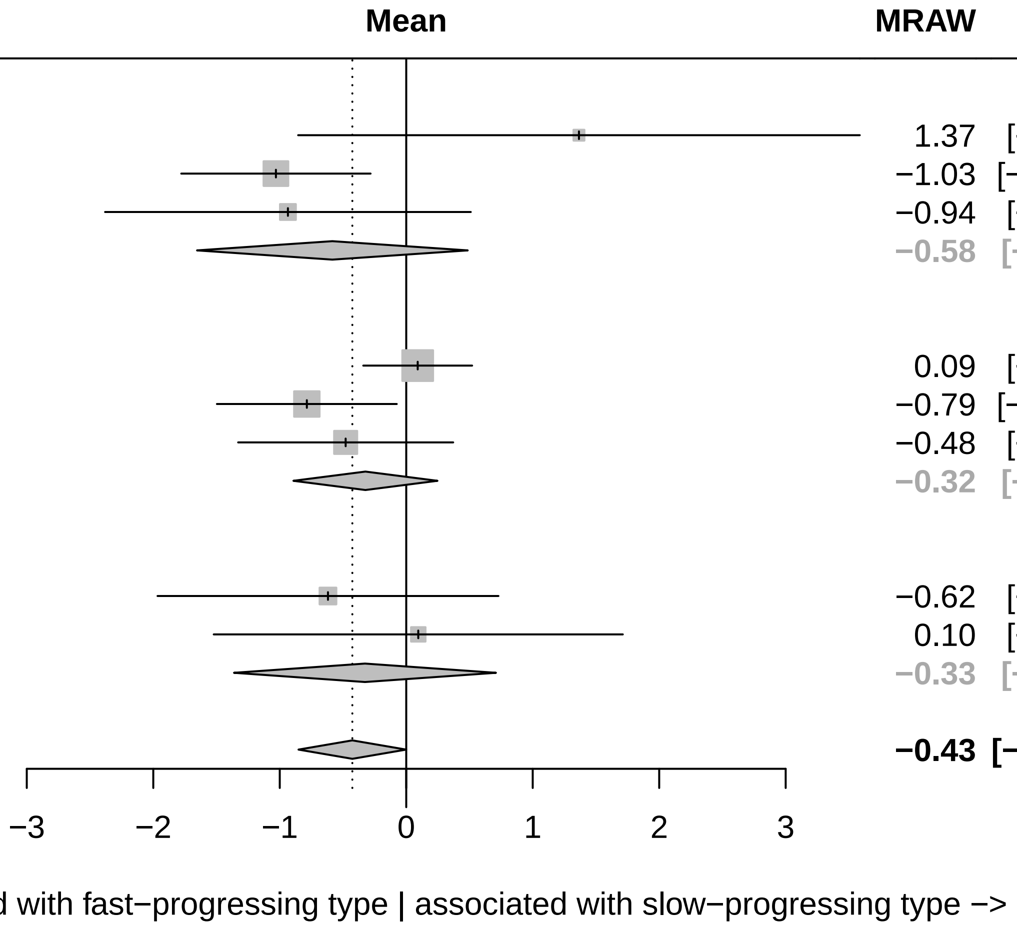

Three−level metaanalysis using random effects to calculate an overall regression coefficient estimate for Anxiety across cohorts.
The dashed line indicates the overall mean estimate. The solid line indicates no effect.

### Forest plot for baseline characteristics of symptom domain Attention

| Study | Total | Mean | SD | MRAW | 95%−CI |
|---|---|---|---|---|---|
| **Cohort = ICEBERG** | | | | | |
| MATTIS Attention | 154 | −2.95 | 12.5929 | −2.95 | [−4.94; −0.96] |
| MMSE Attention | 154 | −1.53 | 9.9091 | −1.53 | [−3.10; 0.03] |
| MOCA Attention | 154 | −1.42 | 8.3050 | −1.42 | [−2.73; −0.11] |
| NMSQ Attention | 152 | −0.59 | 5.0818 | −0.59 | [−1.40; 0.22] |
| **Random effects model** | **614** | | | **−1.34** | **[−2.22; −0.46]** |
| **Cohort = LuxPARK** | | | | | |
| MOCA Attention | 559 | −2.33 | 13.2315 | −2.33 | [−3.42; −1.23] |
| NMSQ Attention | 545 | −0.44 | 4.7993 | −0.44 | [−0.85; −0.04] |
| PDQ39 Attention | 549 | −1.03 | 8.7061 | −1.03 | [−1.76; −0.30] |
| SIQCDE Attention | 233 | 0.11 | 13.0786 | 0.11 | [−1.57; 1.79] |
| Digit Span | 214 | −3.67 | 13.8011 | −3.67 | [−5.52; −1.82] |
| **Random effects model** | **2100** | | | **−1.37** | **[−2.54; −0.20]** |
| **Cohort = PPMI** | | | | | |
| MOCA Attention | 409 | −0.08 | 20.8489 | −0.08 | [−2.10; 1.94] |
| **Random effects model** | **3123** | | | **−1.27** | **[−1.93; −0.62]** |

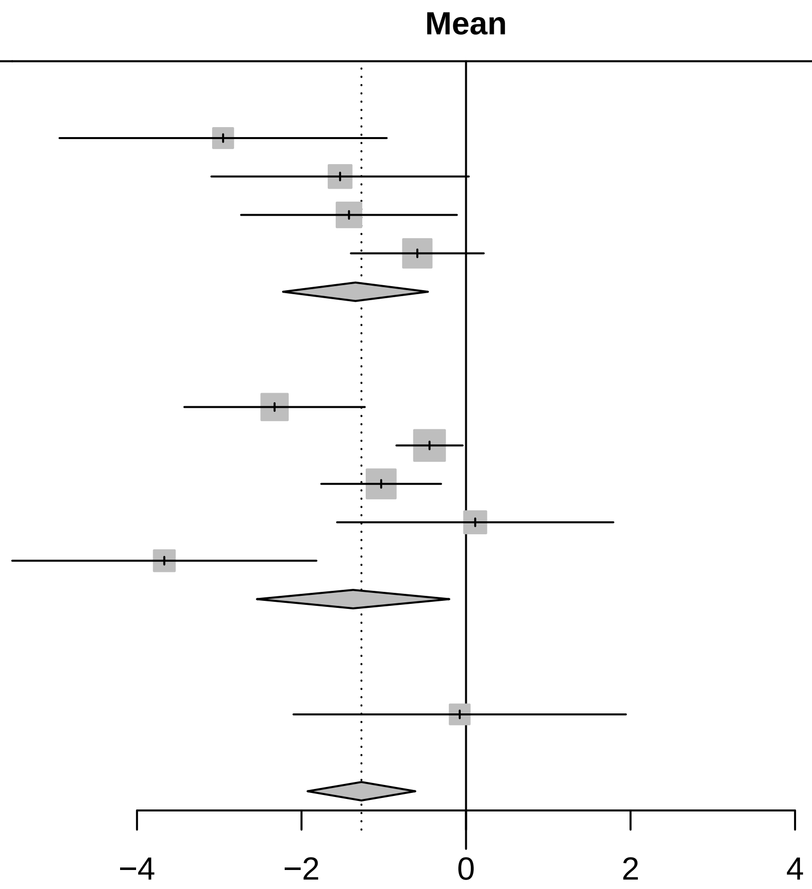

Three−level metaanalysis using random effects to calculate an overall regression coefficient estimate for Attention across cohorts.
The dashed line indicates the overall mean estimate. The solid line indicates no effect.

### Forest plot for baseline characteristics of symptom domain Non motor symptoms

| Study | Total | Mean | SD | MRAW | 95%−CI |
|---|---|---|---|---|---|
| **Cohort = LuxPARK** | | | | | |
| NMSQ | 519 | −1.30 | 12.9026 | −1.30 | [−2.41; −0.19] |
| UPDRS I | 555 | −2.01 | 15.2612 | −2.01 | [−3.28; −0.74] |
| **Random effects model** | **1074** | | | **−1.61** | **[−2.44; −0.77]** |
| **Cohort = ICEBERG** | | | | | |
| NMSQ | 152 | −3.20 | 13.4811 | −3.20 | [−5.35; −1.06] |
| UPDRS I | 154 | −1.88 | 18.1978 | −1.88 | [−4.76; 0.99] |
| **Random effects model** | **306** | | | **−2.73** | **[−4.45; −1.01]** |
| **Cohort = PPMI** | | | | | |
| UPDRS I | 408 | −3.31 | 19.6135 | −3.31 | [−5.21; −1.40] |
| **Random effects model** | **1788** | | | **−2.12** | **[−2.95; −1.28]** |

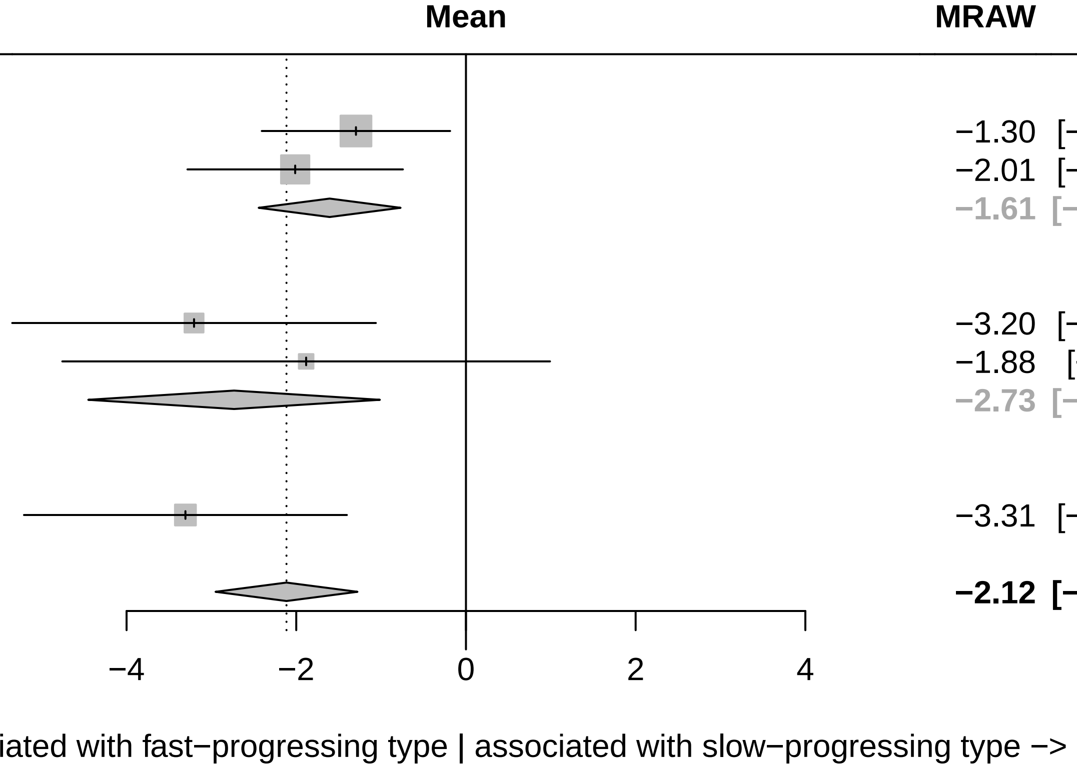

Three−level metaanalysis using random effects to calculate an overall regression coefficient estimate for Non motor symptoms across cohorts. The dashed line indicates the overall mean estimate. The solid line indicates no effect.

### Forest plot for baseline characteristics of symptom domain Autonomic

| Study | Total | Mean | SD | MRAW | 95%−CI |
|---|---|---|---|---|---|
| **Cohort = LuxPARK** | | | | | |
| NMSQ Autonomic | 525 | −0.63 | 11.6412 | −0.63 | [−1.62; 0.37] |
| SCOPA−AUT | 547 | −1.48 | 13.7072 | −1.48 | [−2.63; −0.33] |
| UPDRS I Autonomic | 558 | −1.53 | 14.0669 | −1.53 | [−2.70; −0.37] |
| **Random effects model** | **1630** | | | **−1.15** | **[−1.78; −0.52]** |
| **Cohort = ICEBERG** | | | | | |
| NMSQ Autonomic | 154 | −0.86 | 9.9677 | −0.86 | [−2.43; 0.72] |
| SCOPA−AUT | 153 | 1.92 | 12.9248 | 1.92 | [−0.13; 3.97] |
| UPDRS I Autonomic | 154 | 1.34 | 13.4833 | 1.34 | [−0.79; 3.47] |
| **Random effects model** | **461** | | | **0.69** | **[−1.08; 2.45]** |
| **Cohort = PPMI** | | | | | |
| SCOPA−AUT | 398 | −4.07 | 17.7790 | −4.07 | [−5.82; −2.33] |
| UPDRS I Autonomic | 409 | −2.16 | 14.4778 | −2.16 | [−3.56; −0.75] |
| **Random effects model** | **807** | | | **−3.04** | **[−4.92; −1.17]** |
| **Random effects model** | **2898** | | | **−1.02** | **[−2.22; 0.18]** |

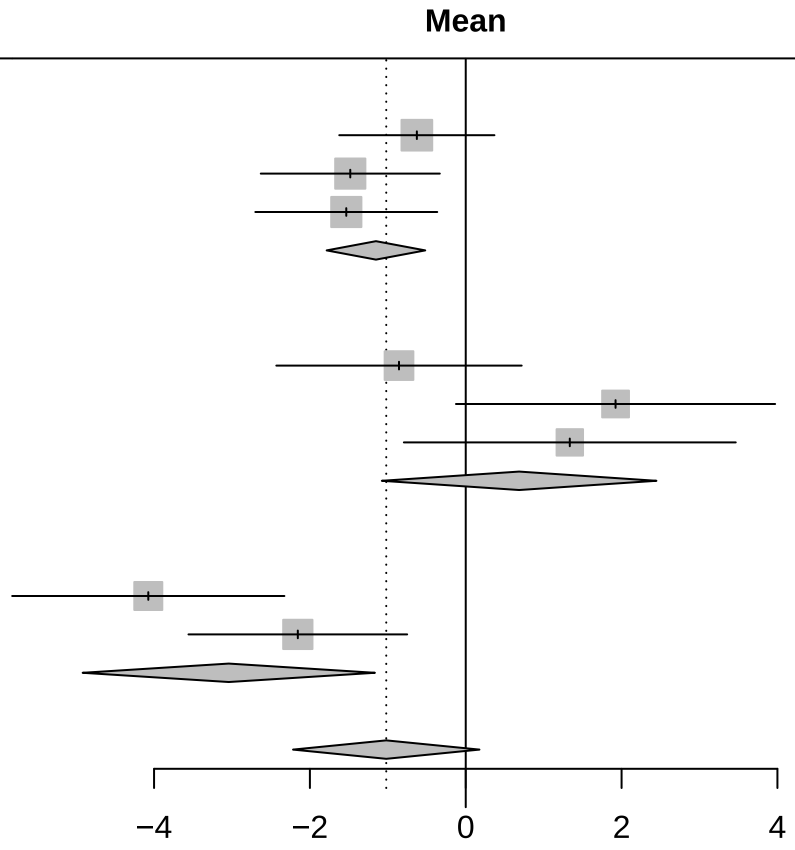

<− Associated with fast−progressing type | associated with slow−progressing type −>

Three−level metaanalysis using random effects to calculate an overall regression coefficient estimate for Autonomic across cohorts.
The dashed line indicates the overall mean estimate. The solid line indicates no effect.

### Forest plot for baseline characteristics of symptom domain Hallucinations

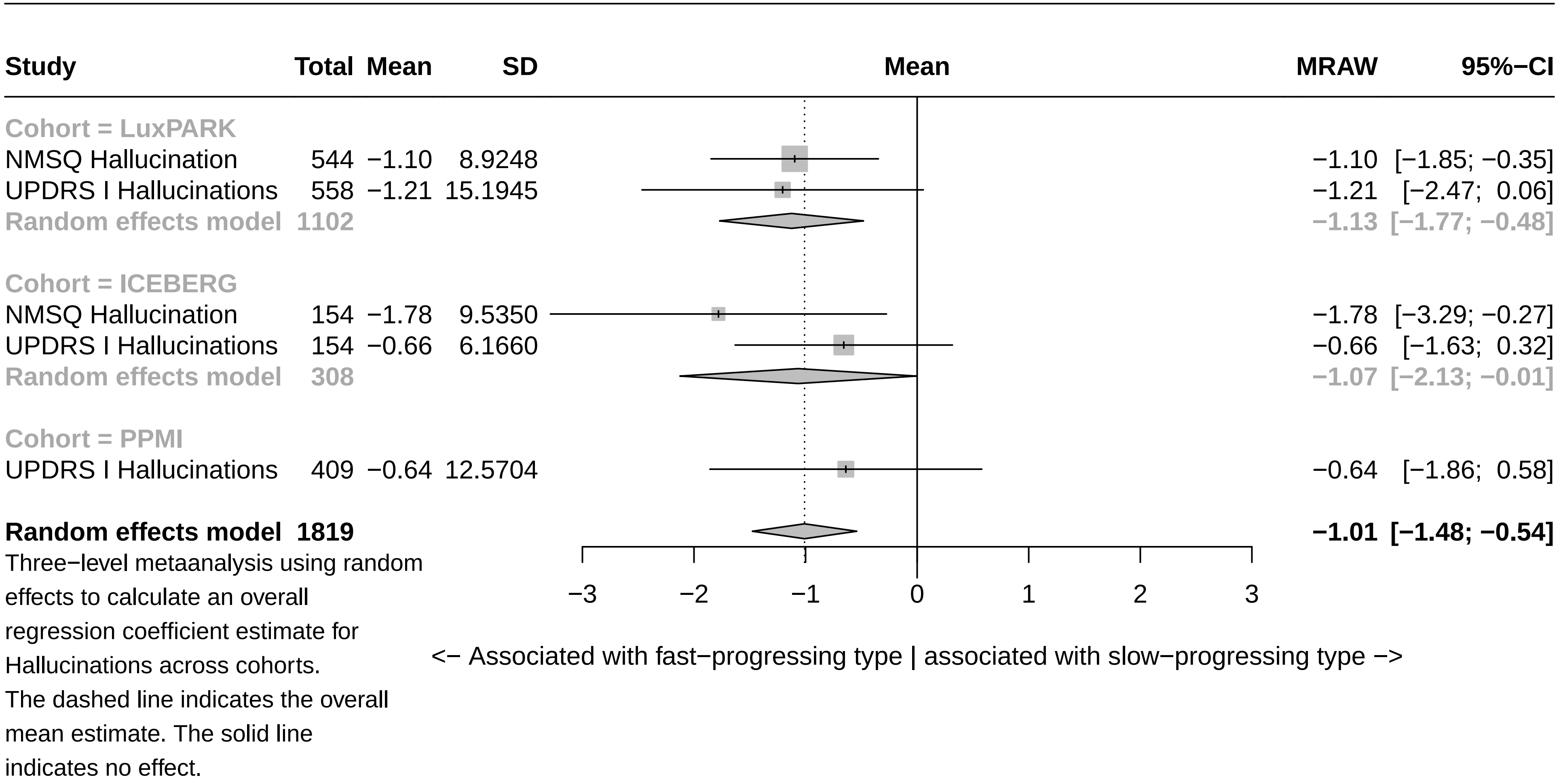

### Forest plot for baseline characteristics of symptom domain Pain

| Study | Total | Mean | SD | MRAW | 95%−CI |
|---|---|---|---|---|---|
| **Cohort = LuxPARK** | | | | | |
| NMSQ Pain | 546 | 0.09 | 5.3917 | 0.09 | [−0.36; 0.54] |
| PDQ39 Pain | 547 | −0.18 | 9.1475 | −0.18 | [−0.95; 0.59] |
| UPDRS I Pain | 558 | −0.46 | 8.5352 | −0.46 | [−1.17; 0.25] |
| **Random effects model** | **1651** | | | **−0.10** | **[−0.45; 0.26]** |
| **Cohort = ICEBERG** | | | | | |
| NMSQ Pain | 152 | −0.59 | 4.6244 | −0.59 | [−1.32; 0.15] |
| UPDRS I Pain | 154 | −2.27 | 10.9099 | −2.27 | [−3.99; −0.55] |
| **Random effects model** | **306** | | | **−1.24** | **[−2.85; 0.36]** |
| **Cohort = PPMI** | | | | | |
| UPDRS I Pain | 408 | −0.43 | 12.3475 | −0.43 | [−1.63; 0.77] |
| **Random effects model** | **2365** | | | **−0.34** | **[−0.72; 0.04]** |

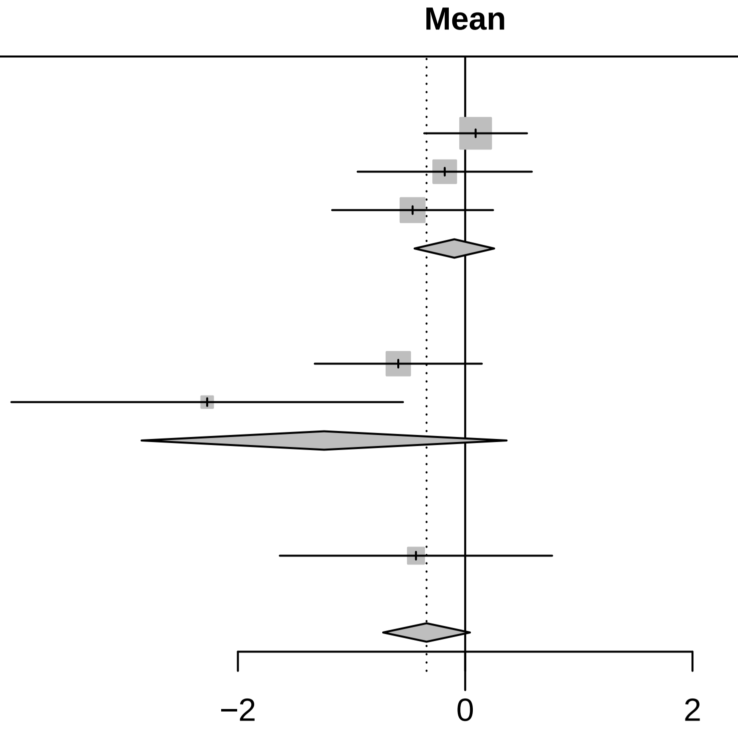

<− Associated with fast−progressing type | associated with slow−progressing type −>

Three−level metaanalysis using random effects to calculate an overall regression coefficient estimate for Pain across cohorts.
The dashed line indicates the overall mean estimate. The solid line indicates no effect.

### Forest plot for baseline characteristics of symptom domain RBD

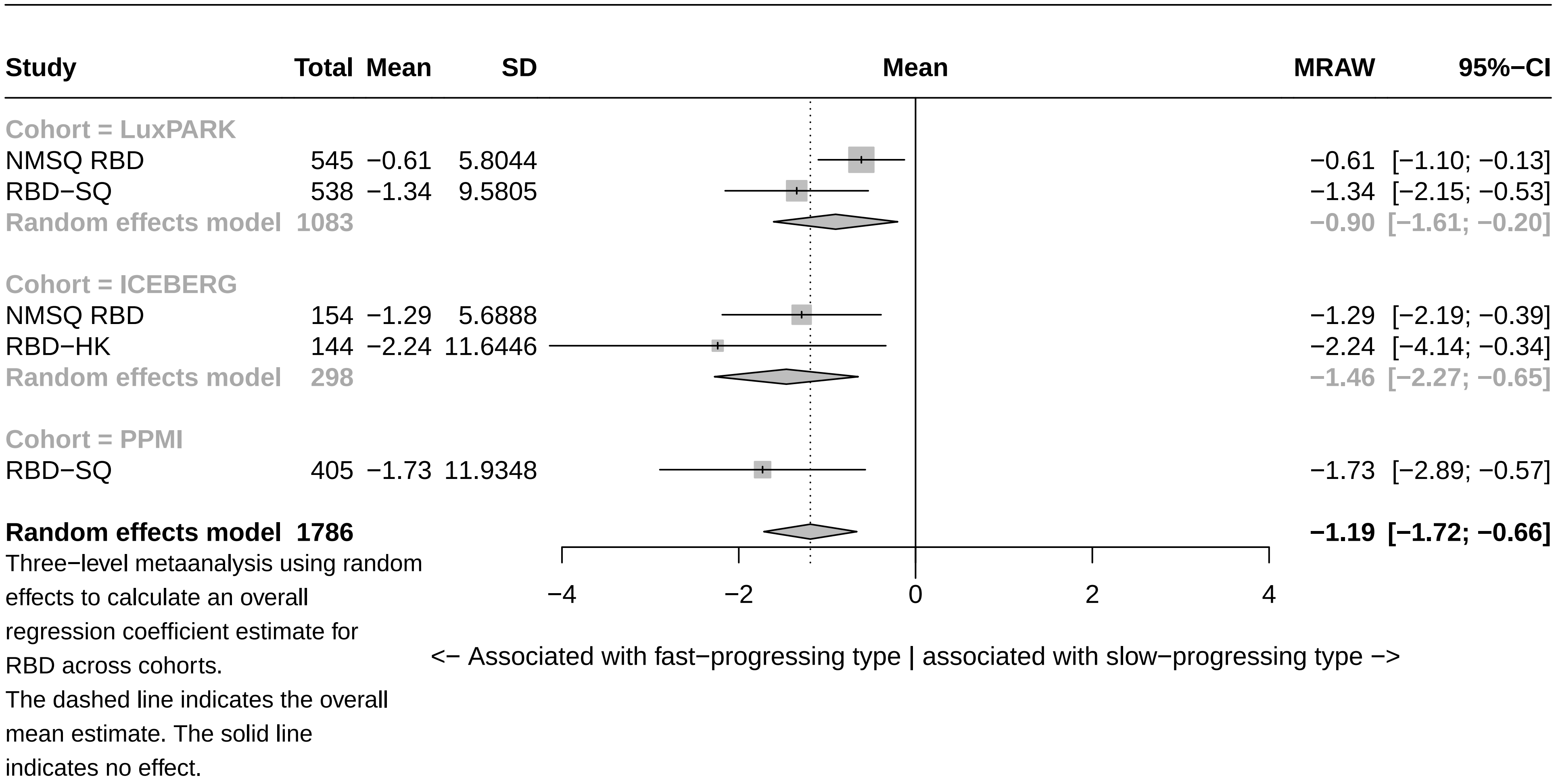

#### Forest plot for baseline characteristics of symptom domain Smell

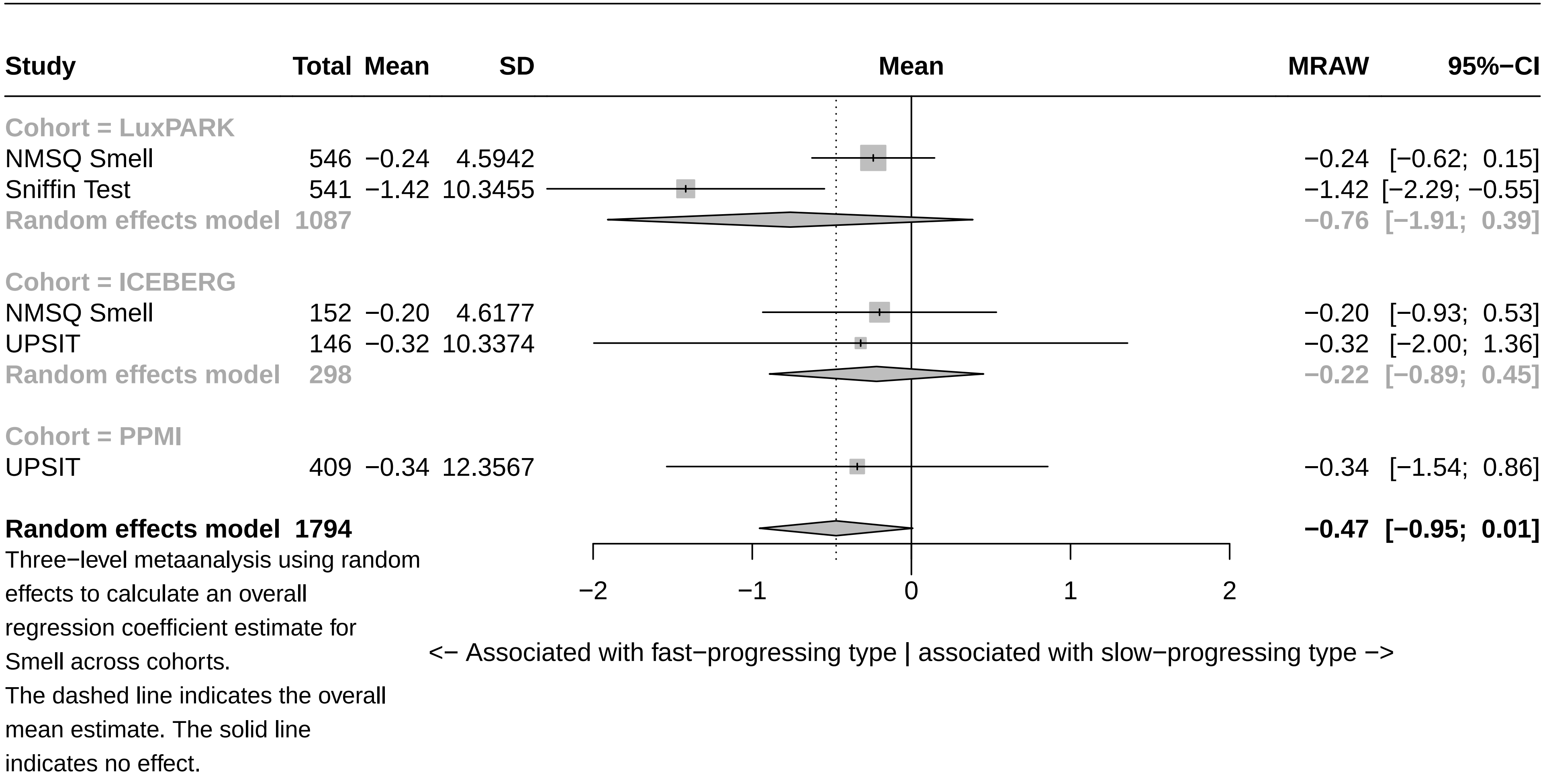

### Forest plot for baseline characteristics of symptom domain Motor symptoms

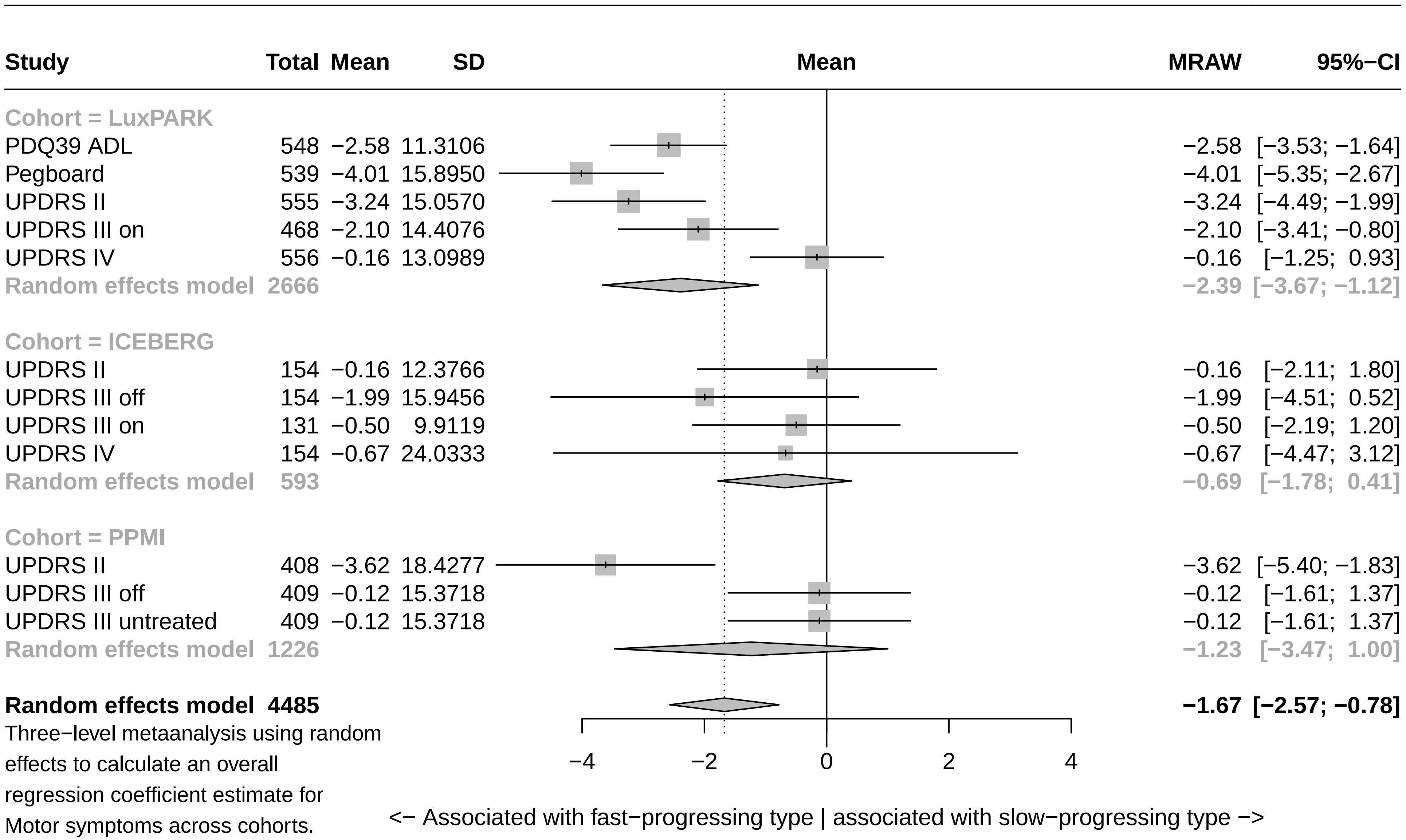

Three−level metaanalysis using random effects to calculate an overall regression coefficient estimate for Motor symptoms across cohorts. The dashed line indicates the overall mean estimate. The solid line indicates no effect.

### Forest plot for baseline characteristics of symptom domain Impulsivity

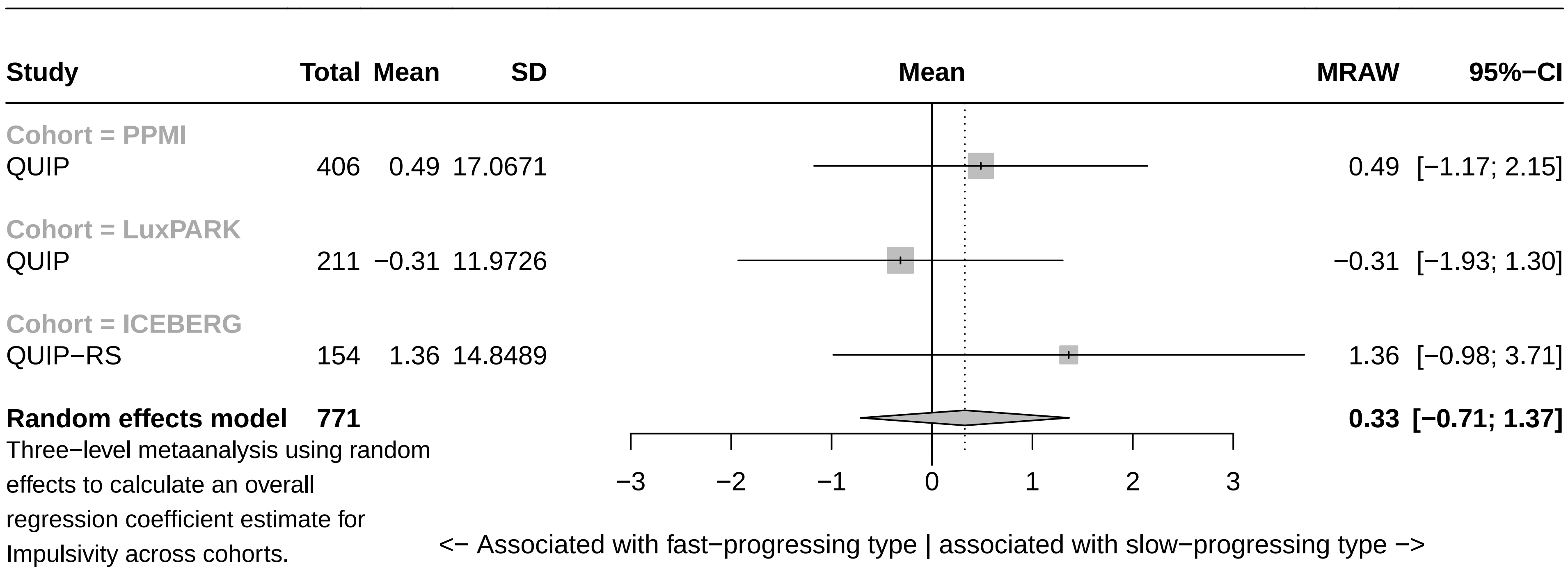

Three−level metaanalysis using random effects to calculate an overall regression coefficient estimate for Impulsivity across cohorts.
The dashed line indicates the overall mean estimate. The solid line indicates no effect.

### Forest plot for baseline characteristics of symptom domain Tremor

| Study | Total | Mean | SD | MRAW | 95%−CI |
|---|---|---|---|---|---|
| **Cohort = ICEBERG** | | | | | |
| TD off | 154 | −1.40 | 10.3429 | −1.40 | [−3.04; 0.23] |
| TD on | 131 | −1.16 | 9.1063 | −1.16 | [−2.71; 0.40] |
| **Random effects model** | **285** | | | **−1.27** | **[−2.40; −0.14]** |
| **Cohort = PPMI** | | | | | |
| TD off | 408 | −0.77 | 14.8601 | −0.77 | [−2.21; 0.68] |
| **Cohort = LuxPARK** | | | | | |
| TD on | 470 | 0.72 | 12.3080 | 0.72 | [−0.39; 1.83] |
| **Random effects model** | **1163** | | | **−0.54** | **[−1.57; 0.50]** |

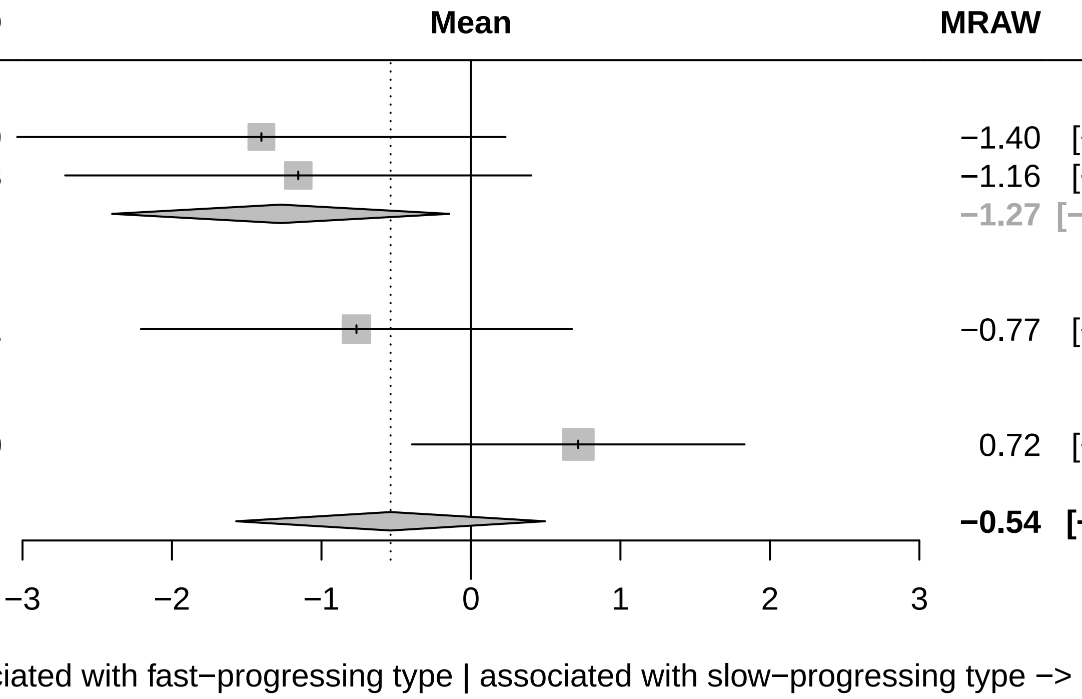

Three−level metaanalysis using random effects to calculate an overall regression coefficient estimate for Tremor across cohorts.
The dashed line indicates the overall mean estimate. The solid line indicates no effect.

### Forest plot for baseline characteristics of symptom domain Fatigue

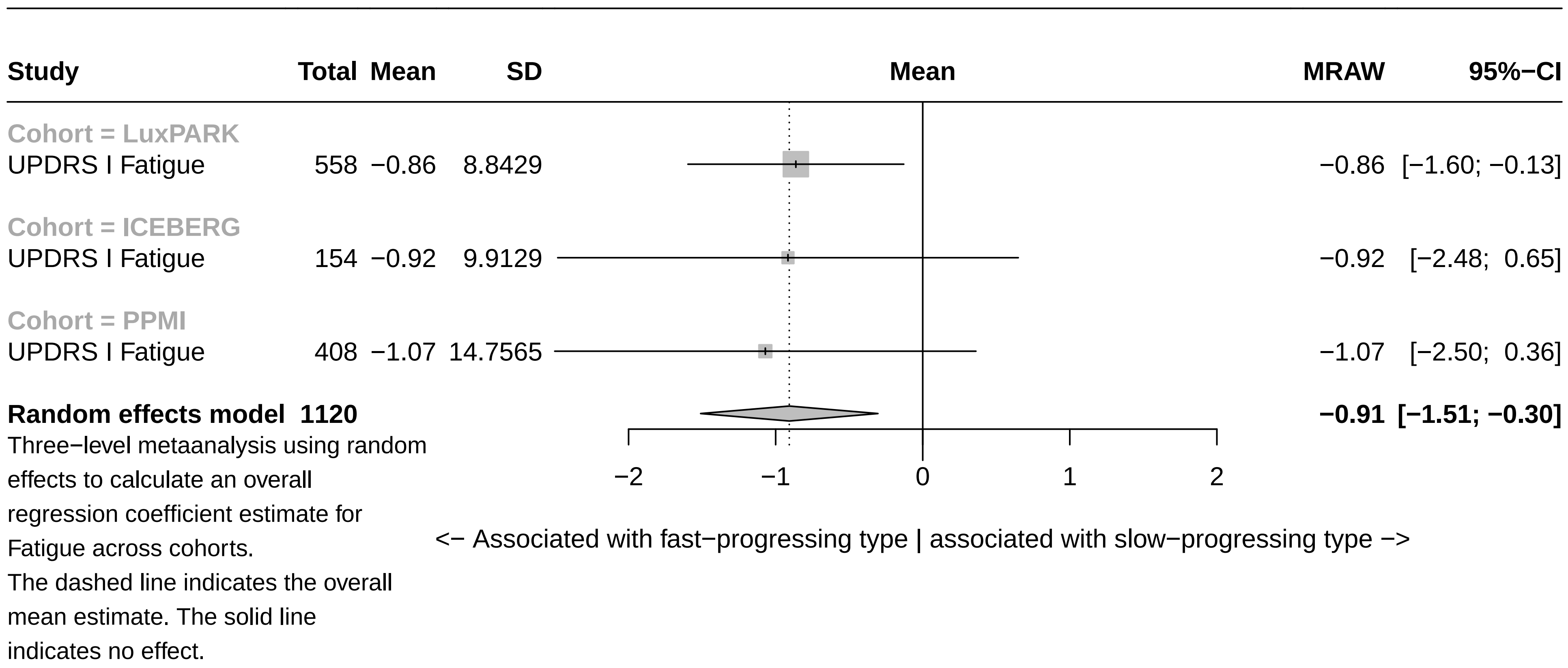

### Forest plots for symptom domain baseline associations (cross-cohort validation)

### Forest plot for baseline characteristics of symptom domain Axial & PIGD (validation)

| Study | Total | Mean | SD | MRAW | 95%−CI |
|---|---|---|---|---|---|
| **Cohort = ICEBERG (validation)** | | | | | |
| UPDRS III axial off | 154 | −0.98 | 15.4066 | −0.98 | [ −3.41; 1.46] |
| UPDRS III axial on | 131 | 0.24 | 13.1173 | 0.24 | [ −2.01; 2.48] |
| GABS Examination | 153 | −0.80 | 16.1203 | −0.80 | [ −3.36; 1.75] |
| GABS Questionaire | 153 | −1.21 | 15.6229 | −1.21 | [ −3.69; 1.26] |
| PIGD off | 154 | −0.27 | 13.3111 | −0.27 | [ −2.37; 1.83] |
| PIGD on | 131 | 0.41 | 12.9408 | 0.41 | [ −1.80; 2.63] |
| **Random effects model** | **876** | | | **−0.37** | **[ −1.32; 0.57]** |
| **Cohort = LuxPARK (validation)** | | | | | |
| UPDRS III axial on | 470 | −3.77 | 16.9217 | −3.77 | [ −5.30; −2.24] |
| FOGAC | 128 | −5.80 | 27.1058 | −5.80 | [−10.50; −1.11] |
| FOGQ | 201 | −2.84 | 15.0671 | −2.84 | [ −4.92; −0.75] |
| NFOGQ | 36 | −5.66 | 15.1159 | −5.66 | [−10.60; −0.72] |
| PDQ39 Mobility | 545 | −2.20 | 12.8964 | −2.20 | [ −3.28; −1.12] |
| PIGD on | 468 | −2.78 | 16.2694 | −2.78 | [ −4.25; −1.31] |
| TuG | 140 | −1.15 | 25.7075 | −1.15 | [ −5.41; 3.11] |
| **Random effects model** | **1988** | | | **−2.87** | **[ −3.62; −2.12]** |
| **Random effects model** | **2864** | | | **−1.83** | **[ −2.70; −0.95]** |

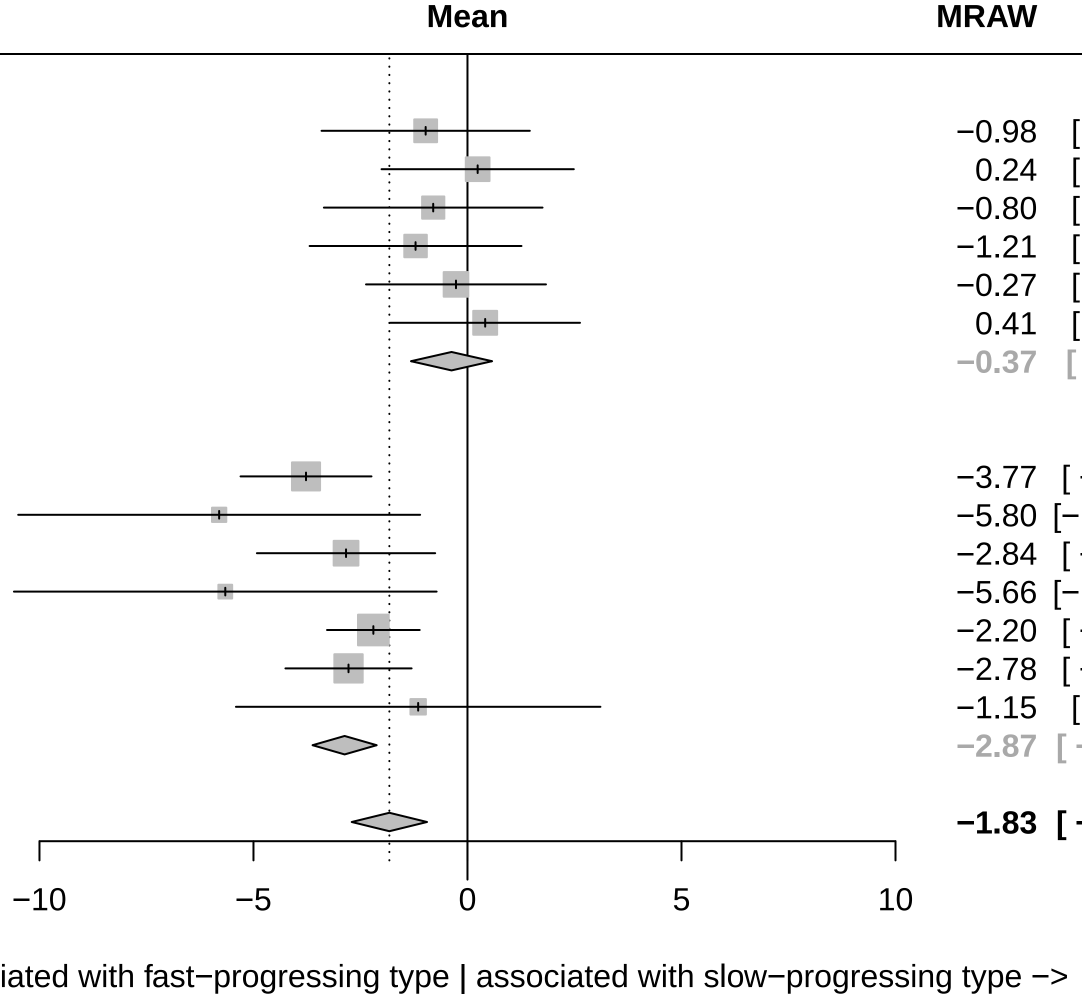

Three−level metaanalysis using random effects to calculate an overall regression coefficient estimate for Axial & PIGD across cohorts. The dashed line indicates the overall mean estimate. The solid line indicates no effect.

### Forest plot for baseline characteristics of symptom domain Depression (validation)

| Study | Total | Mean | SD | MRAW | 95%−CI |
|---|---|---|---|---|---|
| **Cohort = LuxPARK (validation)** | | | | | |
| BDI | 545 | −1.33 | 19.7130 | −1.33 | [−2.98; 0.33] |
| NMSQ Depression | 543 | −0.28 | 6.9264 | −0.28 | [−0.86; 0.30] |
| PDQ39 Depression | 548 | −0.40 | 12.6264 | −0.40 | [−1.46; 0.66] |
| UPDRS I Depression | 558 | −0.27 | 11.7509 | −0.27 | [−1.24; 0.71] |
| **Random effects model** | **2194** | | | **−0.37** | **[−0.81; 0.06]** |
| **Cohort = ICEBERG (validation)** | | | | | |
| HADS depression | 154 | −1.05 | 14.8282 | −1.05 | [−3.39; 1.29] |
| NMSQ Depression | 154 | −0.76 | 7.8135 | −0.76 | [−1.99; 0.48] |
| UPDRS I Depression | 154 | −1.00 | 9.8141 | −1.00 | [−2.55; 0.55] |
| **Random effects model** | **462** | | | **−0.88** | **[−1.77; 0.01]** |
| **Random effects model** | **2656** | | | **−0.47** | **[−0.86; −0.08]** |

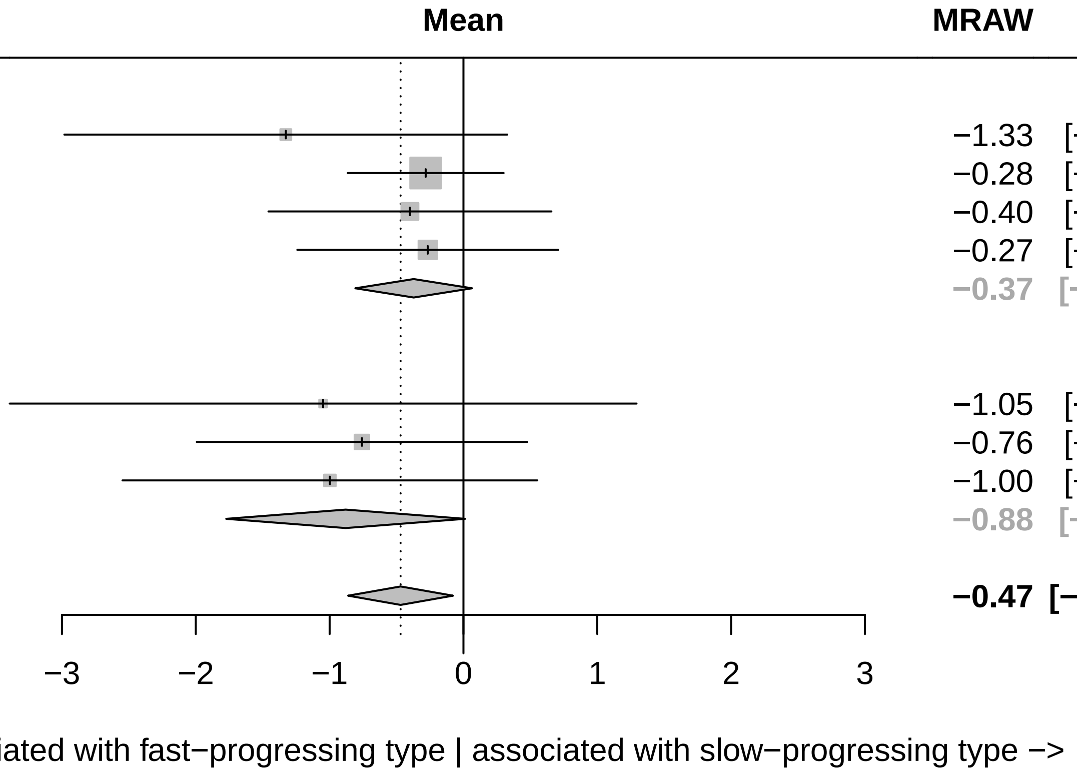

Three−level metaanalysis using random effects to calculate an overall regression coefficient estimate for Depression across cohorts.
The dashed line indicates the overall mean estimate. The solid line indicates no effect.

### Forest plot for baseline characteristics of symptom domain Memory (validation)

| Study | Total | Mean | SD | MRAW | 95%−CI |
|---|---|---|---|---|---|
| **Cohort = LuxPARK (validation)** | | | | | |
| CERAD Words DR | 212 | −0.89 | 11.3786 | −0.89 | [−2.42; 0.65] |
| CERAD Words IR | 216 | −1.80 | 15.0581 | −1.80 | [−3.81; 0.21] |
| CERAD Words Recognition | 217 | 0.70 | 22.9648 | 0.70 | [−2.36; 3.75] |
| MOCA Orientation + Memory | 559 | −3.07 | 14.2284 | −3.07 | [−4.25; −1.89] |
| NMSQ Memory | 545 | −0.59 | 5.4772 | −0.59 | [−1.05; −0.13] |
| PDQ39 Memory | 549 | −1.08 | 10.1624 | −1.08 | [−1.93; −0.23] |
| SIQCDE Memory | 234 | −4.86 | 25.5950 | −4.86 | [−8.14; −1.58] |
| **Random effects model** | **2532** | | | **−1.49** | **[−2.46; −0.52]** |
| **Cohort = ICEBERG (validation)** | | | | | |
| MATTIS Memory | 154 | −0.02 | 12.6948 | −0.02 | [−2.03; 1.98] |
| MMSE Memory | 154 | −0.64 | 9.1126 | −0.64 | [−2.08; 0.80] |
| MOCA Orientation + Memory | 154 | 2.03 | 17.0214 | 2.03 | [−0.66; 4.72] |
| NMSQ Memory | 152 | −0.96 | 6.6616 | −0.96 | [−2.01; 0.10] |
| **Random effects model** | **614** | | | **−0.48** | **[−1.26; 0.31]** |
| **Random effects model** | **3146** | | | **−1.04** | **[−1.76; −0.33]** |

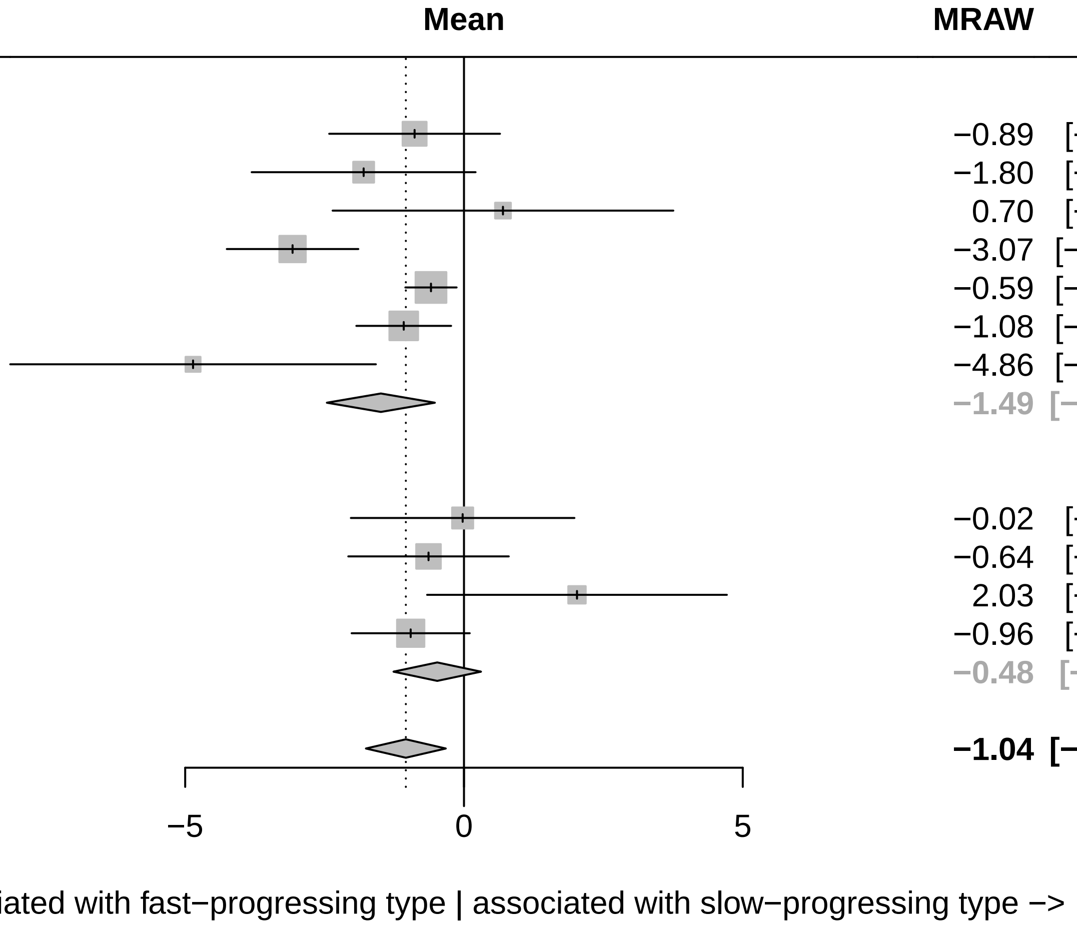

Three−level metaanalysis using random effects to calculate an overall regression coefficient estimate for Memory across cohorts.
The dashed line indicates the overall mean estimate. The solid line indicates no effect.

### Forest plot for baseline characteristics of symptom domain Overall severity (validation)

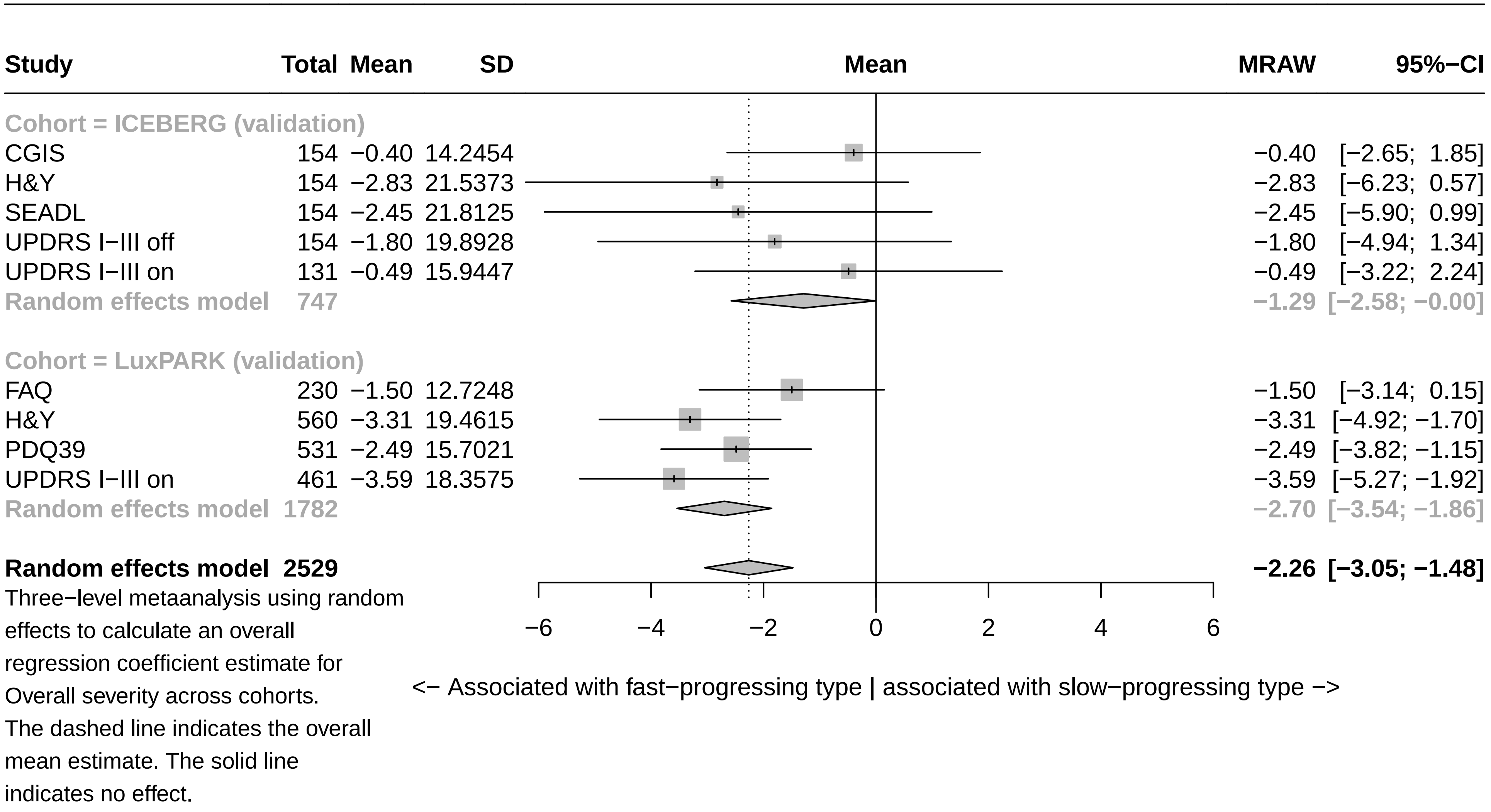

### Forest plot for baseline characteristics of symptom domain Apathy (validation)

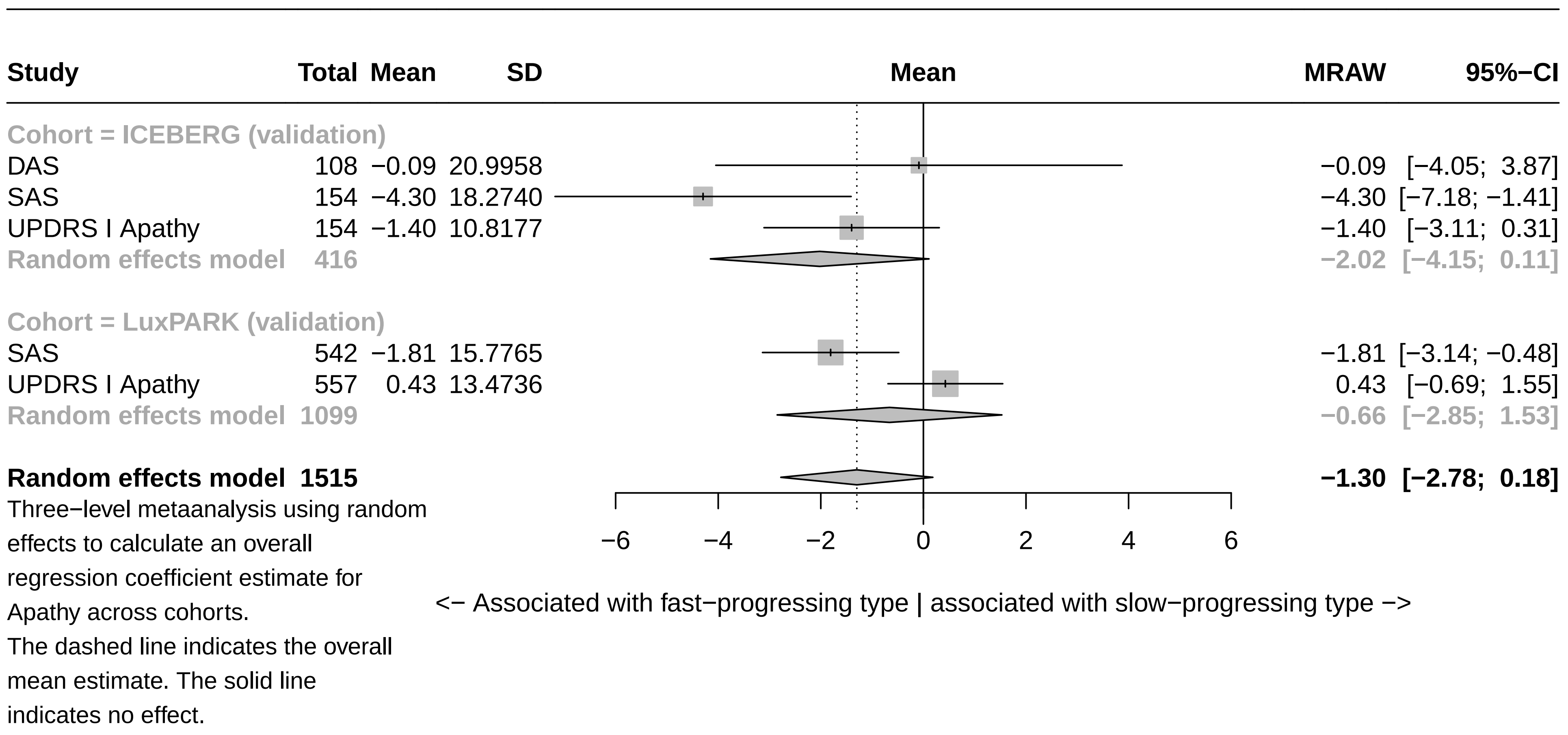

### Forest plot for baseline characteristics of symptom domain Sleep (validation)

| Study | Total | Mean | SD | MRAW | 95%−CI |
|---|---|---|---|---|---|
| **Cohort = ICEBERG (validation)** | | | | | |
| ESS | 143 | 1.09 | 15.6446 | 1.09 | [−1.48; 3.65] |
| NMSQ Sleep | 154 | 0.33 | 10.1584 | 0.33 | [−1.27; 1.94] |
| UPDRS I Sleep | 154 | −0.19 | 12.3587 | −0.19 | [−2.14; 1.77] |
| **Random effects model** | **451** | | | **0.31** | **[−0.81; 1.42]** |
| **Cohort = LuxPARK (validation)** | | | | | |
| NMSQ Sleep | 545 | 0.11 | 8.0388 | 0.11 | [−0.56; 0.79] |
| PDSS | 541 | −0.99 | 15.8621 | −0.99 | [−2.33; 0.34] |
| UPDRS I Sleep | 558 | −0.87 | 12.3135 | −0.87 | [−1.90; 0.15] |
| **Random effects model** | **1644** | | | **−0.45** | **[−1.22; 0.32]** |
| **Random effects model** | **2095** | | | **−0.23** | **[−0.78; 0.33]** |

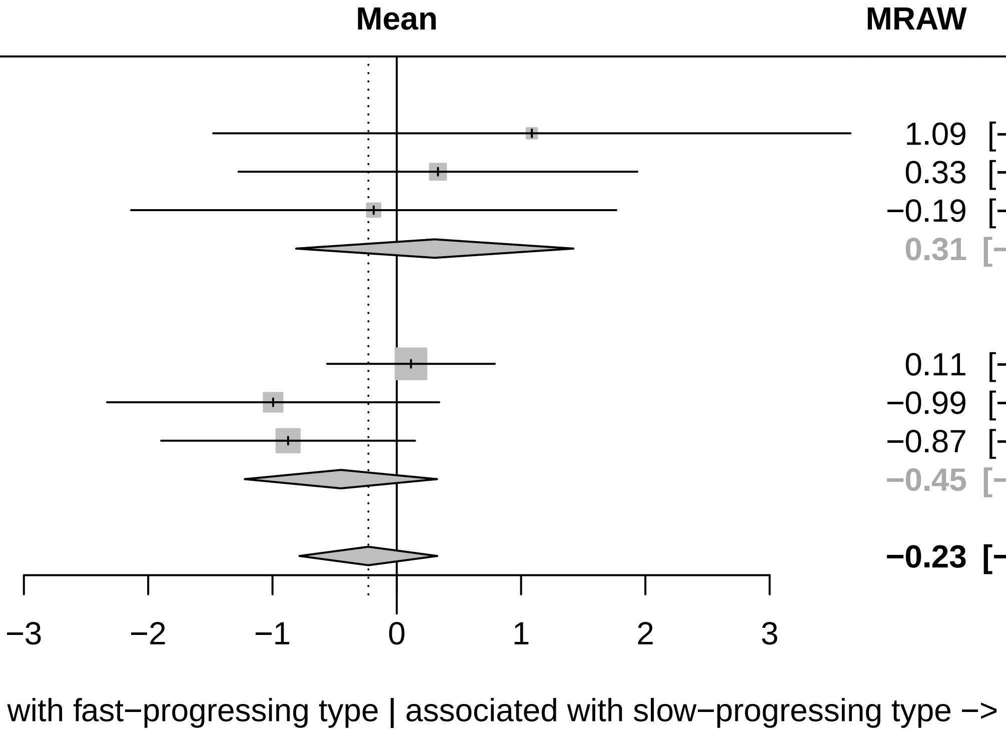

Three−level metaanalysis using random effects to calculate an overall regression coefficient estimate for Sleep across cohorts.
The dashed line indicates the overall mean estimate. The solid line indicates no effect.

### Forest plot for baseline characteristics of symptom domain Overall cognition (validation)

| Study | Total | Mean | SD | MRAW | 95%−CI |
|---|---|---|---|---|---|
| **Cohort = ICEBERG (validation)** | | | | | |
| FAB | 154 | −0.58 | 12.0418 | −0.58 | [−2.48; 1.33] |
| MATTIS | 154 | −1.71 | 14.6102 | −1.71 | [−4.02; 0.59] |
| MMSE | 154 | −1.60 | 8.5313 | −1.60 | [−2.95; −0.25] |
| MoCA | 154 | 0.07 | 13.7346 | 0.07 | [−2.10; 2.24] |
| NMSQ Cognition | 154 | −0.98 | 8.2256 | −0.98 | [−2.28; 0.32] |
| UPDRS I Cognition | 154 | −0.97 | 13.8921 | −0.97 | [−3.16; 1.23] |
| **Random effects model** | **924** | | | **−1.05** | **[−1.75; −0.35]** |
| **Cohort = LuxPARK (validation)** | | | | | |
| FAB | 235 | −1.92 | 11.2959 | −1.92 | [−3.36; −0.47] |
| MoCA | 559 | −3.77 | 14.7988 | −3.77 | [−5.00; −2.55] |
| NMSQ Cognition | 544 | −0.72 | 6.6093 | −0.72 | [−1.28; −0.17] |
| PDQ39 Cognition | 549 | −1.45 | 11.4380 | −1.45 | [−2.41; −0.50] |
| SIQCDE | 232 | −4.61 | 25.8950 | −4.61 | [−7.95; −1.28] |
| UPDRS I Cognition | 558 | −1.64 | 12.1066 | −1.64 | [−2.64; −0.64] |
| **Random effects model** | **2677** | | | **−2.02** | **[−3.03; −1.00]** |
| **Random effects model** | **3601** | | | **−1.55** | **[−2.16; −0.93]** |

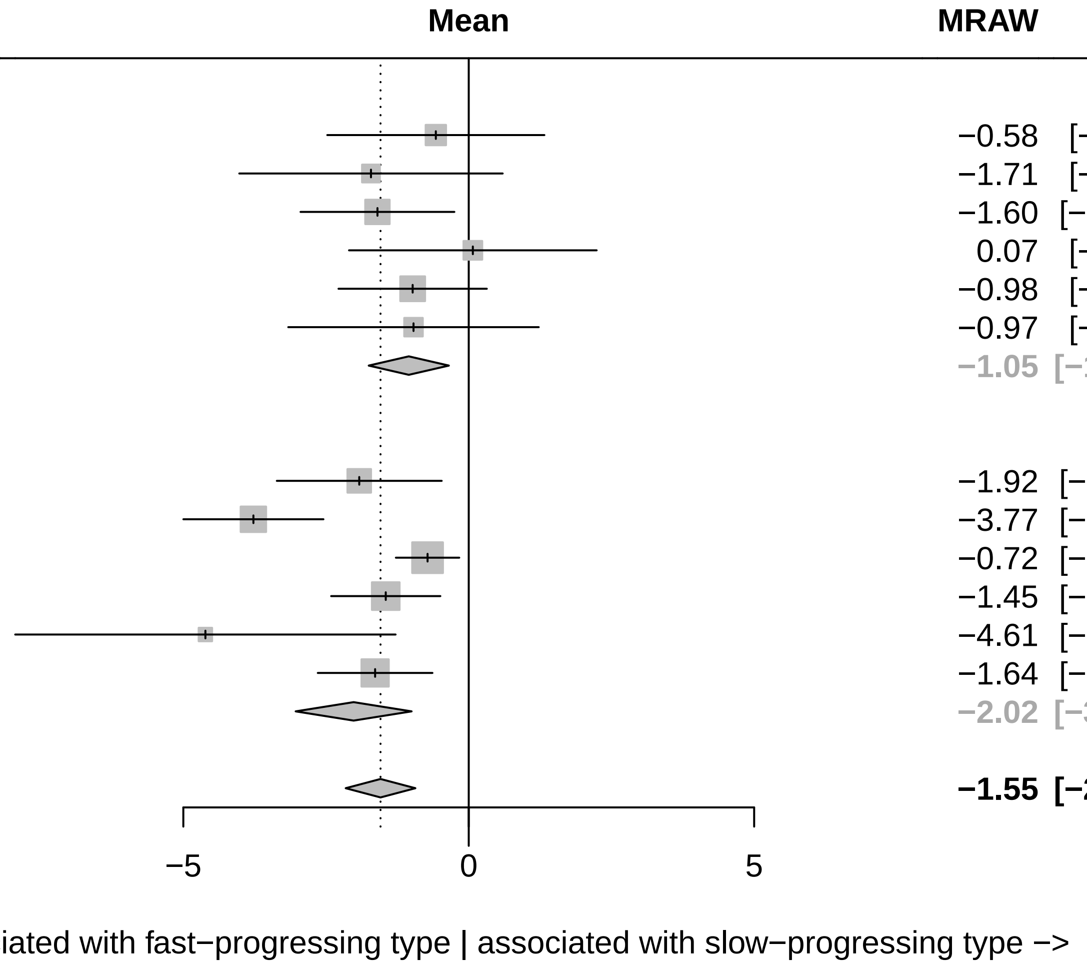

Three−level metaanalysis using random effects to calculate an overall regression coefficient estimate for Overall cognition across cohorts. The dashed line indicates the overall mean estimate. The solid line indicates no effect.

### Forest plot for baseline characteristics of symptom domain Conceptualization (validation)

| Study | Total | Mean | SD | MRAW | 95%−CI |
|---|---|---|---|---|---|
| **Cohort = ICEBERG (validation)** | | | | | |
| FAB Conceptualization | 154 | 1.24 | 13.4546 | 1.24 | [−0.88; 3.37] |
| MATTIS Conzeptualization | 154 | −0.84 | 15.5992 | −0.84 | [−3.30; 1.62] |
| MOCA Abstraction | 154 | 0.59 | 13.7107 | 0.59 | [−1.58; 2.76] |
| **Random effects model** | **462** | | | **0.44** | **[−0.85; 1.73]** |
| **Cohort = LuxPARK (validation)** | | | | | |
| FAB Conceptualization | 236 | −1.04 | 9.0093 | −1.04 | [−2.19; 0.11] |
| MOCA Abstraction | 559 | −0.84 | 6.8609 | −0.84 | [−1.41; −0.27] |
| **Random effects model** | **795** | | | **−0.88** | **[−1.39; −0.37]** |
| **Random effects model** | **1257** | | | **−0.70** | **[−1.18; −0.23]** |

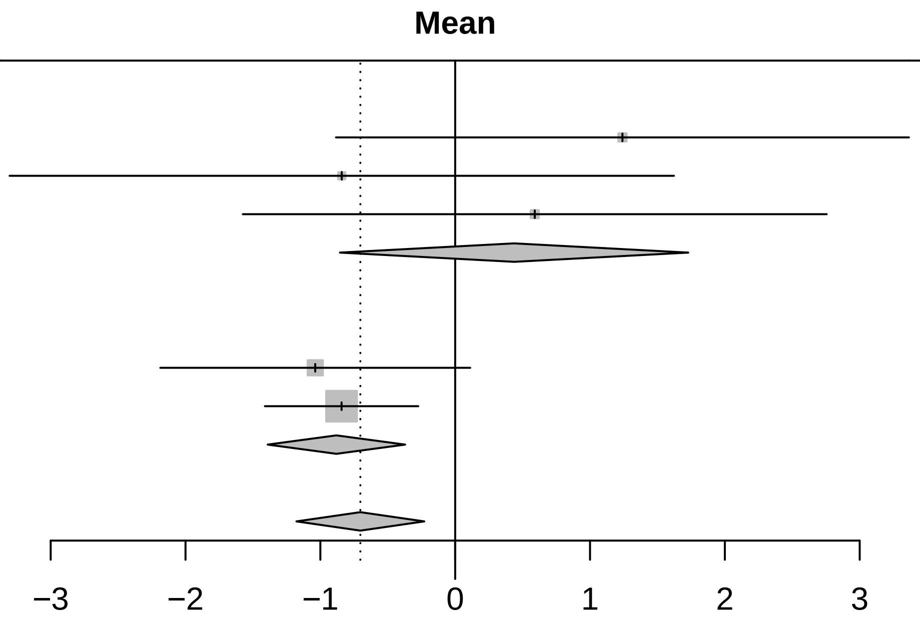

<− Associated with fast−progressing type | associated with slow−progressing type −>

Three−level metaanalysis using random effects to calculate an overall regression coefficient estimate for Conceptualization across cohorts. The dashed line indicates the overall mean estimate. The solid line indicates no effect.

### Forest plot for baseline characteristics of symptom domain Visuo−executive (validation)

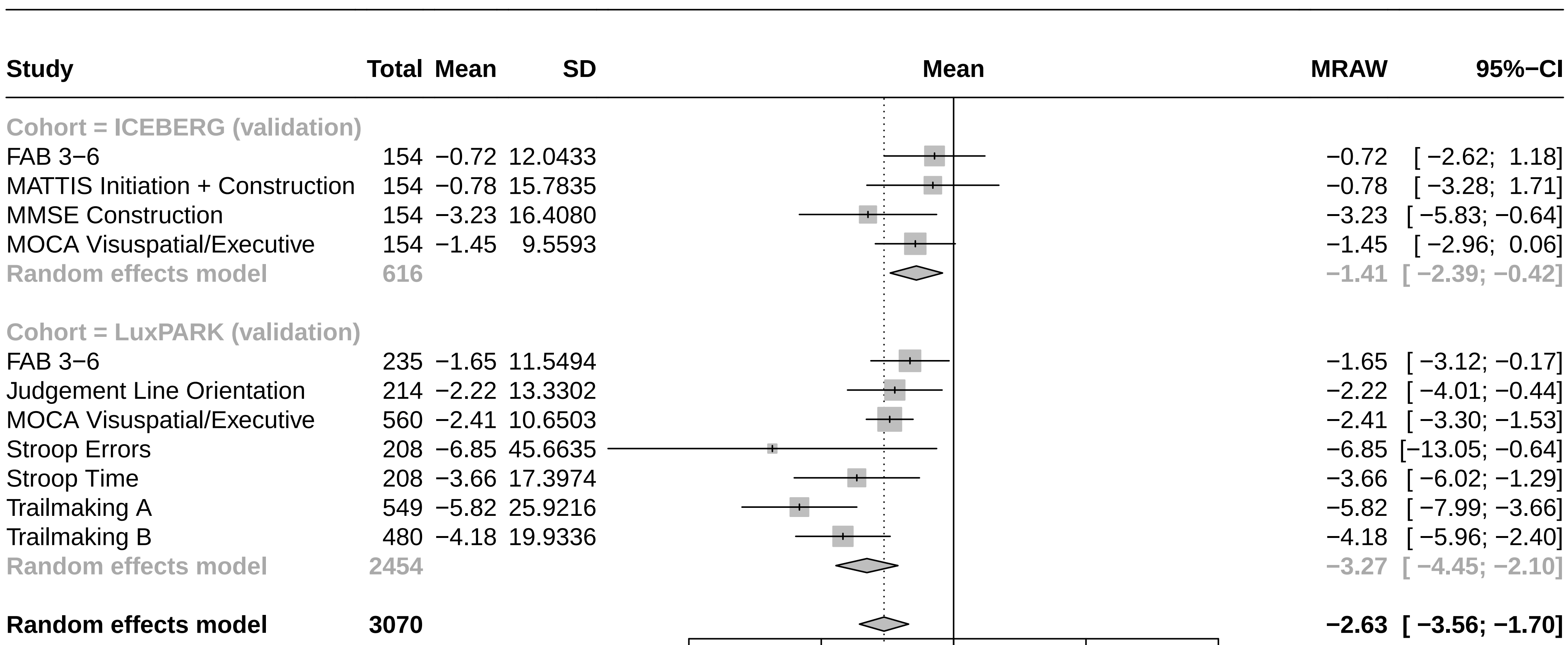

Three−level metaanalysis using random effects to calculate an overall regression coefficient estimate for Visuo−executive across cohorts. The dashed line indicates the overall mean estimate. The solid line indicates no effect.

### Forest plot for baseline characteristics of symptom domain Language (validation)

| Study | Total | Mean | SD | MRAW | 95%−CI |
|---|---|---|---|---|---|
| **Cohort = ICEBERG (validation)** | | | | | |
| FAB VFT | 154 | −0.55 | 10.5285 | −0.55 | [−2.21; 1.11] |
| MMSE Language | 154 | −0.48 | 11.9227 | −0.48 | [−2.36; 1.41] |
| MOCA Language + Naming | 154 | −0.84 | 11.6445 | −0.84 | [−2.68; 1.00] |
| VFT phonematic F | 151 | −1.39 | 15.1994 | −1.39 | [−3.81; 1.03] |
| VFT phonematic S | 143 | −0.53 | 15.4382 | −0.53 | [−3.06; 2.00] |
| VFT semantic supermarket | 151 | −2.42 | 19.5949 | −2.42 | [−5.55; 0.70] |
| **Random effects model** | **907** | | | **−0.84** | **[−1.70; 0.01]** |
| **Cohort = LuxPARK (validation)** | | | | | |
| FAB VFT | 236 | −1.28 | 10.0470 | −1.28 | [−2.56; 0.00] |
| MOCA Language + Naming | 559 | −2.63 | 15.7566 | −2.63 | [−3.94; −1.32] |
| VFT phonematic F | 548 | −3.19 | 17.7775 | −3.19 | [−4.67; −1.70] |
| VFT phonematic S | 221 | −3.32 | 19.1413 | −3.32 | [−5.84; −0.79] |
| VFT semantic sum | 210 | −4.27 | 16.9366 | −4.27 | [−6.56; −1.98] |
| **Random effects model** | **1774** | | | **−2.70** | **[−3.70; −1.71]** |
| **Random effects model** | **2681** | | | **−1.85** | **[−2.62; −1.09]** |

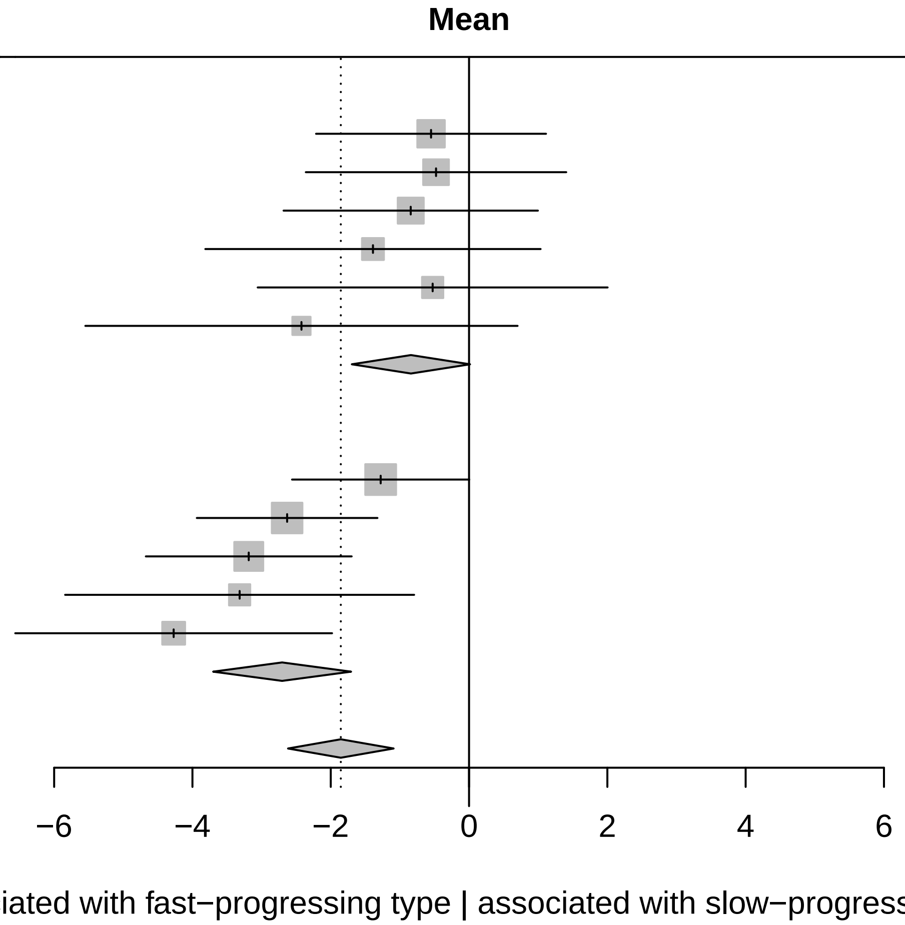

Three−level metaanalysis using random effects to calculate an overall regression coefficient estimate for Language across cohorts.
The dashed line indicates the overall mean estimate. The solid line indicates no effect.

### Forest plot for baseline characteristics of symptom domain Anxiety (validation)

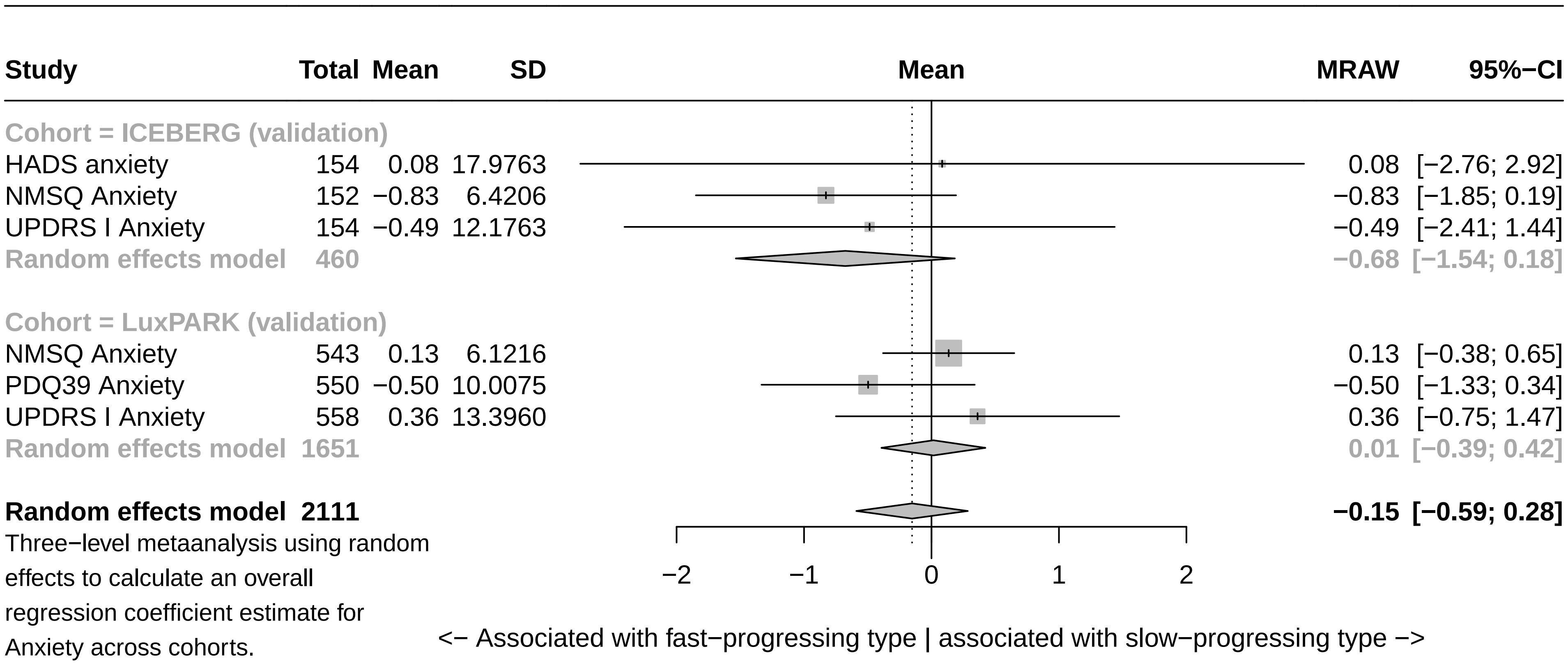

Three−level metaanalysis using random effects to calculate an overall regression coefficient estimate for Anxiety across cohorts.
The dashed line indicates the overall mean estimate. The solid line indicates no effect.

### Forest plot for baseline characteristics of symptom domain Attention (validation)

| Study | Total | Mean | SD | MRAW | 95%−CI |
|---|---|---|---|---|---|
| **Cohort = ICEBERG (validation)** | | | | | |
| MATTIS Attention | 154 | −3.94 | 15.4627 | −3.94 | [−6.38; −1.49] |
| MMSE Attention | 154 | −1.81 | 11.8440 | −1.81 | [−3.68; 0.06] |
| MOCA Attention | 154 | 0.15 | 12.0695 | 0.15 | [−1.76; 2.05] |
| NMSQ Attention | 152 | −0.61 | 6.8590 | −0.61 | [−1.70; 0.48] |
| **Random effects model** | **614** | | | **−1.37** | **[−2.90; 0.16]** |
| **Cohort = LuxPARK (validation)** | | | | | |
| MOCA Attention | 559 | −1.78 | 14.4209 | −1.78 | [−2.97; −0.58] |
| NMSQ Attention | 545 | −0.40 | 5.6128 | −0.40 | [−0.87; 0.07] |
| PDQ39 Attention | 549 | −1.21 | 10.1009 | −1.21 | [−2.05; −0.36] |
| SIQCDE Attention | 233 | −0.84 | 16.8653 | −0.84 | [−3.00; 1.33] |
| Digit Span | 214 | −2.38 | 15.1360 | −2.38 | [−4.41; −0.35] |
| **Random effects model** | **2100** | | | **−1.11** | **[−1.81; −0.40]** |
| **Random effects model** | **2714** | | | **−1.14** | **[−1.74; −0.55]** |

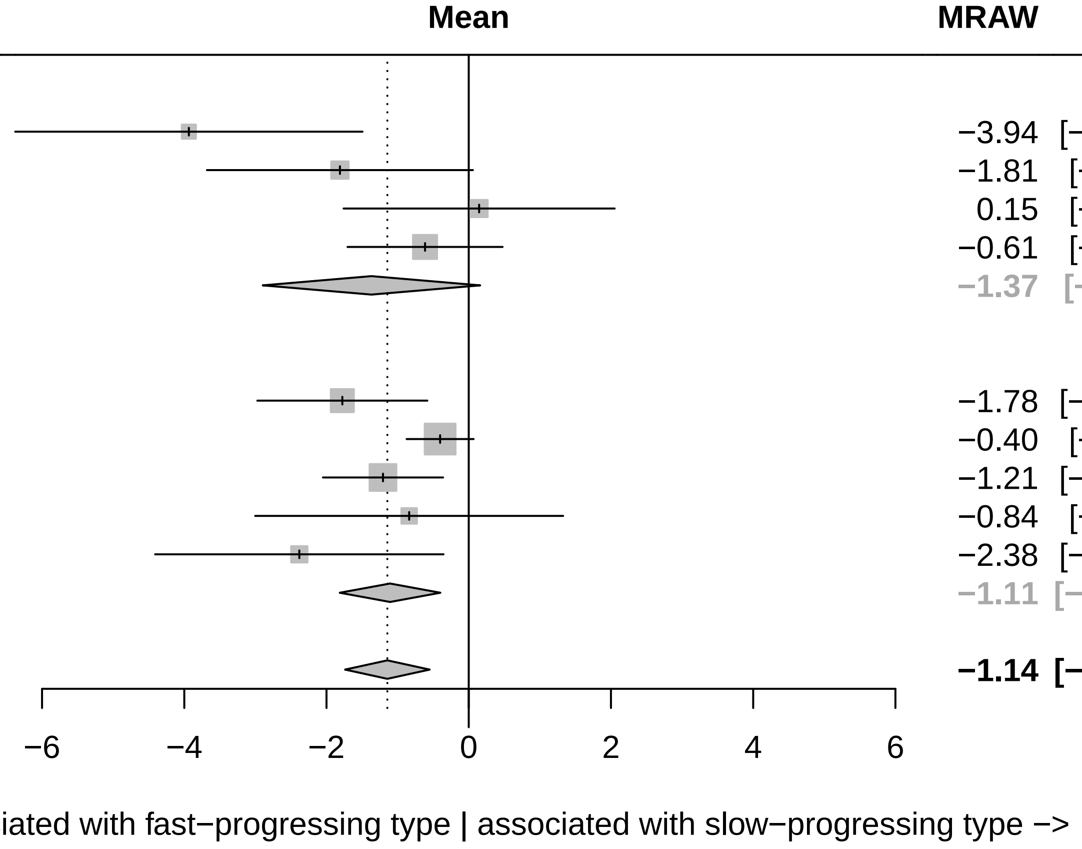

Three−level metaanalysis using random effects to calculate an overall regression coefficient estimate for Attention across cohorts.
The dashed line indicates the overall mean estimate. The solid line indicates no effect.

### Forest plot for baseline characteristics of symptom domain Non motor symptoms (validation)

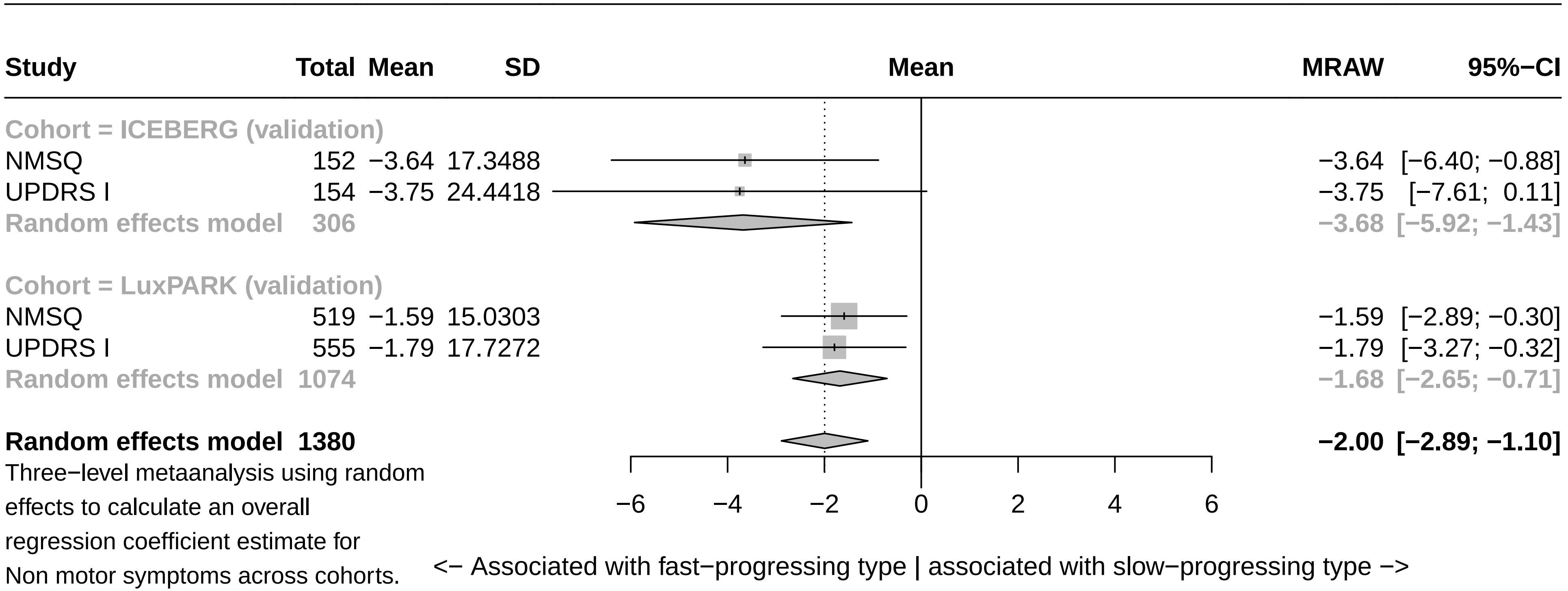

Three−level metaanalysis using random effects to calculate an overall regression coefficient estimate for Non motor symptoms across cohorts. The dashed line indicates the overall mean estimate. The solid line indicates no effect.

### Forest plot for baseline characteristics of symptom domain Autonomic (validation)

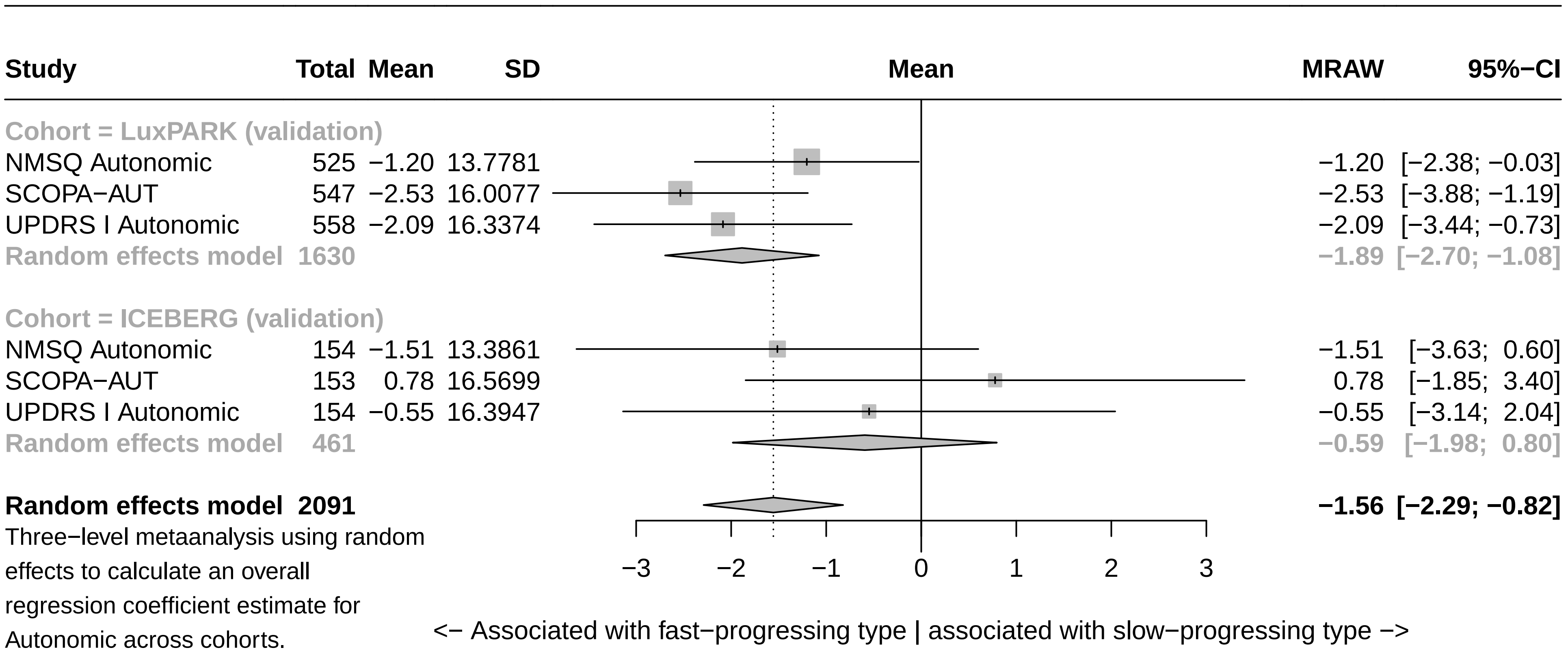

Three−level metaanalysis using random effects to calculate an overall regression coefficient estimate for Autonomic across cohorts.
The dashed line indicates the overall mean estimate. The solid line indicates no effect.

### Forest plot for baseline characteristics of symptom domain Hallucinations (validation)

| Study | Total | Mean | SD | MRAW | 95%−CI |
|---|---|---|---|---|---|
| **Cohort = LuxPARK (validation)** | | | | | |
| NMSQ Hallucination | 544 | −1.11 | 9.9470 | −1.11 | [−1.95; −0.28] |
| UPDRS I Hallucinations | 558 | −0.97 | 17.4361 | −0.97 | [−2.41; 0.48] |
| **Random effects model** | **1102** | | | **−1.08** | **[−1.80; −0.35]** |
| **Cohort = ICEBERG (validation)** | | | | | |
| NMSQ Hallucination | 154 | −1.32 | 10.2670 | −1.32 | [−2.94; 0.30] |
| UPDRS I Hallucinations | 154 | −0.42 | 7.9031 | −0.42 | [−1.67; 0.82] |
| **Random effects model** | **308** | | | **−0.76** | **[−1.75; 0.23]** |
| **Random effects model** | **1410** | | | **−0.96** | **[−1.55; −0.38]** |

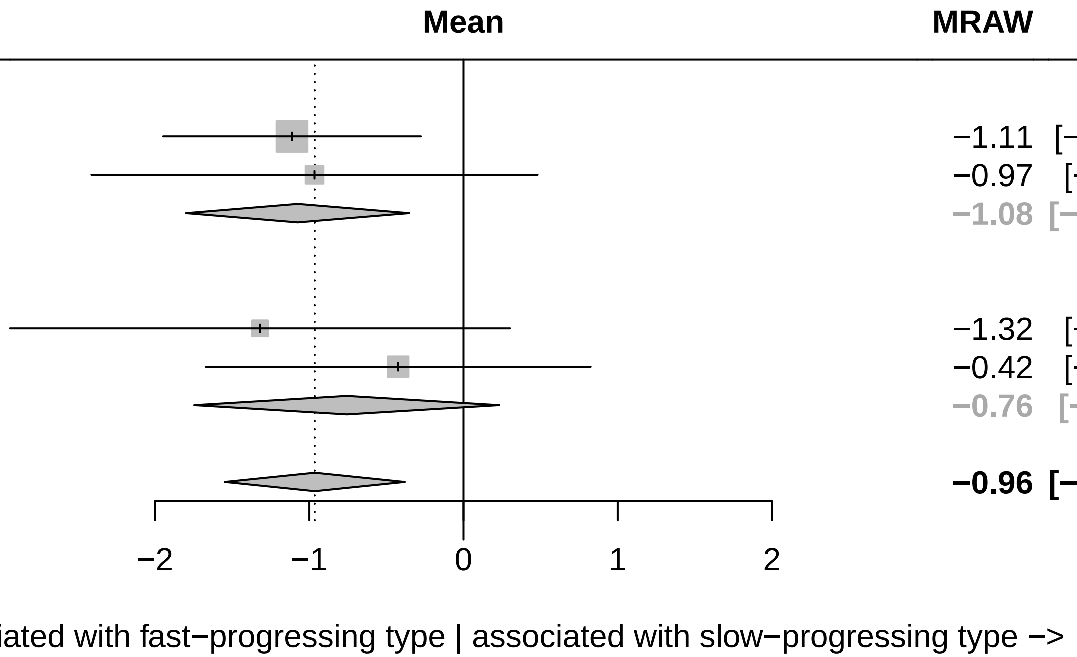

Three−level metaanalysis using random effects to calculate an overall regression coefficient estimate for Hallucinations across cohorts. The dashed line indicates the overall mean estimate. The solid line indicates no effect.

### Forest plot for baseline characteristics of symptom domain Pain (validation)

| Study | Total | Mean | SD | MRAW | 95%−CI |
|---|---|---|---|---|---|
| **Cohort = LuxPARK (validation)** | | | | | |
| NMSQ Pain | 546 | 0.22 | 6.5957 | 0.22 | [−0.33; 0.77] |
| PDQ39 Pain | 547 | −0.21 | 10.8280 | −0.21 | [−1.12; 0.70] |
| UPDRS I Pain | 558 | −0.37 | 10.1509 | −0.37 | [−1.22; 0.47] |
| **Random effects model** | **1651** | | | **−0.01** | **[−0.42; 0.40]** |
| **Cohort = ICEBERG (validation)** | | | | | |
| NMSQ Pain | 152 | −0.44 | 6.3159 | −0.44 | [−1.44; 0.57] |
| UPDRS I Pain | 154 | −2.83 | 14.6445 | −2.83 | [−5.15; −0.52] |
| **Random effects model** | **306** | | | **−1.40** | **[−3.70; 0.90]** |
| **Random effects model** | **1957** | | | **−0.22** | **[−0.67; 0.24]** |

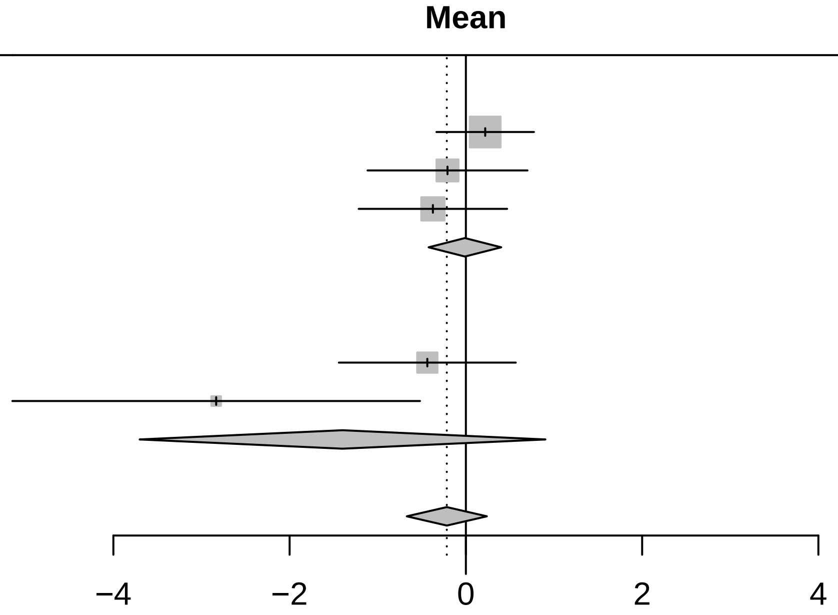

<− Associated with fast−progressing type | associated with slow−progressing type −>

Three−level metaanalysis using random effects to calculate an overall regression coefficient estimate for Pain across cohorts.
The dashed line indicates the overall mean estimate. The solid line indicates no effect.

### Forest plot for baseline characteristics of symptom domain RBD (validation)

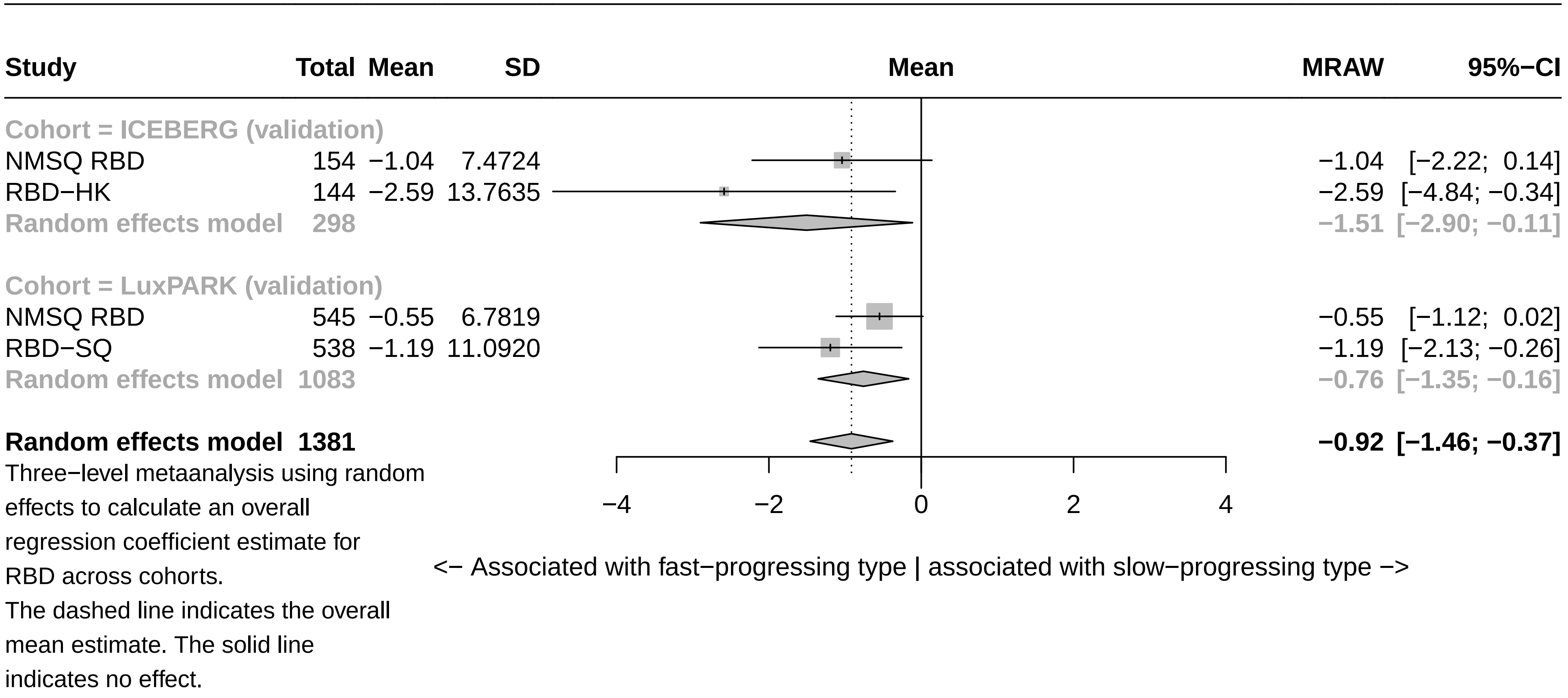

### Forest plot for baseline characteristics of symptom domain Smell (validation)

| Study | Total | Mean | SD | MRAW | 95%−CI |
|---|---|---|---|---|---|
| **Cohort = ICEBERG (validation)** | | | | | |
| NMSQ Smell | 152 | −0.46 | 6.3205 | −0.46 | [−1.47; 0.54] |
| UPSIT | 146 | 0.56 | 13.9305 | 0.56 | [−1.70; 2.82] |
| **Random effects model** | **298** | | | **−0.29** | **[−1.21; 0.62]** |
| **Cohort = LuxPARK (validation)** | | | | | |
| NMSQ Smell | 546 | −0.25 | 5.3725 | −0.25 | [−0.70; 0.21] |
| Sniffin Test | 541 | −1.47 | 12.1039 | −1.47 | [−2.49; −0.45] |
| **Random effects model** | **1087** | | | **−0.77** | **[−1.95; 0.42]** |
| **Random effects model** | **1385** | | | **−0.53** | **[−1.17; 0.11]** |

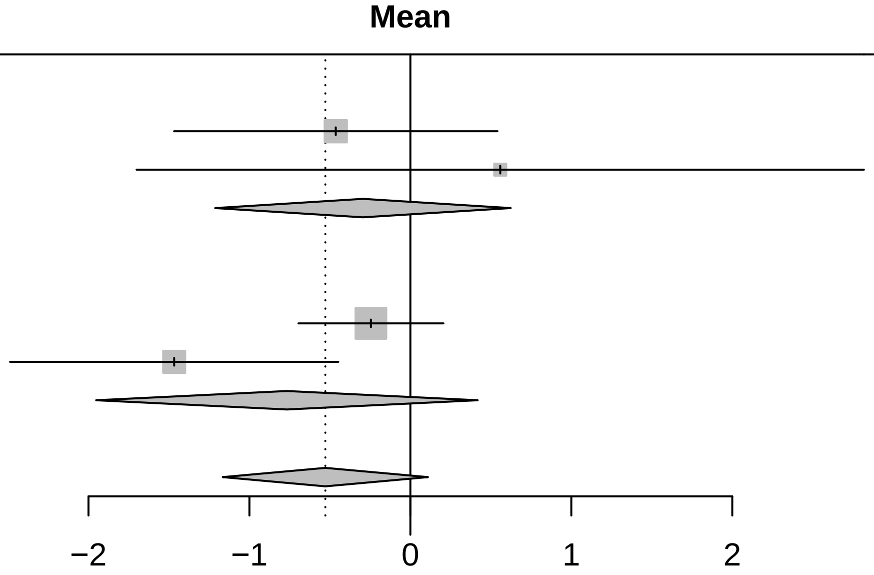

<− Associated with fast−progressing type | associated with slow−progressing type −>

Three−level metaanalysis using random effects to calculate an overall regression coefficient estimate for Smell across cohorts.
The dashed line indicates the overall mean estimate. The solid line indicates no effect.

### Forest plot for baseline characteristics of symptom domain Motor symptoms (validation)

| Study | Total | Mean | SD | MRAW | 95%−CI |
|---|---|---|---|---|---|
| **Cohort = LuxPARK (validation)** | | | | | |
| PDQ39 ADL | 548 | −3.01 | 12.9061 | −3.01 | [−4.09; −1.93] |
| Pegboard | 539 | −5.08 | 18.8444 | −5.08 | [−6.67; −3.48] |
| UPDRS II | 555 | −3.34 | 17.0696 | −3.34 | [−4.76; −1.92] |
| UPDRS III on | 468 | −2.49 | 16.6776 | −2.49 | [−4.00; −0.98] |
| UPDRS IV | 556 | 0.74 | 18.2493 | 0.74 | [−0.78; 2.26] |
| **Random effects model** | **2666** | | | **−2.63** | **[−4.46; −0.81]** |
| **Cohort = ICEBERG (validation)** | | | | | |
| UPDRS II | 154 | 1.01 | 16.5643 | 1.01 | [−1.60; 3.63] |
| UPDRS III off | 154 | −1.53 | 20.9631 | −1.53 | [−4.84; 1.78] |
| UPDRS III on | 131 | 0.02 | 12.8990 | 0.02 | [−2.19; 2.23] |
| UPDRS IV | 154 | −2.18 | 26.0911 | −2.18 | [−6.30; 1.94] |
| **Random effects model** | **593** | | | **−0.23** | **[−1.64; 1.18]** |
| **Random effects model** | **3259** | | | **−1.86** | **[−3.28; −0.45]** |

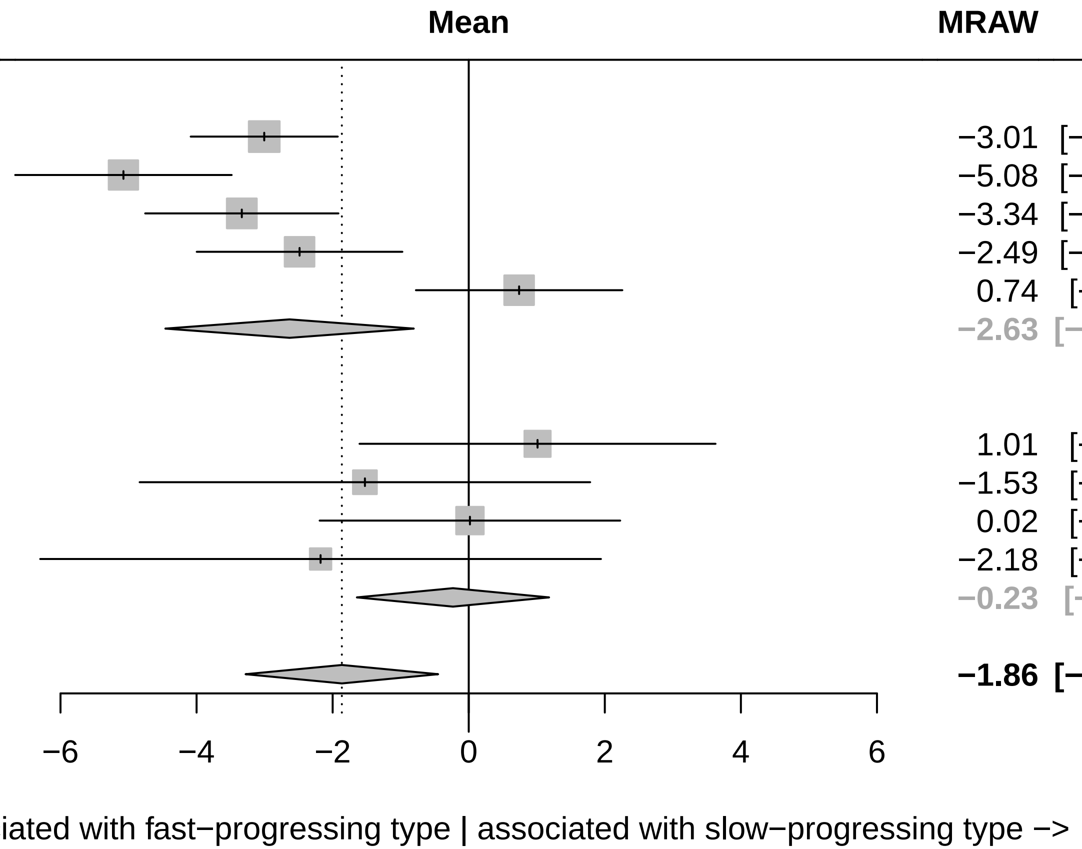

Three−level metaanalysis using random effects to calculate an overall regression coefficient estimate for Motor symptoms across cohorts. The dashed line indicates the overall mean estimate. The solid line indicates no effect.

Forest plot for baseline characteristics of symptom domain Impulsivity (validation)

| Study | Total | Mean | SD | MRAW | 95%−CI |
|---|---|---|---|---|---|
| **Cohort = LuxPARK (validation)** | | | | | |
| QUIP | 211 | −1.31 | 13.5582 | −1.31 | [−3.14; 0.52] |
| **Cohort = ICEBERG (validation)** | | | | | |
| QUIP−RS | 154 | 6.31 | 52.0460 | 6.31 | [−1.91; 14.53] |
| **Random effects model** | **365** | | | **1.40** | **[−5.75; 8.55]** |

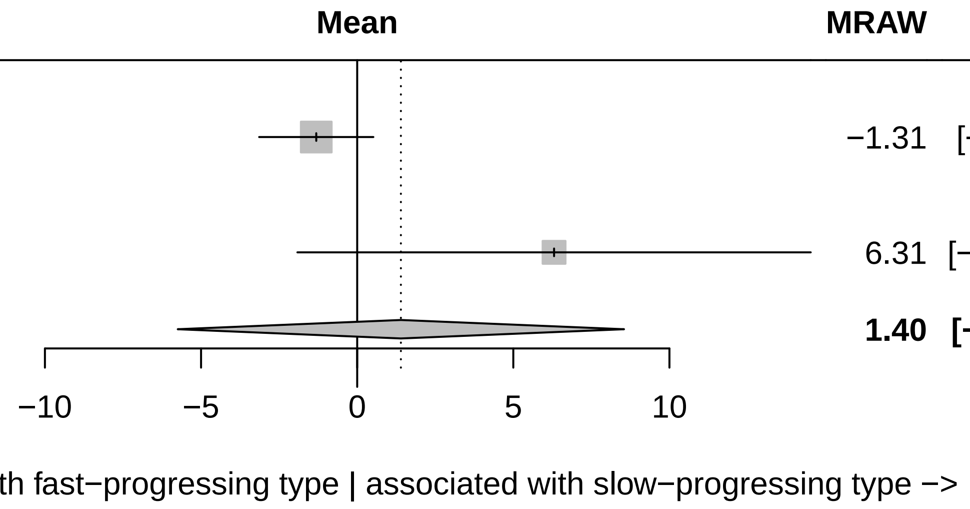

Three−level metaanalysis using random effects to calculate an overall regression coefficient estimate for Impulsivity across cohorts.
The dashed line indicates the overall mean estimate. The solid line indicates no effect.

### Forest plot for baseline characteristics of symptom domain Tremor (validation)

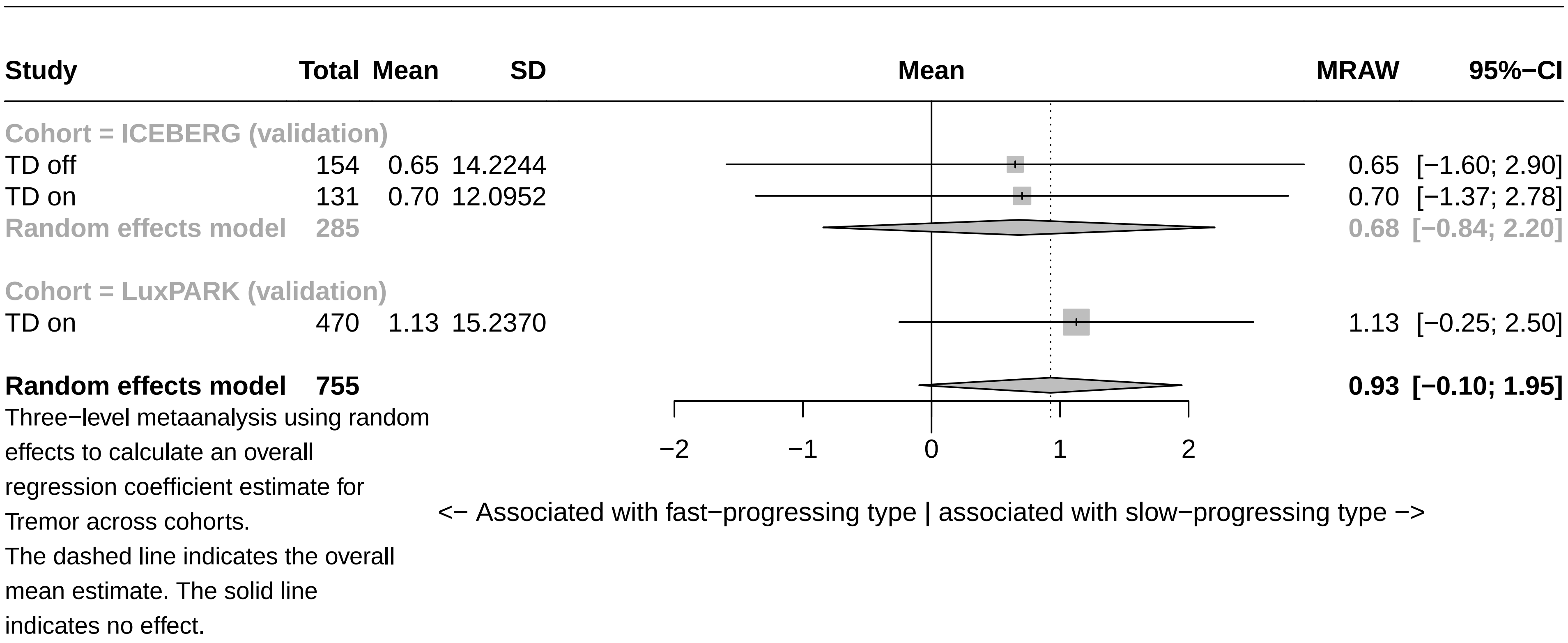

### Forest plot for baseline characteristics of symptom domain Fatigue (validation)

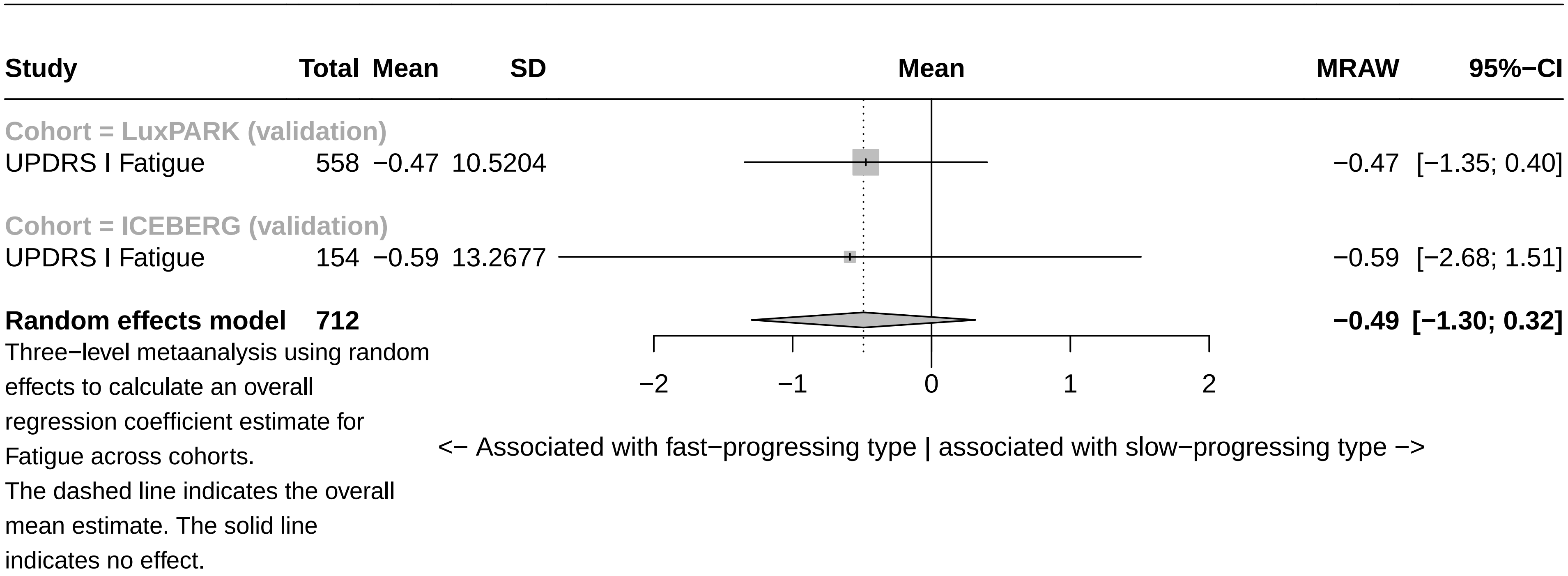
